# Event-Proximity Organ Clocks in Humans Link Multi-organ Biology to Disease and Mortality

**DOI:** 10.64898/2026.11.25.26369010

**Authors:** The MULTI Consortium, Ying Chen, Zhiyuan Song, Qiangqiang Wu, Grace McIlvain, Xia Zhou, Zhou Yu, Seyedmehdi Payabvash, Agustin Ibanez, Andrew Zalesky, Christos Davatzikos, Paul Aisen, Reisa Sperling, Keenan A. Walker, Eleanor M. Simonsick, Susan M. Resnick, Luigi Ferrucci, Junhao Wen

## Abstract

Biological aging clocks^1–7^ have transformed aging research by quantifying individual variation in aging. However, these clocks are trained to estimate chronological age and therefore capture broad aging variance rather than direct biological proximity to adverse clinical events. Here, we introduce **EPOCH** (**E**vent-**P**roximity **O**rgan **C**locks in **H**umans), a general framework that learns organ-and omics-specific signatures of proximity to future disease onset and mortality. We first develop mortality EPOCH clocks from organ-specific imaging, proteomic, and metabolomic features to model proximity to all-cause mortality. These clocks reveal a pervasive, interconnected whole-body genetic and clinical risk architecture linked to systemic disease endpoints. Critically, mortality EPOCH clocks outperform biological aging clocks in predicting future disease onset, while their complementary biology enables the joint modeling of aging resilience, such as individuals with advanced biological aging but unexpectedly low mortality vulnerability. Longitudinal mortality EPOCHs capture within-person longitudinal changes, with longitudinal changes providing information beyond baseline EPOCH. Finally, we demonstrate the broad applicability and generalizability of EPOCH through disease-specific clocks spanning neurodegenerative, respiratory, vascular, and cardiometabolic conditions. Together, EPOCH represents a scalable, organ-and modality-resolved biological clock framework that extends and complements, rather than replaces, existing biological aging clocks for adverse-event prognostication and whole-body modeling.

## Main

Biological aging clocks^1–12^ have transformed biomedicine by providing a quantitative means of measuring biological aging beyond chronological age. These aging clocks, often called first-generation clocks, span epigenetic, imaging, proteomic, metabolomic, phenotypic, and behavioral modalities, and advance the biological aging field by showing that complex human biology can be compressed into an individualized single-scalar estimate of biological age^13,14^. Yet most of these clocks are built to predict chronological age from a single biological snapshot using cross-sectional data, defining the residual between predicted and chronological age as “age deceleration/acceleration” as biological age gaps (BAG). This design has some intrinsic limitations. These clocks are optimized to predict chronological age, a biological process expressed continuously across the lifespan and intertwined with broad, multifaceted variation in human health^15^; therefore, they are not optimized to directly capture risk for specific clinical outcomes (e.g., dementia onset). Rather, disease incidence and mortality are typically evaluated only post hoc, after first-generation clocks have been developed^8^. In addition, cross-sectional age prediction models primarily capture population-level age variance at baseline, making it difficult to distinguish longitudinal dynamic aging from pre-existing inter-individual differences, a challenge recently formalized as the “baseline effect^16^” in brain aging models.

These limitations have motivated the development of the next generation of biological clocks that directly *i*) incorporate adverse clinical events into model training; and *ii*) incorporate longitudinal information (e.g., Cox survival-based “time-to-event” modeling) to estimate each individual’s evolving proximity to, and risk of, future clinical events. Therefore, these clinical endpoint-informed clocks aim to expand the biological aging field from age prediction toward clinically anchored measures of biological risk. For example, DNAm PhenoAge^17^ established an important precedent by deriving a mortality-informed phenotypic age from clinical biomarkers and then training a DNA methylation model to reproduce this risk-enriched phenotype. DNAm GrimAge and GrimAge2^18,19^ went further by using DNA methylation surrogates of plasma proteins, smoking exposure, and demographic factors in Cox-based models of time-to-death, making them canonical survival-trained epigenetic clocks. Beyond DNA methylation, UK Biobank (UKBB) plasma proteomics studies have developed organ-specific mortality models using Cox elastic net^20^, and transcriptomic expected-mortality clocks^21^ have further highlighted the growing interest in modeling lifespan and time-to-death directly. In parallel, pace-of-aging frameworks such as DunedinPACE^22^ and DunedinPACNI^23^ estimate longitudinal aging rate rather than chronological age itself. Together, these approaches move beyond static age prediction and anchor biological clocks to mortality, lifespan, functional decline, or longitudinal aging rate. **Supplementary Tables 1-2** present an overview of the state-of-the-art biological clock research.

Concurrently, a complementary frontier has emerged in biological aging and longevity research that models biological aging in an organ-specific fashion^8,10,20,24^. These organ-specific clocks are built on two central principles. First, organs and biological systems age at different rates^24^, generating substantial heterogeneity across individuals; and second, no organ system is an island^13,25^, because aging in one tissue may reflect, influence, or amplify vulnerability in another. We and others have advanced this first-generation clock framework by developing organ-specific aging clocks from *in vivo* imaging^2,4,6,14,26^, circulating plasma proteomics^5,8,10,20,27–29^, metabolomics^3^, and other behavioral^11^ and clinical phenotypes^24^, enabling biological aging to be mapped across the brain, heart, liver, kidney, immune system, endocrine system, lung, adipose tissue, and other organ systems.

Here, we introduce **EPOCH** (**E**vent-**P**roximity **O**rgan **C**locks in **H**umans), a survival-based framework that generalizes prior second-generation clocks on proximity to future adverse clinical events, including disease onset and mortality, and extends this event-centered approach across multiple organs and omics. Of note, “event-proximity” denotes the relative event-associated hazard captured by the Cox-derived score, rather than an absolute event probability or the literal time remaining until an event. The year-scale EPOCH measures are age-referenced transformations of this relative hazard signal and should not be interpreted as estimates of years to event (**Method 2**). EPOCH does not represent the first mortality-informed clock; prior studies have developed related models, including prior epigenetic clocks^18,19^ and organ-specific proteomic mortality clocks^20^. Therefore, the principal innovation and advance of EPOCH is to unify and extend survival-based modeling into a multi-organ, multi-omics framework that is broadly generalizable and applicable across an array of future clinical events and diverse datasets.

We first established the EPOCH framework in UKBB^30^ by developing multi-organ mortality EPOCH clocks from *in vivo* imaging, plasma proteomics, and metabolomics. We then demonstrated its disease-specific generalizability across 5 major clinical domains (i.e., all-type dementia, asthma, chronic obstructive pulmonary disease (COPD), stroke, and myocardial infarction (MI)) in UKBB (**Method 1** and **Supplementary Table 3**). Together, this work positions EPOCH as organ-and omics-resolved biological clocks that directly connect multi-organ human biology to future risk of adverse clinical events. We disseminate all our research findings, code, and GWAS summary statistics at the MEDICINE portal: https://labs-laboratory.com/medicine/.

## Results

### Mortality EPOCH clocks in the UK Biobank

In UKBB, we fit elastic-net regularized Cox proportional hazards models (**Method 2**) using baseline organ-specific features as predictors and all-cause mortality as the time-to-event outcome (**Fig. 1A**). The derived EPOCH deceleration/acceleration at the year scale (**Method 2**; thereafter EPOCH clocks) quantifies how relatively close an individual’s organ-specific biological profile is to future mortality risk. **Fig. 1A** illustrates the modeling rationale between EPOCH and BAG.

**Figure 1:**
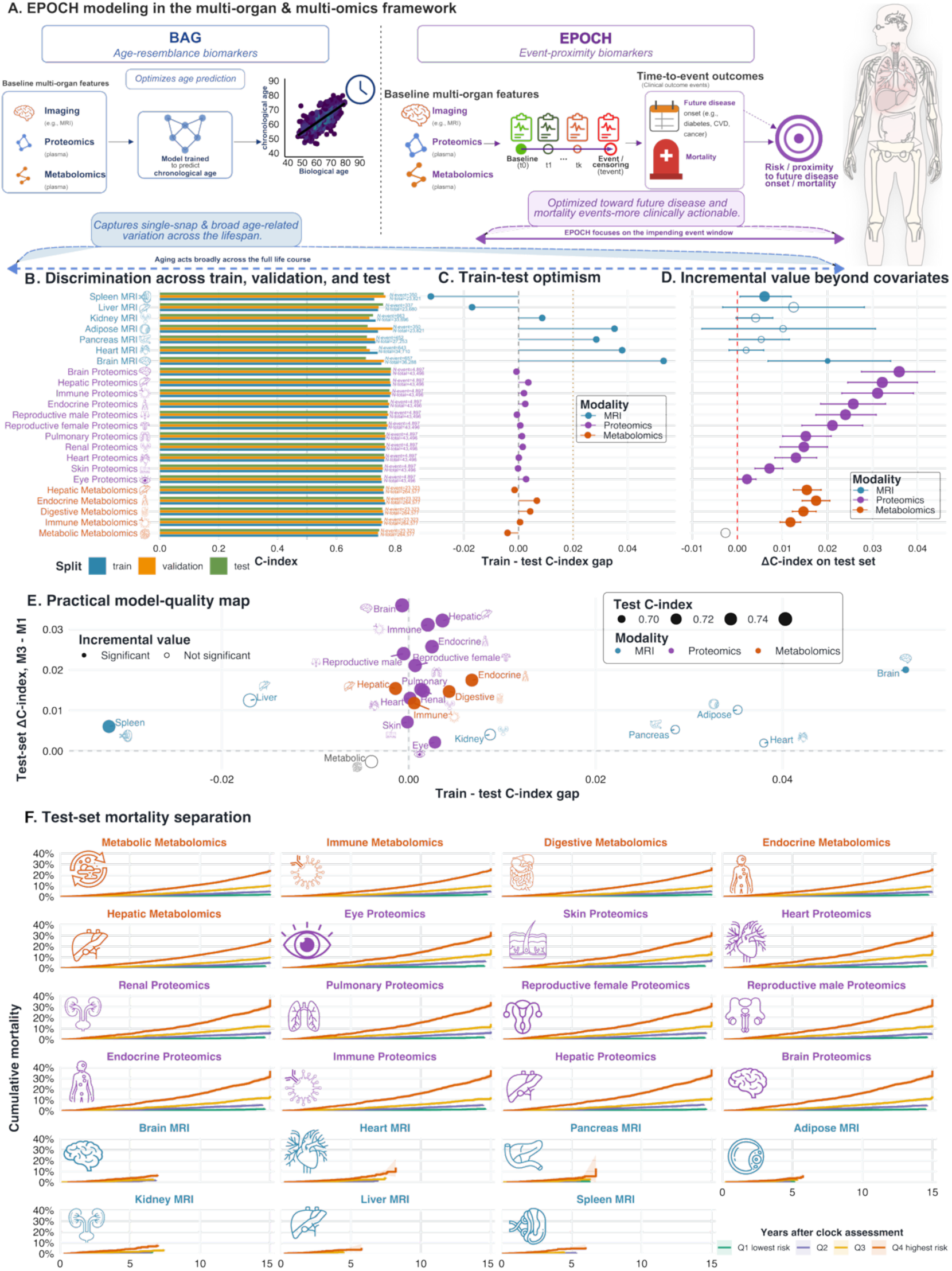
EPOCH modeling framework and model performance of the 23 mortality clocks. **A**) Modeling distinctions between first-generation biological aging clocks (BAGs) and next-generation EPOCH. First-generation clocks are trained to predict chronological age and derive BAGs, thereby capturing broad age-related variation across the lifespan. In contrast, EPOCH leverages baseline multi-organ or multi-omics features together with longitudinal time-to-event outcomes, often spanning more than a decade of follow-up, to learn biomarkers that capture proximity to future clinical events. The life-course schematic illustrates that aging broadly influences health trajectories, whereas direct modeling of future disease and mortality events yields a more focused, clinically actionable biomarker framework. **B-C**) C-index values for MRI-, proteomics-, and metabolomics-based mortality EPOCH across training, validation, and held-out test sets. Similar discrimination across splits indicates generalizable event-risk prediction, whereas large differences between training and test performance would suggest potential overfitting. **D**) Test-set improvement in C-index after adding each EPOCH score (M3) to the covariate-only baseline model (M1). Positive ΔC-index values indicate that the clock contributes predictive information beyond demographic and clinical covariates. Filled points indicate statistically significant incremental value, whereas open points indicate nonsignificant improvement (e.g., metabolomics-based metabolic EPOCH). **E**) Integrated assessment of model utility by jointly displaying train-test optimism and test-set incremental value. **F**) Kaplan-Meier cumulative mortality curves in the held-out test set stratified by training-defined EPOCH risk quartiles. Across modality clocks, higher-risk quartiles generally show greater cumulative mortality over follow-up, demonstrating prospective mortality-risk separation. Quartiles were defined using the clock score distribution in the training set and then applied to the test set.

We first evaluated the model performance for 7 MRI-, 11 plasma proteomics-, and 5 metabolomics-based models. In the held-out test dataset, the EPOCH clocks achieved moderate discrimination for all-cause mortality, with C-indices ranging from 0.700 to 0.785 and a median of 0.759. Proteomics-based clocks (*N∼*50,000) showed the most consistent performance, with test C-indices from 0.751 to 0.785, led by brain, hepatic, and immune proteomics-based EPOCH clocks. Metabolomics-based clocks (*N>*270,000) also generalized well, with test C-indices from 0.746 to 0.762. In contrast, MRI-based clocks showed more heterogeneous performance, likely reflecting relatively smaller sample sizes (20,000<*N*<40,000) and shorter follow-up (around 5 years) in the imaging subsets (**Fig. 1B**). Train-test differences were generally small, particularly for proteomics and metabolomics clocks (**Fig. 1C**). Incremental prediction value showed that 17 of 23 clocks significantly improved test-set discrimination beyond covariate-baseline models (**Fig. 1D-E**). Finally, we observed that the metabolomics-based metabolic EPOCH clock showed poor year-scale calibration, with deceleration/acceleration estimates falling outside a biologically interpretable range (>100 years; **Supplementary Figure 1**), which was discarded for downstream analyses. Kaplan-Meier analyses in the held-out test datasets further confirmed that mortality EPOCH clocks stratified future mortality risk (**Fig. 1F**). Calibration slopes were close to the ideal value of 1 for the molecular EPOCHs, with mean slopes of 0.990±0.016 for metabolomics, 0.991±0.009 for proteomics, and 0.951±0.134 for MRI-based clocks. **Extended Data Figure 1** shows the distribution overlaps of the EPOCH clocks across training, validation, and test datasets. We also investigated the biological pathways underlying EPOCH using the brain proteomics-based EPOCH as a representative example (**Supplementary Figure 2**), highlighting neuronal development, axonogenesis, neurogenesis, and synaptic transmission and plasticity, with preferential enrichment in brain and central nervous system tissues.

We developed 22 mortality EPOCH clocks and demonstrated their incremental predictive value for all-cause mortality. In downstream analyses, we further characterized their genetic architecture.

### Genetic architecture of the mortality EPOCH clocks

We conducted GWAS (**Method 3a**) for the 22 mortality EPOCH clocks (19,008<N<210,481 participants with European ancestry) and identified 365 (P-value<5×10^-^8/22) genomic locus-EPOCH pairs. The strongest locus burden was observed for metabolomics mortality clocks (201 loci), followed by proteomics (123 loci) and MRI (41 loci) (**Fig. 2A**). We denoted the genomic loci using their top lead SNPs and cytogenetic regions defined by FUMA (**Supplementary Note 1**), considering linkage disequilibrium (LD). **Supplementary Figure 25** illustrates brain-body genetic signals associated with the brain proteomics mortality EPOCH, highlighting convergence across neurodegenerative, vascular, immune/inflammatory, and metabolic pathways that may jointly contribute to mortality risk and echoing the proteomic evidence (**Supplementary Figure 2**). The LDSC^31^ intercept (LDSC*_b_*=1.025 [1.011, 1.038]) of the EPOCH clocks was close to 1, and the LDSC ratio was small, indicating no severe inflation due to population stratification. **Supplementary Table 4** shows the detailed statistics from LDSC. **Extended Data Figure 2** presents the trumpet plots of the effective allele frequency vs. the *β* coefficients.

**Figure 2:**
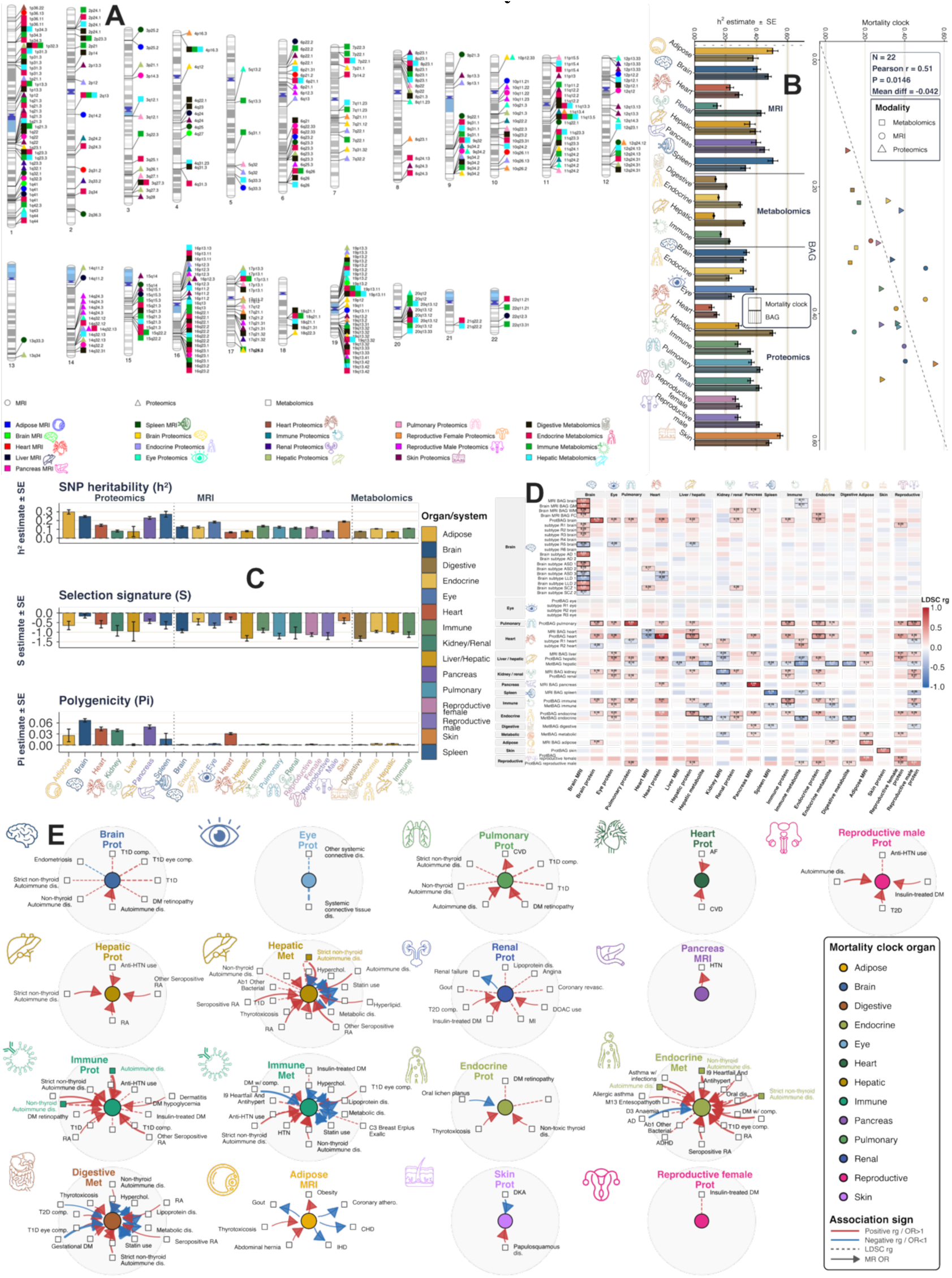
Genetic architecture of the 22 mortality EPOCH clocks. **A**) Cytogenetic regions associated with the 22 EPOCH mortality clocks, with significant genomic loci identified using the genome-wide significance threshold (two-sided P-value<5×10^-8^/22). **B**) Matched comparison of mortality EPOCH and BAG SNP-based heritability estimates (*h^2^*) across organ-specific pairs. GCTA/HEreg SNP heritability estimates were compared between mortality EPOCH clocks and their corresponding BAGs across 22 matched organ pairs. Bars show organ-specific *h^2^* estimates ± SE, with mortality EPOCH clocks shown as solid bars and BAGs shown as striped bars using consistent organ/system colors. The paired scatter plot summarizes concordance between BAG *h^2^* and mortality EPOCH *h^2^* estimates across MRI, proteomic, and metabolomic clocks, with the dashed line indicating equality between paired estimates. **C**) SBayesS was used to estimate three genetic-architecture parameters for 22 mortality EPOCH clocks across MRI, proteomic, and metabolomic modalities: SNP-based heritability (*h^2^*), selection signature (*S*), and polygenicity (*Pi*). Bars show parameter estimates ± SE for each organ-specific EPOCH clock, grouped by modality and colored by organ/system using a consistent palette. Vertical dotted lines separate MRI, proteomics, and metabolomics clocks. **D**) LDSC genetic correlation (*r_g_*) was estimated between 22 organ-specific mortality EPOCH clocks and 47 previously developed AI biomarkers, including multi-organ BAGs and disease subtypes. Rows represent AI biomarkers and columns represent mortality EPOCH clocks, with both axes grouped by organ/system. Tile color indicates the magnitude and direction of LDSC genetic correlation: red indicates positive and blue indicates negative. Significant cells are annotated as: * nominal two-sided P < 0.05; ** Bonferroni correction across 22 mortality EPOCH clocks, two-sided P < 0.05/22 = 2.27 × 10^-3^; and *** Bonferroni correction across all 22 × 47 mortality EPOCH-AI biomarker tests, two-sided P < 0.05/1034 = 4.84 × 10^-5^. **E**) A hub-and-spoke network was used to summarize genetic relationships between 22 organ-specific mortality EPOCH clocks and 527 disease endpoints (DEs) using both LDSC genetic correlation and bidirectional Mendelian randomization (MR). Each mortality EPOCH clock is shown as a central hub, with associated DEs arranged as satellite nodes surrounding it. Dotted edges indicate non-directional LDSC genetic correlations, whereas solid arrows indicate MR-supported causal directions from EPOCH to DE (*EPOCH2DE*) or from DE to EPOCH (*DE2EPOCH*); bidirectional MR relationships are shown as paired curved arrows. Red edges indicate positive associations, defined as *r_g_* > 0 for LDSC or odds ratio (OR) > 1 for MR, and blue edges indicate negative associations. We present only Bonferroni-corrected significant associations, using the number of valid DEs tested separately in the LDSC and MR analyses as the correction denominator (two-sided P < 0.05/number of valid DEs).

We estimated the SNP-based heritability (*h*^2^*_SNP_*) for the 22 mortality EPOCH clocks using the GCTA^32^ (**Method 3b**), with estimates ranging from 0.11 to 0.55 across clocks. We next compared these estimates with the matched BAGs, whose heritability estimates ranged from 0.14 to 0.50. Overall, mortality EPOCH and BAG clocks showed a moderate correlation in heritability estimates (Pearson’s *r*=0.51), indicating partially shared but non-identical genetic architectures (**Fig. 2B** and **Supplementary Table 5**). In addition, we used SBayesS^33^ to estimate two additional genetic parameters: polygenicity (*Pi*) and selection signature (*S*). Across clocks, *Pi* ranged from 0.0003 to 0.0669 with a mean of 0.013±0.020, while *S* was consistently negative, with a mean of-0.84±0.34 (**Fig. 2C**), suggesting that mortality EPOCH clocks are polygenic traits shaped by negative selection, with larger effects tending to occur among lower-frequency variants (**Extended Data Figure 2**). **Supplementary Figure 26** presents the comparative results of 3 heritability methods, including GCTA, SBayesS, and LDSC (**Supplementary Tables 4**-**6**).

We then performed genetic correlations (*r_g_*; **Method 3c**) between the 22 mortality EPOCH clocks and 46 previously developed multi-organ AI biomarkers from our previous studies, including first-generation BAGs^2–6^ and disease subtypes^34–37^ (**Supplementary Note 2** denotes the details of these AI biomarkers). We found that 33 associations survived Bonferroni correction across all tests (two-sided P<0.05/22/46). In total, 20 of the 33 Bonferroni-significant associations linked mortality EPOCH clocks to AI biomarkers from the same organ or system, whereas 13 represented cross-organ connections. For example, strong within-organ correlations were observed between the pulmonary proteomics mortality EPOCH and pulmonary ProtBAG (*r_g_=*0.85±0.04; P-value=1.22×10^-104^). Cross-organ signals were also observed, such as the hepatic proteomics mortality EPOCH with heart ProtBAG (*r_g_=*0.43±0.10; P-value=1.02×10^-5^) (**Fig. 2D**).

We finally performed a two-layer analysis (**Method 3d**) to depict the relationships between the 22 mortality EPOCH clocks and 527 disease endpoints (DEs) using FinnGen and PGC GWAS summary statistics (**Fig. 2E**). For genetic correlation, 52 EPOCH-DE associations survived Bonferroni correction (P<0.05/527=9.49×10^-5^), spanning 11 mortality EPOCH clocks and 30 unique DEs. These associations were enriched for immune, endocrine, hepatic, pulmonary, renal, and brain-related axes, such as ADHD with the endocrine metabolomics mortality EPOCH clock (*r_g_=*0.18±0.04; P-value=8.08×10^-6^), autoimmune disease, type 1 diabetes and diabetic complications, hypertension/antihypertensive use, MI, gout, and retinopathy (**Extended Data Figure 3**). For bidirectional Mendelian randomization, we identified 108 Bonferroni-corrected potentially causal signals, including 24 EPOCH→DE signals and 84 DE→EPOCH signals, with 13 bidirectional clock-disease pairs. EPOCH→DE signals were concentrated in metabolomics and MRI clocks and involved lipid/metabolic, statin-use, obesity, gout, renal failure, and cardiovascular endpoints, whereas DE→EPOCH signals were broader and included autoimmune/inflammatory, cardiometabolic, hypertension, AD, retinopathy, and renal-related endpoints (**Extended Data Figure 4**). Although we provide detailed sensitivity analyses for the MR signals, these results should be interpreted cautiously and should not be taken as definitive evidence of causality.

Together, these results show that the 22 mortality EPOCH clocks are heritable, polygenic, and genetically linked across organs and to diverse diseases. We next compared EPOCH with first-generation BAGs in comparable survival analyses to test our central hypothesis that event-proximity training improves prediction of future disease onset.

### Mortality EPOCH clocks show stronger prediction power of future disease onset than matched BAGs, but the two frameworks model different facets of human biology

To enable a fair comparison despite differences in model training targets, we performed a cross-domain disease onset prediction analysis, contrasting BAGs, trained on chronological age, with EPOCH clocks, trained on proximity to mortality. Crucially, the mortality-trained EPOCH is deliberately used to predict disease rather than mortality itself, so as to avoid obvious circularity^38^ (**Method 4a**). The main comparative effect was defined as the joint log-HR difference between mortality EPOCH and BAG, with values greater than zero indicating stronger mortality EPOCH effects and values less than zero indicating stronger BAG effects.

Our comparative survival results strongly favored mortality EPOCH (**Fig. 3A**). Across the DEs, most associations showed positive joint log-HR differences, indicating that mortality EPOCH retained stronger conditional disease prediction effects than matched BAGs when both were evaluated jointly. This pattern was especially prominent for proteomics and metabolomics EPOCH clocks, where large positive differences were observed across cardiovascular, respiratory, renal, endocrine/metabolic, liver, immune, and other systemic DEs. Only a small subset of DEs fell below zero, indicating stronger BAG effects (**Supplementary Figure 27**). **Supplementary Figure 28** confirms the EPOCH dominance using data only from the held-out test dataset to further rule out data leakage from the training data. Our results, therefore, support the central premise that EPOCH clocks capture future disease-relevant biology beyond conventional age-acceleration measures, although the two sets of clocks capture different facets of human biology by design.

**Figure 3:**
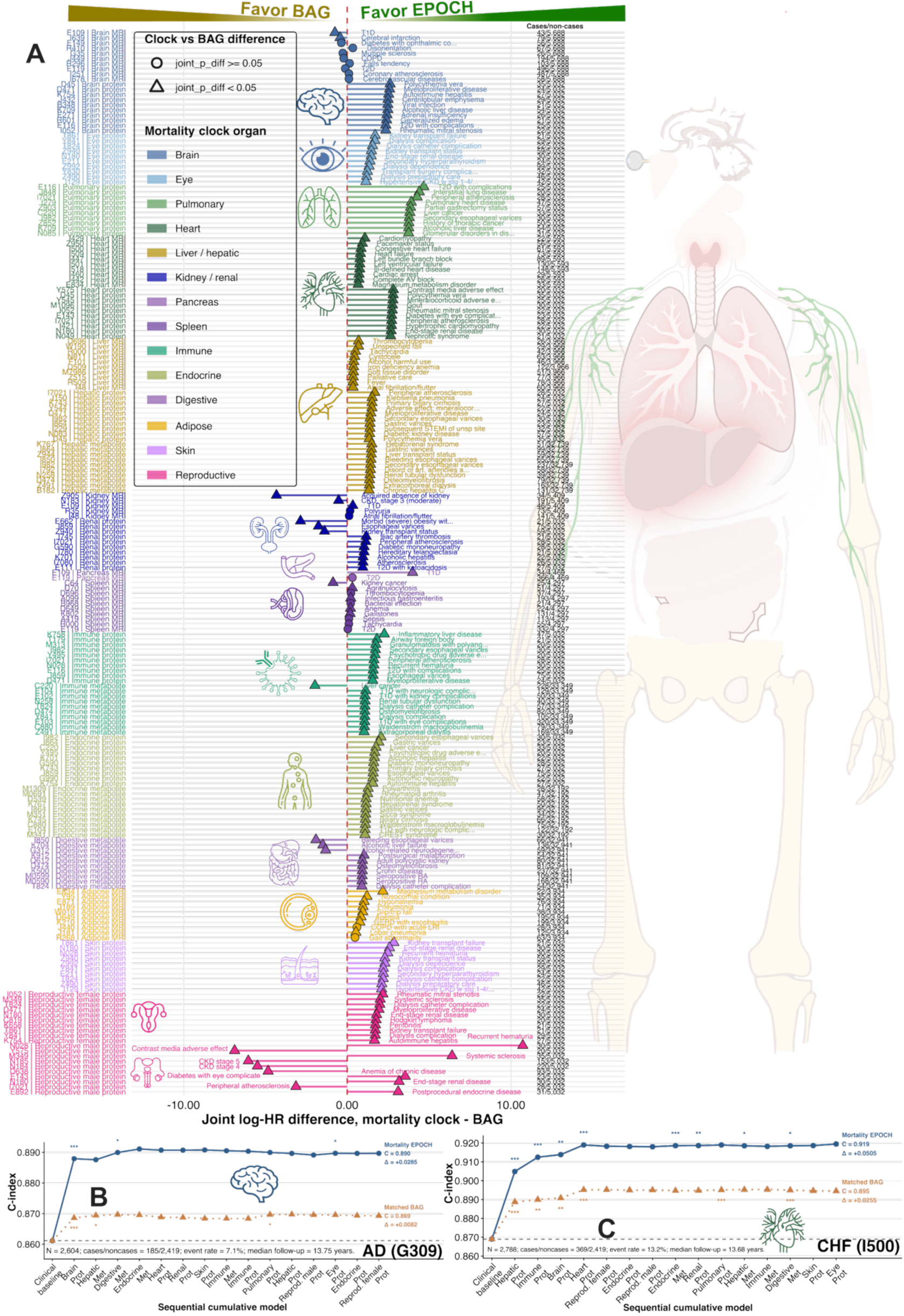
The 22 mortality EPOCH clocks predict future disease onset significantly better than respective BAGs. **A**) For each matched mortality EPOCH-BAG pair, future disease onset associations were first selected using a fair comparison rule requiring either the EPOCH or matched BAG association to pass Bonferroni correction across DEs and clocks/BAGs (P < 0.05/[number of DEs × number of clocks/BAGs]). The plot shows the top 10 disease endpoints per mortality EPOCH–BAG pair, ranked by the absolute joint log-HR difference between EPOCH and BAG effects from the joint Cox model. The x-axis shows the joint log-HR difference, *β_EPOCH_* − *β_BAG_*; values greater than 0 indicate stronger conditional disease association for the mortality EPOCH clock, whereas values less than 0 indicate stronger association for the matched BAG. Points and connecting segments are colored by mortality EPOCH organ/system. Circles indicate no nominally significant EPOCH-BAG difference, while triangles indicate nominally significant differences (two-sided P<0.05). The y-axis lists ICD-10 codes and mortality EPOCH clock; shortened disease names are annotated next to each point, and the far-right column reports the number of cases/non-cases for each disease endpoint. **B**) Cumulative survival analysis for AD (G309). Harrell’s C-index is shown as the 14 proteomics and metabolomics mortality EPOCH clocks are sequentially added to a clinical baseline Cox model. The baseline model includes age, sex, smoking status, BMI, diastolic blood pressure, and systolic blood pressure. The dashed curves denote results for the corresponding organ-specific BAGs, ordered according to their own significance-based ranking. Step-wise significance was denoted with asterisks based on sequential likelihood-ratio tests comparing each cumulative model with the immediately preceding model: P<0.05 (*), P<0.01 (**), P<0.001 (***). **C**) Cumulative survival analysis for congestive heart failure (CHF; I500).

We further showed that multi-organ mortality EPOCH clocks derived from proteomics and metabolomics (with sufficient samples after merging the organ populations) provided additive prediction power in cumulative survival analyses (**Method 4b**). **Fig. 3B-C** showcase two DEs in which mortality EPOCH clocks produced large predictive gains over the clinical baseline and outperformed the corresponding BAG results. In the example of AD, the mortality EPOCH clocks increased the 5-fold cross-validated C-index from the clinical baseline to 0.890 (ΔC = +0.0285), whereas the matched BAGs reached 0.869 (ΔC = +0.0082). **Extended Data Figure 5** and **Supplementary Note 3** present the brain-body risk architecture of the mortality EPOCH clocks. **Supplementary Figure 29** showcases a comparative analysis for predicting Alzheimer’s onset between the brain proteomic mortality EPOCH and its underlying 53 brain-enriched proteins. In addition, we compared the brain proteomic mortality EPOCH with 10 conventional mortality biomarkers (**Supplementary Figure 30**). Finally, we also compared the brain MRI EPOCH with 9 AI-derived neuroimaging subtypes^34^ in both survival time-to-event and logistic regression analyses (**Supplementary Figure 31**). The consistent predictive advantage of EPOCH across both analytical frameworks suggests that its superior prognostic performance is not attributable solely to concordance between EPOCH’s survival-based training objective and downstream survival analyses.

Conversely, EPOCH may complement rather than replace BAG, as the two frameworks are optimized for distinct modeling targets, rationales, and contexts. In **Supplementary Note 4** and **Extended Data Figure 6**, we illustrate this complementarity by jointly modeling BAG and EPOCH to identify discordant aging states and investigate candidate resilience to aging-related vulnerability (**Method 4c**). Individuals with high brain proteomics-based BAG but unexpectedly low EPOCH emerge as candidate resilient agers, characterized by distinct genetic signatures, preserved brain structure, and enrichment of lipid-and membrane-transport pathways, including HDL remodeling and ATP-binding cassette (ABC) transporter activity.

Together, these results show that the 22 mortality EPOCH clocks capture a whole-body risk architecture and outperform first-generation biological aging clocks. We next evaluated their clinical validity using longitudinal follow-up data in the 4 metabolomics clocks with sufficient sample sizes at the follow-up visit.

### Mortality EPOCH clocks capture longitudinal changes and cross-organ interactions

We further evaluated longitudinal changes in mortality EPOCH clocks using the 4 metabolomics-based models, which had sufficient sample sizes at follow-up assessment approximately 3-6 years after baseline assessment (**Method 4d**).

We first observed that mortality EPOCH was higher at the follow-up assessment than at baseline across all 4 metabolomics clocks. The largest longitudinal increase was observed for the digestive mortality EPOCH clock (Δ=3.14 years) (**Fig. 4A**). Second, longitudinal changes in all the 4 mortality EPOCH clocks provided incremental predictive value beyond baseline clocks for subsequent all-cause mortality in a landmark survival analysis. The endocrine metabolomics clock’s longitudinal change showed the strongest association with mortality risk (HR=1.07 [1.05-1.09]; P-value=3.70×10^-15^) (**Fig. 4B** & **Supplementary Table 7**). Finally, we found that longitudinal EPOCH changes also predicted subsequent disease onset. The endocrine clock predicted future COPD (HR=1.32 [1.19-1.47]; P-value=5.80×10^-7^) and MI (HR=1.29 [1.17-1.43]; P-value=1.18×10^-6^) (**Fig. 4C** & **Supplementary Table 8**).

**Figure 4:**
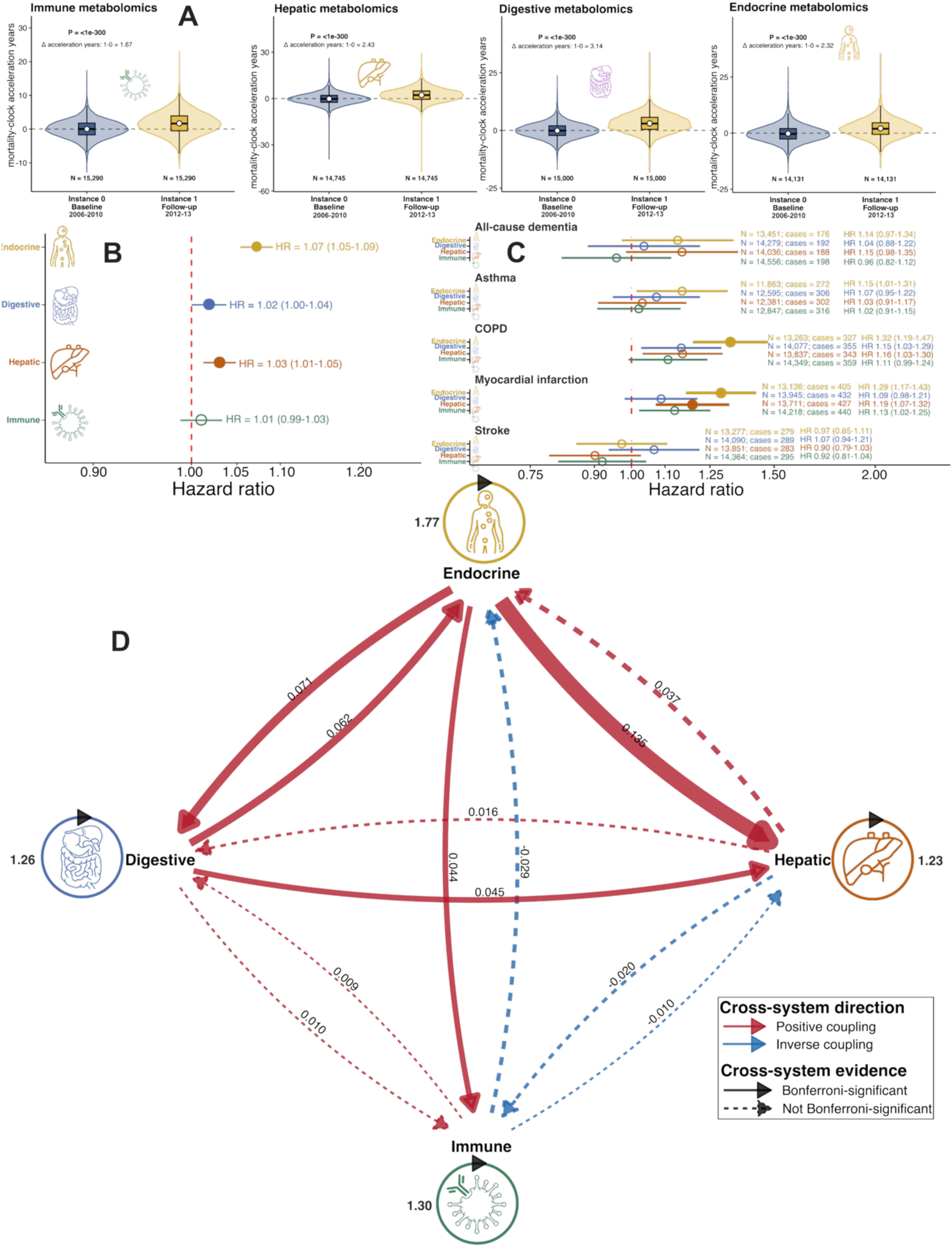
Longitudinal validity of the 4 metabolomics mortality EPOCH clocks. **A**) Violin and embedded box plots compare metabolomics mortality EPOCH acceleration years between UK Biobank instance 0 baseline assessment (2006-2010) and instance 1 follow-up assessment (2012-2013) among participants with paired measurements for each metabolomic system. Compared to proteomics and MRI data, four metabolomics mortality EPOCHs with sufficiently large sample sizes at instance 1 are shown. Sample sizes are shown below each distribution. The annotated Δ value represents the mean within-participant change from instance 0 to instance 1, and the *P* value is from a linear mixed-effects model testing the visit effect with participant-level random intercepts. **B**) Forest plot showing the hazard ratio (HR) for all-cause mortality associated with a 1-year increase in longitudinal change in metabolomics mortality EPOCH acceleration years, defined as instance 1 follow-up minus instance 0 baseline. Landmark Cox models started at the instance 1 metabolomics assessment and tested whether Δ acceleration years provided incremental mortality-prediction value beyond baseline EPOCH acceleration years and baseline chronological age. Solid points indicate significant HRs (two-sided P<0.05/4), and horizontal lines indicate 95% confidence intervals (CI) for the four metabolomics mortality EPOCHs. **C**) Forest plot showing associations between longitudinal change in metabolomics mortality EPOCH acceleration years and incident disease onset across 5 algorithmically-defined DEs: all-cause dementia, asthma, COPD, myocardial infarction (MI), and stroke. Δ acceleration years were defined as follow-up instance 1 minus baseline instance 0 for endocrine, digestive, hepatic, and immune metabolomics mortality EPOCHs. Cox models tested the association between each Δ metabolomics EPOCH and future disease onset after instance 1, adjusting for age at instance 1, sex, smoking, BMI, blood pressure, and baseline mortality EPOCH acceleration. Points indicate HRs per 1-SD higher Δ metabolomics EPOCH, and horizontal lines indicate 95% confidence intervals. Larger filled points and thicker confidence intervals denote Bonferroni-significant associations, using P < 0.05/4/5 = 0.0025 for 4 metabolomics EPOCHs and 5 disease endpoints. Sample size, number of incident cases, and HR with 95% CI are annotated for each association. **D**) Directed cross-system arrows represent associations between each baseline metabolomics mortality EPOCH and subsequent annualized change in another system, with all four baseline EPOCHs entered simultaneously and adjusted for age, sex, BMI, diastolic blood pressure, and systolic blood pressure. Arrow width is proportional to the absolute *β* coefficient, red arrows indicate positive coupling, and blue arrows indicate inverse coupling. Solid arrows denote associations that remained significant after Bonferroni correction, whereas dashed arrows denote associations that did not meet the Bonferroni threshold. Closed self-loops represent within-system longitudinal persistence, quantified by the association between each baseline EPOCH and its corresponding follow-up EPOCH (not comparable to cross-organ *β* values). Numerical labels indicate *β* estimates. Overall, the network highlights an endocrine-centered temporal architecture, including strong endocrine-to-hepatic coupling and reciprocal coupling between endocrine and digestive mortality EPOCH clocks.

Finally, we investigated whether mortality-proximity propagates across organ systems using data from both instances using a temporal coupling analysis (**Method 4e**). Baseline endocrine EPOCH showed the strongest prospective association with subsequent hepatic EPOCH progression and also predicted changes in the digestive and immune systems, suggesting that endocrine-related metabolic dysregulation may indicate broader multisystem deterioration (**Fig. 4D**). The strong within-system results additionally indicate substantial longitudinal persistence of each EPOCH (**Supplementary Table 9**). We also performed an exploratory analysis of associations between longitudinal EPOCH changes and medication history (**Supplementary Figure 32**).

To summarize, the longitudinal analyses show that mortality EPOCH clocks dynamically track changes in event-proximal biology over time. We next tested the broad applicability and generalizability of EPOCH by extending it to disease-specific clocks and mortality clocks to diverse populations and mortalities.

### Applicability and generalizability of the EPOCH framework

We next evaluated both the applicability and generalizability of EPOCH across different disease domains, including all-cause dementia, asthma, COPD, stroke, and MI.

We found that disease-specific EPOCH clocks in UKBB improved test-set discrimination beyond baseline covariate-baseline models across multiple diseases, with the most consistent gains arising from proteomics and metabolomics models (**Fig. 5A**). Across the 47 significant and clinically interpretable disease-specific EPOCH clocks, incremental test-set ΔC-index gains were observed across all diseases, with representative maxima including dementia brain proteomics (ΔC=0.0366), COPD heart MRI (ΔC=0.0783), asthma reproductive female proteomics (ΔC=0.0277), and stroke endocrine proteomics (ΔC=0.0237). Some rare-event or imaging-based disease clocks should be interpreted more cautiously because of year-scale calibration (e.g., COPD clocks with a hollow triangle) (**Supplementary Figures 33-37**).

**Figure 5:**
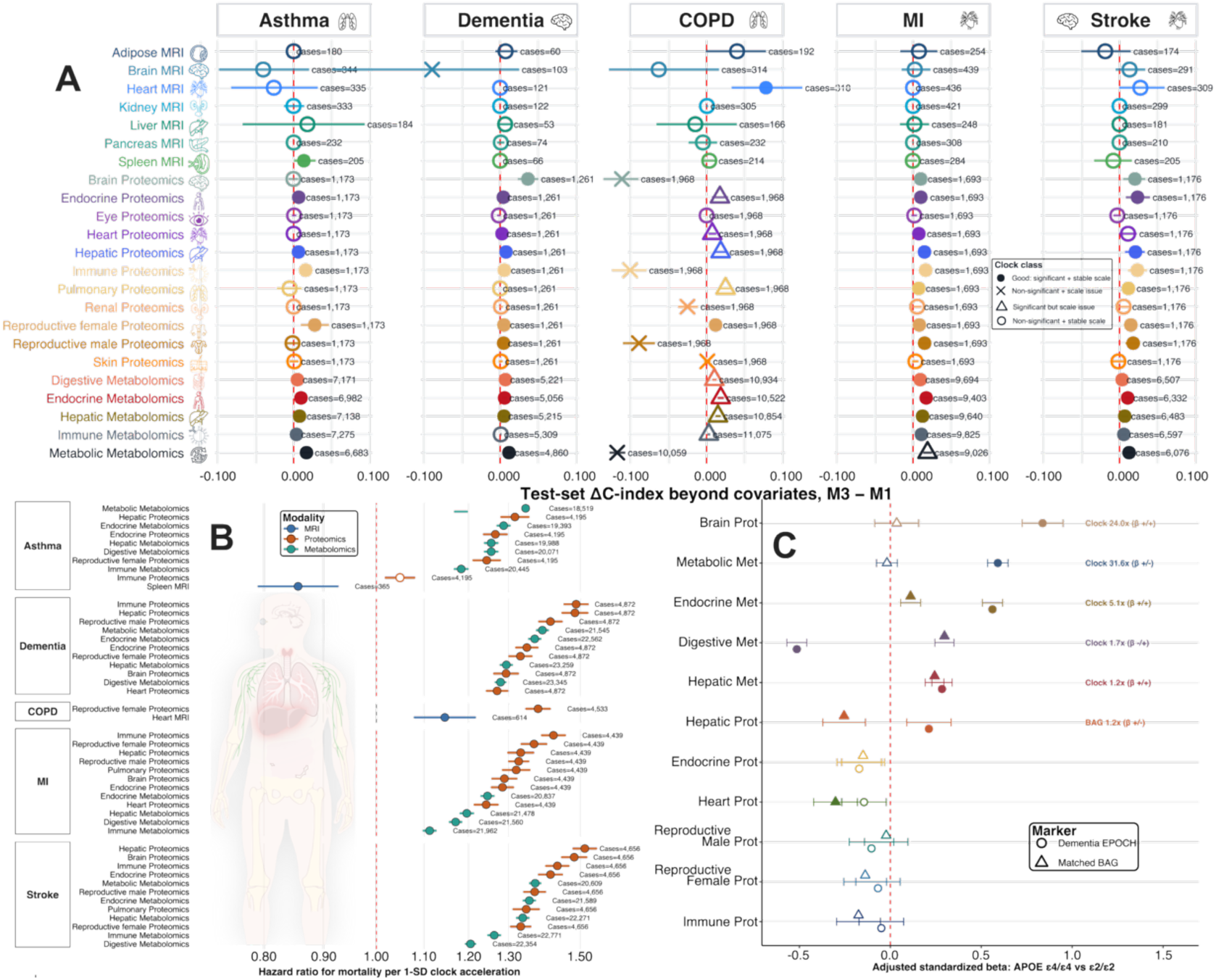
Applicability and generalizability of EPOCH clocks. **A**) Forest plot compares the incremental predictive value of 23 disease EPOCH clocks for future onset of asthma, dementia, COPD, myocardial infarction (MI), and stroke. The x-axis shows the test-set ΔC-index (M3 model with the EPOCH clock minus M1 model with covariates only). Horizontal bars indicate 95% confidence intervals (CI) for ΔC-index. Colors denote organ-specific EPOCH clocks, and right-side annotations show the number of cases for each disease clock. We selected the 47 filled circles indicating clocks with significant positive incremental value and stable year-scale calibration as significant disease clocks, which were used for downstream analyses. Open circles indicate non-significant clocks with stable scaling; triangles indicate significant clocks with year-scale issues; and crosses indicate non-significant clocks with year-scale issues. The red dashed vertical line marks ΔC-index=0, where the EPOCH clock provides no incremental discrimination beyond covariates. **B**) Forest plot showing associations between disease EPOCH clocks and subsequent all-cause mortality across the 5 disease clocks. Each point represents the hazard ratio for mortality per 1-SD higher disease EPOCH, with horizontal bars indicating 95% CI. Colors denote clock modality, including MRI, proteomics, and metabolomics. Filled circles indicate Bonferroni-significant mortality associations using two-sided P < 0.05/47/5 = 2.13 × 10^-4^. Right-side annotations show the number of mortality cases/deaths included for each clock. **C**) Forest plot comparing *APOE* ε4/ε4 versus ε2/ε2 effects on 11 dementia EPOCH clocks and their matched BAGs. The x-axis shows adjusted standardized beta estimates; positive values indicate higher clock/BAG burden among *APOE* ε4/ε4 carriers relative to *APOE* ε2/ε2 carriers, whereas negative values indicate lower burden. Circles represent dementia EPOCH clocks and triangles represent matched BAGs; horizontal bars show 95% CI. Filled symbols indicate Bonferroni-significant associations using P < 0.05/11/2 = 2.27×10^-3^, while open symbols indicate non-significant associations. Fold labels are shown for EPOCH-BAG pairs where at least one marker is significant and indicate whether the EPOCH clock or matched BAG has the larger absolute *APOE* effect size, with *β* signs denoting the direction of EPOCH/BAG effects.

We next evaluated whether the 47 disease clocks predicted future all-cause mortality. The strongest mortality associations were observed for stroke and dementia EPOCH clocks. For example, the stroke hepatic proteomics clock showed the largest mortality association (HR=1.51 [1.48-1.55]; P-value=8.31×10^-280^). Disease clocks for MI, COPD, and asthma also predicted mortality, including MI immune proteomics (HR=1.42 [1.39-1.45]; P-value=1.32×10^-196^) and asthma metabolomics-based metabolic (HR=1.35 [1.33-1.36]; P-value<1×10^-300^). At the disease level, median mortality HRs per 1-SD EPOCH acceleration were highest for stroke clocks (median HR=1.36), followed by dementia (median HR=1.35), MI (median HR=1.29), COPD (median HR=1.26), and asthma (median HR=1.26). Proteomics clocks showed the strongest and most consistent mortality associations overall, with a median HR=1.33 and the largest incremental C-index gains (**Fig. 5B**). **Supplementary Figure 38** compares the disease-specific EPOCH clocks with ten conventional biomarkers for predicting all-cause mortality.

We then tested whether the 11 dementia EPOCH clocks were linked to APOE carrier status by comparing homozygous *APOE* ε4/ε4 carriers with *APOE* ε2/ε2 carriers. Six of the 11 dementia EPOCH clocks were Bonferroni-significant (P<0.05/11/2=2.27×10^-3^), including brain proteomics (*β*=0.84±0.06; P-value=4.64×10^-45^), metabolomics-based metabolic (*β*=0.59±0.03; P-value=1.32×10^-91^), endocrine metabolomics (*β*=0.56±0.03; P-value=1.91×10^-86^), digestive metabolomics (*β*=-0.51±0.03; P-value=1.64×10^-74^), hepatic metabolomics (*β*=0.28±0.03; P-value=1.72×10^-23^), and hepatic proteomics (*β*=0.21±0.06; P-value=6.81×10^-4^) (**Fig. 5C**). Compared with matched BAGs, dementia EPOCH had the larger absolute effect in 7 of 11 matched pairs. We also performed genetic analyses on the 47 disease EPOCH clocks (**Extended Data Figure 7-8**, **Supplementary Note 5**).

Finally, in UKBB, we conducted an exploratory mediation analysis (**Method 5**) to evaluate whether associations between molecular disease EPOCH clocks (at baseline) and observed mortality (after imaging visit) were statistically mediated through the seven MRI mortality EPOCH clocks (at second imaging visit) (**Extended Data Figure 9**). The results identified both disease-concordant and cross-organ mediation patterns, highlighting a distributed multi-organ architecture linking molecular disease susceptibility to subsequent mortality and providing a framework for investigating brain-body interactions in human aging and disease. We present the detailed findings with an expanded interpretation provided in **Supplementary Note 6**.

## Discussion

EPOCH is an organ-resolved biological clock framework that enriches first-generation biological aging clocks by shifting from chronological age prediction to direct modeling of future risk of adverse clinical events. Specifically, EPOCH is trained on prospective time-to-event clinical outcomes to quantify individualized risk before clinical manifestation. To summarize, EPOCH captures event-proximity biology beyond conventional biological aging clocks, provides a whole-body framework for brain-body mechanistic and biological discovery, and can be applied to and generalized across diverse populations and settings.

### EPOCH directly models proximity to future clinical events rather than chronological age

EPOCH complements biological aging research by shifting the modeling target from chronological age to future clinical event proximity. This event-centered design has direct clinical implications. By learning from pre-event biology, EPOCH provides individualized estimates of future mortality and disease risk before overt clinical onset, supporting earlier detection, longitudinal monitoring^39^, and prevention. Importantly, EPOCH outperformed first-generation biological aging clocks in predicting future disease endpoints, affirming that survival-based event-proximity training captures disease-relevant biology that is not fully represented by generic age acceleration. Previous work also supported this claim. For example, a recent study by Mavrommatis et al.^40^ benchmarked 14 epigenetic clocks and concluded that second-and third-generation clocks significantly outperform first-generation clocks in disease predictions. Goeminne and colleagues^41^ also confirmed that their mortality clocks, a very similar survival-based clock construction to EPOCH, not only better predict mortality hazard but also the hazards of various age-related diseases.

The modeling distinction of BAG compared to EPOCH is critical because, by design, biological aging clocks provide a single cross-sectional snapshot intended to summarize global variation in aging, even though aging is inherently multidimensional, heterogeneous, and connected to numerous aspects of human health and lifespan^24,42^. For example, an individual may not appear globally “older” by a conventional BAG model, yet may already carry organ-or modality-specific biological signatures of approaching dementia, myocardial infarction, stroke, COPD, or asthma^13^. Moreover, EPOCH can be recalculated at repeated visits, while incorporating longitudinal outcome information by design. It enables dynamic tracking of preclinical progression, risk stabilization, and potential intervention response over time^43^.

### EPOCH complements rather than replaces biological aging clocks

Relatedly, EPOCH, or any other next-generation clocks, is not intended to supersede the first-generation BAG approaches, which remain highly valuable for characterizing broad inter-individual variation in aging biology and for studying mechanisms associated with accelerated or resilient aging^10^. The two frameworks address related but distinct biological questions because they are optimized toward different targets. BAGs quantify the extent to which an individual’s biological profile resembles that of chronologically older or younger individuals, thereby capturing generalized and widespread age-related variation. In contrast, EPOCH is explicitly trained on the timing of adverse clinical events and is therefore designed to preferentially capture biological variation that is directly proximal to future disease onset or mortality. Accordingly, the stronger prognostic performance of EPOCH (**Fig. 3**) should not be interpreted as evidence that biological ageing clocks are universally inferior, but as a partial consequence of aligning the learning target more directly with the clinical outcome of interest. Indeed, concordance and discordance between BAG and EPOCH may themselves be biologically informative and clinically valuable, distinguishing generalized biological aging from event-proximal vulnerability and potentially revealing states of resilience^44^ in which age-related biology is not accompanied by elevated future clinical risk (**Extended Data Figure 6**). To summarize, rather than replacing the BAG framework, EPOCH expands the biological clock family along a complementary event-proximity axis, bridging fundamental aging biology with precision risk stratification, prevention, and ultimately the extension of healthy lifespan.

### EPOCH establishes a whole-body framework for human aging and longevity

Another contribution of EPOCH is its organ-resolved, cross-organ modeling framework. Rather than compressing aging into a single global score, EPOCH decomposes future clinical risk across organs and omics modalities, enabling localization of risk to specific biological systems and molecular layers. This is particularly important for complex diseases, where the relevant biology may not reside solely in the primary clinically affected organ^45,46^. For example, dementia and stroke risk may be reflected not only in brain-derived clocks, but also in immune, hepatic, endocrine, cardiometabolic and pulmonary clocks (**Extended Data Figure 7G**). The mortality EPOCH clocks further reinforced a whole-body risk architecture, in which organ-specific clocks were interconnected through shared phenotypic and genetic associations with systemic DEs (**Fig. 2D-E**). These findings reinforce a brain-body model of human aging and disease: future clinical events emerge from distributed, genetically influenced vulnerabilities and susceptibility across organ systems rather than from isolated organ-and tissue-specific dysfunction. Consistent with their organism-wide regulatory roles, the endocrine and immune systems particularly showed some of the strongest signals (**Fig. 2E** and **Fig. 3A**). This pattern is biologically plausible because both systems coordinate communication across multiple organs, shape systemic homeostasis, and influence diverse age-related disease processes.

Another underexplored question in systems neuroscience is how aging-related changes in one organ system are coordinated with, or potentially influence, aging processes in others. Tian et al.^24^ previously used first-generation BAGs to demonstrate the brain-body connection using data from two time points. Here, we extended this cross-organ connection to metabolomics-derived mortality clocks. The endocrine system occupies a central position within the cross-organ network, exhibiting multiple and comparatively strong connections with the hepatic, digestive, and immune systems (**Fig. 4D**). These findings may further strengthen that endocrine metabolic aging may represent an important organizing axis of brain-body and multi-organ mortality risk, while also highlighting that cross-system relationships are heterogeneous, directional, and not uniform^47^, reinforcing our recent notion that “no organ is an island^48^”.

## Limitations

Several limitations warrant consideration. First, although the findings were supported by multiple independent cohorts, validation in an additional population-scale resource with similarly deep multi-organ and multi-omics phenotyping as in UKBB will be important to establish broader generalizability. Second, the genetic analyses were conducted predominantly in individuals of European ancestry, underscoring the need for replication and extension across more diverse ancestral populations. Third, longer-term longitudinal follow-up with repeated multi-organ and multi-omics measurements will be required to evaluate the temporal synchronization of organ-specific trajectories and to fully test the proposed medical digital twin framework^25^. Fourth, despite adjustment for a broad range of covariates, residual confounding and reverse causality cannot be completely excluded. Finally, future studies should extend the EPOCH framework across a wider portion of the lifespan to characterize how event proximity and related risk trajectories evolve from early adulthood through late life.

## Methods

### Method 1: The MULTI Consortium

The MULTI Consortium is an ongoing initiative to integrate and consolidate multi-organ data and multi-omics data. Building on existing consortia and studies, MULTI aims to curate and harmonize the data to model human aging and disease at scale across organs, omics, and lifespan. Refer to **Supplementary Table 3** for comprehensive information, including the complete list of data analyzed and their respective sample characteristics. Participants provided written informed consent to the corresponding studies. The MULTI study is approved by the Institutional Review Board at Columbia University (AAAV6751).

### UK Biobank

UKBB^30^ is a population-based research initiative comprising around 500,000 individuals from the United Kingdom between 2006 and 2010. Ethical approval for the UKBB study has been secured, and information about the ethics committee can be found here: https://www.ukbiobank.ac.uk/learn-more-about-uk-biobank/governance/ethics-advisory-committee. The core biomedical data used to develop EPOCH include *in vivo* imaging, plasma proteomic and metabolomic profiles, and adverse clinical variables, including longitudinal information on disease diagnoses and mortality events. UKBB was used as the primary dataset for developing the mortality and disease-specific EPOCH clocks.

### FinnGen

The FinnGen^49^ study is a large-scale genomics initiative that has analyzed over 500,000 Finnish biobank samples and correlated genetic variation with health data to understand disease mechanisms and predispositions. The project is a collaboration between research organizations and biobanks within Finland and international industry partners. For the benefit of research, FinnGen generously made its GWAS findings accessible to the wider scientific community (https://www.finngen.fi/en/access_results). This research utilized the publicly released GWAS summary statistics (version R9), which became available on May 11, 2022, after harmonization by the consortium. No individual data were used in the current study.

FinnGen published the R9 version of GWAS summary statistics via REGENIE software (v2.2.4)^50^, covering 2272 DEs, including 2269 binary traits and 3 quantitative traits. The GWAS model encompassed covariates like age, sex, the initial 10 genetic principal components, and the genotyping batch. Genotype imputation was referenced to the population-specific SISu v4.0 panel. We included GWAS summary statistics for 521 FinnGen DEs in our analyses.

### Psychiatric Genomics Consortium

PGC^51^ is an international collaboration of researchers studying the genetic basis of psychiatric disorders. PGC aims to identify and understand the genetic factors contributing to various psychiatric disorders such as schizophrenia, bipolar disorder, major depressive disorder, and others. The GWAS summary statistics were acquired from the PGC website (https://pgc.unc.edu/for-researchers/download-results/), underwent quality checks, and were harmonized to ensure seamless integration into our analysis. No individual data were used from PGC. Each study detailed its specific GWAS models and methodologies, and the consortium consolidated the release of GWAS summary statistics derived from individual studies. In the current study, we included summary data for 6 brain diseases.

### Method 2: The EPOCH framework based on organ-resolved survival models Principle

EPOCH was designed as an event-proximal or time-to-event clock framework for estimating how close an individual’s organ-specific biological profile is to a future clinical event (i.e., disease onset or mortality). In contrast to the first-generation BAG framework, which is trained to predict chronological age and then interpreted *post hoc* in relation to disease or mortality (as demonstrated in prior studies^1–6^), EPOCH directly learns the relationship between organ-specific features and time-to-event outcomes. Thus, the estimand is not biological age per se, but relative prospective proximity to an adverse endpoint, including all-cause mortality or incident disease onset (**Fig. 1A**). Of note, EPOCH builds on regularized Cox survival modeling, as used in several existing second-generation clocks (**Supplementary Table 2**), while extending this approach into an organ-resolved framework across different disease domains and mortality. For all-cause mortality, the event is death after the sample date of imaging, proteomics, or metabolomics. For disease-specific clocks, the event is the first incident diagnosis of the target disease after baseline, with participants who already have the target disease at or before baseline excluded.

### Modeling approaches

For the participant, time zero was defined as the date at which the organ-specific features were measured. In UKBB, for imaging-derived EPOCH clocks, time zero corresponded to the imaging assessment date; for plasma omics clocks, time zero corresponded to the baseline omics assessment date. The event date was defined as the date of death for mortality clocks or the first diagnosis date of the target disease for disease-specific clocks. We excluded participants with missing time zero, events occurring before or on time zero, or follow-up shorter than the minimum allowed duration. Participants were administratively censored at the pre-specified censoring date if the event did not occur before censoring.

For each organ system and omics modality, an organ-specific feature matrix was constructed using only features belonging to that organ system and modality (i.e., the same features used in our BAG work). Three omics layers were included, including *i*) organ-specific imaging features, *ii*) organ-enriched proteins, and *iii*) organ-associated metabolites. Non-organ covariates were included to adjust for potential confounding. The baseline covariate set included age and sex, and additional covariates included BMI, systolic and diastolic blood pressure, smoking status, assessment center, intracranial volume, or other modality-specific technical covariates when applicable.

EPOCH was trained using a Cox proportional hazards model with elastic-net regularization^52,53^. Model parameters were estimated by minimizing the negative Cox partial log-likelihood with an elastic-net penalty on organ-specific features, where a hyperparameter (*λ*) controls overall regularization strength and another (*ρ*) controls the balance between L1 and L2 penalties. The L1 component encourages sparse organ-specific feature selection, while the L2 component stabilizes estimates in the presence of correlated biomarker features. To ensure the clock retains organ-specific features under regularization, we ensure a minimum number of nonzero organ-feature coefficients.

#### Outcome

The primary Cox linear predictor was the raw EPOCH risk score (*η_i_*). To derive covariate-adjusted EPOCH acceleration (at the year scale), we derived covariate-adjusted EPOCH deceleration/acceleration by residualizing *η_i_* against non-organ covariates in the training set, then scaling the residuals to z-score (for downstream analyses) and year units. Let *Z_i_* denote the covariates used for residualization. A linear model was fit in the training set:

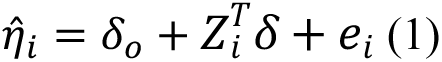

The residual *e_i_* represents the event-proximal organ signal after removing the expected risk-score component attributable to non-organ covariates. EPOCH (de)acceleration was then defined as:

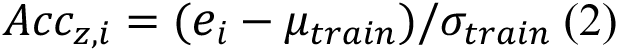

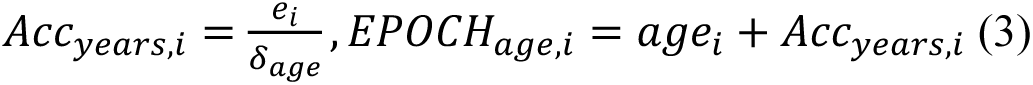

where *μ_train_* and *σ_train_* are the mean and standard deviation of training-set residuals, and *δ_a_*_g*e*_ is the fitted age coefficient from the residualization model. If *δ_a_*_g*e*_ was near zero, the year-scale acceleration exploded (as shown in the metabolomics-based metabolic mortality EPOCH clock in **Supplementary Figure 1**). Of note, the risk score can still be predictive in such a case, and the standardized acceleration z-score can still be usable, but the conversion into “years” is not reliable because the year scaling calibration is unstable (far beyond 100 years old).

#### Model evaluation

To determine whether organ-specific EPOCH features provided predictive power beyond demographic and clinical covariates, we evaluated 4 comparable Cox survival models using the same training, validation, and held-out test splits: *i*) the first model, *M0*, included age and sex only and served as a minimal demographic benchmark; *ii*) the second model, *M1*, included all retained non-organ covariates and served as the primary covariate baseline; *iii*) the third model, *M2*, included only organ-specific features, thereby quantifying the standalone event-proximal signal carried by the target organ or modality; and *iv*) the fourth model, *M3*, was the full EPOCH model and included both non-organ covariates and organ-specific features. We define the incremental model prediction power as:

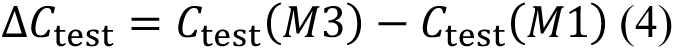

A positive Δ*C*_)*+)_indicates that organ-specific features improve discrimination beyond the full-covariate baseline. Uncertainty in Δ*C*_)*+)_was estimated using paired bootstrap resampling of the held-out test set, preserving the paired predictions from M3 and M1 for each participant.

#### Data split

For each EPOCH model, the dataset was split into training, validation, and hold-out test (20%) sets. The remaining 80% was further split into training and validation sets for a 5-fold cross-validation to select elastic-net hyperparameters, following our previous open-access, reproducible frameworks^54–56^. Additionally, splitting was stratified by event status to preserve event balance across splits. Candidate models were trained on the training split and evaluated on the validation split using Harrell’s concordance index^57^ (C-index). The model with the highest validation C-index was selected. After hyperparameter selection, the final model was refit using the combined training and validation data and was evaluated once on the held-out test set. In the held-out test set, predictive performance was characterized beyond Harrell’s C-index using bootstrap 95% confidence intervals, calibration slope, observed-to-expected risk, IPCW Brier scores, and calibration plots comparing predicted with Kaplan-Meier-observed risk across risk strata.

### Method 3: Genetic analyses

We used the genotype and imputed genotype data from UKBB for all genetic analyses. Our quality-check pipeline focused on European ancestry in the UKBB (6,477,810 SNPs passed quality checks). We summarize our genetic quality-check steps. First, we excluded related individuals (up to 2^nd^-degree) from the complete UKBB sample using the KING software for family relationship inference^58^. We then removed duplicated variants from all 22 autosomal chromosomes. Individuals whose genetically identified sex did not match their self-acknowledged sex were removed. Other exclusion criteria were: *i*) individuals with more than 3% of missing genotypes; *ii*) variants with minor allele frequency (MAF; dosage mode) of less than 1%; *iii*) variants with larger than 3% missing genotyping rate; *iv*) variants that failed the Hardy-Weinberg test at 1.0×10^-10^. To further adjust for population stratification^59^, we derived the first 40 genetic principal components using the FlashPCA software^60^. Details of the genetic quality check protocol are described elsewhere^1–6,25,34–37,45,48,61–63^.

#### **(a)** GWAS

We applied a linear mixed model regression to the European ancestry populations using fastGWA^64^ implemented in GCTA^32^. Our GWASs adjusted for common covariates, including age, dataset status (training/validation/test datasets), age-squared, sex, interactions of age with sex, BMI, waist circumference, standing height, weight, and the first 40 genetic principal components, as well as organ-specific covariates. We applied a stringent threshold based on Bonferroni correction at the genome-wide significance level (5×10^-8^/number of clocks) to annotate the significant independent genomic loci. For all GWASs, genomic loci were annotated using FUMA^65^, as detailed in **Supplementary Note 1**.

#### **(b)** Three key genetic parameters

We first calculated the SNP-based heritability (*h*^2^*_SNP_*) using GCTA^32^, which employs raw individual genotype data to generate a genetic relationship matrix, addressing the “missing heritability” issue^32^. GCTA-GREML^32^ was the default implementation, and GCTA-HEreg was used to overcome scalability issues in metabolomics clocks, which have a larger sample size (*N*>270,000). Additionally, two additional methods using only GWAS summary statistics were also used as secondary confirmation. First, we estimated *h^2^* using the LDSC^31^ software, leveraging only GWAS summary statistics. To this end, we used precomputed LD scores from European ancestry in the 1000 Genomes dataset. Second, we also used SBayesS^33,66^ to estimate *h*^2^*_SNP_.* Notably, disparities in SNP-based *h^2^* estimates were observed between the software tools, which is in line with our own findings^2,67^ and those from other studies^68,69^. Besides SNP-based heritability, SBayesS^33^ also estimates two additional key genetic parameters: *i*) polygenicity (*Pi*), and the relationship between minor allele frequency (MAF) and effect size (*S*) as *ii*) natural selection signatures. We used the software’s pre-computed sparse LD correlation matrix derived from European ancestry by Zeng et al.^33^.

#### **(c)** Genetic correlation

We estimated the genetic correlation (*g_c_*) using LDSC^31^ between these EPOCH clocks and *i*) 46 previously developed AI biomarkers (including first-generation BAGs^2–6^ and disease subtypes^34–37^ from our previous work detailed in **Supplementary Note 2**) and *ii*) 527 disease endpoints from the FinnGen and PGC datasets (**Supplementary Table 10**). We employed precomputed LD scores from the 1000 Genomes of European ancestry, maintaining default settings for other parameters in LDSC. It’s worth noting that LDSC corrects for sample overlap, ensuring an unbiased genetic correlation estimate^70^.

#### (d) Two-sample bidirectional Mendelian randomization

We performed two-sample bidirectional Mendelian randomization by linking the EPOCH clocks and 525 DEs from FinnGen^49^ and PGC^51^ (2 DEs from PGC did not provide allele frequency information). In total, 2 networks were established: i) *EPOCH2DE* and ii) *DE2EPOCH*. We performed systematic quality-checking procedures to ensure unbiased exposure/outcome variable and instrumental variable (IV) selection.

We used the *TwoSampleMR* package^71^ to infer the causal relationships. We employed five distinct Mendelian randomization methods, including the inverse variance weighted (IVW) method, Egger, weighted median, simple mode, and weighted mode estimators. The STROBE-MR Statement^72^ guided our analyses to increase transparency and reproducibility, encompassing the selection of exposure and outcome variables, reporting statistics, and implementing sensitivity checks to identify potential violations of underlying assumptions. First, we performed an unbiased quality check on the GWAS summary statistics. Notably, the absence of population overlapping bias^73^ was confirmed, given that FinnGen and UKBB participants largely represent populations of European ancestry without explicit overlap with UKBB. PGC GWAS summary data were ensured to exclude UKBB participants. Furthermore, all consortia’s GWAS summary statistics were based on or lifted to GRCh37. Subsequently, we selected the effective exposure variables by assessing the statistical power of the exposure GWAS summary statistics in terms of instrumental variables (IVs), ensuring that the number of IVs exceeded 7 before harmonizing the data. Crucially, the function “*clump_data*” was applied to the exposure GWAS data, considering LD. The function “*harmonise_data*” was then used to harmonize the GWAS summary statistics of the exposure and outcome variables. Bonferroni correction was applied across all tested traits based on the number of effective DEs, resulting in a relatively conservative adjustment.

Finally, we conducted multiple sensitivity analyses. First, we conducted a heterogeneity test to scrutinize potential violations of the IV’s assumptions. To assess horizontal pleiotropy, which indicates the IV’s exclusivity assumption^74^, we utilized a funnel plot, single-SNP Mendelian randomization methods, and the Egger estimator. Furthermore, we performed a leave-one-out analysis, systematically excluding one instrument (SNP/IV) at a time, to gauge the sensitivity of the results to individual SNPs.

### Method 4: Survival analyses using baseline EPOCH clocks

After deriving the EPOCH clocks, we evaluated their predictive value through different sets of cross-domain analyses to avoid circularity^38^: *i*) mortality EPOCH clocks for predicting incident DEs, and *ii*) disease-specific EPOCH for estimating future risk of all-cause mortality.

#### (a) Mortality EPOCH for disease prediction and disease EPOCH for mortality risk

For the 22 mortality EPOCH clocks at baseline, we employed a Cox proportional hazards model while adjusting for covariates to test the associations of the 22 mortality EPOCH clocks with the time to incident ICD-based single DEs. Notably, we excluded individuals with any prior disease diagnosis (except the disease of interest) to ensure the analysis was restricted to a disease-free population. To train the model, the “time” variable was determined by calculating the difference between the date of diagnosis of the disease for cases (or the censoring date for non-cases) and the date of attending the assessment center. Participants who were diagnosed with a specific disease of interest after enrolling in the study were classified as cases; non-cases were defined as participants without any disease diagnoses.

To evaluate whether mortality EPOCH clocks capture future disease risk beyond first-generation BAGs, we compared 22 mortality EPOCH clocks with their corresponding organ-specific BAGs across incident ICD-based DEs. Each comparison was performed within a matched mortality EPOCH-BAG pair. DEs were analyzed only when the number of incident cases and non-cases exceeded the prespecified minimum threshold (*N*>20). Within each DE tested, both the mortality EPOCH and BAG were standardized to z-scores, allowing hazard ratios to be interpreted per 1-SD higher score. Different models were evaluated for fair comparisons: *i*) the covariate-only model, *ii*) the EPOCH-only and *iii*) the BAG-only models quantified the separate associations of each score with incident disease, while *iv*) the joint model tested whether the mortality EPOCH and BAG contributed independent information when modeled together. The primary pairwise comparison was based on the difference between the *iv*) joint-model log-hazard coefficients, where joint log-HR Δ*β* > 0 indicates a stronger conditional association for the mortality EPOCH clock than for the matched BAG, and Δ*β* < 0 indicates a stronger conditional association for the BAG. A Wald test was used to test the null hypothesis, accounting for the covariance between the two coefficient estimates in the joint Cox model.

#### **(b)** Cumulative survival analyses for predicting future disease onset

We also evaluated the cumulative predictive value of 15 mortality EPOCH clocks, comprising 11 proteomics clocks and 4 metabolomics clocks, which overlap with each other for a sufficient number of participants but not with MRI data (i.e., too small sample sizes after merging the populations), for predicting ICD-defined DEs.

Cox proportional hazards models were fitted using a complete-case analysis sample containing nonmissing values for all 15 mortality EPOCH clocks (dropping MRI clocks due to high missing rates) and all included covariates. The clinical covariate model included baseline age, sex, smoking status, body mass index, diastolic blood pressure, and systolic blood pressure. All mortality EPOCH clocks were standardized to a mean of zero and standard deviation of one, such that hazard ratios corresponded to a 1-SD increase in each clock. A covariate-only Cox model was fitted first, after which mortality EPOCH clocks were added sequentially to construct cumulative models from one to all 15 clocks.

For the cumulative analysis’s clock order, individual clocks surviving the Bonferroni-corrected threshold of (P<0.05/15) were entered first and ranked by decreasing hazard ratio; clocks not meeting this threshold were appended in a random order. At each step, we reported the hazard ratio and 95% confidence interval for the newly added clock, the model log-likelihood, partial Akaike information criterion, apparent Harrell concordance index, change in concordance relative to the covariate-only model, and change relative to the preceding cumulative model. Predictive discrimination was evaluated using stratified 5-fold cross-validation to mitigate overfitting issues as more clocks/features were added. To provide a matched benchmark against first-generation BAGs, the cumulative analysis was repeated using the 15 organ-and modality-matched BAGs. By default, BAGs were ranked using their own disease-specific individual BAG hazard ratios and P-values, thereby allowing the BAG models an independently optimized entry order.

#### (c) Joint analysis using BAG and EPOCH

To distinguish generalized biological aging from event-proximal vulnerability, we quantified discordance between brain proteomic BAG and brain proteomic mortality EPOCH. We first standardized both and then regressed EPOCH on BAG using ordinary least squares, and the participant-level residual was retained as the continuous EPOCH-BAG discordance phenotype. Negative residuals indicate lower mortality proximity than expected for an individual’s degree of biological aging, whereas positive residuals indicate higher-than-expected mortality proximity.

Categorical phenotypes were defined from joint tails of BAG and the standardized residual. Candidate resilient agers (CRA) had high BAG but low EPOCH-BAG residual, whereas concordant unfavorable agers (CUA) had high BAG and high residual. Concordant favorable agers (CFA) had low BAG and low residual, and latent vulnerability agers (LVA) had low BAG and high residual. Definitions based on the 10%, 20%, and 25% tails were generated; the 25% definition was used for the downstream neuroimaging analyses to increase the available sample size. Of note, these thresholds were selected pragmatically for mechanistic characterization, while recognizing that the specific cutoffs are inherently arbitrary.

To maximize statistical power, GWAS was performed using the continuous EPOCH-BAG residual rather than the categorical CRA phenotype. Genetic variants were tested for association with the residual using fastGWA. Because the sign of a GWAS beta depends on the coded effect allele, directional biological interpretation was based on alignment with molecular-QTL effects rather than solely on labeling loci as resilience or vulnerability based on the *beta* coefficient sign. Directional eQTL analyses were performed using data from GTEx v8’s eQTL mappings.

The gene expression and gene-set enrichment analyses specifically focused on these resilience-related genes. Multiple SNP-gene mappings were first collapsed within each gene and GTEx tissue. A strict resilience-associated expression gene was defined as a gene with at least one tissue showing exclusively resilience-direction eQTL evidence (promoting lower residual) and no tissue showing an opposing or mixed direction. The resilience-associated genes were used for the cross-tissue expression heatmap and FUMA GENE2FUNC enrichment using default settings.

Brain-wide association analyses compared CRA with CUA, providing a contrast between individuals with similarly elevated brain proteomic BAG but lower versus higher event-proximal vulnerability. CRA participants were coded as 1 and CUA participants as 0, with CFA, LVA, and other participants excluded. Diffusion MRI fractional anisotropy (FA) measures, T1-weighted regional gray matter volumes, and functional MRI connectivity features were analyzed separately using logistic regression. Models were adjusted for age at imaging, sex, and standardized brain proteomic BAG. Positive coefficients indicate that higher values of an imaging feature were associated with greater odds of CRA, whereas negative coefficients indicate greater odds of CUA. Multiple testing was controlled separately within each imaging modality using the Benjamini-Hochberg false-discovery-rate (FDR) procedure, considering relatively small sample sizes between proteomics and neuroimaging populations.

#### (d) Longitudinal modeling of the metabolomics mortality EPOCH clocks

Longitudinal analyses were performed for the 4 metabolomics-based mortality EPOCH clocks with sufficient follow-up data. Follow-up metabolomics data from instance 1 (2012-2013) were restricted to the same organ-specific metabolite features used by the corresponding baseline instance 0 model (2006-2010). The pretrained baseline metabolomics mortality EPOCH models were applied to instance 1 without retraining, generating follow-up EPOCH clock estimates.

Participant-level longitudinal change was defined as the follow-up EPOCH value minus the baseline value (ΔEPOCH_1−0_). Because longitudinal change is only observable among participants who survived and returned for follow-up, all subsequent survival analyses used a landmark design^75^ starting at the instance 1 sample date. In this design, cases were defined as participants who died or had a disease diagnosis after the instance 1 sample date and before administrative censoring, whereas non-cases were participants alive or censored after instance 1.

We first compared paired distributions of mortality EPOCH clocks between baseline and follow-up for each organ system. We then investigated whether longitudinal change (ΔEPOCH_1−0_) predicted subsequent all-cause mortality or disease. Cox proportional hazards models were fit from the instance 1 landmark date to death/diagnosis or censoring. The primary model included baseline EPOCH, longitudinal change (ΔEPOCH_1−0_), and chronological age, allowing us to test whether the longitudinal change in EPOCH provided additional mortality/disease-prediction information beyond baseline EPOCH level and age.

#### (e) Cross-organ temporal coupling network in the 4 metabolomics mortality EPOCH clocks

Cross-organ longitudinal temporal coupling was evaluated among the endocrine, digestive, hepatic, and immune metabolomics-based mortality EPOCH clocks using participants with complete baseline and follow-up measurements for all 4 clocks. For each participant, annualized change is denoted as ΔEPOCH_1−0_. Cross-organ coupling was assessed using multivariable linear regression models in which ΔEPOCH_1−0_ in each EPOCH was regressed simultaneously on the standardized baseline EPOCH values of all 4 EPOCH clocks. Models were adjusted for baseline age, sex, body mass index, diastolic blood pressure, and systolic blood pressure. The coefficient for each off-diagonal predictor-outcome pair quantified whether a higher baseline EPOCH in one system was associated with faster subsequent change in another system, independent of the remaining baseline EPOCH clocks and covariates. Of note, our analyses aim to answer whether greater event proximity in one system at baseline predicts faster subsequent change in another system, conditional on the other baseline clocks and measured covariates, which does not claim causality and statistically differs from the structural equation modeling approach used in Tian et al^24^. Within-organ longitudinal persistence was evaluated in separate lagged follow-up models in which the follow-up value of each EPOCH was regressed on all 4 standardized baseline EPOCH clocks and the same covariates. For within-organ associations, we did not use the coefficients from the cross-organ change-score models (above-mentioned) as within-organ coupling, as the baseline value is mathematically embedded in the change outcome and may induce regression-to-the-mean and mathematical-coupling effects.

### Method 5: Statistical mediation analysis linking molecular disease EPOCHs, MRI mortality EPOCHs and observed mortality

We investigated whether disease-related molecular EPOCH clocks measured at baseline were statistically linked to subsequent all-cause mortality through later organ-specific MRI mortality EPOCH clocks (measured at second imaging visit). The mediation framework was designed to preserve the temporal ordering of the available measurements: baseline molecular disease EPOCH acceleration was treated as the exposure (*X*), MRI mortality EPOCH acceleration obtained at imaging visit 2 as the mediator (*M*), and all-cause mortality occurring after the imaging assessment as the outcome (*Y*). Imaging date was used as the survival-analysis landmark. Follow-up extended from this landmark until death or administrative censoring, and participants without a death event contributed their complete censored follow-up.

#### A two-stage mediational model

For each model, *X* and *M* were standardized, and the models were adjusted for age, sex, and BMI. The exposure-to-mediator *a* path was estimated using OLS with HC3 robust standard errors; the mediator-to-mortality *b* path was estimated using a Cox proportional hazards model, where *b* represents the log-hazard coefficient for a 1-SD higher MRI mortality EPOCH, conditional on the molecular disease EPOCH and covariates. The indirect association was quantified as *a*×*b* on the Cox log-hazard scale. Its uncertainty was assessed using a *delta*-method approximation and 1,000 participant-level nonparametric bootstrap replicates, in which both the OLS and Cox models were refitted. Bootstrap percentile 95% confidence intervals were reported as complementary evidence. Because Cox coefficients are non-collapsible, we did not estimate a proportion mediated and interpreted *a*×*b* as a statistical indirect association rather than a causal natural indirect effect. Multiple testing was controlled across all 315 prespecified models using Bonferroni correction (P<0.05/315=1.59×10^-4^).

## Data Availability Statement

The GWAS summary statistics for the 22 mortality EPOCH and 47 disease-specific EPOCH clocks from UKBB are publicly accessible via the MEDICINE portal (https://labs-laboratory.com/medicine/) and also available at Synapse (https://www.synapse.org/Synapse:syn76450542). GWAS summary data for the DEs were downloaded from the official websites of FinnGen (R9: https://www.finngen.fi/en/access_results) and PGC (https://pgc.unc.edu/for-researchers/download-results/). Individual data from the UK Biobank can be requested through proper registration at https://www.ukbiobank.ac.uk/.

## Acknowledgments

This research is supported through funding from the NIH-supported MULTI Consortium (W.J.; grant number: RF1AG092412) and Cure Alzheimer’s Fund (W.J.). The MULTI consortium (W.J.; UK Biobank Application Number: 647044) aims to integrate multi-organ imaging and multi-omics data to advance our understanding of human aging and disease. We want to express our sincere gratitude to the UK Biobank team for their invaluable contribution to advancing clinical research in our field (https://www.ukbiobank.ac.uk/). We want to acknowledge the participants and investigators of the FinnGen study and the PGC consortium, and we thank FinnGen (https://www.finngen.fi/en) and PGC (https://pgc.unc.edu/) for their generosity in sharing the GWAS summary statistics with the scientific community.

## Authors’ Contributions Statement

Dr. Wen has full access to all the study data and is responsible for its integrity and accuracy.

*Study concept and design*: W.J.

*Drafting of the manuscript*: W.J.

*Critical revision of the manuscript for important intellectual* content: All.

*Analysis*: W.J. ran all analyses. W.J., C. Y., and S.Z. developed the MEDICINE portal.

## Competing Interests Statement

All authors state no competing interests in this study.

## Code Availability Statement

The software and resources used in this study are all publicly available:

• Lifelines (v0.27.8): https://lifelines.readthedocs.io/en/latest/, survival analysis;

• GCTA (v1.94.1): https://yanglab.westlake.edu.cn/software/gcta/#Overview, fastGWA;

• LDSC (git version aa33296): https://github.com/bulik/ldsc, genetic correlation;

• FUMA (v1.8.0): https://fuma.ctglab.nl/, Gene mapping, genomic locus annotation;

• TwoSampleMR (v0.5.6): https://mrcieu.github.io/TwoSampleMR/index.html, Mendelian randomization;

• GCTB (v2.0.5): https://cnsgenomics.com/software/gctb/; SBayesS.

### The MULTI Consortium

Junhao Wen^1,2,3,4,15^, Andrew Zalesky^10^, Christos Davatzikos^11^, Paul Aisen^12^, Reisa Sperling^13^, Keenan A. Walker^14^, Eleanor M. Simonsick^14^, Susan M. Resnick^14^, Luigi Ferrucci^14^

^1^Laboratory of AI and Biomedical Science (LABS), Columbia University, New York, NY, USA

^2^Department of Biomedical Engineering, Columbia University, New York, NY, USA

^3^Department of Computer Science, Columbia University, New York, NY, USA

^4^Department of Radiology, Columbia University, New York, NY, USA

^10^Department of Psychiatry, Melbourne Medical School, The University of Melbourne, Melbourne, Victoria, Australia

^11^Artificial Intelligence in Biomedical Imaging Laboratory (AIBIL), Center for AI and Data Science for Integrated Diagnostics (AI^2^D), Perelman School of Medicine, University of Pennsylvania, Philadelphia, PA, USA

^12^Alzheimer’s Therapeutic Research Institute, Keck School of Medicine of the University of Southern California, San Diego, CA 92121, USA

^13^Department of Neurology, Massachusetts General Hospital, Brigham and Women’s Hospital, and Harvard Medical School, Boston, MA, USA

^14^National Institute on Aging, National Institutes of Health, Baltimore, MD, USA

^15^New York Genome Center (NYGC), New York, NY, USA

## Extended Data Figure

**Extended Data Figure 1:**
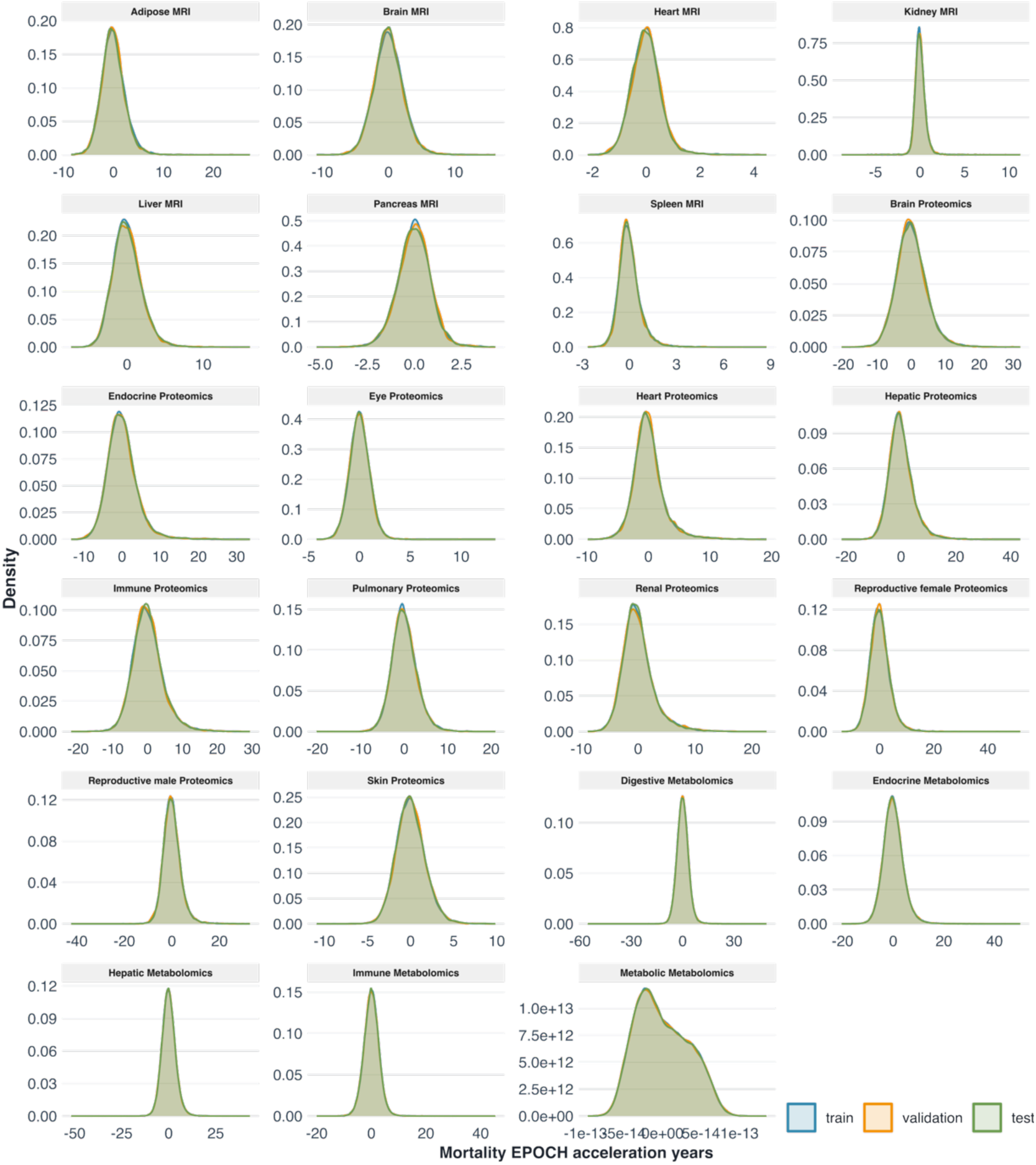
Distribution of mortality EPOCH clock acceleration across training, validation, and test datasets. Density plots show the distributions of acceleration-year estimates for 23 mortality EPOCH clocks across the training, validation, and held-out test datasets. Each panel represents one organ-specific EPOCH, spanning MRI-, proteomics-, and metabolomics-derived models. Across most clocks, the train, validation, and test distributions overlapped substantially, supporting stable out-of-sample calibration of mortality EPOCH acceleration estimates. One exception was the metabolomics EPOCH clock, whose acceleration estimates did not scale to a biologically interpretable year range and exhibited numerical instability. Therefore, the metabolomics mortality EPOCH clock was excluded from subsequent follow-up analyses.

**Extended Data Figure 2:**
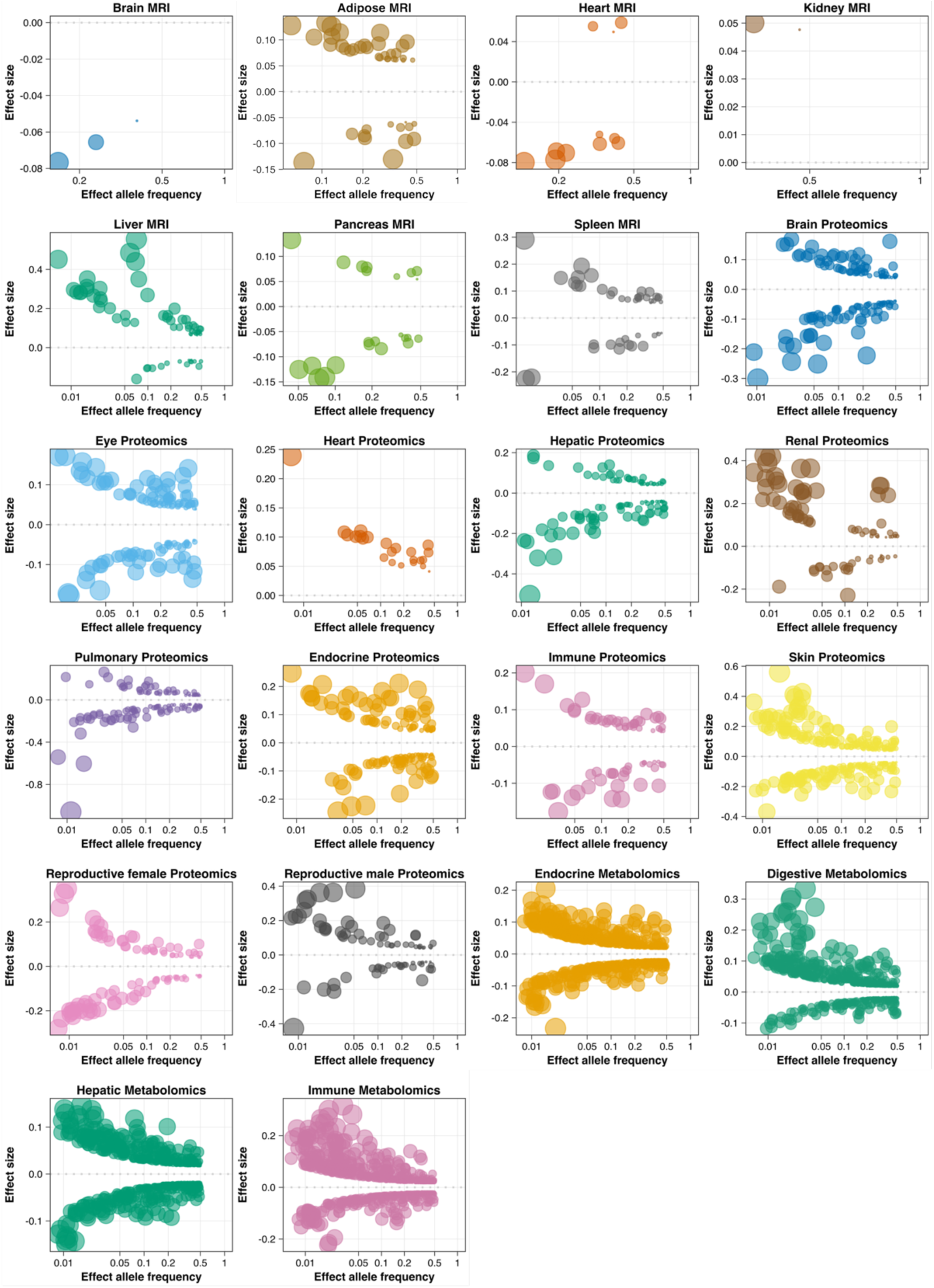
Trumpet plots of the effect allele frequency vs. the *β* coefficient of the 22 mortality EPOCH clocks. Trumpet plots show the relationship between effect allele frequency and GWAS *β* coefficient for independent significant variants associated with mortality EPOCH clock year-acceleration. Each panel represents one organ-specific mortality EPOCH clock across MRI-, proteomics-, and metabolomics-derived models. Points denote FUMA-defined independent significant SNPs merged with fastGWA effect estimates; bubble size reflects the available GWAS effect size for each variant. The dashed horizontal line marks *β* = 0. Across clocks, lower-frequency variants generally showed larger absolute effect sizes, consistent with the expected trumpet-shaped genetic architecture. The metabolomics-based metabolic mortality EPOCH clock was excluded from this figure and downstream analyses because its acceleration estimates did not scale to a biologically interpretable year range.

**Extended Data Figure 3:**
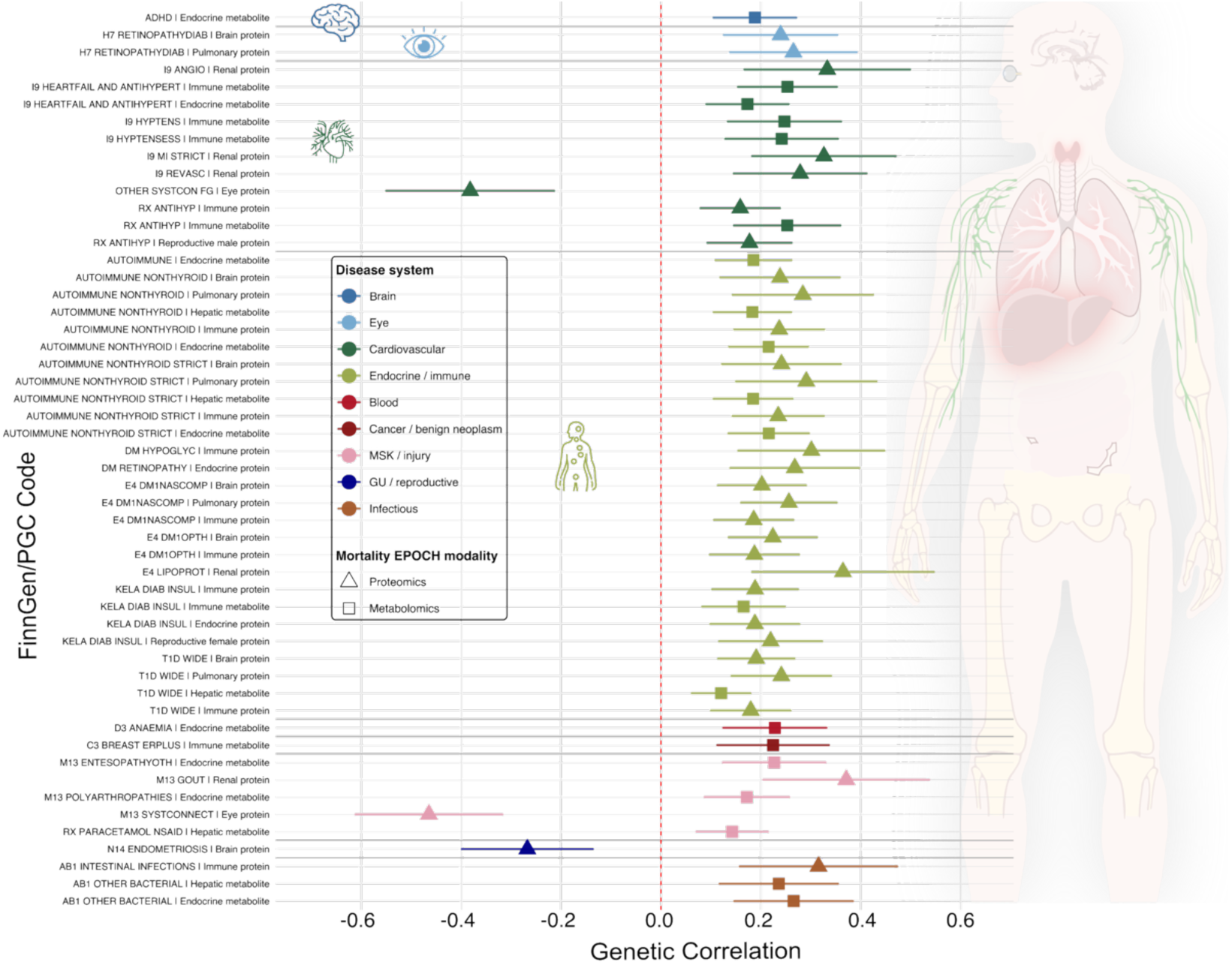
Genetic correlations link mortality EPOCH clocks to disease endpoints across the brain-body systems. Bonferroni-significant LDSC genetic correlations between the 22 mortality EPOCH clocks and disease endpoints from FinnGen and PGC GWAS summary statistics are shown. Each row represents one significant mortality EPOCH-DE pair passing the disease endpoint Bonferroni correction (two-sided P-value<0.05/527). Points indicate LDSC genetic-correlation estimates (*r_g_*), and horizontal bars represent 95% confidence intervals calculated from the standard error. Colors denote disease systems, and point shapes indicate the EPOCH clock modality, including MRI, proteomics, and metabolomics. Significant associations span neuropsychiatric, cardiovascular, endocrine/immune, blood, cancer, musculoskeletal, genitourinary/reproductive, and infectious disease domains, highlighting shared genetic architecture between mortality-proximal organ clocks and diverse disease liabilities. The anatomical silhouette provides a brain-body context for the multi-organ EPOCH framework.

**Extended Data Figure 4:**
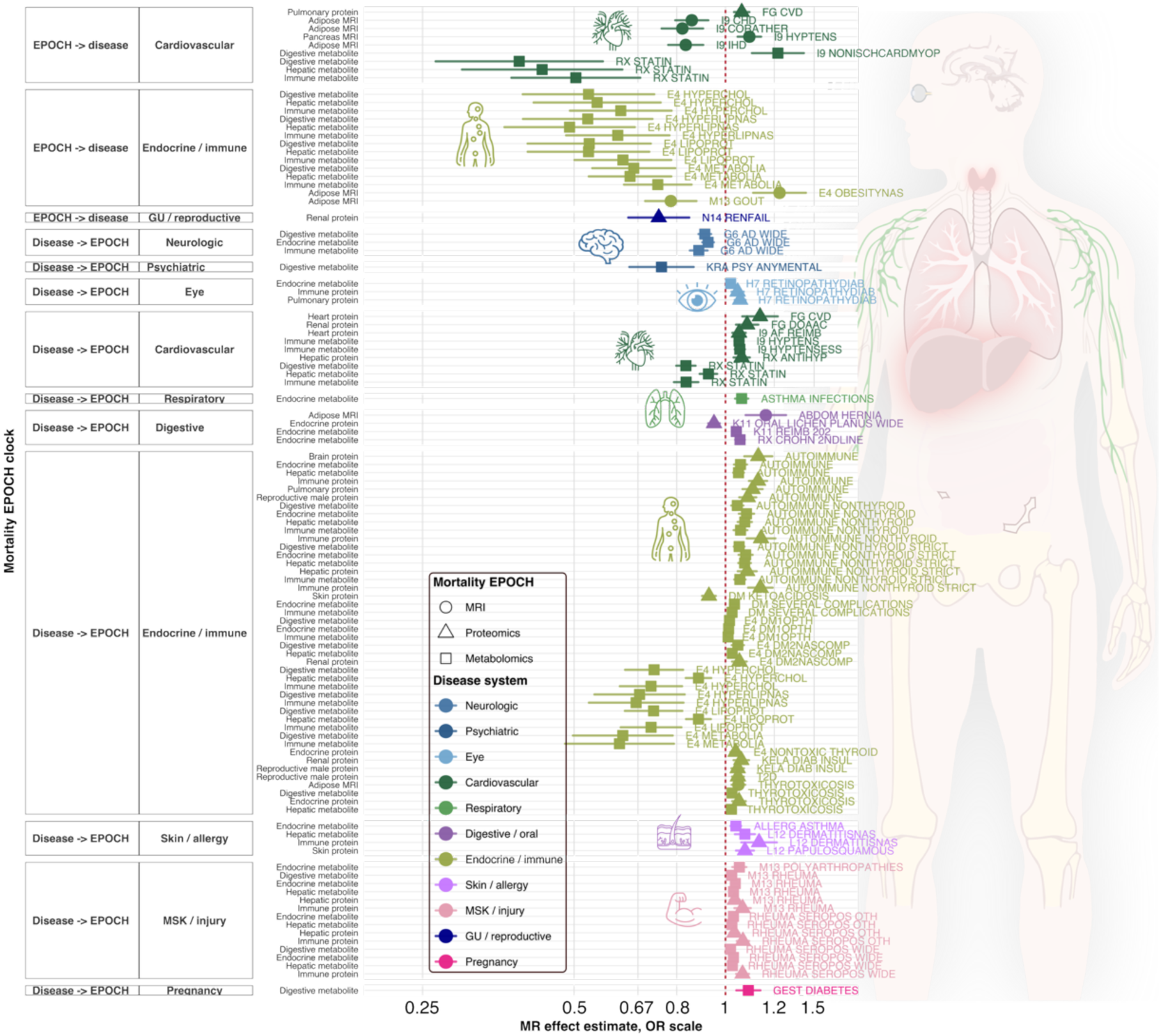
Bidirectional Mendelian randomization links mortality EPOCH clocks to disease endpoints across brain-body systems. Bidirectional Mendelian randomization associations between the 22 mortality EPOCH clocks and disease endpoints from FinnGen and PGC GWAS summary statistics are shown. Only significant IVW MR associations that passed correction for the number of disease endpoints are displayed. The upper blocks show EPOCH → DE analyses (P-value<0.05/524 DEs), testing whether genetically predicted mortality EPOCH clocks are associated with disease risk, whereas the lower blocks show DE → EPOCH analyses (P-value<0.05/213 DEs), testing whether genetically predicted disease liability is associated with mortality EPOCH clock variation. Points indicate MR effect estimates on the odds-ratio scale, and horizontal bars represent 95% confidence intervals. The red dashed vertical line marks OR = 1. Colors denote disease systems, and point shapes indicate the mortality EPOCH modality, including MRI, proteomics, and metabolomics. Disease endpoint ICD codes are annotated to the right of each association.

**Extended Data Figure 5:**
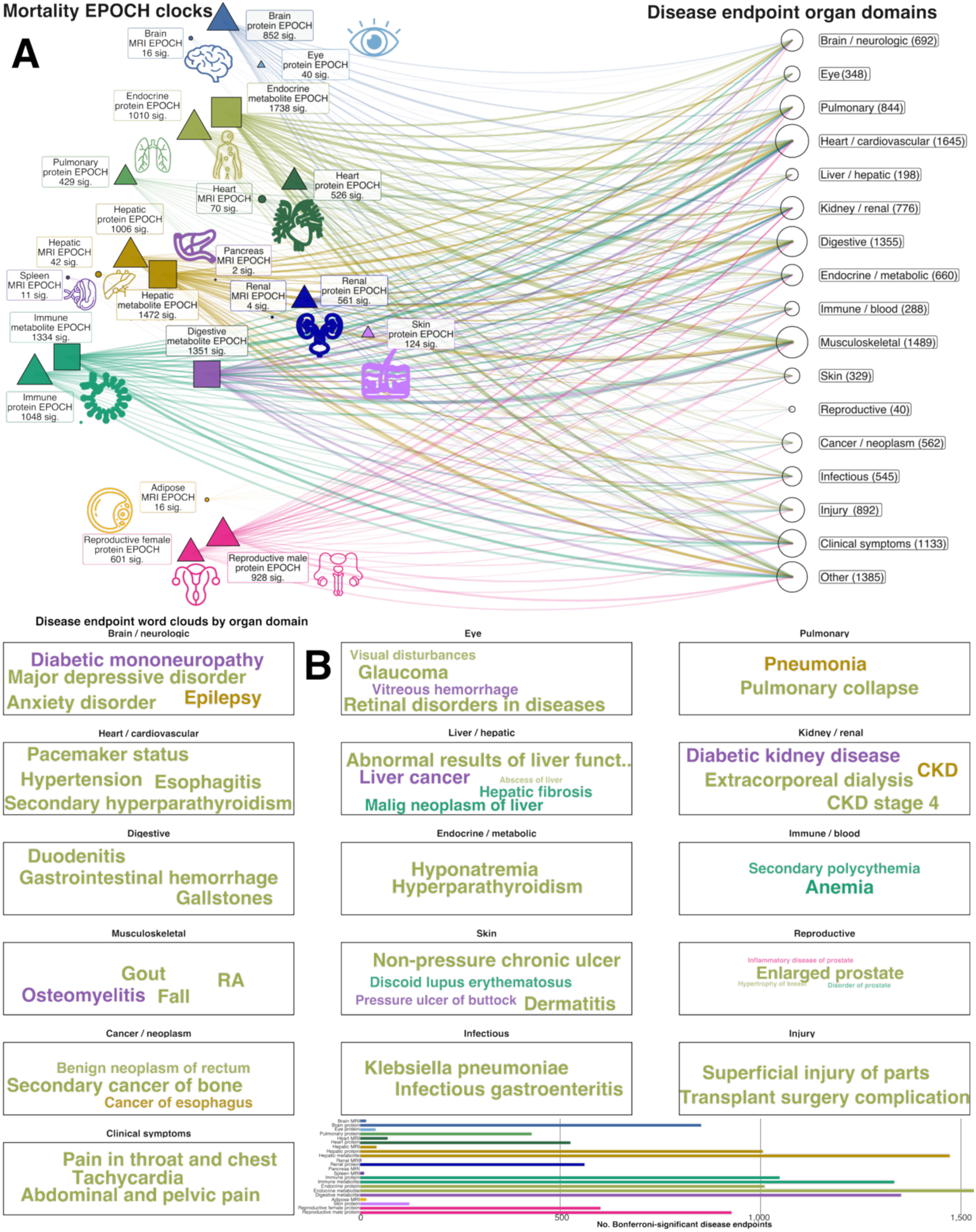
Mortality EPOCH reveals a brain-body, whole-body architecture of future disease risk. **A**) Organ-specific mortality EPOCH clocks are positioned on a simplified human anatomy layout according to their biological source, including MRI-, proteomics-, and metabolomics-derived clocks. Disease endpoints are grouped into organ-informed domains on the right. Curves connect each mortality EPOCH clock to disease endpoint domains with Bonferroni-significant survival associations (two-sided P-value < 0.05/3471/22). Node size reflects the number of significant disease endpoints, and curve density highlights the breadth of within-organ and cross-organ disease-risk connectivity. Together, this panel shows that mortality EPOCH clocks capture a distributed brain-body risk architecture rather than isolated organ-specific signals. **B**) Disease endpoint word clouds and clock-level burden across the whole body. Word clouds show disease endpoint names grouped by organ-informed disease domains. Word color indicates the dominant mortality EPOCH organ/system associated with that endpoint, using the same color scheme as Panel A. Larger words represent endpoints with stronger or more recurrent mortality EPOCH associations. The bar plot at the bottom summarizes the total number of Bonferroni-significant disease endpoints associated with each mortality EPOCH clock, highlighting which organ-and omics-derived clocks contribute most broadly to future disease risk prediction.

**Extended Data Figure 6:**
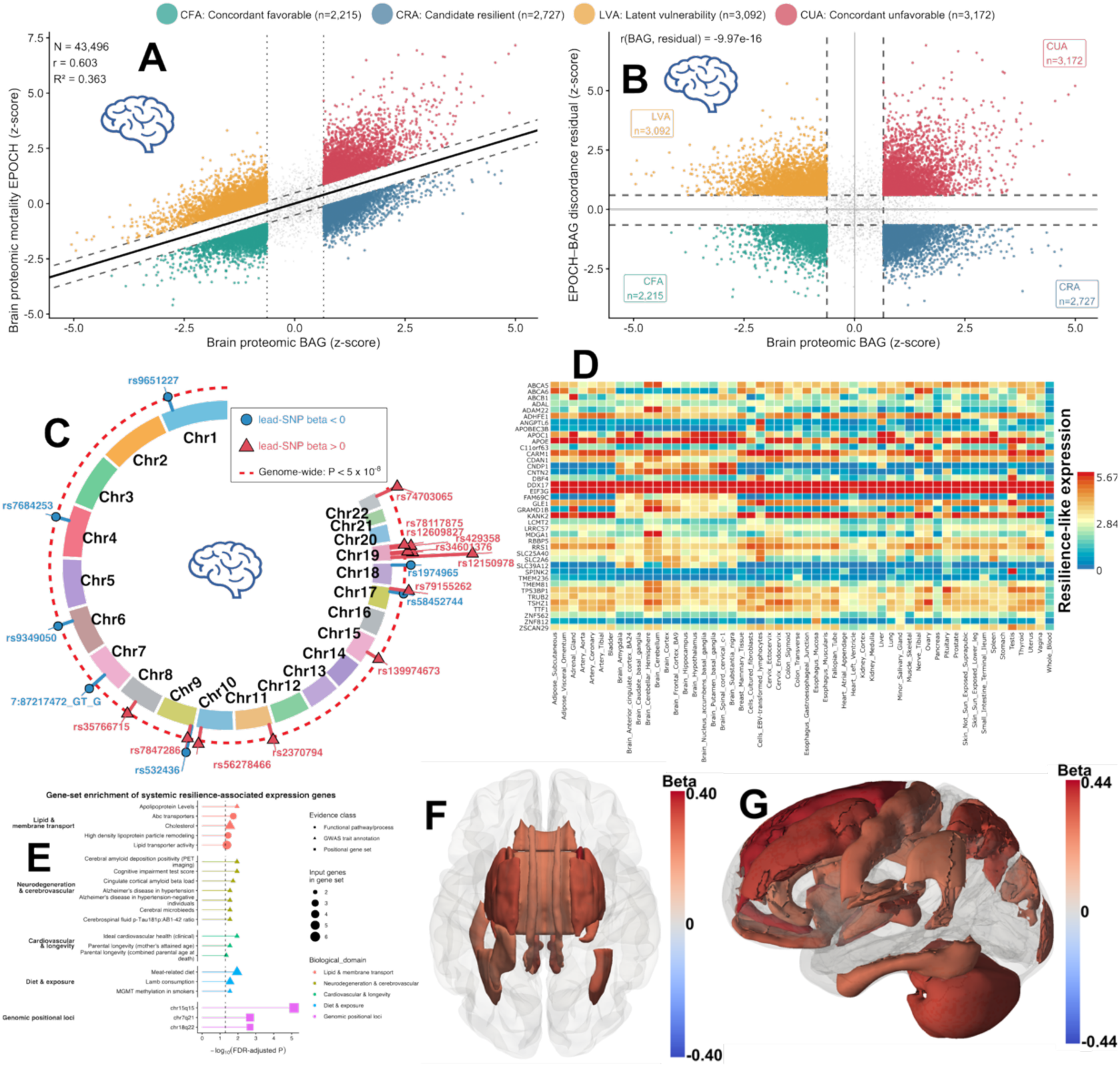
Multiscale characterization of brain proteomic EPOCH-BAG discordance reveals candidate resilience-associated genetic, biological pathways, and neuroimaging signatures. **A)** Relationship between brain proteomic BAG and EPOCH. The fitted regression captures the component of EPOCH shared with BAG, whereas vertical deviation from the regression line represents EPOCH-BAG discordance. Individuals in the distributional tails are highlighted according to the four aging phenotypes subsequently defined from BAG and EPOCH residuals. **B)** Definition of candidate resilient and comparison aging phenotypes. Brain proteomic BAG is plotted against the standardized EPOCH residual conditional on BAG. Using the 25th and 75th percentiles, participants were classified as concordant favorable agers (CFA; low BAG, low residual), candidate resilient agers (CRA; high BAG, low residual), latent vulnerability agers (LVA; low BAG, high residual), or concordant unfavorable agers (CUA; high BAG, high residual). CRA therefore identifies individuals exhibiting lower mortality-proximal EPOCH than expected for their degree of advanced biological aging, whereas CUA represents individuals with concordantly elevated biological aging and event-proximal vulnerability. **C**) Genetic architecture of EPOCH-BAG discordance. Circular Manhattan plot of the genome-wide association study of the continuous EPOCH-BAG residual (for statistical power considerations). Genome-wide significant loci are highlighted and annotated by top lead variants. The sign of the GWAS effect refers to the effect allele, with alleles associated with lower residual corresponding to the resilience-like phenotypic direction (beta<0) and alleles associated with higher residual corresponding to the vulnerability-like direction (beta>0). **D**) Cross-tissue expression profiles of resilience-associated genes. GTEx v8 tissue-expression heatmap for genes whose genetically aligned expression direction was associated with lower EPOCH-BAG residual. Rows represent resilience-associated genes and columns represent GTEx tissues, illustrating heterogeneous expression patterns spanning brain and peripheral organ systems rather than a purely brain-restricted transcriptional architecture. **E**) Functional characterization of systemic resilience-associated genes. GENE2FUNC enrichment analysis of the systemic resilience-associated gene set using protein-coding genes as the reference background. Enriched annotations are organized by biological domain, highlighting convergence on lipid and membrane transport, neurodegenerative and cerebrovascular phenotypes, cardiovascular and longevity-related biology, and related systemic processes. GWAS Catalog annotations are interpreted as prior trait-association enrichment rather than independent mechanistic pathways. **F**) White matter microstructural correlates of candidate resilient aging. Brain visualization of diffusion MRI fractional anisotropy (FA) features associated with CRA relative to CUA in logistic-regression models adjusted for age at imaging, sex, and brain proteomic BAG. Only tracts significant after within-modality Benjamini-Hochberg false-discovery-rate correction are displayed. Colors represent the signed standardized feature regression coefficient; positive values indicate that higher FA is associated with greater odds of CRA, whereas negative values indicate greater odds of CUA. **G**) Gray matter structural correlates of candidate resilient aging.

**Extended Data Figure 7:**
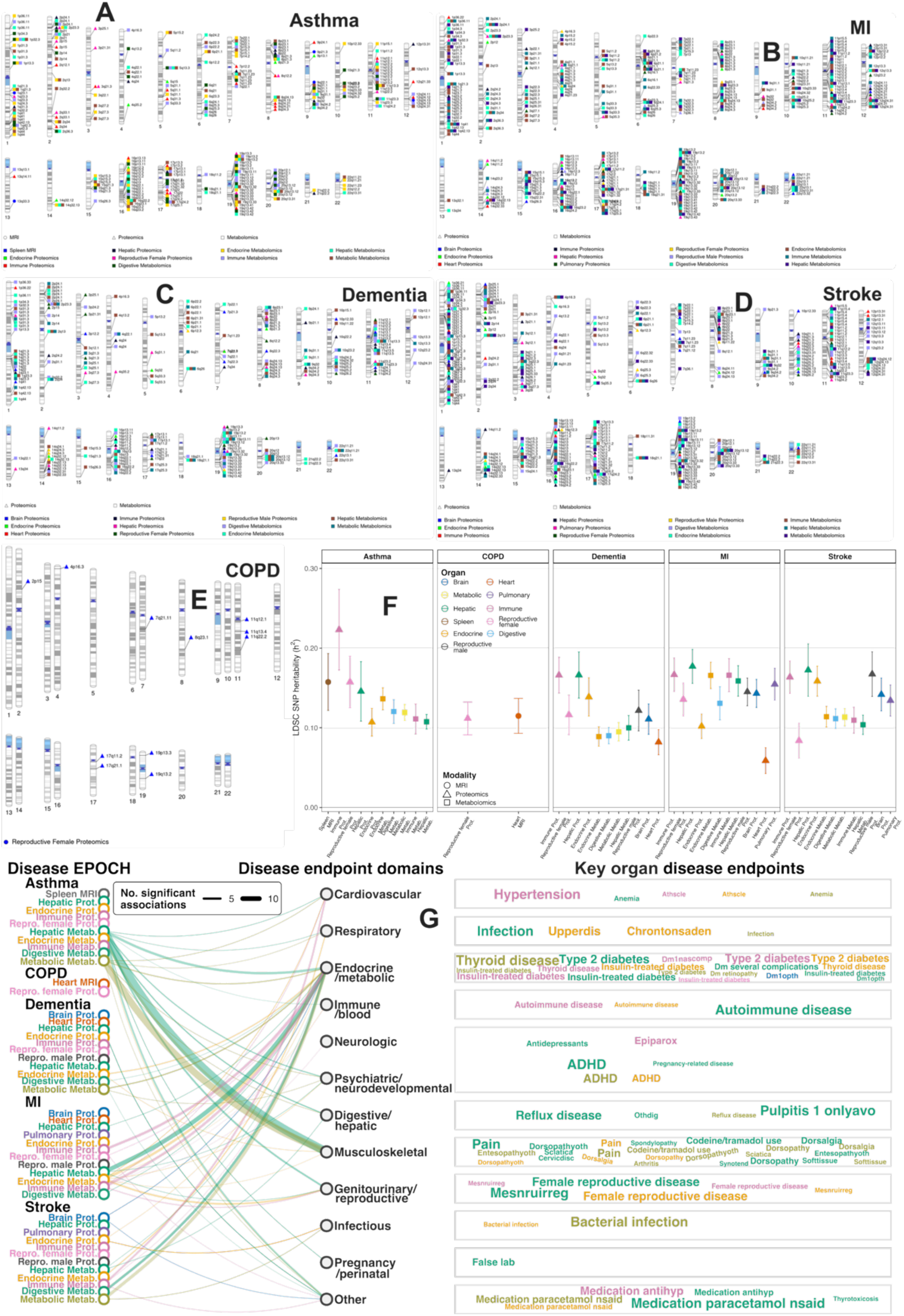
**Genetic architecture of 47 disease EPOCH clocks across brain-body systems A-E**) Chromosome ideogram plot showing genome-wide significant loci associated with organ-specific EPOCH clocks spanning asthma, MI, dementia, stroke, and COPD. Colors denote the contributing EPOCH clock, spanning MRI-, proteomics-, and metabolomics-derived organ systems. **F**) LDSC-based SNP heritability estimates are shown for the 47 disease EPOCH clocks across five disease outcomes: asthma, COPD, dementia, myocardial infarction, and stroke. Points represent SNP heritability estimates, and error bars indicate standard errors. Colors denote the organ or biological system used to derive each EPOCH clock, while point shapes indicate data modality, including MRI-, proteomics-, and metabolomics-based clocks. **G**) Network visualization summarizes genetic correlations between disease EPOCH clocks and disease endpoints in each organ/clinical domain (two-sided P-value<0.05/527/22). Clock labels on the left are colored by organ or system, and curves connect clocks to disease domains on the right. The accompanying word cloud panels highlight representative disease endpoints within each domain, with larger words indicating stronger or more frequent signals. All GWAS summary statistics are publicly disseminated at our MEDICINE portal: https://labs-laboratory.com/medicine/.

**Extended Data Figure 8:**
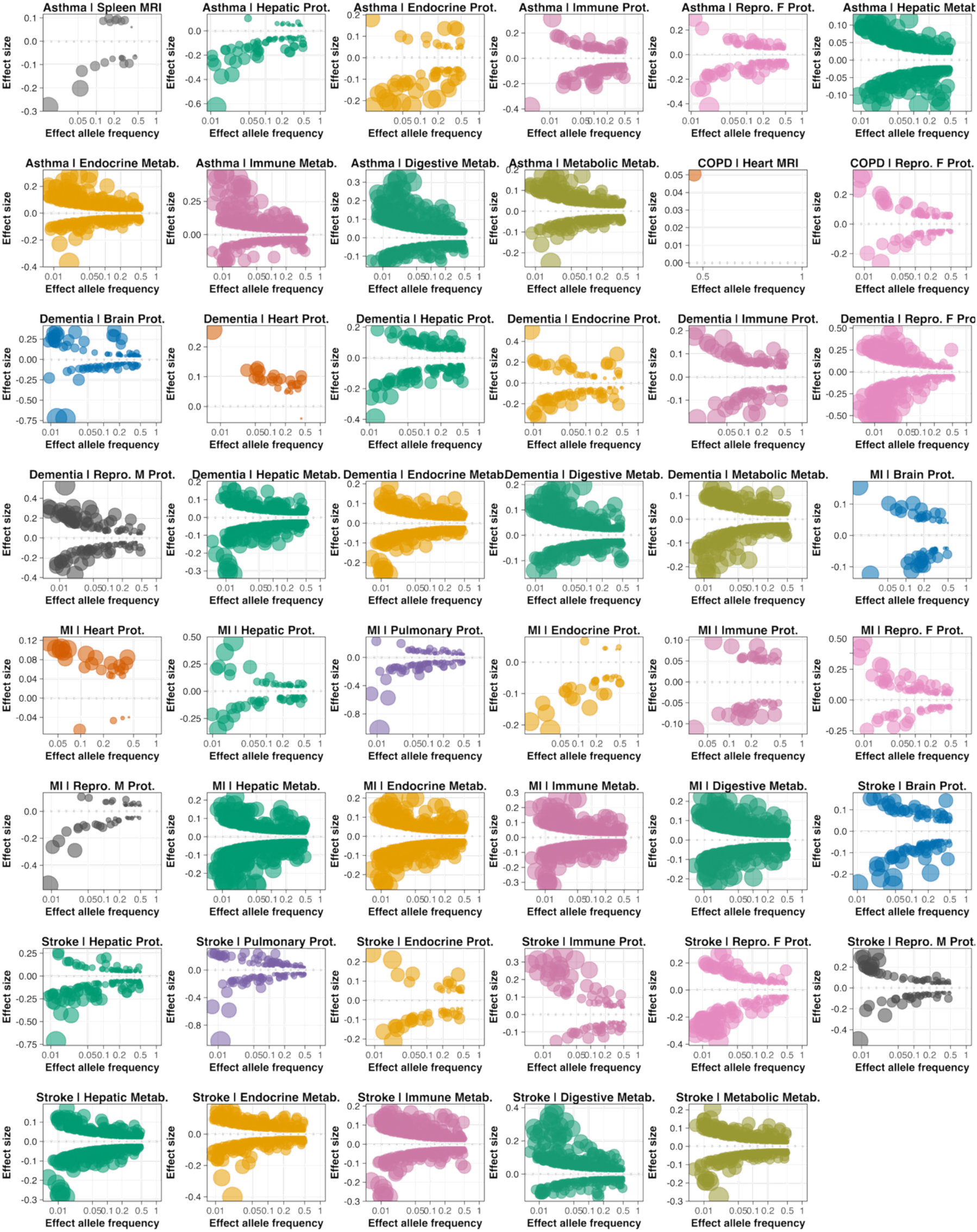
Trumpet plots of the effect allele frequency vs. the *β* coefficient of the 47 disease EPOCH clocks. Trumpet plots show the relationship between effect allele frequency and GWAS *β* coefficient for independent significant variants associated with disease EPOCH clock year-acceleration. Each panel represents one organ-specific disease EPOCH clock across MRI-, proteomics-, and metabolomics-derived models. Points denote FUMA-defined independent significant SNPs merged with fastGWA effect estimates; bubble size reflects the available GWAS effect size for each variant. The dashed horizontal line marks *β* = 0. Across clocks, lower-frequency variants generally showed larger absolute effect sizes, consistent with the expected trumpet-shaped genetic architecture.

**Extended Data Figure 9:**
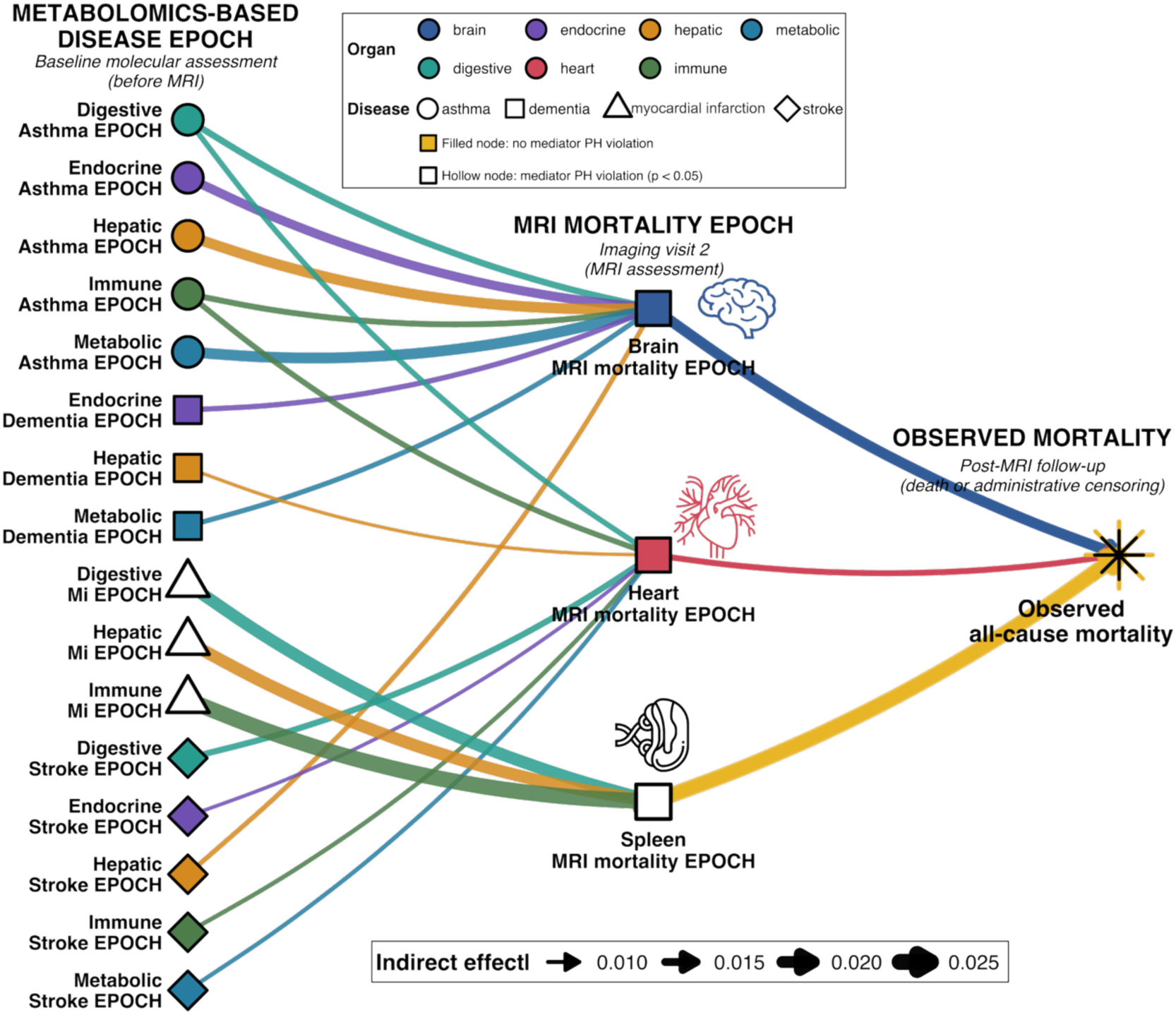
Within-and cross-organ mediation network linking metabolomics-based disease EPOCHs to mortality through MRI mortality EPOCHs. Network representation of the Bonferroni-significant statistical mediation pathways connecting baseline molecular-based disease EPOCH clocks (proteomics-based clock did not survive multiple comparisons), MRI mortality EPOCH clocks measured subsequently at the imaging visit, and all-cause mortality during post-imaging follow-up. The analysis considered 45 molecular disease EPOCHs and 7 MRI mortality EPOCHs, yielding 315 prespecified exposure-mediator combinations. Only the 18 pathways surviving Bonferroni correction (two-sided P-value<0.05/315=1.59×10^-4^) are displayed; all 18 were additionally supported by bootstrap 95% confidence intervals excluding zero. Colors denote the organ system from which each EPOCH was derived, and shapes denote the disease endpoint represented by the baseline metabolomics-based EPOCH. Solid directed edges indicate the statistical mediation sequence from the molecular disease EPOCH to the MRI mortality EPOCH and from the MRI mortality EPOCH to subsequent observed mortality. For each pathway, the thickness of the solid line (*a*) denotes the standardized OLS coefficient relating the baseline disease EPOCH to the subsequent MRI mortality EPOCH, whereas the next solid line (*b*) denotes the Cox log-hazard coefficient relating the MRI mortality EPOCH to post-imaging mortality conditional on the disease EPOCH and covariates. The indirect association was quantified as *a*×*b* on the Cox log-hazard scale. Hollow spleen-associated nodes indicate evidence that the proportional-hazards assumption for the MRI mediator was violated (P_PH_<0.05); these pathways should therefore be interpreted with additional caution.

## Notes

### Competing Interest Statement

The authors have declared no competing interest.

### Author Declarations

Participants provided written informed consent to the corresponding studies. The MULTI Consortium is approved by the Institutional Review Board at Columbia University (AAAV6751).

## References

1. MULTI Consortium et al. Sleep chart of biological ageing clocks in middle and late life. Nature 1–11 (2026) doi:10.1038/s41586-026-10524-5.

2. Wen, J. et al. The genetic architecture of biological age in nine human organ systems. Nat Aging 1–18 (2024) doi:10.1038/s43587-024-00662-8.

3. The MULTI Consortium et al. Multi-organ metabolome biological age implicates cardiometabolic conditions and mortality risk. Nat Commun 16, 4871 (2025).

4. The MULTI Consortium et al. MRI-based multi-organ clocks for healthy aging and disease assessment. Nat Med 1–11 (2025) doi:10.1038/s41591-025-03999-8.

5. Wen, J. Refining the generation, interpretation and application of multi-organ, multi-omics biological aging clocks. Nat Aging 1–17 (2025) doi:10.1038/s43587-025-00928-9.

6. Wen, J. et al. The genetic architecture of multimodal human brain age. Nat Commun 15, 2604 (2024).

7. The MULTI Consortium. Sex-specific biological aging clocks across organs and omics. Nature Medicine (2026) doi:10.1038/s41591-026-04662-6.

8. Oh, H. S.-H. et al. Organ aging signatures in the plasma proteome track health and disease. Nature 624, 164–172 (2023).

9. Argentieri, M. A. et al. Proteomic aging clock predicts mortality and risk of common age-related diseases in diverse populations. Nat Med 30, 2450–2460 (2024).

10. Oh, H. S.-H. et al. Plasma proteomics links brain and immune system aging with healthspan and longevity. Nat Med 31, 2703–2711 (2025).

11. Hernandez, H. et al. The exposome of healthy and accelerated aging across 40 countries. Nat Med 31, 3089–3100 (2025).

12. Legaz, A. et al. The exposome of brain aging across 34 countries. Nat Med 32, 1838–1851 (2026).

13. Zalesky, A., Wen, J. & Tian, Y. E. From whole-body to organ-specific biological age clocks. Nat Aging 6, 961–969 (2026).

14. Cole, J. H. et al. Brain age predicts mortality. Mol Psychiatry 23, 1385–1392 (2018).

15. Poganik, J. R. et al. Biological age is increased by stress and restored upon recovery. Cell Metabolism 35, 807–820.e5 (2023).

16. Smith, S. M., Miller, K. L. & Nichols, T. E. Characterising ongoing brain aging and baseline effects from cross-sectional data. doi:10.1162/IMAG.a.39.

17. Levine, M. E. et al. An epigenetic biomarker of aging for lifespan and healthspan. Aging (Albany NY) 10, 573–591 (2018).

18. Lu, A. T. et al. DNA methylation GrimAge strongly predicts lifespan and healthspan. Aging (Albany NY) 11, 303–327 (2019).

19. Lu, A. T. et al. DNA methylation GrimAge version 2. Aging (Albany NY) 14, 9484–9549 (2022).

20. Goeminne, L. J. E. et al. Plasma protein-based organ-specific aging and mortality models unveil diseases as accelerated aging of organismal systems. Cell Metab 37, 205–222.e6 (2025).

21. Tyshkovskiy, A. et al. Universal transcriptomic hallmarks of mammalian ageing and mortality. Nature 654, 173–188 (2026).

22. Belsky, D. W. et al. DunedinPACE, a DNA methylation biomarker of the pace of aging. Elife 11, e73420 (2022).

23. Whitman, E. T. et al. DunedinPACNI estimates the longitudinal Pace of Aging from a single brain image to track health and disease. Nat Aging 5, 1619–1636 (2025).

24. Tian, Y. E. et al. Heterogeneous aging across multiple organ systems and prediction of chronic disease and mortality. Nat Med 1–11 (2023) doi:10.1038/s41591-023-02296-6.

25. Wen, J. Towards a multi-organ, multi-omics medical digital twin. *Nat*. Biomed. Eng 1–4 (2025) doi:10.1038/s41551-025-01474-w.

26. Ren, P. et al. Imaging-based organ-specific aging clock predicts human diseases and mortality. *npj Digit*. Med. 9, 278 (2026).

27. Wang, Y. et al. Organ-specific proteomic aging clocks predict disease and longevity across diverse populations. Nat Aging 6, 162–180 (2026).

28. Kivimäki, M. et al. Proteomic organ-specific ageing signatures and 20-year risk of age-related diseases: the Whitehall II observational cohort study. The Lancet Digital Health 7, e195–e204 (2025).

29. Xiong, J. et al. Multi-omic underpinnings of heterogeneous aging across multiple organ systems. Cell Genomics 0, (2025).

30. Bycroft, C. et al. The UK Biobank resource with deep phenotyping and genomic data. Nature 562, 203–209 (2018).

31. Bulik-Sullivan, B. K. et al. LD Score regression distinguishes confounding from polygenicity in genome-wide association studies. Nat Genet 47, 291–295 (2015).

32. Yang, J., Lee, S. H., Goddard, M. E. & Visscher, P. M. GCTA: A Tool for Genome-wide Complex Trait Analysis. Am J Hum Genet 88, 76–82 (2011).

33. Zeng, J. et al. Widespread signatures of natural selection across human complex traits and functional genomic categories. Nat Commun 12, 1164 (2021).

34. Wen, J. et al. Neuroimaging endophenotypes reveal underlying mechanisms and genetic factors contributing to progression and development of four brain disorders. *Nat*. Biomed. Eng 1–18 (2025) doi:10.1038/s41551-025-01412-w.

35. The MULTI Consortium et al. Multi-organ AI endophenotypes chart the heterogeneity of brain, eye and heart pan-disease. Nat. Mental Health 1–28 (2026) doi:10.1038/s44220-025-00560-x.

36. Wen, J. et al. Characterizing Heterogeneity in Neuroimaging, Cognition, Clinical Symptoms, and Genetics Among Patients With Late-Life Depression. JAMA Psychiatry (2022) doi:10.1001/jamapsychiatry.2022.0020.

37. Wen, J. et al. Genetic and clinical correlates of two neuroanatomical AI dimensions in the Alzheimer’s disease continuum. Transl Psychiatry 14, 1–14 (2024).

38. Kriegeskorte, N., Simmons, W. K., Bellgowan, P. S. F. & Baker, C. I. Circular analysis in systems neuroscience: the dangers of double dipping. Nat. Neurosci. 12, 535–540 (2009).

39. Schüssler-Fiorenza Rose, S. M., et al. A longitudinal big data approach for precision health. Nat Med 25, 792–804 (2019).

40. Mavrommatis, C. et al. An unbiased comparison of 14 epigenetic clocks in relation to 174 incident disease outcomes. Nat Commun 16, 11164 (2025).

41. Goeminne, L. J. E. et al. Plasma protein-based organ-specific aging and mortality models unveil diseases as accelerated aging of organismal systems. Cell Metabolism 37, 205–222.e6 (2025).

42. Moqri, M. et al. Biomarkers of aging for the identification and evaluation of longevity interventions. Cell 186, 3758–3775 (2023).

43. Kuo, P.-L. et al. Longitudinal changes in epigenetic clocks predict survival in the InCHIANTI cohort. Nat Aging 6, 534–540 (2026).

44. Ferrucci, L., Wilson, D. M., Donegà, S. & Gorospe, M. The energy–splicing resilience axis hypothesis of aging. Nat Aging 2, 182–185 (2022).

45. The MULTI Consortium et al. Brain–heart–eye axis revealed by multi-organ imaging genetics and proteomics. *Nat*. Biomed. Eng 1–23 (2025) doi:10.1038/s41551-025-01506-5.

46. Ibáñez, A. et al. Computational whole-body-exposome models for global precision brain health. Nat Commun 16, 11078 (2025).

47. López-Otín, C., Blasco, M. A., Partridge, L., Serrano, M. & Kroemer, G. Hallmarks of aging: An expanding universe. Cell 186, 243–278 (2023).

48. Wen, J. Multiorgan biological age shows that no organ system is an island. Nat Aging 1–2 (2024) doi:10.1038/s43587-024-00690-4.

49. Kurki, M. I. et al. FinnGen provides genetic insights from a well-phenotyped isolated population. Nature 613, 508–518 (2023).

50. Mbatchou, J. et al. Computationally efficient whole-genome regression for quantitative and binary traits. Nat Genet 53, 1097–1103 (2021).

51. O’Donovan, M. C. What have we learned from the Psychiatric Genomics Consortium. World Psychiatry 14, 291–293 (2015).

52. Simon, N., Friedman, J., Hastie, T. & Tibshirani, R. Regularization Paths for Cox’s Proportional Hazards Model via Coordinate Descent. J Stat Softw 39, 1–13 (2011).

53. Zou, H. & Hastie, T. Regularization and Variable Selection Via the Elastic Net. J. R. Stat. Soc. Ser. B. Stat. Methodol. 67, 301–320 (2005).

54. Wen, J. et al. Convolutional neural networks for classification of Alzheimer’s disease: Overview and reproducible evaluation. Medical Image Analysis 63, 101694 (2020).

55. Samper-González, J. et al. Reproducible evaluation of classification methods in Alzheimer’s disease: Framework and application to MRI and PET data. NeuroImage 183, 504–521 (2018).

56. Wen, J. et al. Reproducible Evaluation of Diffusion MRI Features for Automatic Classification of Patients with Alzheimer’s Disease. Neuroinformatics 19, 57–78 (2021).

57. Harrell, F. E., Lee, K. L. & Mark, D. B. Multivariable prognostic models: issues in developing models, evaluating assumptions and adequacy, and measuring and reducing errors. Stat Med 15, 361–387 (1996).

58. Manichaikul, A. et al. Robust relationship inference in genome-wide association studies. Bioinformatics 26, 2867–2873 (2010).

59. Price, A. L., Zaitlen, N. A., Reich, D. & Patterson, N. New approaches to population stratification in genome-wide association studies. Nat Rev Genet 11, 459–463 (2010).

60. Abraham, G., Qiu, Y. & Inouye, M. FlashPCA2: principal component analysis of Biobank-scale genotype datasets. Bioinformatics 33, 2776–2778 (2017).

61. Wen, J. et al. Genomic loci influence patterns of structural covariance in the human brain. Proceedings of the National Academy of Sciences 120, e2300842120 (2023).

62. Wen, J. Pan-disease dimensions in the brain, eye and heart capture shared and specific heterogeneity. Nat. Mental Health 1–2 (2026) doi:10.1038/s44220-025-00566-5.

63. Wen, J. Too little or too much sleep is linked to faster ageing throughout the body. Nature (2026) doi:10.1038/d41586-026-01564-y.

64. Jiang, L. et al. A resource-efficient tool for mixed model association analysis of large-scale data. Nat Genet 51, 1749–1755 (2019).

65. Watanabe, K., Taskesen, E., van Bochoven, A. & Posthuma, D. Functional mapping and annotation of genetic associations with FUMA. Nat Commun 8, 1826 (2017).

66. Zeng, J. et al. Signatures of negative selection in the genetic architecture of human complex traits. Nat Genet 50, 746–753 (2018).

67. Wen, J. et al. Neuroimaging-AI Endophenotypes of Brain Diseases in the General Population: Towards a Dimensional System of Vulnerability. 2023.08.16.23294179 Preprint at 10.1101/2023.08.16.23294179 (2023).

68. Elliott, L. T. et al. Genome-wide association studies of brain imaging phenotypes in UK Biobank. Nature 562, 210–216 (2018).

69. Zhao, B. et al. Genome-wide association analysis of 19,629 individuals identifies variants influencing regional brain volumes and refines their genetic co-architecture with cognitive and mental health traits. Nat Genet 51, 1637–1644 (2019).

70. Bulik-Sullivan, B. et al. An atlas of genetic correlations across human diseases and traits. Nat Genet 47, 1236–1241 (2015).

71. Hemani, G. et al. The MR-Base platform supports systematic causal inference across the human phenome. eLife 7, e34408 (2018).

72. Skrivankova, V. W. et al. Strengthening the Reporting of Observational Studies in Epidemiology Using Mendelian Randomization: The STROBE-MR Statement. JAMA 326, 1614–1621 (2021).

73. Sanderson, E. et al. Mendelian randomization. Nat Rev Methods Primers 2, 1–21 (2022).

74. Bowden, J. et al. A framework for the investigation of pleiotropy in two-sample summary data Mendelian randomization. Stat Med 36, 1783–1802 (2017).

75. Dafni, U. Landmark analysis at the 25-year landmark point. Circ Cardiovasc Qual Outcomes 4, 363–371 (2011).

