## supplement for "Event-Proximity Organ Clocks in Humans Link Multi-organ Biology to Disease and Mortality"

### Online Supplementary Materials

**Note 1: The definition of genomic loci, independent significant SNP, lead SNP, candidate SNP**
**Note 2: The 46 AI biomarkers across organs and omics layers**
**Note 3: Brain-body risk architecture of the 22 mortality EPOCH clocks with systemic disease endpoints in survival analyses**
**Note 4: Joint analyses of BAG and EPOCH to study resilient aging**
**Note 5: The genetic architecture of the 47 disease-specific EPOCH clocks**
**Note 6: Cross-organ disease-mortality mediation reveals selective convergence on brain, heart and spleen MRI mortality EPOCHs**
**Figure 1: Relationship between mortality EPOCH clocks and paired biological aging clocks across organs and modalities**
**Figure 2: Feature architecture and biological interpretation of the brain proteomics mortality EPOCH**
**Figure 3-24: Circular Manhattan plot of phenome-wide associations in GWAS Catalog for the 22 mortality EPOCH clocks**
**Figure 25: Genetic architecture of brain proteomics mortality EPOCH converges on four interacting biological systems relevant to lifespan**
**Figure 26: Cross-method consistency of SNP heritability estimates for 22 mortality EPOCH clocks**
**Figure 27: Comparisons between the 22 mortality EPOCH clocks and their corresponding BAGs**
**Figure 28: The 22 mortality EPOCH clocks predict future disease onset in the holdout test dataset**
**Figure 29: Comparison of brain proteomic EPOCH with its constituent proteins for Alzheimer's disease onset**
**Figure 30: Brain proteomics mortality EPOCH provides incremental prediction of Alzheimer's disease and heart failure beyond conventional clinical biomarkers**
**Figure 31: Brain MRI mortality EPOCH predicts incident mental and nervous-system disorders (ICD code: G and F) beyond disease-specific AI-derived brain MRI subtypes across survival and fixed-horizon analyses**
**Figure 32: Medication history association with longitudinal changes in metabolomics mortality EPOCH clocks**
**Figure 33: Model performance of asthma EPOCH clocks across organs and modalities**
**Figure 34: Model performance of COPD EPOCH clocks across organs and modalities**
**Figure 35: Model performance of dementia EPOCH clocks across organs and modalities**
**Figure 36: Model performance of MI EPOCH clocks across organs and modalities**
**Figure 37: Model performance of stroke EPOCH clocks across organs and modalities**
**Figure 38: Comparing disease-specific EPOCH clocks against conventional mortality-related biomarkers for all-cause mortality**
**Table 1. First-generation clocks**
**Table 2. Second-generation and outcome-informed clocks**
**Table 3: The characteristics of participants consolidated via the MULTI study**
**Table 4: LDSC statistics for scrutinizing the GWAS of the 22 mortality EPOCH clocks**
**Table 5: GCTA statistics for comparative SNP-based heritability estimates between the 22 mortality EPOCH clocks and corresponding BAGs**

**Table 6: SBayesS statistics for the three key genetic parameters**

**Table 7: Landmark survival analysis to predict subsequent mortality risk for the 4 metabolomics mortality EPOCH clocks' longitudinal changes**

**Table 8: Landmark survival analysis to predict subsequent disease risk for the 4 metabolomics mortality EPOCH clocks' longitudinal changes**

**Table 9: Detailed statistics for the within-organ and cross-organ temporal coupling analysis**

**Table 10: Disease endpoints from FinnGen and PGC**

**Note 1: The definition of genomic loci, independent significant SNP, lead SNP, candidate SNP**

FUMA defined the significant independent SNPs, lead SNPs, candidate SNPs, and genomic risk loci as follows (https://fuma.ctglab.nl/tutorial#snp2gene):

*Independent significant SNPs*

They are defined as SNPs with $P \leq 5\times10^{-8}$ that are independent of each other at the user-defined $r^2$ (set to 0.6 in the current study). We further describe *candidate SNPs* as those in linkage disequilibrium (LD) with independent significant SNPs. FUMA then queries each candidate SNP in the GWAS Catalog to check whether any clinical traits have been reported to be associated with previous GWAS studies.

*Lead SNPs*

Lead SNPs are defined as independent significant SNPs that are also independent of each other at $r^2<0.1$. If multiple independent significant SNPs are correlated at $r^2\geq0.1$, then the one with the lowest individual *P*-value becomes the lead SNP. If $r^2$ threshold is set to 0.1 for the independent significant SNPs, then they would constitute the identical set as the lead SNPs. FUMA thus advises setting $r^2$ to 0.6 or higher.

*Genomic risk loci*

FUMA defines genomic risk loci to include all independent signals physically close or overlapping in a single locus. First, independent significant SNPs dependent on each other at $r^2\geq0.1$ are assigned to the same genomic risk locus. Then, independent significant SNPs with less than the user-defined distance (250 kilobases by default) of one another are merged into the same genomic risk locus – the distance between two LD blocks of two independent significant SNPs is the distance between the closest points from each LD block. Each locus is represented by the SNP within the locus with the lowest *P*-value.

**Note 2: The 46 AI biomarkers across organs and omics layers**

In our recent effort to develop AI-based biomarkers from large-scale multimodal data, we developed 46 AI biomarkers from *in vivo* imaging, plasma proteomics, and plasma metabolomics. These biomarkers span multiple organ systems and molecular layers and can be broadly grouped into two categories: biological aging clocks and disease subtypes.

**a) 26 biological aging clocks**.

We developed 26 BAGs using a unified machine-learning framework with nested cross-validation[1]. BAG was defined as the difference between AI-predicted biological age and chronological age. These clocks comprised *i*) 3 brain MRI-based BAGs (MRIBAGs) derived from multimodal brain MRI, including grey matter MRIBAG based on regional grey matter volume, white matter MRIBAG based on white-matter tract features, and functional connectivity MRIBAG based on resting-state functional MRI[2]; 6 additional MRIBAGs derived from organ-specific MRI features of the heart, liver, adipose tissue, spleen, kidney and pancreas[3]; *ii*) 11 proteomics-based BAGs (ProtBAGs) derived from organ-enriched plasma proteins defined using the Human Protein Atlas, spanning the female reproductive, male reproductive, pulmonary, brain, eye, hepatic, renal, endocrine, immune, heart, and skin systems[4]; and *iii*) 5 metabolomics-based BAGs (MetBAGs) derived from organ-associated plasma metabolites, spanning the endocrine, digestive, hepatic, immune, and metabolic systems[5]. Refer to the original reference papers for detailed information.

Here is a descriptive summary of these biomarkers:

- **Brain MRIBAG**: brain age derived from gray matter imaging features from multimodal MRI;
- **Brain GM MRIBAG**: brain age derived from gray matter volumetric features from T1-weighted MRI;
- **Brain WM MRIBAG**: brain age derived from white matter fractional anisotropy (FA) tract features from diffusion MRI;
- **Brain FC MRIBAG**: brain age derived from functional connectivity (FC) features from resting-state functional MRI;
- **Heart MRIBAG**: heart age derived from imaging features from cardiac MRI;
- **Liver MRIBAG**: liver age derived from imaging features from liver MRI;
- **Adipose MRIBAG**: adipose age derived from imaging features from several abdominal MRIs;
- **Spleen MRIBAG**: spleen age derived from imaging features from spleen MRI;
- **Kidney MRIBAG**: kidney age derived from imaging features from kidney MRI;
- **Pancreas MRIBAG**: pancreas age derived from imaging features from pancreas MRI;
- **Female/male reproductive ProtBAG**: female/male reproductive system age derived from organ-enriched proteins defined by the HPA platform;
- **Pulmonary ProtBAG**: pulmonary age derived from organ-enriched proteins defined by the HPA platform;
- **Brain ProtBAG**: brain age derived from organ-enriched proteins defined by the HPA platform;
- **Eye ProtBAG**: eye age derived from organ-enriched proteins defined by the HPA platform;
- **Hepatic ProtBAG**: hepatic age derived from organ-enriched proteins defined by the HPA platform;

- **Renal ProtBAG**: renal age derived from organ-enriched proteins defined by the HPA platform;
- **Endocrine ProtBAG**: endocrine age derived from organ-enriched proteins defined by the HPA platform;
- **Immune ProtBAG**: immune age derived from organ-enriched proteins defined by the HPA platform;
- **Heart ProtBAG**: heart age derived from organ-enriched proteins defined by the HPA platform;
- **Skin ProtBAG**: skin age derived from organ-enriched proteins defined by the HPA platform;
- **Endocrine MetBAG**: endocrine age derived from organ-associated metabolites;
- **Digestive MetBAG**: digestive age derived from organ-associated metabolites;
- **Hepatic MetBAG**: hepatic age derived from organ-associated metabolites;
- **Immune MetBAG**: immune age derived from organ-associated metabolites;
- **Metabolic MetBAG**: metabolic age derived from organ-associated metabolites.

**b) 20 disease subtypes**.

We also developed 20 disease subtype models using our weakly supervised clustering framework[6] to dissect disease heterogeneity in the brain, eye, and heart from *in vivo* imaging data. These included 9 brain-specific subtypes[7] for AD (AD1-2), late-life depression (LLD1-2), schizophrenia (SCZ1-2), and autism spectrum disorder (ASD1-3), as well as 11 pan-disease subtypes[8] capturing shared and distinct patterns of disease heterogeneity across organs, including 6 brain subtypes (PanBrain1-6), 3 retinal subtypes (PanEye1-3), and 2 heart subtypes (PanHeart1-2). Refer to the original reference papers for detailed information.

Here is a descriptive summary of these biomarkers:

- **PanBrain1:** Pan-disease brain subtype 1 shows a pronounced temporal-lobe and inferior-cerebellar volume loss and relative enlargement in the superior cerebellum;
- **PanBrain2:** Pan-disease brain subtype 2 is characterized by diffuse atrophy together with enlarged cerebellum volume;
- **PanBrain3:** Pan-disease brain subtype 3 shows widespread global brain atrophy with relative enlargement in the frontal region;
- **PanBrain4:** Pan-disease brain subtype 4 shows increased brain volume mainly in the frontal and occipital lobes, largely sparing the temporal poles and deep subcortical structures;
- **PanBrain5:** Pan-disease brain subtype 5 shows enlarged brain volume in the frontal and temporal lobes;
- **PanBrain6:** Pan-disease brain subtype 6 shows frontal-lobe atrophy with enlarged volume in deep subcortical structures;
- **PanEye1:** Pan-disease eye subtype 1 shows the most pronounced retinal thinning between the external limiting membrane and the inner-outer photoreceptor segment layer;
- **PanEye2:** Pan-disease eye subtype 2 shows globally thinner retina, especially reduced overall macular thickness, together with thickening of the outer nuclear and outer plexiform layer;
- **PanEye3:** Pan-disease eye subtype 3 shows greater macular thickness and increased vertical cup-to-disc ratio;

- **PanHeart1:** Pan-disease heart subtype 1 shows a prominent increase in left-ventricular myocardial wall thickness;
- **PanHeart2:** Pan-disease heart subtype 2 shows globally increased cardiac and aortic functional measures, with reduced left- and right-atrial ejection fraction;
- **AD1:** AD subtype 1 shows global diffuse atrophy;
- **AD2:** AD subtype 2 shows a focal medial temporal lobe atrophy;
- **SCZ1**: SCZ subtype 1 shows global/widespread brain atrophy, especially lower grey matter volume across cortical and subcortical regions;
- **SCZ2:** SCZ subtype 2 shows expanded basal ganglia volume, with relatively preserved cortical anatomy;
- **LLD1:** LLD subtype 1 shows augmented subcortical volume;
- **LLD2:** LLD subtype 2 shows overall brain atrophy;
- **ASD1:** ASD subtype 1 shows reduced overall brain volume;
- **ASD2:** ASD subtype 2 shows expanded subcortical volume;
- **ASD3:** ASD subtype 3 is associated with increased cortical volume.

**Note 3: Brain-body risk architecture of the 22 mortality EPOCH clocks with systemic disease endpoints in survival analyses**

To further characterize the disease relevance of mortality EPOCH, we examined the 22 mortality EPOCH clocks alone across future incident disease endpoints. This analysis revealed a broad brain-body risk architecture, with more than 13,000 Bonferroni-significant EPOCH-DE associations spanning diverse organ domains. The strongest signal was observed for molecular clocks, especially metabolomics and proteomics EPOCH clocks (with notably much larger sample sizes than MRI), whereas MRI-based clocks showed fewer but more organ-specific associations. Nearly all significant associations were risk-increasing (HR>1), indicating that higher mortality EPOCH acceleration generally reflects a systemic event-proximal state associated with elevated future disease burden.

As shown in **Extended Data Figure 5**, mortality EPOCH clocks are associated with a wide range of disease endpoints, including cardiovascular, digestive, musculoskeletal, clinical symptom, pulmonary, kidney/renal, neurologic, endocrine/metabolic, cancer/neoplasm, infectious, immune/blood, skin, and reproductive endpoints. The largest disease-domain burdens were observed for cardiovascular, musculoskeletal, digestive, clinical symptom, pulmonary, kidney/renal, neurologic, and endocrine/metabolic endpoints, suggesting that mortality EPOCH does not merely capture mortality risk in isolation but instead indexes a broad preclinical vulnerability to future multi-system disease. Representative endpoints included hypertension, pacemaker status, pneumonia, CKD, diabetic kidney disease, liver cancer, hepatic fibrosis, anemia, gout, rheumatoid arthritis, dermatitis, infectious gastroenteritis, abdominal/pelvic pain, and transplant-related complications.

The clock-level architecture further showed both organ-specific and cross-organ patterns. For example, brain and eye EPOCH clocks were connected to neurologic, psychiatric, and ophthalmic endpoints, while pulmonary, renal, hepatic, and cardiovascular EPOCH clocks showed expected links to respiratory, kidney, liver, and heart-related outcomes. At the same time, several clocks, particularly endocrine, hepatic, digestive, and immune metabolomics EPOCH clocks, as well as immune, endocrine, hepatic, reproductive, and brain proteomics EPOCH clocks, showed extensive cross-domain connectivity. These widespread links support the interpretation that mortality EPOCH captures coordinated brain-body disease susceptibility rather than isolated organ dysfunction.

**Note 4: Joint analyses of BAG and EPOCH to study resilient aging**
Phenotypic (**Supplementary Figure 1**) and genetic (**Fig. 2B**) correlations indicate that BAG and EPOCH share partially overlapping, but distinct, biological architectures, echoing their different modeling targets. Rather than replacing BAG, EPOCH provides a complementary dimension of event-proximal vulnerability.

To demonstrate this, we used the brain proteomics-based BAG and EPOCH clocks (showing a moderate positive correlation with each other) as an example to identify biologically informative discordant/concordant states. In particular, BAG-high/EPOCH-low individuals may represent candidate resilient agers, showing advanced age-related biology without correspondingly high mortality proximity, whereas BAG-low/EPOCH-high individuals may reflect latent vulnerability despite relatively youthful age-related profiles (**Method 4c**).

Brain proteomic BAG and mortality EPOCH showed substantial but incomplete concordance, indicating that generalized biological aging and event-proximal vulnerability capture overlapping but distinct biological information (**Extended Data Figure 6A**). Residualizing EPOCH on BAG revealed marked inter-individual discordance, with some participants showing substantially lower or higher EPOCH than expected for their degree of biological aging. Using the joint tails of BAG and the EPOCH-BAG residual, we further defined four aging states, including candidate resilient agers (CRA), who exhibited high BAG but unexpectedly low event-proximal vulnerability, in contrast to concordant unfavorable agers (CUA) with high values for both dimensions (**Extended Data Figure 6B**).

GWAS of the EPOCH-BAG residual identified 19 genomic risk loci, indicating a distributed genetic architecture underlying discordance between generalized biological aging and event-proximal vulnerability (**Extended Data Figure 6C**). The top lead variants included coded effect alleles associated with both lower and higher residuals, motivating subsequent molecular-QTL alignment to infer resilience-associated expression directions. Directional GTEx v8 eQTL mapping identified 40 systemic resilience-associated expression genes, which showed heterogeneous expression across brain and peripheral tissues (**Extended Data Figure 6D**). Functional enrichment converged particularly on lipid and membrane-transport biology, including high-density lipoprotein particle remodeling, lipid transporter activity and ABC transporter pathways, with additional trait enrichments related to cardiovascular health, neurodegeneration and longevity (**Extended Data Figure 6E**).

Comparing CRA with CUA revealed a broad neuroimaging signature of candidate resilience (**Extended Data Figure 6F-G**). Of 48 diffusion MRI features, 23 fractional anisotropy tracts survived within-modality BH correction, all in the CRA-associated direction, with the strongest effects observed in the right anterior limb of the internal capsule and right medial lemniscus; bilateral cerebellar peduncles, corona radiata, and external capsules were also implicated. Similarly, 36 of 119 T1 gray-matter volumes survived within-modality BH correction, again with all significant coefficients positive, indicating larger regional volumes in CRA relative to CUA. Together, these findings suggest that lower-than-expected event-proximal vulnerability despite advanced brain proteomic aging is accompanied by relatively preserved white matter microstructure and gray matter structure. (**Extended Data Figure 6F-G**).

To sum up, EPOCH and BAG quantify complementary dimensions of human aging. BAG captures generalized age-related biological variation, whereas EPOCH captures biological variation aligned with proximity to adverse clinical events. Their discordance identifies individuals in whom biological aging and event-proximal vulnerability are uncoupled, providing a framework to investigate resilience and latent susceptibility.

**Note 5: The genetic architecture of the 47 disease-specific EPOCH clocks**

The disease-specific EPOCH clocks showed broad genetic connectivity with external disease endpoints, supporting the idea that event-proximity clocks capture inherited vulnerability to disease processes rather than only phenotypic prediction from baseline data. Across FinnGen and PGC DEs, the strongest signals were dominated by metabolomics-based EPOCH clocks, particularly hepatic, metabolic, endocrine, digestive and immune clocks. Asthma EPOCH clocks showed genetic correlations with pain, dorsalgia, joint pain, analgesic use, autoimmune endpoints and ADHD; MI EPOCH clocks were strongly connected to type 2 diabetes (T2D), diabetes medication use and diabetes complications; stroke EPOCH clocks were linked to antihypertensive use, heart failure/hypertension, autoimmune endpoints and diabetes-related traits; and dementia EPOCH clocks showed correlations with autoimmune, endocrine and diabetes-related endpoints. These patterns suggest that disease-specific EPOCH clocks do not merely recapitulate the target disease itself. Instead, they encode broader genetic liabilities spanning inflammation, metabolism, vascular function, endocrine regulation and systemic disease burden.

These results are particularly relevant to brain-body biology. For neurological and vascular outcomes such as dementia and stroke, the genetic signals were not confined to brain-specific axes. Instead, dementia and stroke EPOCH clocks shared architecture with peripheral immune, hepatic, endocrine, digestive and metabolic disease liabilities. This supports a model in which brain disease risk is still being positioned at the center, yet also embedded in systemic biology: cardiometabolic dysfunction, immune activation, endocrine dysregulation and inflammatory disease burden may create a genetically mediated background in which neurodegeneration and cerebrovascular disease emerge. Thus, the EPOCH framework positions brain disorders not as isolated brain-local processes, but as outcomes of distributed organ-system vulnerability.

When linked with previously developed AI biomarkers, the disease-specific EPOCH clocks showed strong genetic overlap with organ-specific BAGs and disease-subtype biomarkers, indicating that EPOCH clocks retain meaningful links to established biological-aging architecture while extending it toward future clinical events. The strongest modality- and organ-aligned correlations included MI heart proteomics EPOCH with Heart ProtBAG, dementia heart proteomics EPOCH with Heart ProtBAG, MI and stroke pulmonary proteomics EPOCH with Pulmonary ProtBAG, stroke brain proteomics EPOCH with Brain ProtBAG, and asthma spleen MRI EPOCH with spleen MRIBAG. These results suggest that EPOCH clocks preserve organ-specific biological-aging information but also reinterpret it through the lens of event proximity. In other words, organ aging and event risk are genetically related but not identical. BAGs quantify deviation from expected biological age, whereas EPOCH clocks identify the organ-specific genetic architecture most relevant to future disease trajectories.

The strongest and most widespread genetic correlations were observed between the disease-specific EPOCH clocks and the 22 mortality EPOCH clocks. This result is expected but biologically informative. Disease EPOCH clocks were highly correlated with mortality EPOCH clocks from the same organ and modality, such as MI heart proteomics with heart mortality EPOCH, MI pulmonary proteomics with pulmonary mortality EPOCH, stroke pulmonary proteomics with pulmonary mortality EPOCH, dementia immune proteomics with immune mortality EPOCH, and dementia digestive metabolomics with digestive mortality EPOCH. These same-organ correlations indicate that disease-specific and mortality-specific event-proximity clocks share a common genetic basis of organ vulnerability. At the same time, many correlations crossed organ boundaries, especially among immune, hepatic, endocrine, metabolic, and reproductive-system mortality clocks, suggesting that mortality risk is organized by system-wide genetic axes rather than by isolated organs.

Together, these results support a multi-scale model of EPOCH genetic architecture. First, disease-specific EPOCH clocks connect to external clinical and disease endpoints, revealing shared inherited liabilities across cardiometabolic, immune, inflammatory, pain-related, and neuropsychiatric domains. Second, they connect to existing AI-derived BAGs and disease-subtype biomarkers, showing that event-proximity clocks are rooted in organ-aging biology but are not reducible to age acceleration. Third, they connect strongly to mortality EPOCH clocks, indicating that disease-specific event risk and all-cause mortality risk share a multi-organ genetic foundation. This architecture reinforces the central concept of brain-body interconnection: future disease risk is not encoded within a single organ, but emerges from genetically coordinated vulnerabilities across the brain, immune system, liver, endocrine system, cardiopulmonary axis and systemic metabolism.

**Note 6: Cross-organ disease-mortality mediation reveals selective convergence on brain, heart and spleen MRI mortality EPOCHs**

The statistical mediation analysis revealed a highly structured rather than diffuse relationship between baseline molecular disease EPOCHs and subsequent mortality. Of the 315 molecular disease EPOCH-MRI mortality EPOCH combinations, 18 indirect associations survived stringent Bonferroni correction. All 18 also had bootstrap confidence intervals excluding zero, indicating that the independently resampled estimates supported the significant delta-method results.

All 18 Bonferroni-significant exposures were metabolomics-based disease EPOCHs. None of the proteomics-derived disease EPOCHs survived the multiple correction, potentially due to the limited sample sizes of the proteomics data (N~500k). The significant baseline molecular signals originated from five organ systems, including digestive, endocrine, hepatic, immune, and metabolic, and converged on 3 of the 7 MRI mortality EPOCHs: brain, heart, and spleen. Eight of the 18 significant pathways involved the brain MRI mortality EPOCH, seven involved the heart MRI mortality EPOCH, and three involved the spleen MRI mortality EPOCH. Similarly, the brain and heart MRI populations are overall larger than the other organ imaging data (e.g., liver).

The strongest recurring disease-specific pattern involved asthma. Seven Bonferroni-significant pathways involved an asthma EPOCH. Five distinct organ-specific metabolomic asthma EPOCHs, digestive, endocrine, hepatic, immune, and metabolic, were linked to the brain MRI mortality EPOCH. In addition, digestive and immune asthma EPOCHs were linked to the heart MRI mortality EPOCH. The convergence onto the brain was particularly consistent. For example, hepatic metabolomics asthma EPOCH acceleration was positively associated with subsequent brain MRI mortality EPOCH acceleration ($a$=0.055), while the brain MRI mortality EPOCH was strongly associated with mortality in the conditional Cox model ($b$=0.358; HR=1.43). Their resulting indirect association was $a \times b$=0.0198, with a bootstrap 95% CI of 0.0131-0.0275. Metabolic asthma showed a similar pattern ($a$=0.059, $b$=0.343, $a \times b$=0.0201), as did endocrine, digestive, and immune asthma EPOCHs.

These recurrent associations suggest that molecular signatures of asthma-related disease proximity across several peripheral physiological systems are associated with a common later brain MRI phenotype that itself carries substantial mortality information. Importantly, this should not be interpreted as evidence that asthma causes brain ageing or that the brain causally mediates asthma-related mortality. Rather, the pattern identifies a reproducible temporal statistical bridge between earlier systemic molecular disease susceptibility and a later brain-derived mortality phenotype.

A second topology involved stroke-related metabolomic EPOCHs. Digestive, endocrine, immune, and metabolic stroke EPOCHs were each significantly mediated through the heart MRI mortality EPOCH. Despite originating from distinct molecular organ systems, these pathways showed similar effect structures. For example, the digestive stroke EPOCH had $a$=0.052 with the heart MRI mortality EPOCH, while the heart mediator had $b$=0.260, yielding an indirect log-hazard association of 0.0135. Corresponding indirect estimates for endocrine, immune, and metabolic stroke EPOCHs were approximately 0.0100, 0.0110, and 0.0109, respectively. Hepatic stroke constituted an exception to this predominant heart pattern, showing significant mediation through the brain MRI mortality EPOCH.

Thus, the network suggests that metabolomic states associated with proximity to stroke across multiple physiological systems converge particularly strongly on subsequent cardiac mortality-related imaging features. This is consistent with a cross-organ disease framework in which the molecular manifestation of a clinically brain-defined disease may be embedded in broader cardiovascular and systemic physiology.

Three significant pathways were involved in dementia-related molecular EPOCHs. Endocrine and metabolic metabolomic dementia EPOCHs were mediated through the brain MRI mortality EPOCH, whereas the hepatic metabolomics dementia EPOCH was mediated through the heart MRI mortality EPOCH. Disease-specific EPOCHs appear to encode distributed molecular states whose downstream mortality associations can emerge through anatomically distinct organs.

The spleen MRI mortality EPOCH findings should be interpreted with additional caution because the proportional hazards assumption for the mediator was violated in the significant MI-related mediation models. Schoenfeld residual tests indicated that the association between the spleen MRI mortality EPOCH and subsequent mortality was not constant over follow-up time. Although these spleen-mediated associations remained significant after Bonferroni correction and were supported by bootstrap confidence intervals, they warrant confirmation in sensitivity analyses that allow the mediator effect to vary with time (**Supplementary File 11**).

**Figure 1: Relationship between mortality EPOCH clocks and paired biological aging clocks across organs and modalities**

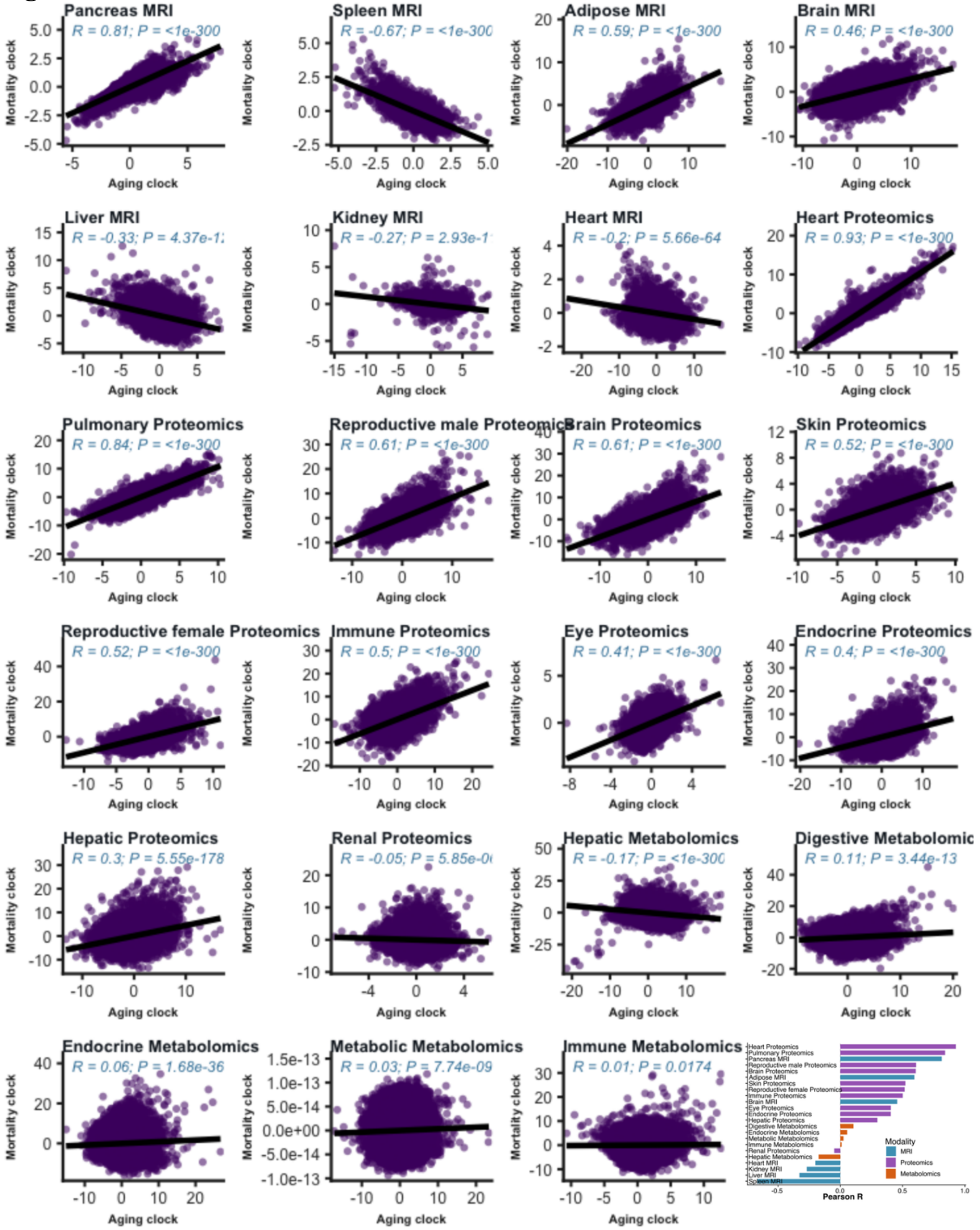


Scatterplots show the relationship between 22 mortality EPOCH clocks and their paired biological age gaps (BAGs) across matched organ systems and data modalities. For each panel, the x-axis shows the paired aging clock/BAG acceleration, and the y-axis shows the corresponding mortality EPOCH acceleration. Each point represents one participant, and the black line indicates the fitted linear trend. Pearson correlation coefficients and P values are shown within each panel. Several proteomics-based mortality EPOCH clocks, including heart, pulmonary, reproductive male, brain, and immune proteomics, showed strong positive correlations with their paired BAGs, indicating substantial shared age-related variation. In contrast, several MRI- and metabolomics-based clocks showed weaker or inverse correlations, suggesting that mortality EPOCH captures event-proximal risk signals that are not simply equivalent to biological age acceleration. The inset bar plot summarizes Pearson correlations across all clock pairs, ranked by correlation strength and colored by modality. Overall, these analyses demonstrate that mortality EPOCH is related to, but distinct from, conventional organ-specific BAGs, supporting its role as a complementary framework for modeling future mortality risk. Of note, because the metabolic metabolomics EPOCH clock showed an abnormal year-scale range on the y-axis, suggesting poor calibration of the acceleration estimates, we excluded this clock from downstream association analyses.

**Figure 2: Feature architecture and biological interpretation of the brain proteomics mortality EPOCH**

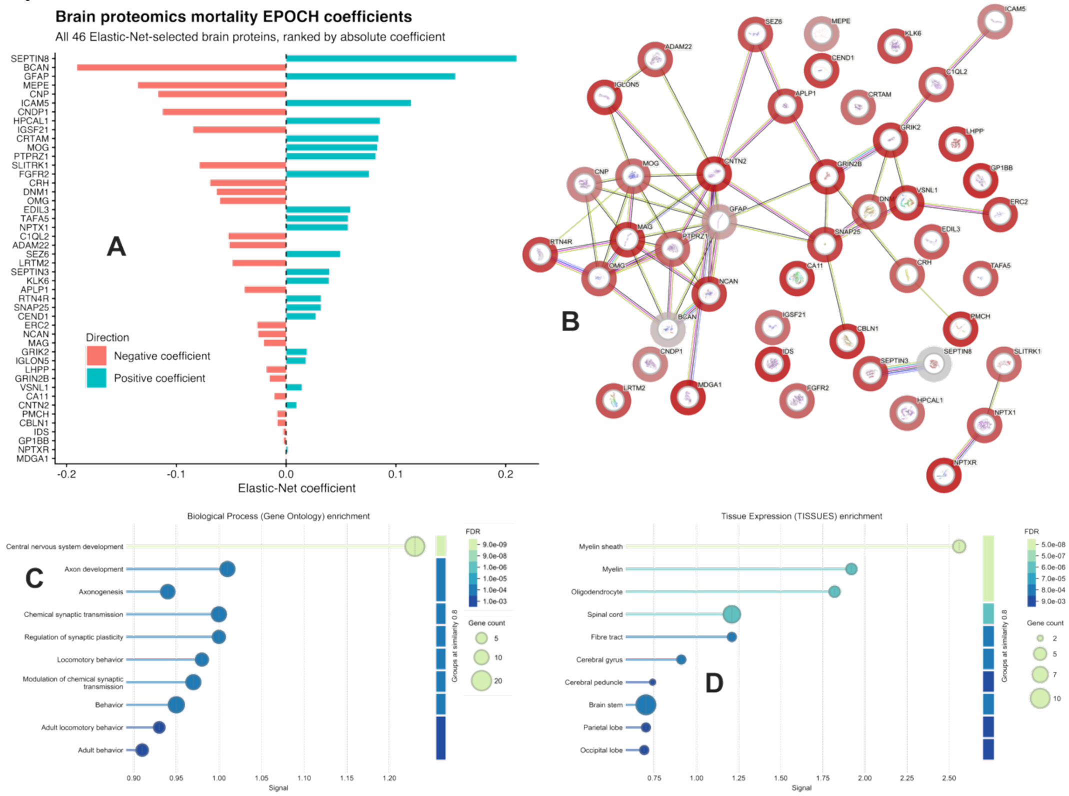


**A**) Elastic-Net Cox coefficients of the 53 brain proteomic features retained in the mortality EPOCH, ranked by absolute coefficient magnitude. Positive coefficients indicate that higher protein levels contribute to a higher model-predicted relative mortality hazard, whereas negative coefficients contribute to a lower predicted relative hazard, conditional on the covariates in the multivariable model. **B**) STRING protein-association network of the non-zero brain proteomic features retained by Elastic Net. Nodes represent proteins and edges represent STRING-supported functional or physical associations; the different edge colors denote distinct evidence channels. **C**) Gene Ontology Biological Process enrichment of the selected proteins. Enriched processes prominently included central nervous system development, axon development, neurogenesis, chemical synaptic transmission, regulation of synaptic plasticity, and modulation of chemical synaptic transmission, indicating convergence on neuronal development and synaptic function. **D**) Tissue-expression enrichment of the selected proteins using the STRING TISSUES resource, demonstrating preferential enrichment in brain and other central nervous system-related tissues. In C-D, horizontal position represents the STRING enrichment signal, point size represents the number of contributing proteins, and color represents the false discovery rate (FDR).

**Figure 3: Circular Manhattan plot of phenome-wide associations in GWAS Catalog for the spleen MRI mortality EPOCH clock**

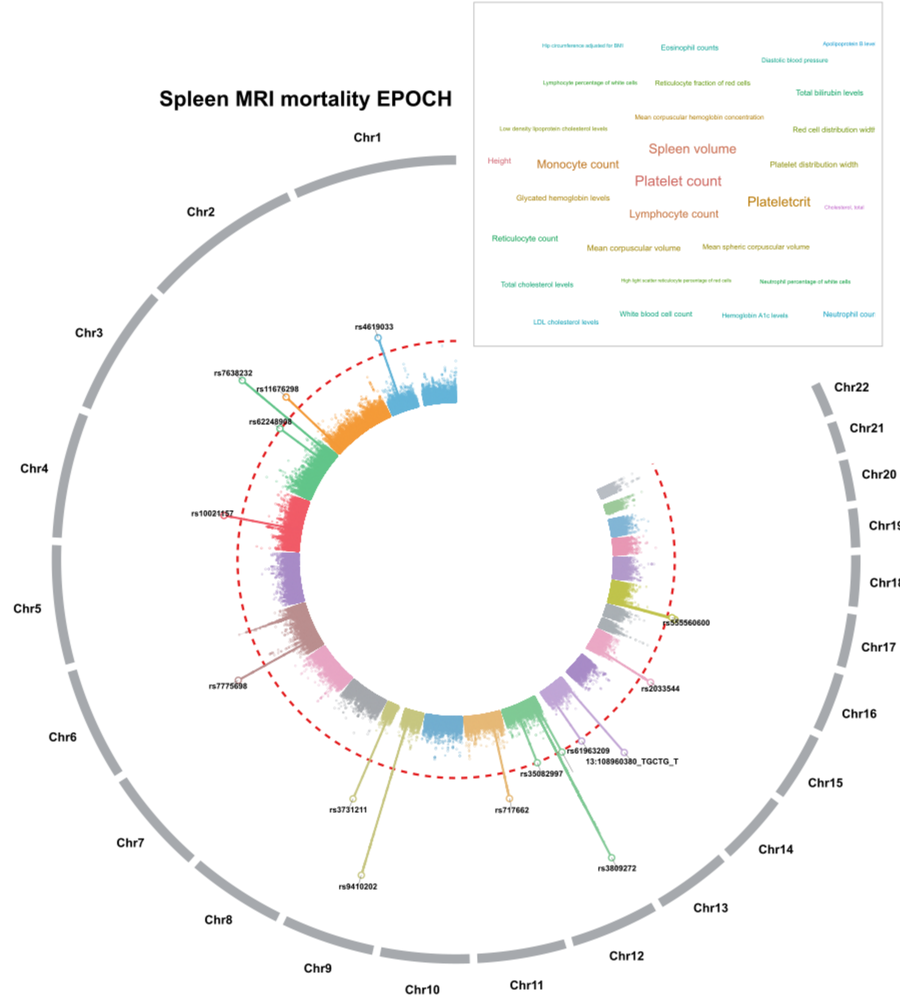


Genome-wide association results from fastGWA are displayed across chromosomes 1-22, with variants positioned according to genomic location and radial distance representing ($-\log_{10}(P)$). Chromosomes are shown in alternating colors, and red-colored dashed circles indicate the genome-wide significance threshold (P-value$<5\times10^{-8}$). The locus-level top SNP from each FUMA-defined genomic risk locus is highlighted and labeled using the SNP ID. The word cloud summarizes GWAS Catalog phenotypic categories previously associated with these loci, with text size proportional to the number of distinct loci linked to each category.

**Figure 4: Circular Manhattan plot of phenome-wide associations in GWAS Catalog for the skin proteomics mortality EPOCH clock**

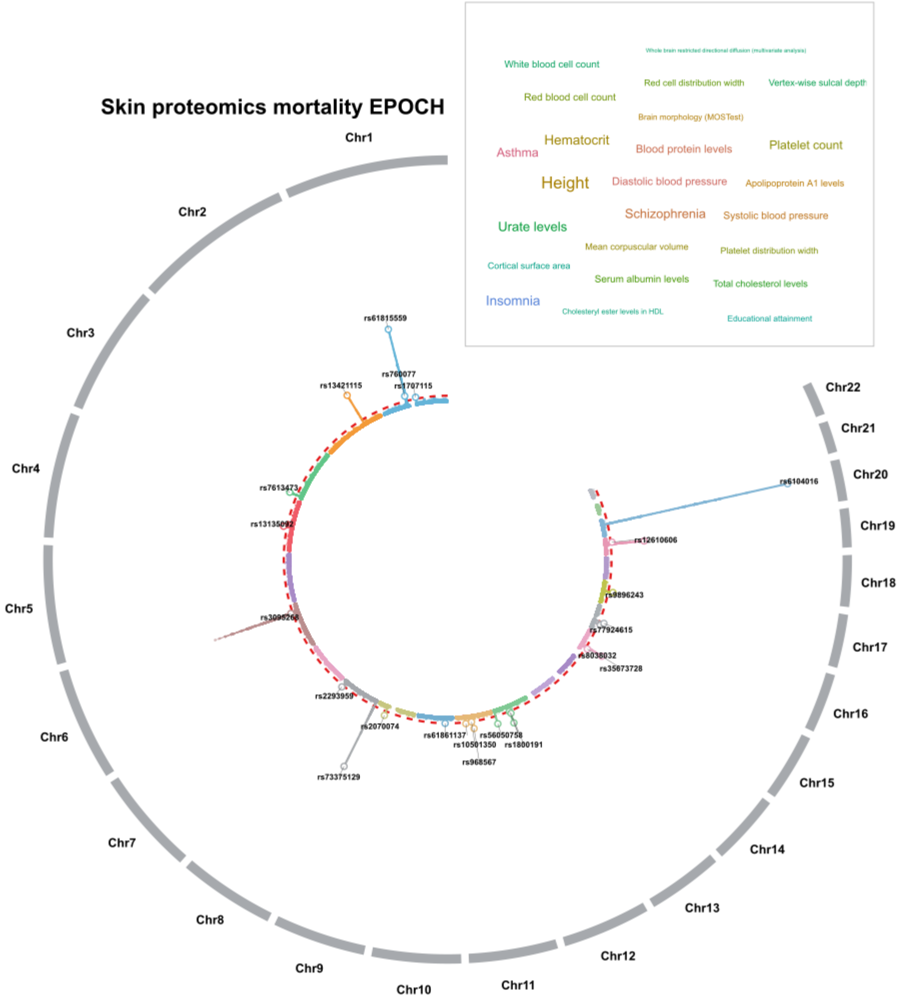


Genome-wide association results from fastGWA are displayed across chromosomes 1-22, with variants positioned according to genomic location and radial distance representing ($-\log_{10}(P)$). Chromosomes are shown in alternating colors, and red-colored dashed circles indicate the genome-wide significance threshold (P-value$<5\times10^{-8}$). The locus-level top SNP from each FUMA-defined genomic risk locus is highlighted and labeled using the SNP ID. The word cloud summarizes GWAS Catalog phenotypic categories previously associated with these loci, with text size proportional to the number of distinct loci linked to each category.

**Figure 5: Circular Manhattan plot of phenome-wide associations in GWAS Catalog for the reproductive (male) proteomics mortality EPOCH clock**

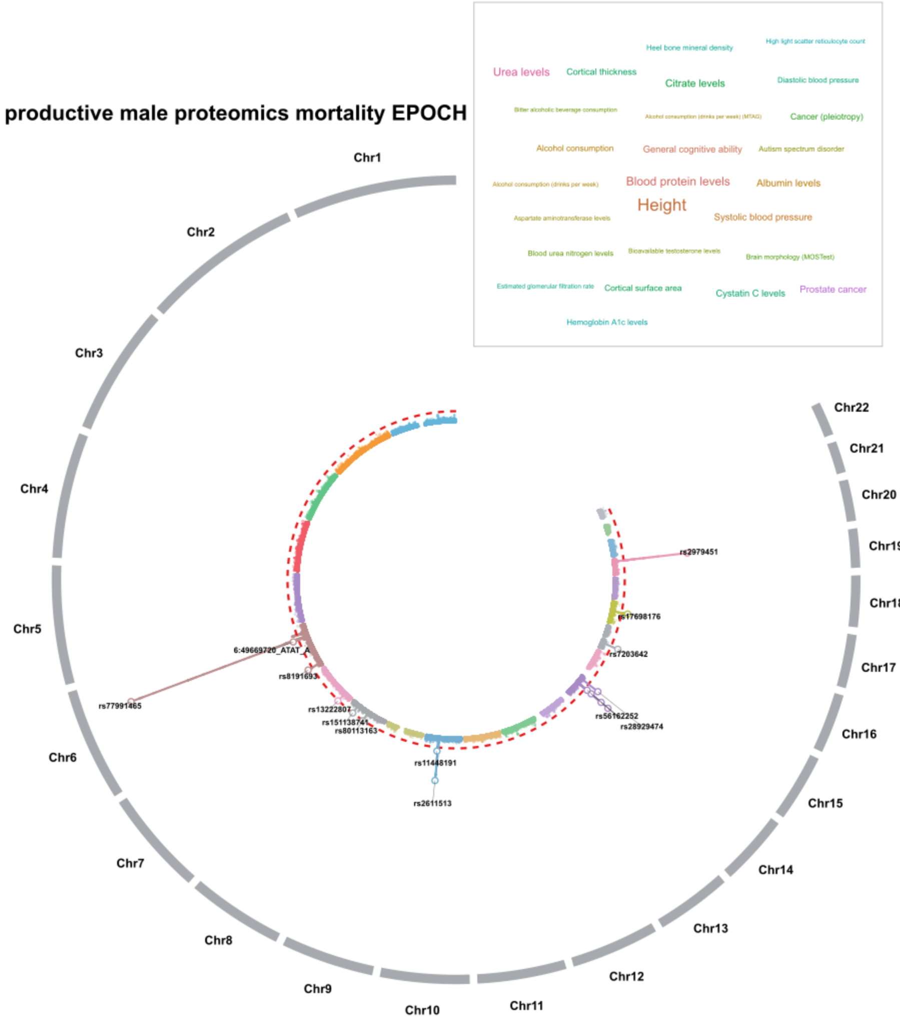


Genome-wide association results from fastGWA are displayed across chromosomes 1-22, with variants positioned according to genomic location and radial distance representing ($-\log_{10}(P)$). Chromosomes are shown in alternating colors, and red-colored dashed circles indicate the genome-wide significance threshold (P-value<$5\times10^{-8}$). The locus-level top SNP from each FUMA-defined genomic risk locus is highlighted and labeled using the SNP ID. The word cloud summarizes GWAS Catalog phenotypic categories previously associated with these loci, with text size proportional to the number of distinct loci linked to each category.

**Figure 6: Circular Manhattan plot of phenome-wide associations in GWAS Catalog for the reproductive (female) proteomics mortality EPOCH clock**

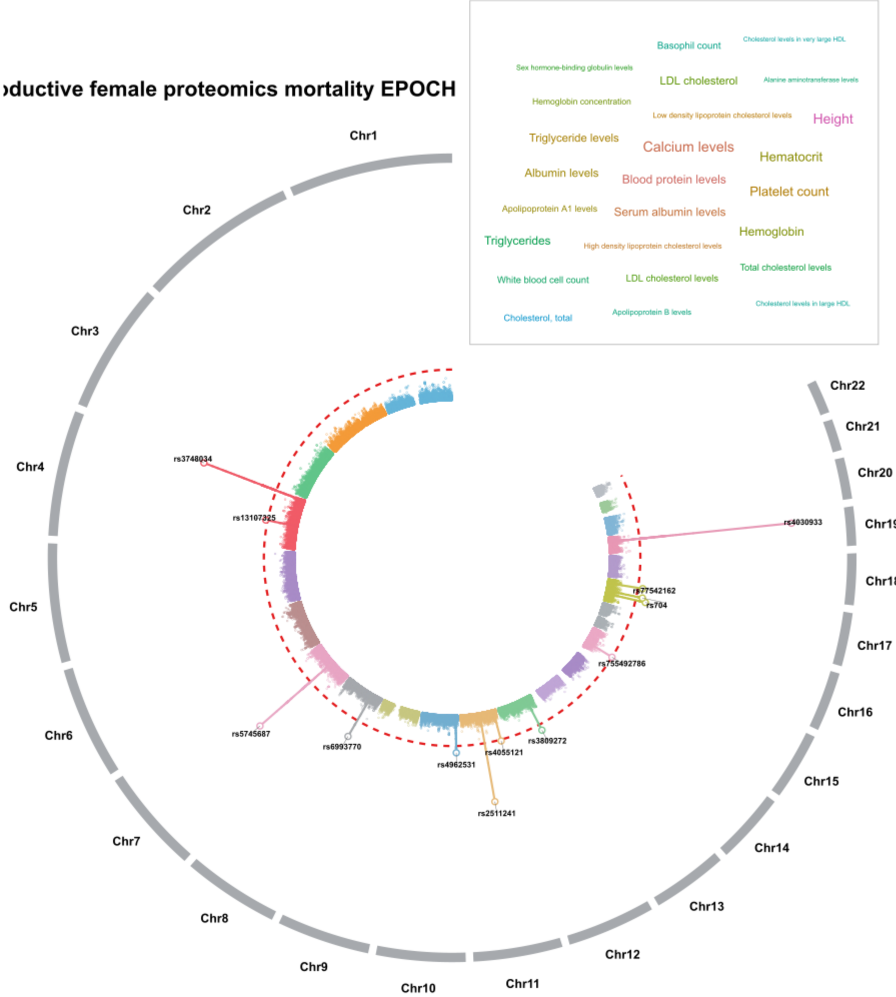


Genome-wide association results from fastGWA are displayed across chromosomes 1-22, with variants positioned according to genomic location and radial distance representing ($-\log_{10}(P)$). Chromosomes are shown in alternating colors, and red-colored dashed circles indicate the genome-wide significance threshold (P-value<$5\times10^{-8}$). The locus-level top SNP from each FUMA-defined genomic risk locus is highlighted and labeled using the SNP ID. The word cloud summarizes GWAS Catalog phenotypic categories previously associated with these loci, with text size proportional to the number of distinct loci linked to each category.

**Figure 7: Circular Manhattan plot of phenome-wide associations in GWAS Catalog for the renal proteomics mortality EPOCH clock**

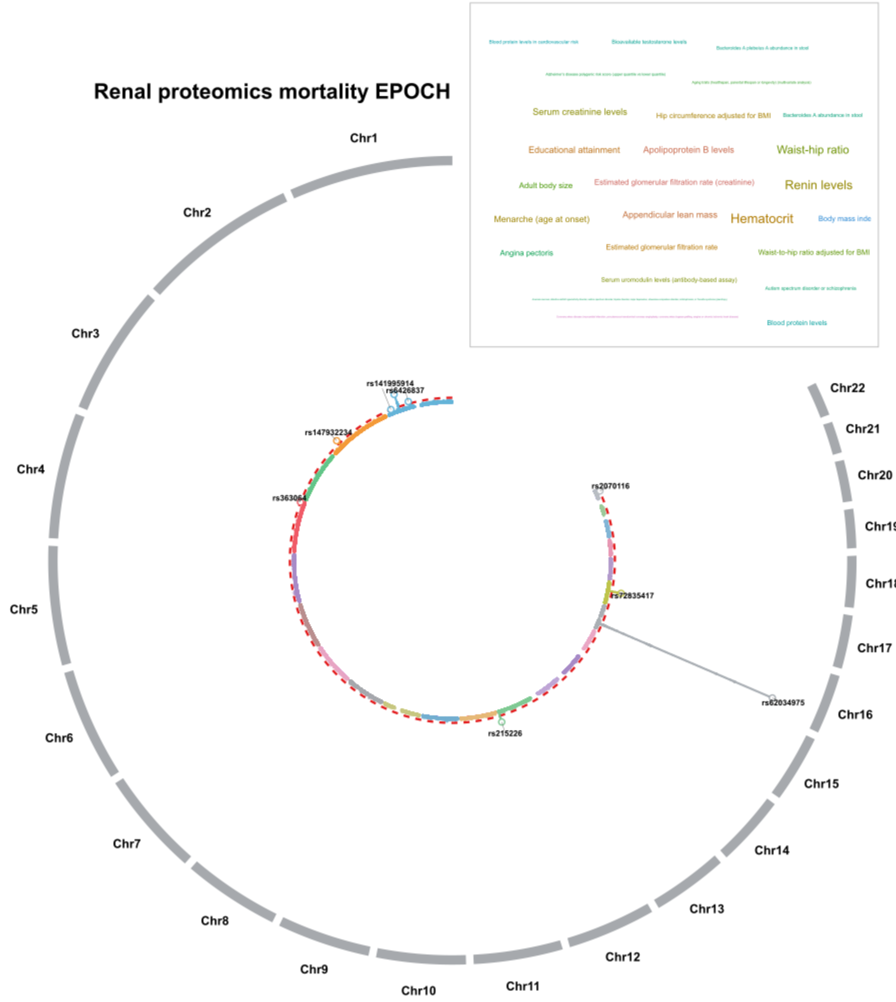


Genome-wide association results from fastGWA are displayed across chromosomes 1-22, with variants positioned according to genomic location and radial distance representing ($-\log_{10}(P)$). Chromosomes are shown in alternating colors, and red-colored dashed circles indicate the genome-wide significance threshold (P-value$<5\times10^{-8}$). The locus-level top SNP from each FUMA-defined genomic risk locus is highlighted and labeled using the SNP ID. The word cloud summarizes GWAS Catalog phenotypic categories previously associated with these loci, with text size proportional to the number of distinct loci linked to each category.

**Figure 8: Circular Manhattan plot of phenome-wide associations in GWAS Catalog for the pulmonary proteomics mortality EPOCH clock**

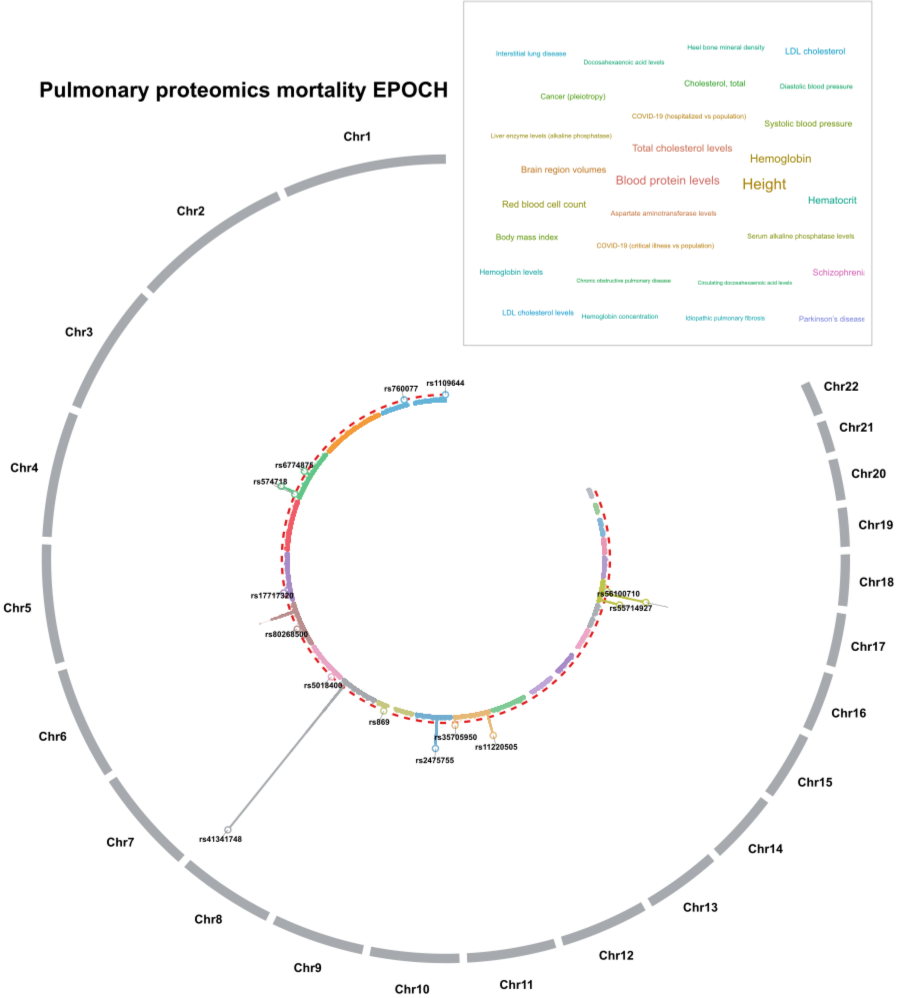


Genome-wide association results from fastGWA are displayed across chromosomes 1-22, with variants positioned according to genomic location and radial distance representing ($-\log_{10}(P)$). Chromosomes are shown in alternating colors, and red-colored dashed circles indicate the genome-wide significance threshold (P-value<$5 \times 10^{-8}$). The locus-level top SNP from each FUMA-defined genomic risk locus is highlighted and labeled using the SNP ID. The word cloud summarizes GWAS Catalog phenotypic categories previously associated with these loci, with text size proportional to the number of distinct loci linked to each category.

**Figure 9: Circular Manhattan plot of phenome-wide associations in GWAS Catalog for the pancreas MRI mortality EPOCH clock**

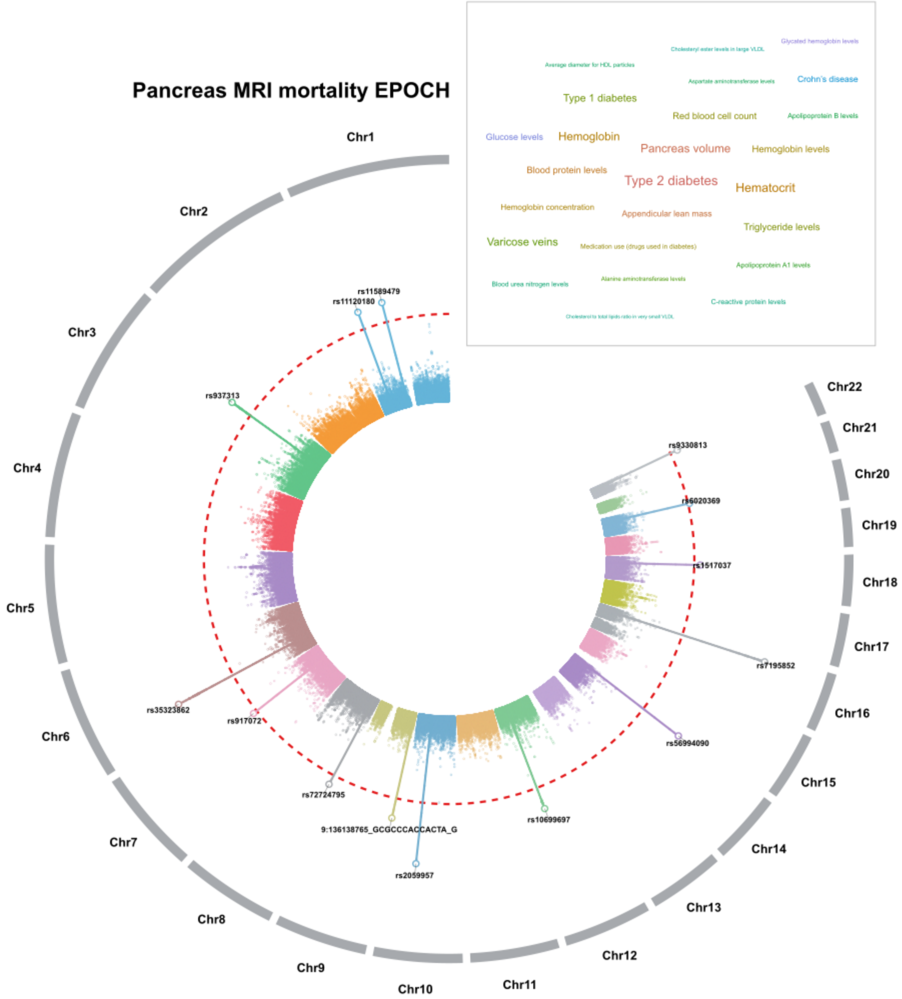


Genome-wide association results from fastGWA are displayed across chromosomes 1-22, with variants positioned according to genomic location and radial distance representing ($-\log_{10}(P)$). Chromosomes are shown in alternating colors, and red-colored dashed circles indicate the genome-wide significance threshold (P-value<$5 \times 10^{-8}$). The locus-level top SNP from each FUMA-defined genomic risk locus is highlighted and labeled using the SNP ID. The word cloud summarizes GWAS Catalog phenotypic categories previously associated with these loci, with text size proportional to the number of distinct loci linked to each category.

**Figure 10: Circular Manhattan plot of phenome-wide associations in GWAS Catalog for the liver MRI mortality EPOCH clock**

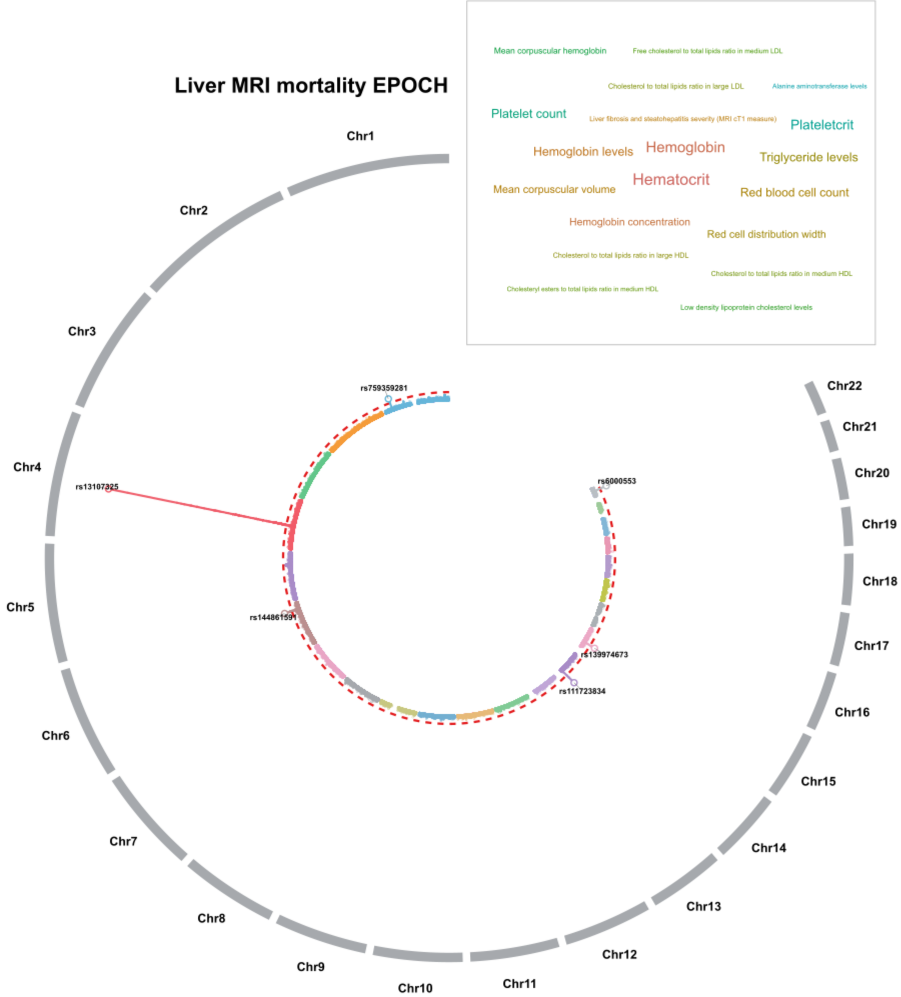


Genome-wide association results from fastGWA are displayed across chromosomes 1-22, with variants positioned according to genomic location and radial distance representing ($-\log_{10}(P)$). Chromosomes are shown in alternating colors, and red-colored dashed circles indicate the genome-wide significance threshold (P-value<$5 \times 10^{-8}$). The locus-level top SNP from each FUMA-defined genomic risk locus is highlighted and labeled using the SNP ID. The word cloud summarizes GWAS Catalog phenotypic categories previously associated with these loci, with text size proportional to the number of distinct loci linked to each category.

**Figure 11: Circular Manhattan plot of phenome-wide associations in GWAS Catalog for the kidney MRI mortality EPOCH clock**

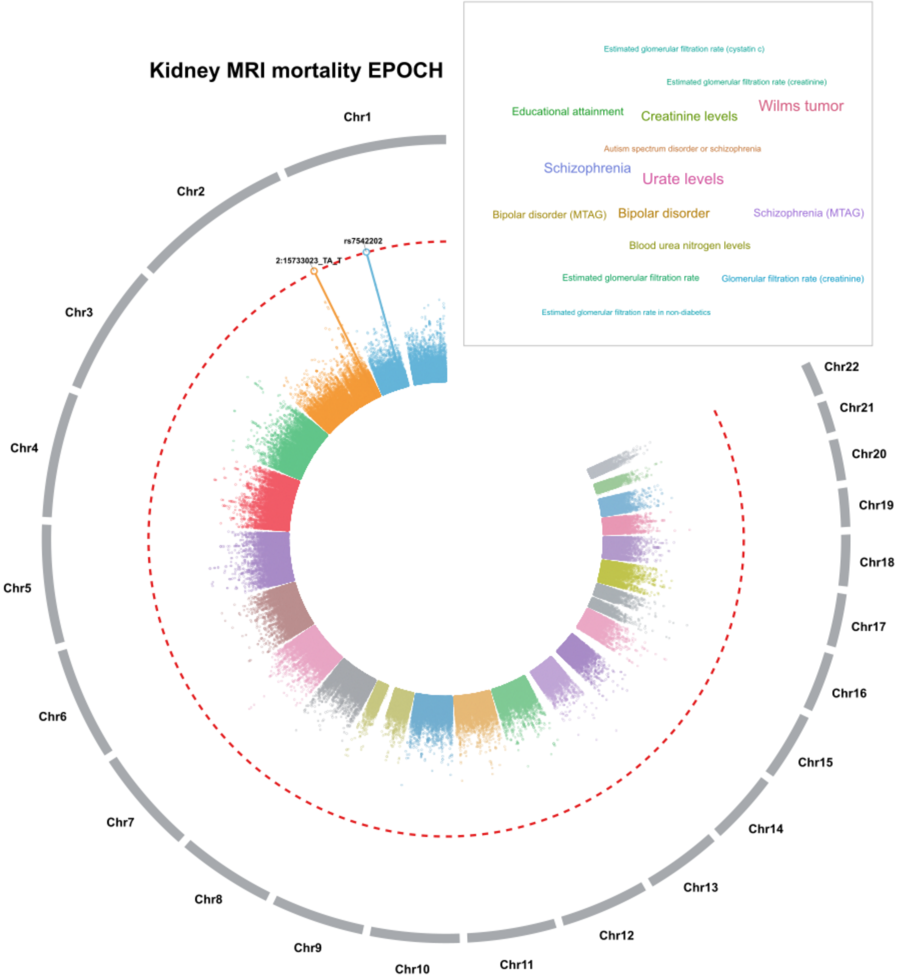


Genome-wide association results from fastGWA are displayed across chromosomes 1-22, with variants positioned according to genomic location and radial distance representing ($-\log_{10}(P)$). Chromosomes are shown in alternating colors, and red-colored dashed circles indicate the genome-wide significance threshold (P-value<$5\times10^{-8}$). The locus-level top SNP from each FUMA-defined genomic risk locus is highlighted and labeled using the SNP ID. The word cloud summarizes GWAS Catalog phenotypic categories previously associated with these loci, with text size proportional to the number of distinct loci linked to each category.

**Figure 12: Circular Manhattan plot of phenome-wide associations in GWAS Catalog for the immune proteomics mortality EPOCH clock**

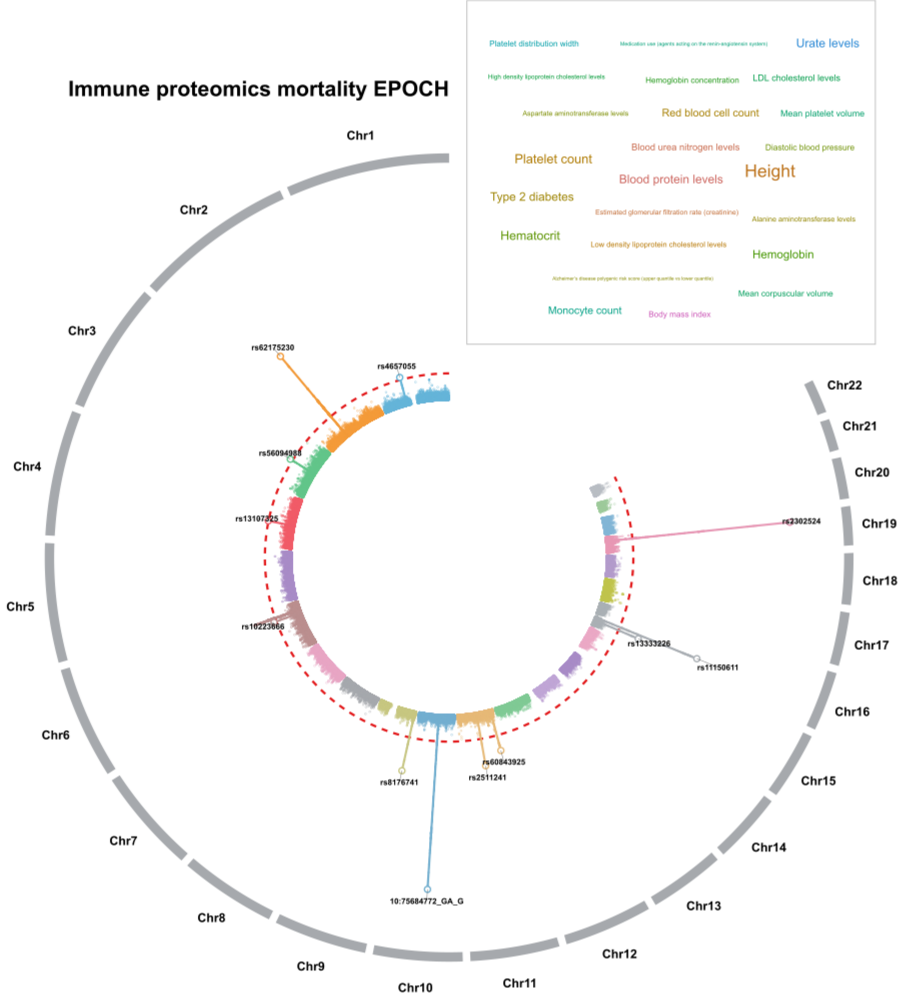


Genome-wide association results from fastGWA are displayed across chromosomes 1-22, with variants positioned according to genomic location and radial distance representing ($-\log_{10}(P)$). Chromosomes are shown in alternating colors, and red-colored dashed circles indicate the genome-wide significance threshold (P-value$<5\times10^{-8}$). The locus-level top SNP from each FUMA-defined genomic risk locus is highlighted and labeled using the SNP ID. The word cloud summarizes GWAS Catalog phenotypic categories previously associated with these loci, with text size proportional to the number of distinct loci linked to each category.

**Figure 13: Circular Manhattan plot of phenome-wide associations in GWAS Catalog for the immune metabolomics mortality EPOCH clock**

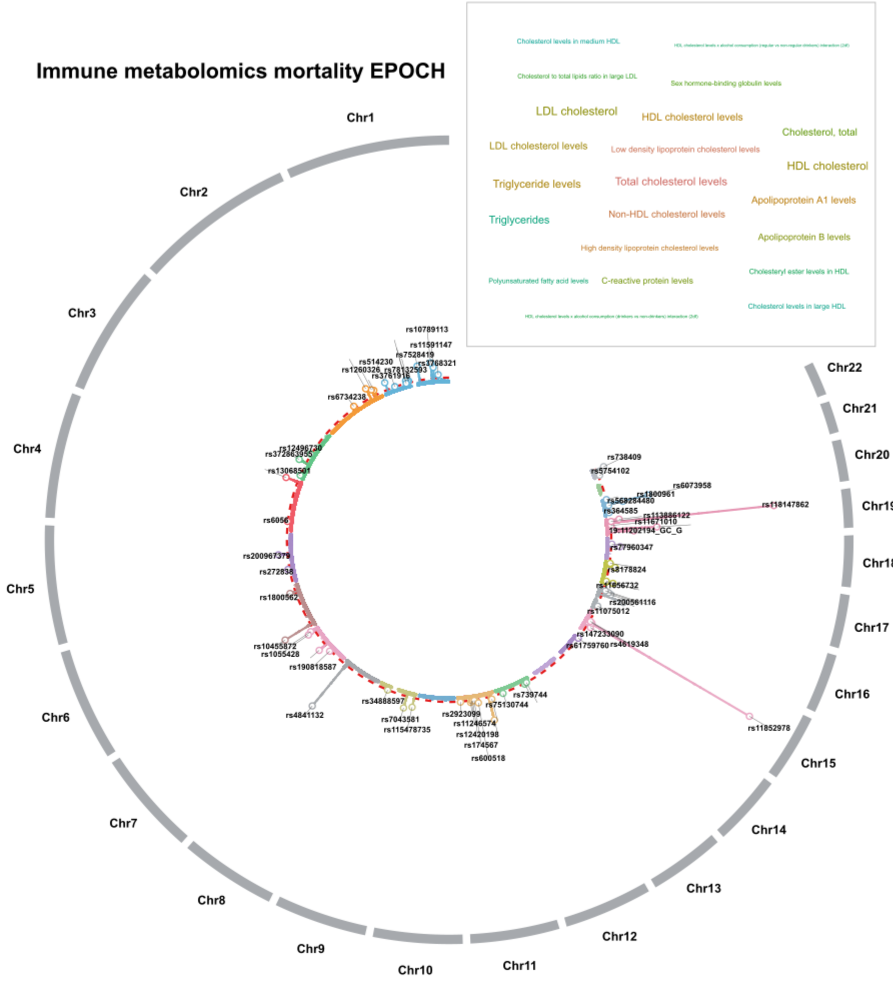


Genome-wide association results from fastGWA are displayed across chromosomes 1-22, with variants positioned according to genomic location and radial distance representing ($-\log_{10}(P)$). Chromosomes are shown in alternating colors, and red-colored dashed circles indicate the genome-wide significance threshold (P-value<$5\times10^{-8}$). The locus-level top SNP from each FUMA-defined genomic risk locus is highlighted and labeled using the SNP ID. The word cloud summarizes GWAS Catalog phenotypic categories previously associated with these loci, with text size proportional to the number of distinct loci linked to each category.

**Figure 14: Circular Manhattan plot of phenome-wide associations in GWAS Catalog for the hepatic proteomics mortality EPOCH clock**

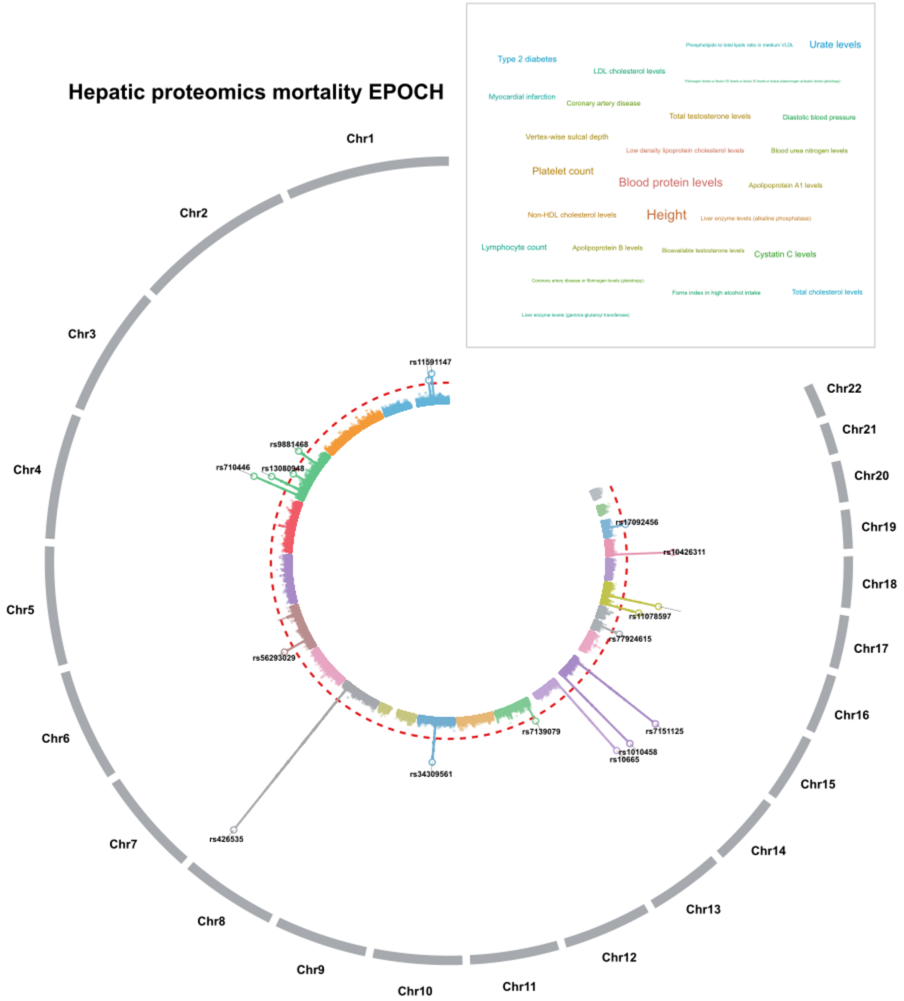


Genome-wide association results from fastGWA are displayed across chromosomes 1-22, with variants positioned according to genomic location and radial distance representing ($-\log_{10}(P)$). Chromosomes are shown in alternating colors, and red-colored dashed circles indicate the genome-wide significance threshold (P-value$<5\text{x}10^{-8}$). The locus-level top SNP from each FUMA-defined genomic risk locus is highlighted and labeled using the SNP ID. The word cloud summarizes GWAS Catalog phenotypic categories previously associated with these loci, with text size proportional to the number of distinct loci linked to each category.

**Figure 15: Circular Manhattan plot of phenome-wide associations in GWAS Catalog for the hepatic metabolomics mortality EPOCH clock**

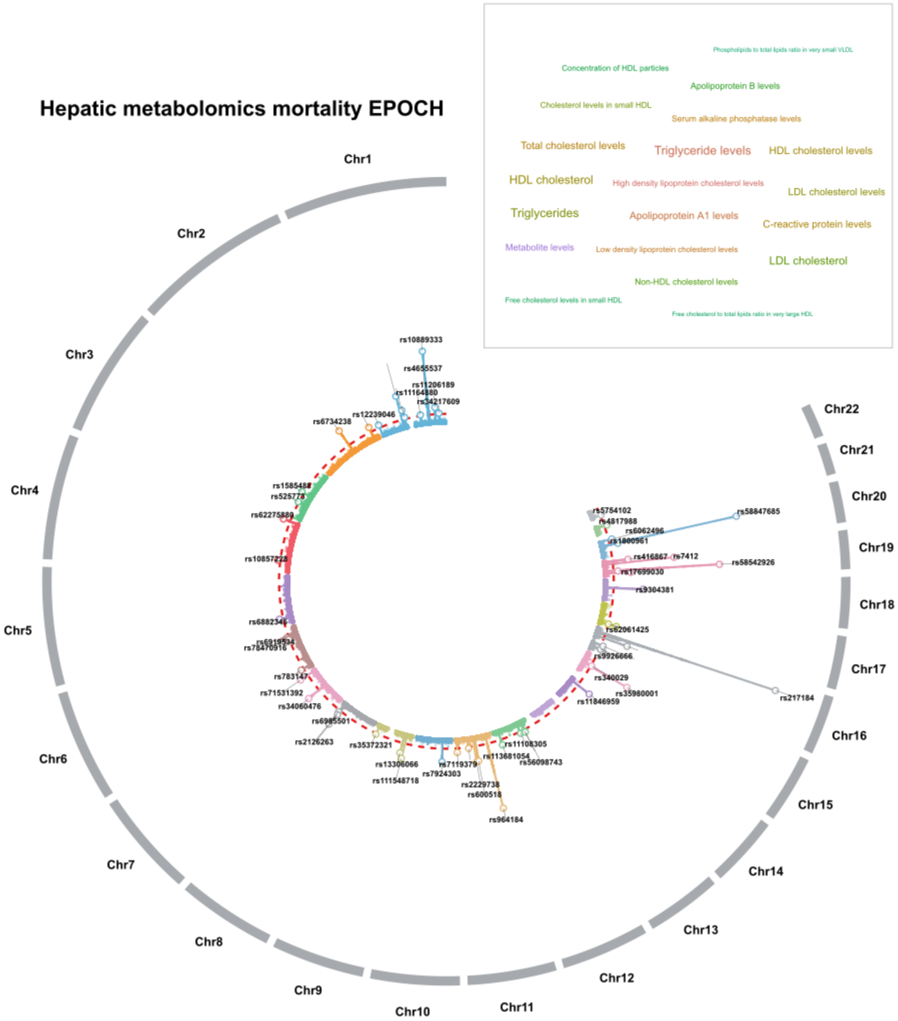


Genome-wide association results from fastGWA are displayed across chromosomes 1-22, with variants positioned according to genomic location and radial distance representing ($-\log_{10}(P)$). Chromosomes are shown in alternating colors, and red-colored dashed circles indicate the genome-wide significance threshold (P-value<$5\times10^{-8}$). The locus-level top SNP from each FUMA-defined genomic risk locus is highlighted and labeled using the SNP ID. The word cloud summarizes GWAS Catalog phenotypic categories previously associated with these loci, with text size proportional to the number of distinct loci linked to each category.

**Figure 16: Circular Manhattan plot of phenome-wide associations in GWAS Catalog for the heart proteomics mortality EPOCH clock**

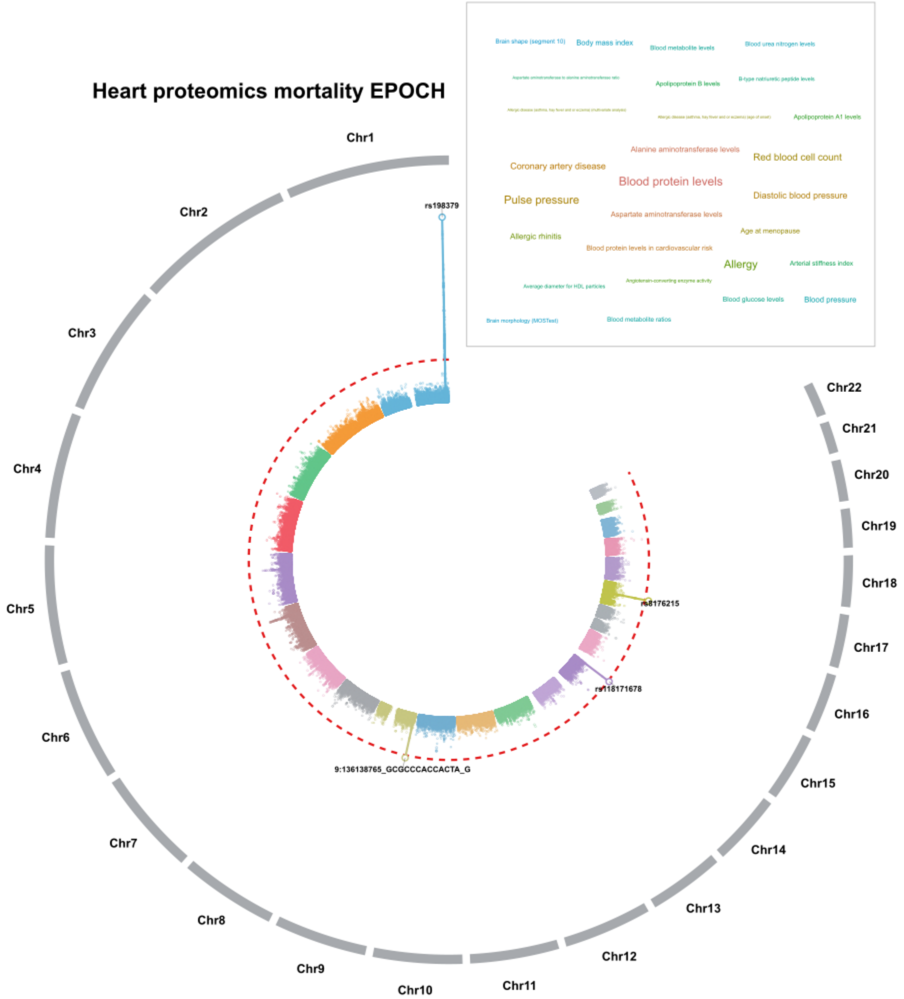


Genome-wide association results from fastGWA are displayed across chromosomes 1-22, with variants positioned according to genomic location and radial distance representing ($-\log_{10}(P)$). Chromosomes are shown in alternating colors, and red-colored dashed circles indicate the genome-wide significance threshold (P-value$<5\times10^{-8}$). The locus-level top SNP from each FUMA-defined genomic risk locus is highlighted and labeled using the SNP ID. The word cloud summarizes GWAS Catalog phenotypic categories previously associated with these loci, with text size proportional to the number of distinct loci linked to each category.

**Figure 17: Circular Manhattan plot of phenome-wide associations in GWAS Catalog for the heart MRI mortality EPOCH clock**

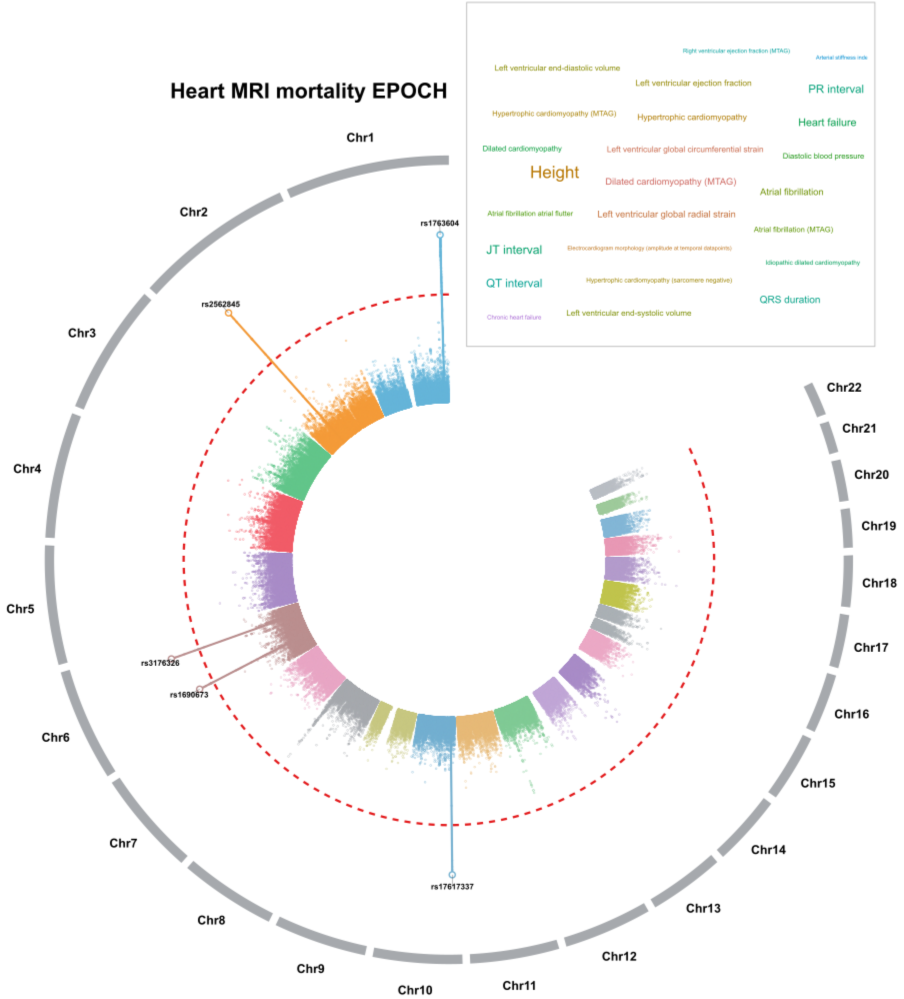


Genome-wide association results from fastGWA are displayed across chromosomes 1-22, with variants positioned according to genomic location and radial distance representing ($-\log_{10}(P)$). Chromosomes are shown in alternating colors, and red-colored dashed circles indicate the genome-wide significance threshold (P-value<$5\times10^{-8}$). The locus-level top SNP from each FUMA-defined genomic risk locus is highlighted and labeled using the SNP ID. The word cloud summarizes GWAS Catalog phenotypic categories previously associated with these loci, with text size proportional to the number of distinct loci linked to each category.

**Figure 18: Circular Manhattan plot of phenome-wide associations in GWAS Catalog for the eye proteomics mortality EPOCH clock**

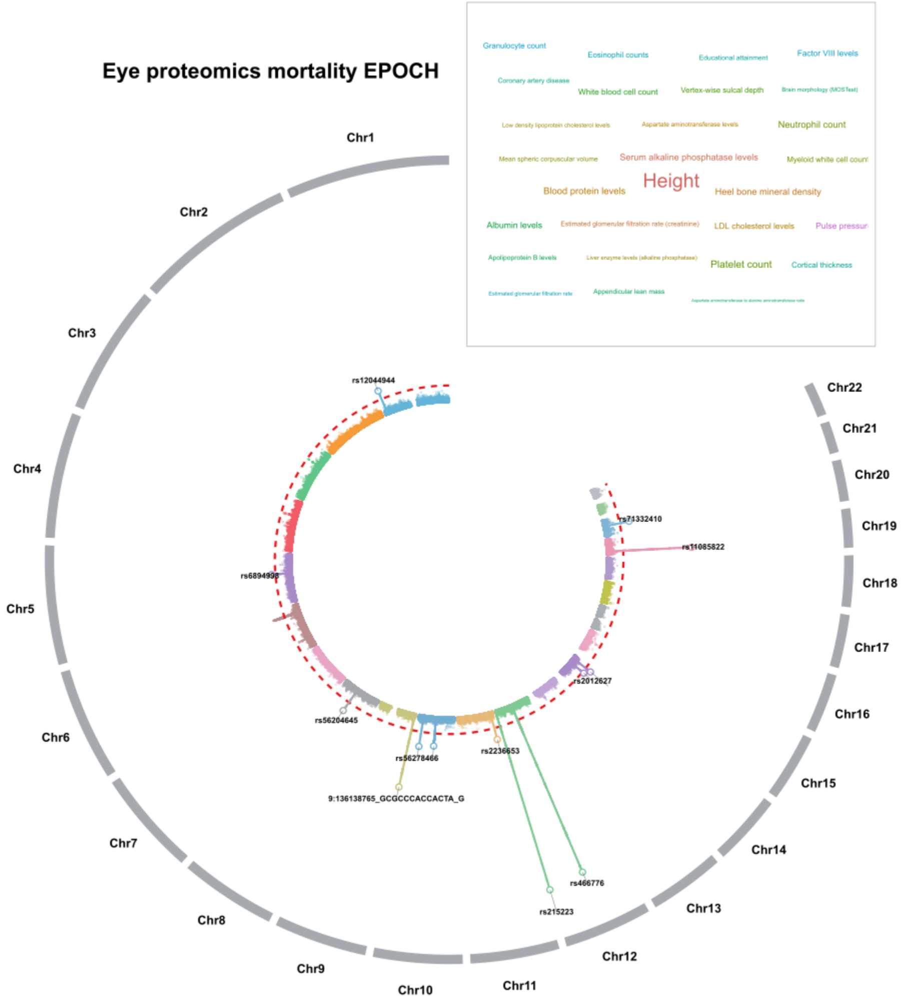


Genome-wide association results from fastGWA are displayed across chromosomes 1-22, with variants positioned according to genomic location and radial distance representing ($-\log_{10}(P)$). Chromosomes are shown in alternating colors, and red-colored dashed circles indicate the genome-wide significance threshold (P-value$<5\text{x}10^{-8}$). The locus-level top SNP from each FUMA-defined genomic risk locus is highlighted and labeled using the SNP ID. The word cloud summarizes GWAS Catalog phenotypic categories previously associated with these loci, with text size proportional to the number of distinct loci linked to each category.

**Figure 19: Circular Manhattan plot of phenome-wide associations in GWAS Catalog for the endocrine proteomics mortality EPOCH clock**

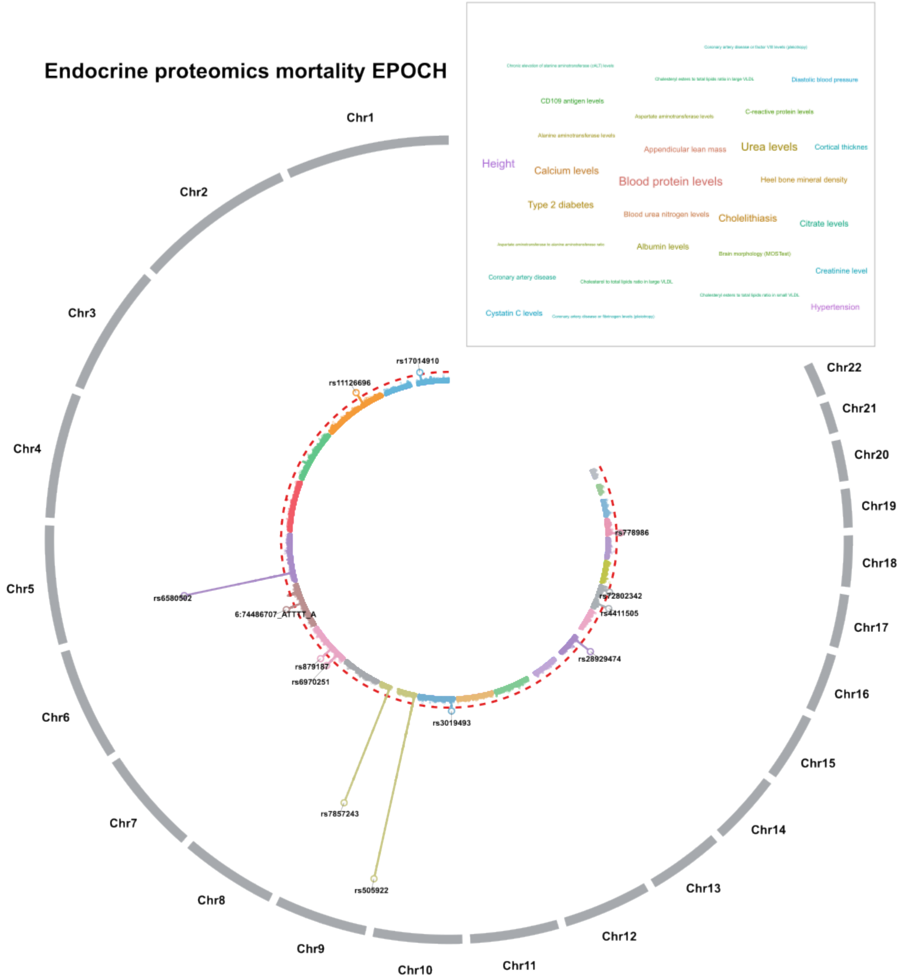


Genome-wide association results from fastGWA are displayed across chromosomes 1-22, with variants positioned according to genomic location and radial distance representing ($-\log_{10}(P)$). Chromosomes are shown in alternating colors, and red-colored dashed circles indicate the genome-wide significance threshold (P-value<$5\times10^{-8}$). The locus-level top SNP from each FUMA-defined genomic risk locus is highlighted and labeled using the SNP ID. The word cloud summarizes GWAS Catalog phenotypic categories previously associated with these loci, with text size proportional to the number of distinct loci linked to each category.

**Figure 20: Circular Manhattan plot of phenome-wide associations in GWAS Catalog for the endocrine metabolomics mortality EPOCH clock**

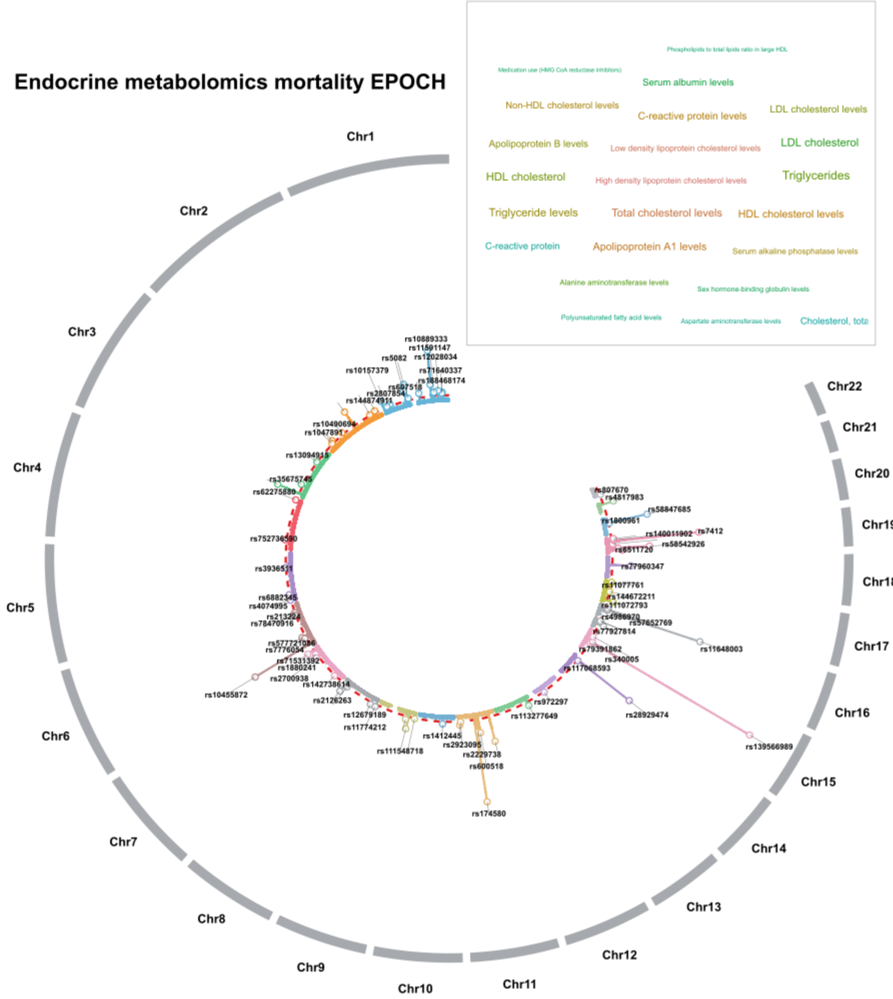


Genome-wide association results from fastGWA are displayed across chromosomes 1-22, with variants positioned according to genomic location and radial distance representing ($-\log_{10}(P)$). Chromosomes are shown in alternating colors, and red-colored dashed circles indicate the genome-wide significance threshold (P-value<$5\times10^{-8}$). The locus-level top SNP from each FUMA-defined genomic risk locus is highlighted and labeled using the SNP ID. The word cloud summarizes GWAS Catalog phenotypic categories previously associated with these loci, with text size proportional to the number of distinct loci linked to each category.

**Figure 21: Circular Manhattan plot of phenome-wide associations in GWAS Catalog for the digestive metabolomic MRI mortality EPOCH clock**

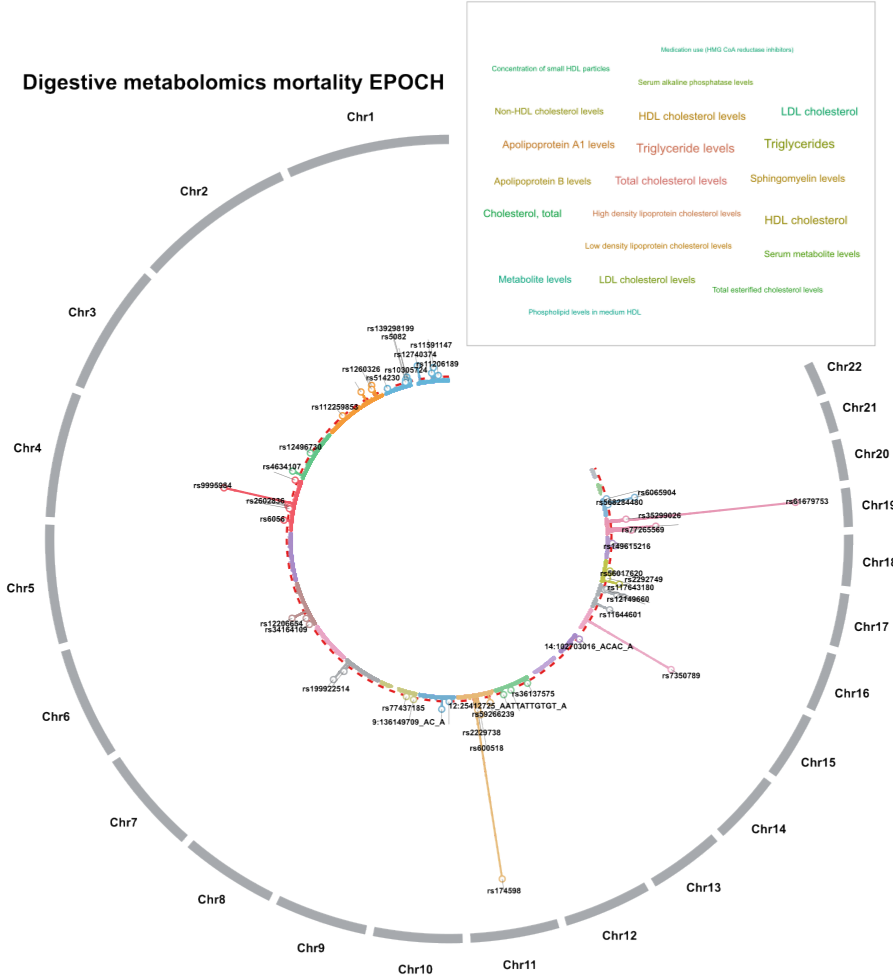


Genome-wide association results from fastGWA are displayed across chromosomes 1-22, with variants positioned according to genomic location and radial distance representing ($-\log_{10}(P)$). Chromosomes are shown in alternating colors, and red-colored dashed circles indicate the genome-wide significance threshold (P-value<$5\times10^{-8}$). The locus-level top SNP from each FUMA-defined genomic risk locus is highlighted and labeled using the SNP ID. The word cloud summarizes GWAS Catalog phenotypic categories previously associated with these loci, with text size proportional to the number of distinct loci linked to each category.

**Figure 22: Circular Manhattan plot of phenome-wide associations in GWAS Catalog for the brain proteomics mortality EPOCH clock**

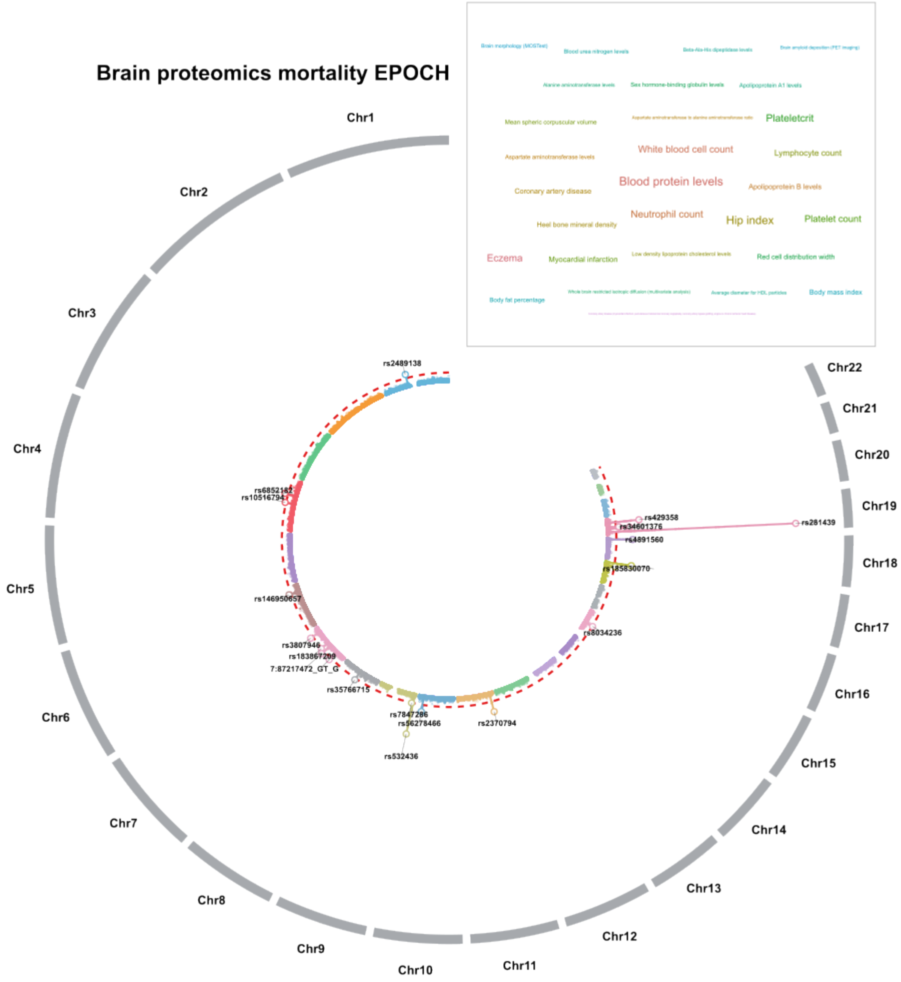


Genome-wide association results from fastGWA are displayed across chromosomes 1-22, with variants positioned according to genomic location and radial distance representing ($-\log_{10}(P)$). Chromosomes are shown in alternating colors, and red-colored dashed circles indicate the genome-wide significance threshold (P-value<$5x10^{-8}$). The locus-level top SNP from each FUMA-defined genomic risk locus is highlighted and labeled using the SNP ID. The word cloud summarizes GWAS Catalog phenotypic categories previously associated with these loci, with text size proportional to the number of distinct loci linked to each category.

**Figure 23: Circular Manhattan plot of phenome-wide associations in GWAS Catalog for the brain MRI mortality EPOCH clock**

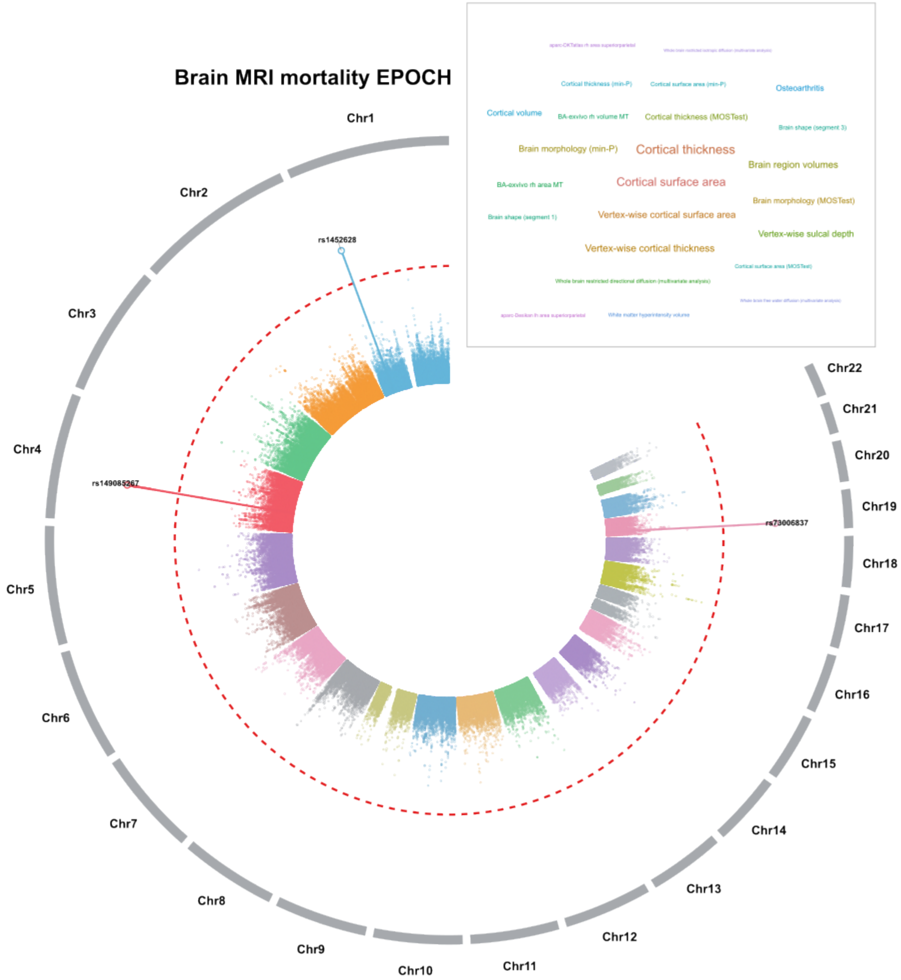


Genome-wide association results from fastGWA are displayed across chromosomes 1-22, with variants positioned according to genomic location and radial distance representing ($-\log_{10}(P)$). Chromosomes are shown in alternating colors, and red-colored dashed circles indicate the genome-wide significance threshold (P-value<$5 \times 10^{-8}$). The locus-level top SNP from each FUMA-defined genomic risk locus is highlighted and labeled using the SNP ID. The word cloud summarizes GWAS Catalog phenotypic categories previously associated with these loci, with text size proportional to the number of distinct loci linked to each category.

**Figure 24: Circular Manhattan plot of phenome-wide associations in GWAS Catalog for the adipose MRI mortality EPOCH clock**

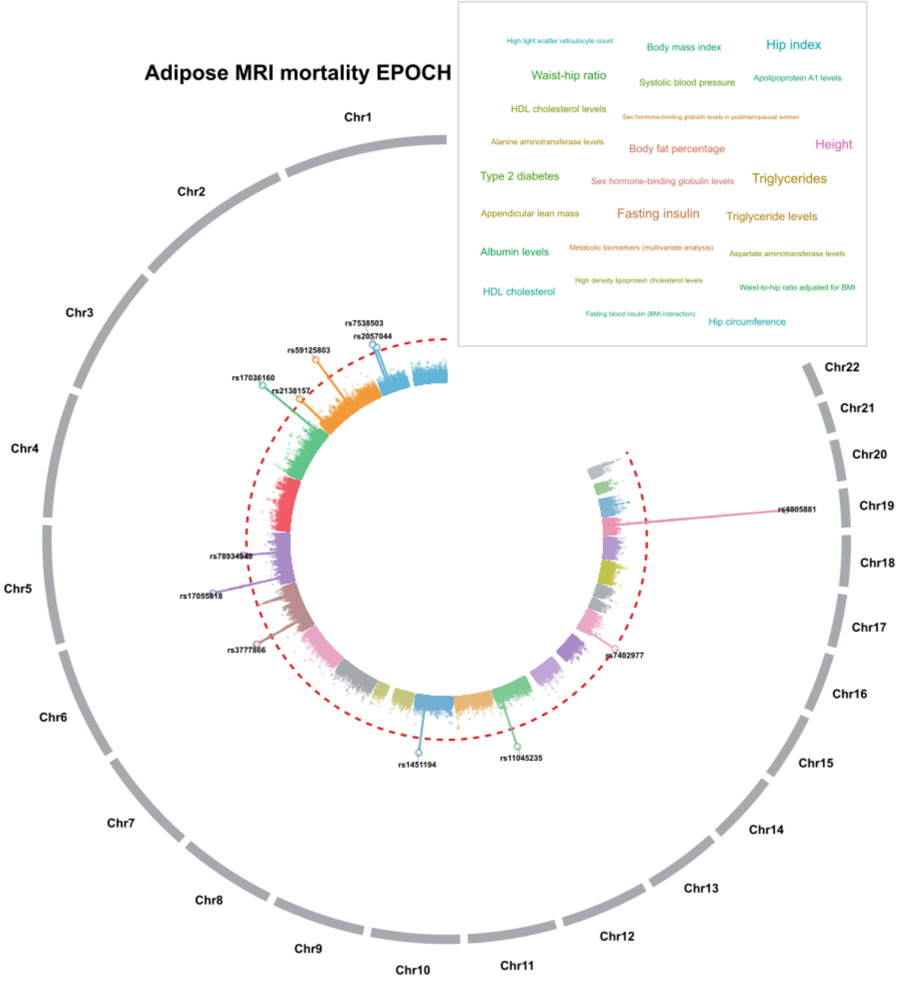


Genome-wide association results from fastGWA are displayed across chromosomes 1-22, with variants positioned according to genomic location and radial distance representing ($-\log_{10}(P)$). Chromosomes are shown in alternating colors, and red-colored dashed circles indicate the genome-wide significance threshold (P-value<$5\times10^{-8}$). The locus-level top SNP from each FUMA-defined genomic risk locus is highlighted and labeled using the SNP ID. The word cloud summarizes GWAS Catalog phenotypic categories previously associated with these loci, with text size proportional to the number of distinct loci linked to each category.

**Figure 25: Genetic architecture of brain proteomics mortality EPOCH converges on four interacting biological systems relevant to lifespan**

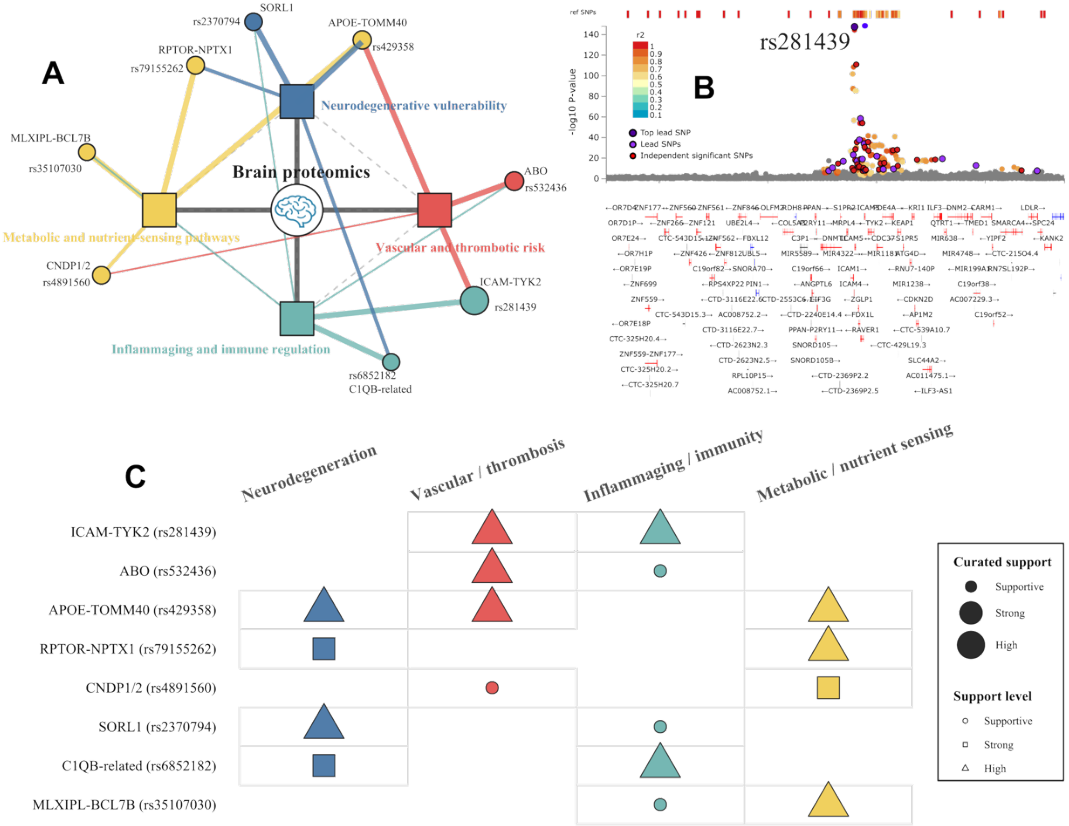


**A**) Conceptual network representation of mortality-relevant genetic pathways. The brain proteomics mortality EPOCH is positioned at the center of 4 interacting biological systems: i) neurodegenerative vulnerability, ii) vascular and thrombotic risk, iii) inflammaging and immune regulation, and iv) metabolic and nutrient-sensing biology. Representative genome-wide significant loci and mapped genes are connected to the systems they support, including the APOE-TOMM40-APOC1 region, ICAM-TYK2, ABO, SORL1, RPTOR-NPTX1, CNDP1/CNDP2, MLXIPL-BCL7B, and C1QB-related signals. Colored lines denote locus-to-system relationships, while dashed lines indicate conceptual interactions among the 4 systems. Where line thickness varies, it reflects curated qualitative support from convergent positional mapping, eQTL evidence, chromatin interactions, and prior GWAS-trait associations. **B**) Regional Manhattan plot for the inflammaging and immunity locus (rs281429) and the mapped genes. **C**) The matrix summarizes the relative support linking each representative locus to the four biological systems. Darker or larger cells indicate stronger curated evidence based on the combined FUMA annotations and prior-trait overlap in GWAS Catalog. Several loci show pleiotropic relationships across systems. For example, APOE links neurodegeneration, lipid metabolism, and vascular risk, whereas ICAM-TYK2 connects immune activation with endothelial and vascular injury. Together, these results illustrate that the genetic basis of the brain proteomics mortality EPOCH reflects a coordinated brain-vascular-immune-metabolic axis that may contribute to differences in lifespan and mortality risk.

**Figure 26: Cross-method consistency of SNP heritability estimates for 22 mortality EPOCH clocks**

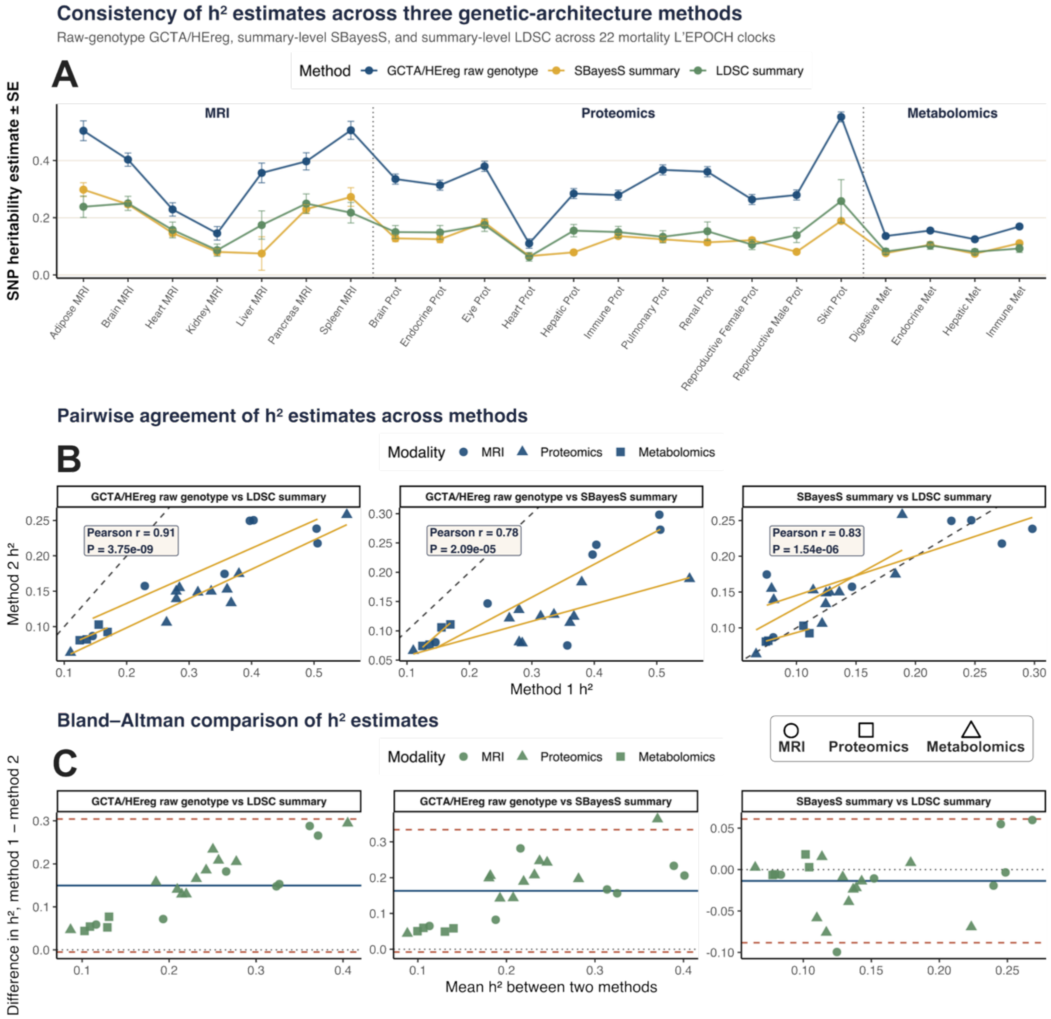


**A**) Heritability estimates across three genetic-architecture methods. Line plot comparing SNP heritability ($h^2$) estimates for 22 mortality EPOCH clocks using three complementary approaches: raw-genotype GCTA/HEreg, summary-level SBayesS, and summary-level LDSC. Clocks are grouped by modality, including MRI-, proteomics-, and metabolomics-derived EPOCH clocks. Points represent $h^2$ estimates, and error bars indicate standard errors. Across modalities, the three methods showed broadly consistent relative patterns, although raw-genotype GCTA/HEreg generally produced higher estimates than summary-level methods. Of note, GCTA-HEreg was used for clocks for which standard GCTA-GREML failed to converge or produced unstable variance-component estimates. This occurred among metabolomics EPOCH clocks, likely reflecting differences in phenotype scaling, distributional properties, or residual variance structure. **B**) Pairwise agreement of heritability estimates across methods. Scatterplots show pairwise correlations of $h^2$estimates between GCTA/HEreg and LDSC, GCTA/HEreg and SBayesS, and SBayesS and LDSC across mortality EPOCH clocks. **C**) Bland-Altman plots assess agreement between each pair of heritability estimation methods by comparing the average $h^2$ estimate against the between-method difference. The solid horizontal line indicates the mean difference, and dashed lines indicate limits of agreement. These analyses highlight systematic differences in scale between raw-genotype and summary-level methods while confirming broadly consistent clock-level heritability patterns across MRI, proteomics, and metabolomics mortality EPOCH clocks.

**Figure 27: Comparisons between the 22 mortality EPOCH clocks and their corresponding BAGs**

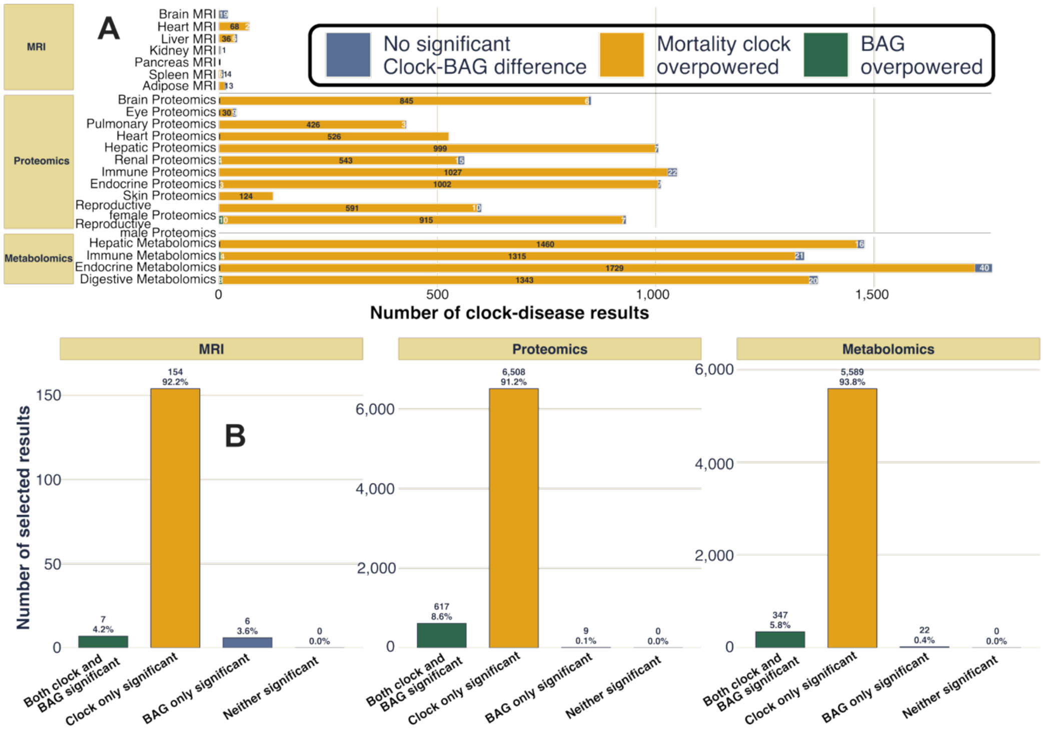


**A**) For all EPOCH-BAG pairs, stacked bars are stratified by whether there was no significant EPOCH-BAG difference, a stronger mortality EPOCH effect, or a stronger BAG effect. **B**) For all EPOCH-BAG pairs, bars show the number and percentage of selected results in each modality classified as both EPOCH and BAG significant, EPOCH only significant, BAG only significant, or neither significant. This highlights that most selected associations were driven by mortality EPOCH-only significance, particularly among proteomic and metabolomic clocks.

**Figure 28: The 22 mortality EPOCH clocks predict future disease onset in the holdout test dataset**

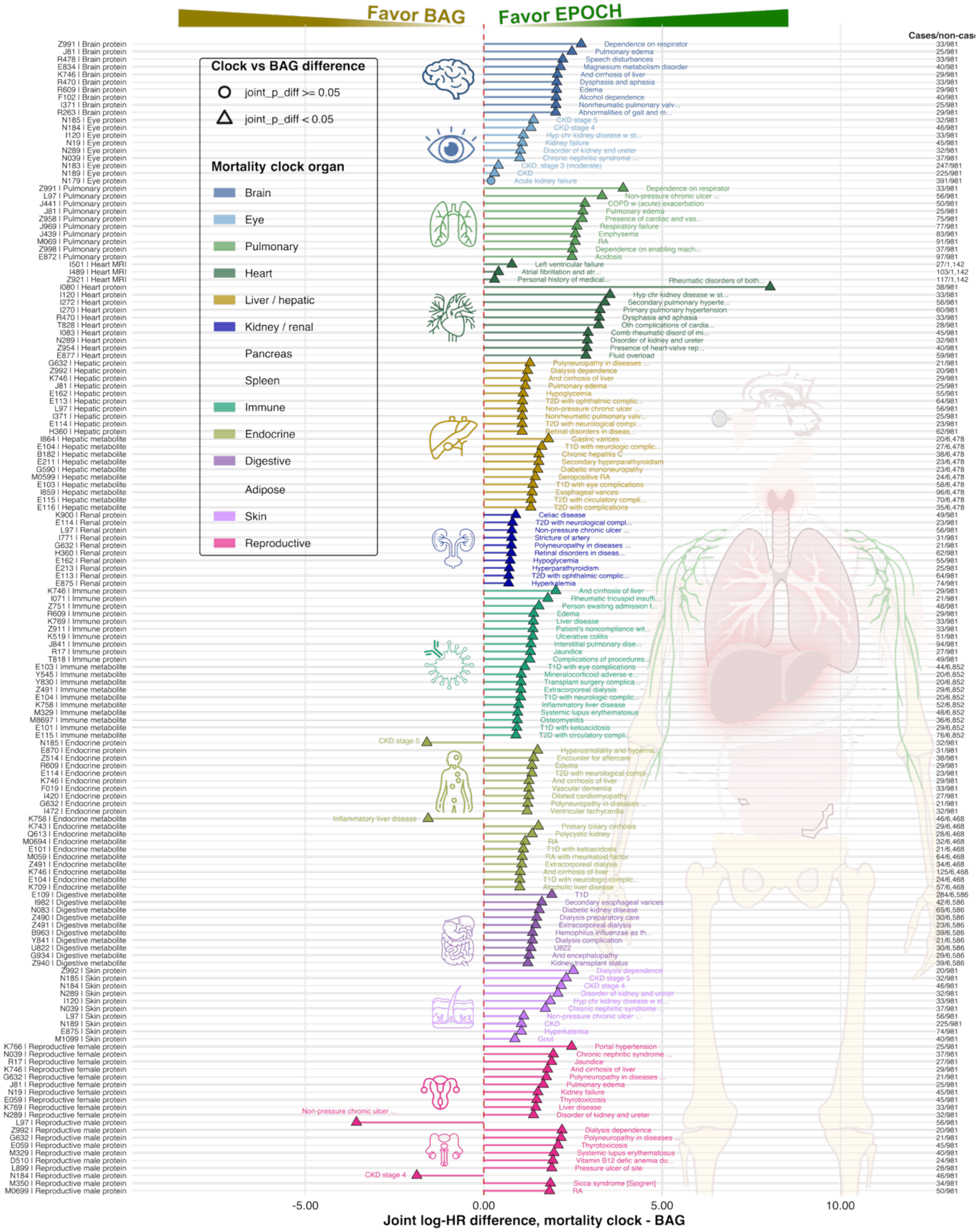


Mortality EPOCH retains superior disease-onset associations over matched BAGs in the holdout test participants. Forest plots show the top 10 disease endpoints per mortality clock, ranked by the absolute difference between the joint Cox regression coefficients for mortality EPOCH acceleration and its matched biological age gap (BAG). Analyses were restricted to each clock's original held-out mortality-model test set and used disease-free controls, resulting in substantially smaller numbers of incident cases than in the full-sample analysis. The joint log-HR values greater than 1 indicate a stronger conditional association for EPOCH, whereas values below 1 indicate a stronger association for BAG. Triangles denote nominally significant differences between EPOCH and BAG in the joint model ($P<0.05$). Despite the approximately five-fold reduction in sample size and corresponding loss of statistical power, the predominance of EPOCH over matched BAGs persisted across organ systems and molecular modalities, supporting the robustness of EPOCH's clinically anchored disease-risk information beyond the mortality-clock development cohort. Cases and disease-free non-cases included in each comparison are shown at right.

**Figure 29: Comparison of brain proteomic EPOCH with its constituent proteins for Alzheimer's disease onset**

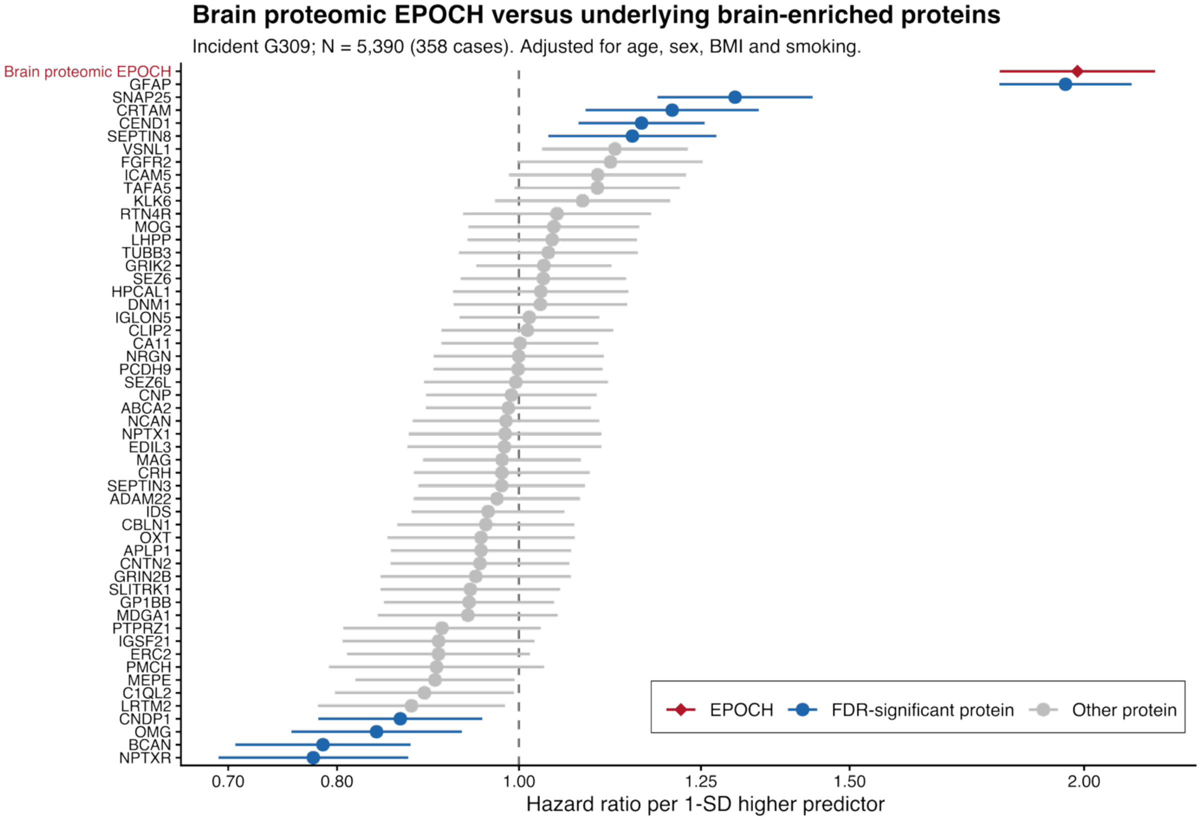


Forest plot comparing the brain proteomic mortality EPOCH with each of its 53 underlying brain-enriched proteins for prediction of incident Alzheimer's disease (G309). Hazard ratios (HRs) and 95% confidence intervals are shown per 1-SD higher predictor from separate Cox proportional-hazards models adjusted for age, sex, BMI, and smoking status, using the same analysis sample (*N*=5,390; 358 incident cases). The brain proteomic EPOCH showed a strong association with subsequent Alzheimer's disease onset (HR = 1.98, 95% CI: 1.80-2.18), exceeding most individual constituent proteins, with GFAP showing a comparable effect. EPOCH is shown in red, proteins passing Benjamini-Hochberg correction across the 53 proteins (FDR < 0.05) in blue, and remaining proteins in gray. The dashed vertical line denotes HR=1.

**Figure 30: Brain proteomics mortality EPOCH provides incremental prediction of Alzheimer's disease and heart failure beyond conventional clinical biomarkers**

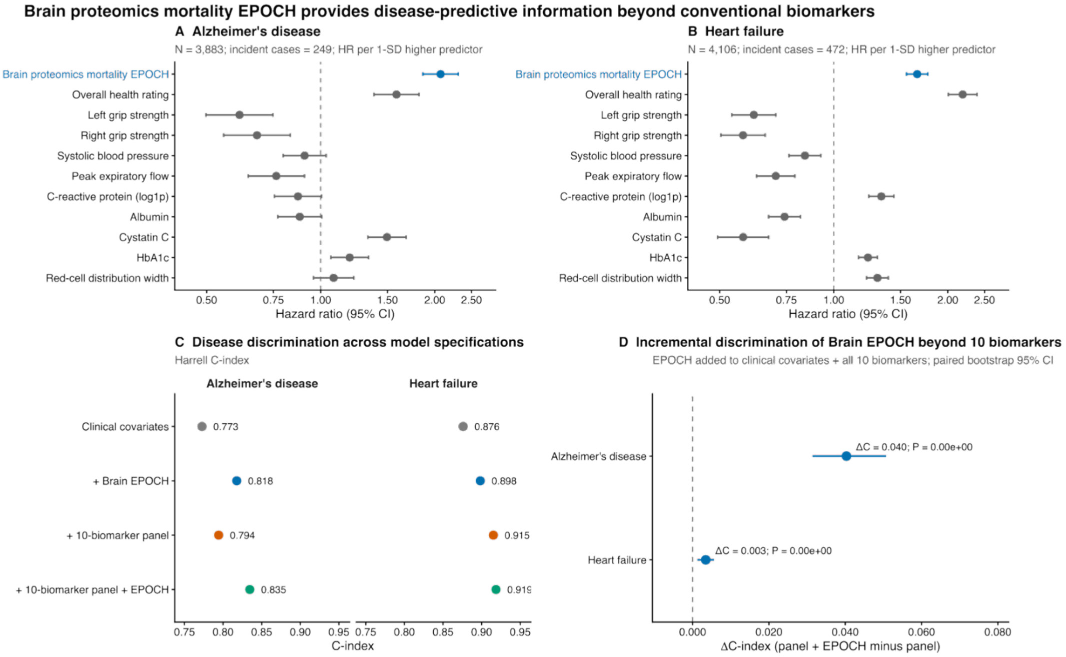


**A**) Associations of the brain proteomics mortality EPOCH and 10 conventional biomarkers with incident Alzheimer's disease. Points indicate hazard ratios (HRs) per 1-SD higher predictor and horizontal lines indicate 95% confidence intervals from Cox proportional hazards models. The vertical dashed line denotes HR = 1. **B**) Corresponding associations with incident heart failure using the same modeling strategy and common-sample design. **C**) Discrimination of multivariable models for Alzheimer's disease and heart failure, quantified using Harrell's C-index. Models include baseline clinical covariates alone, baseline covariates plus the brain proteomics mortality EPOCH, baseline covariates plus the combined 10-biomarker panel, and baseline covariates plus both the 10-biomarker panel and the brain EPOCH. **D**) Incremental discrimination gained by adding the brain proteomics mortality EPOCH to the complete 10-biomarker model. Points represent the paired difference in C-index (ΔC) between the model containing the 10 conventional biomarkers plus EPOCH and the model containing the 10 biomarkers alone; horizontal lines indicate 95% confidence intervals derived from participant-level paired bootstrap resampling. The vertical dashed line denotes no incremental improvement (ΔC=0). All analyses used an identical complete-case population within each disease to ensure direct comparison among predictors and models.

**Figure 31: Brain MRI mortality EPOCH predicts incident mental and nervous-system disorders (ICD code: G and F) beyond disease-specific AI-derived brain MRI subtypes across survival and fixed-horizon analyses**

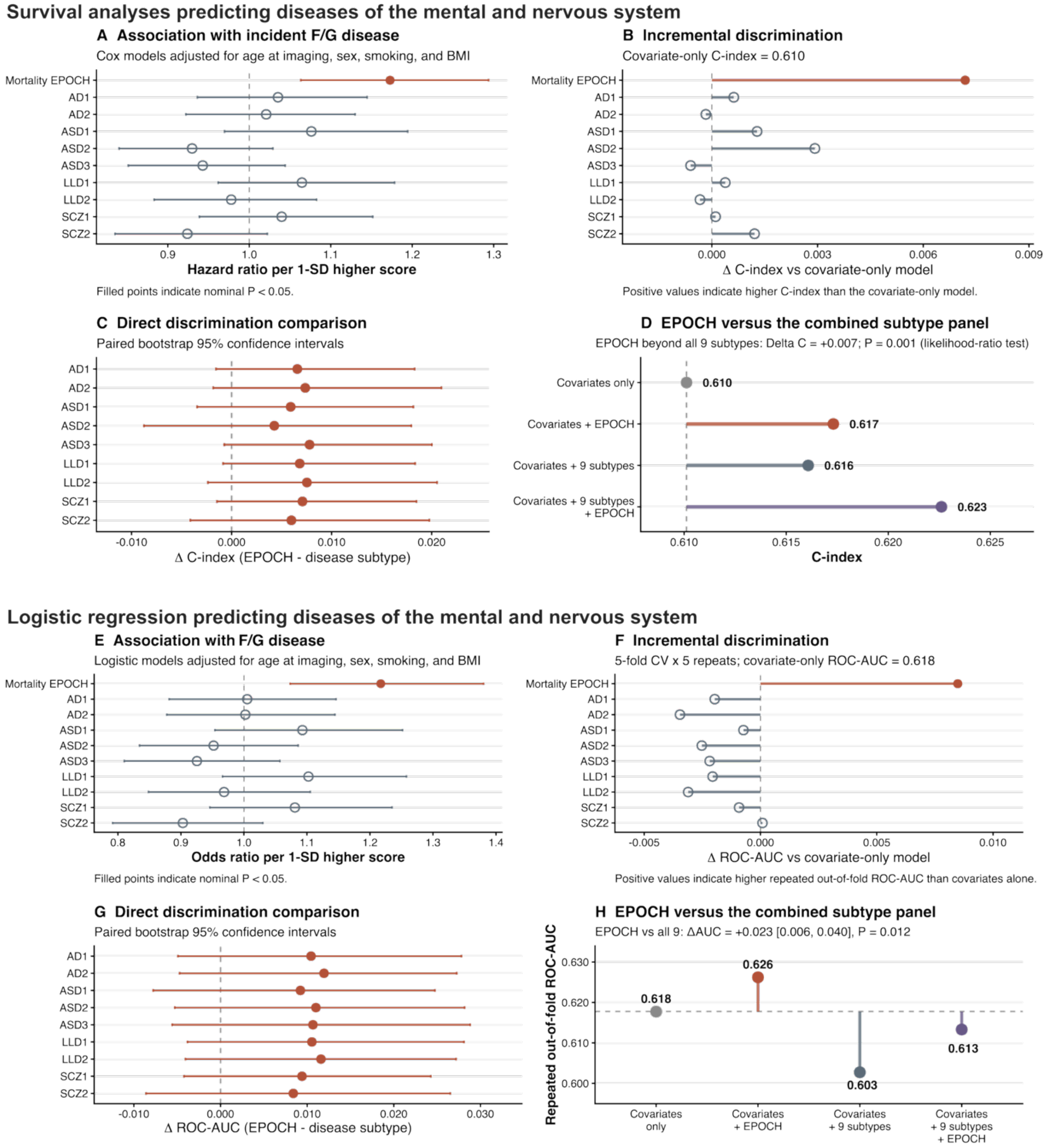


All analyses were performed in participants from the held-out EPOCH test set and restricted to common complete-case samples to ensure identical comparisons across biomarkers. Incident disease was defined as the first recorded ICD-10 diagnosis within the mental and behavioral disorders (F*) or nervous-system disorders (G*) chapters after brain MRI. The 9 disease-specific AI biomarkers comprised AD1-2, ASD1-3, LLD1-2, and SCZ1-2. **A**) Associations of the brain MRI mortality EPOCH and individual disease-subtype scores with incident F/G disease, adjusted for age at imaging, sex, smoking, and BMI. Points indicate hazard ratios per 1-SD higher score and horizontal lines indicate 95% confidence intervals; filled points denote nominal P<0.05. In the common sample (N=6,327; 381 events), mortality EPOCH was significantly associated with subsequent F/G disease (HR=1.17, 95% CI 1.06-1.29, P=0.0015), whereas none of the individual disease-subtype scores reached significance. **B**) Incremental discrimination relative to the covariate-only Cox model, denoted by ΔC-index. **C**) Direct paired-bootstrap comparison of discrimination between EPOCH and each disease subtype, expressed as ΔC-index (EPOCH minus subtype) with 95% bootstrap confidence intervals; positive values favor EPOCH, although the individual pairwise confidence intervals included zero. **D**) Comparison of the covariate-only model, covariates plus EPOCH, covariates plus all nine disease subtypes, and covariates plus all nine subtypes and EPOCH. EPOCH provided significant incremental model information beyond the combined nine-subtype panel (likelihood-ratio P=0.0013). **E**) Complementary logistic regression analyses were designed to evaluate whether the predictive signal of EPOCH persisted when disease status was modeled independently of a survival-analysis framework (as by EPOCH). In the common sample (N=6,296; 235 cases), mortality EPOCH was significantly associated with the disease status (OR=1.22, 95% CI 1.07-1.38, P=0.0022), whereas none of the nine subtype scores was individually significant. **F**) Incremental repeated out-of-fold ROC-AUC relative to the covariate-only logistic model, estimated using repeated five-fold cross-validation. EPOCH achieved the highest individual adjusted discrimination (ROC-AUC=0.626 versus 0.618 for covariates alone). **G**) Paired-bootstrap differences in cross-validated ROC-AUC between EPOCH and each disease subtype; positive values favor EPOCH. **H**) Cross-validated discrimination of the covariate-only, EPOCH, combined nine-subtype, and nine-subtype-plus-EPOCH models. EPOCH showed higher discrimination than the combined nine-subtype panel (ΔROC-AUC=0.023, 95% CI 0.006-0.040, bootstrap P=0.012) and remained independently informative after adjustment for all nine subtype scores (likelihood-ratio P=0.0013).

**Figure 32: Medication history association with longitudinal changes in metabolomics mortality EPOCH clocks**

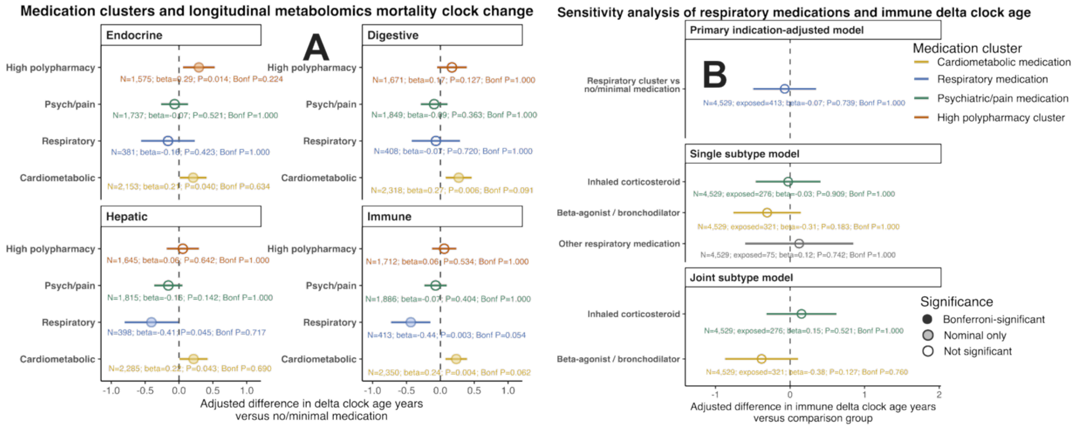


**A**) Forest plots show associations between baseline medication-history categories, compared to non-medication history participants, and longitudinal changes (instance 1 – instance 0) in four metabolomics mortality EPOCH clocks, including endocrine, digestive, hepatic, and immune metabolomics clocks. Points represent estimated effect sizes, and horizontal bars indicate confidence intervals. Medication categories include high polypharmacy, psychotropic, respiratory, and cardiometabolic medication use. Although several medication classes showed nominal associations with changes in metabolomics mortality EPOCH clocks, none survived correction for multiple comparisons. **B**). Respiratory-medication history was further decomposed into medication subtypes using primary indication-adjusted, single-subtype, and joint-subtype models. These analyses evaluated whether inhaled corticosteroids, beta-agonists/bronchodilators, or other respiratory medications were independently associated with longitudinal changes in metabolomics mortality EPOCH clocks. Nominal associations observed in the primary models were attenuated after accounting for additional covariates and respiratory medication subtypes, and no association survived multiple-comparisons correction. Together, these sensitivity analyses indicate that medication-history signals are weak and likely reflect confounding by indication, disease burden, or correlated medication use rather than robust medication-specific effects on metabolomics mortality EPOCH trajectories.

**Figure 33: Model performance of asthma EPOCH clocks across organs and modalities**

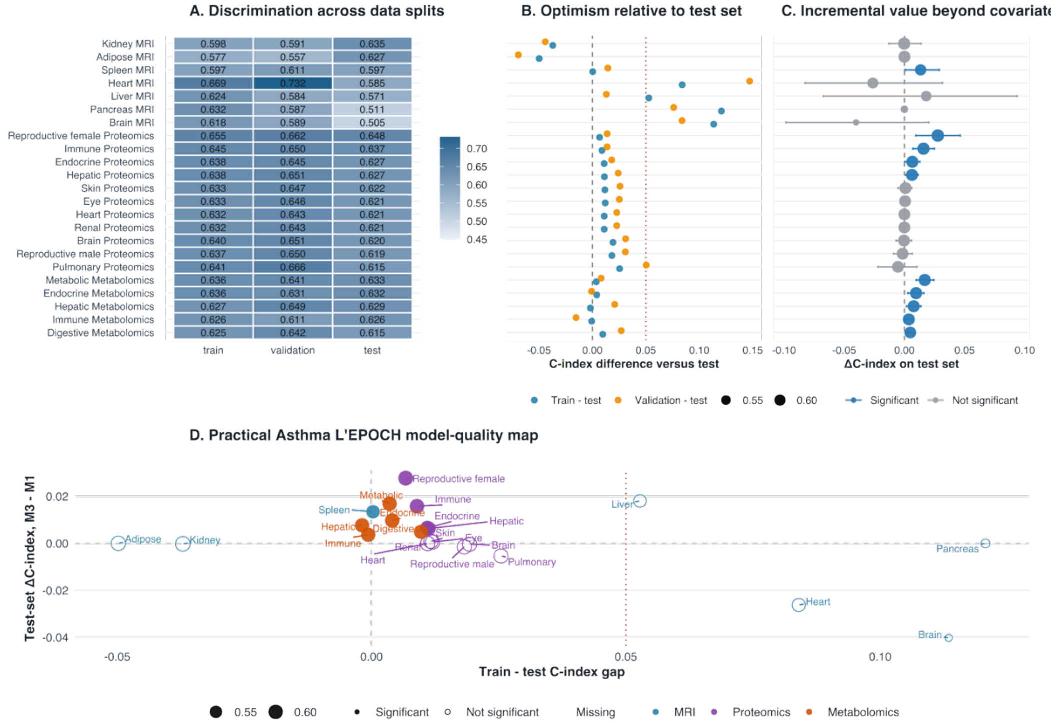


**A**) Heatmap showing C-index values for asthma EPOCH clocks across the training, validation, and held-out test datasets. Rows represent organ-specific asthma EPOCH clocks, including MRI-, proteomics-, and metabolomics-derived models. Overall, most clocks showed moderate discrimination, with most clocks maintaining comparable performance across data splits, supporting reproducible out-of-sample prediction of future asthma risk. **B**) Dot plot showing the difference in C-index between the training or validation datasets and the held-out test dataset. Values near zero indicate limited optimism and stable generalization, whereas positive values indicate higher apparent performance in training or validation than in the test set. **C**) Forest plot showing the change in test-set C-index after adding each asthma EPOCH clock to the covariate-only model. Points indicate ΔC-index estimates, and horizontal bars indicate uncertainty intervals. Blue points denote models with statistically significant incremental value, while gray points indicate non-significant improvements. **D**) Scatterplot summarizing model quality by jointly comparing generalization and incremental value. The x-axis shows the train-test C-index gap, with values closer to zero indicating better generalization, while the y-axis shows the test-set ΔC-index beyond covariates, with positive values indicating added predictive utility. Points are colored by modality and labeled by organ/system. Filled circle points denote the clocks that were selected for downstream analyses.

**Figure 34: Model performance of COPD EPOCH clocks across organs and modalities**

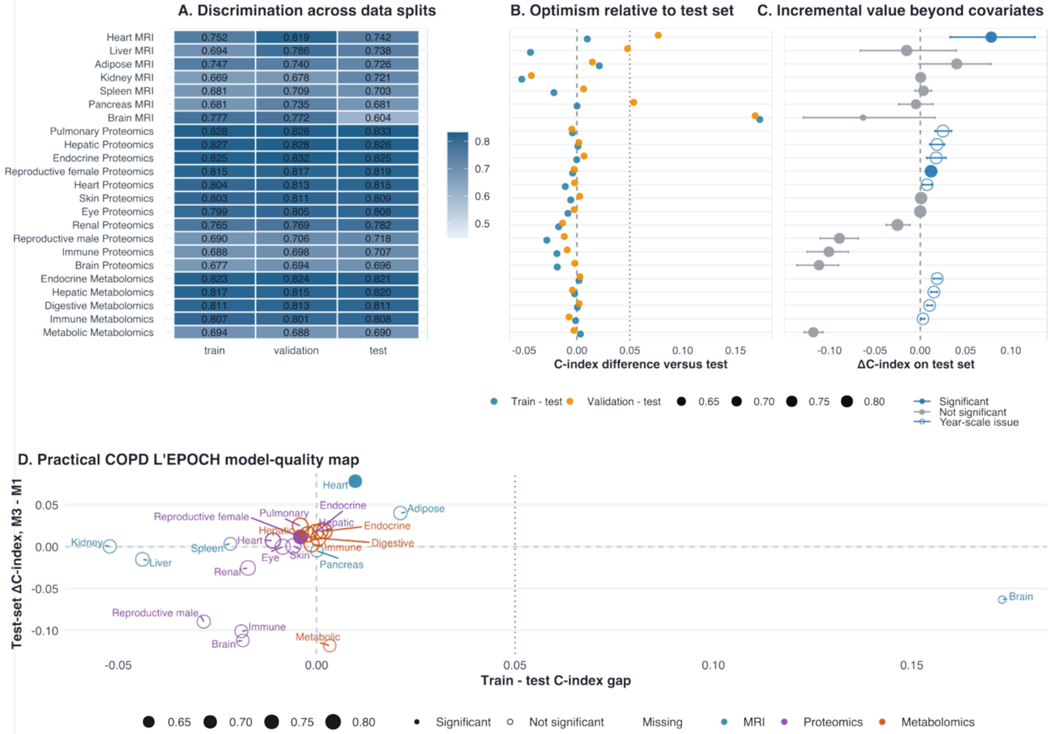


**A**) Heatmap showing C-index values for COPD EPOCH clocks across the training, validation, and held-out test datasets. Rows represent organ-specific COPD EPOCH clocks, including MRI-, proteomics-, and metabolomics-derived models. **B**) Dot plot showing the difference in C-index between the training or validation datasets and the held-out test dataset. Values near zero indicate limited optimism and stable generalization, whereas positive values indicate higher apparent performance in training or validation than in the test set. **C**) Forest plot showing the change in test-set C-index after adding each COPD EPOCH clock to the covariate-only model. Points indicate ΔC-index estimates, and horizontal bars indicate uncertainty intervals. Filled blue points denote models with statistically significant incremental value, whereas gray points indicate non-significant improvements. Open blue points denote models with statistically significant incremental value but clear year-scaling issues, as shown in **Supplementary Figure 1**. **D**) Scatterplot summarizing model quality by jointly comparing generalization and incremental value. The x-axis shows the train-test C-index gap, with values closer to zero indicating better generalization, while the y-axis shows the test-set ΔC-index beyond covariates, with positive values indicating added predictive utility. Points are colored by modality and labeled by organ/system. Filled circle points denote the clocks that were selected for downstream analyses.

**Figure 35: Model performance of dementia EPOCH clocks across organs and modalities**

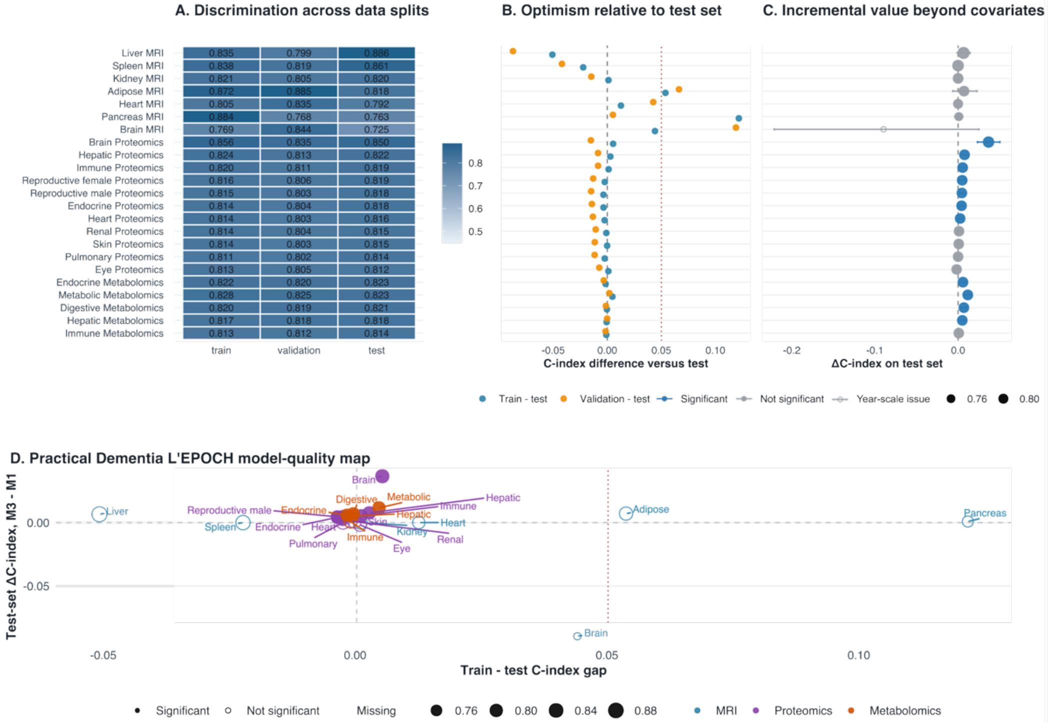


**A**) Heatmap showing C-index values for dementia EPOCH clocks across the training, validation, and held-out test datasets. Rows represent organ-specific dementia EPOCH clocks, including MRI-, proteomics-, and metabolomics-derived models. **B**) Dot plot showing the difference in C-index between the training or validation datasets and the held-out test dataset. Values near zero indicate limited optimism and stable generalization, whereas positive values indicate higher apparent performance in training or validation than in the test set. **C**) Forest plot showing the change in test-set C-index after adding each dementia EPOCH clock to the covariate-only model. Points indicate ΔC-index estimates, and horizontal bars indicate uncertainty intervals. Filled blue points denote models with statistically significant incremental value, whereas gray points indicate non-significant improvements. Open blue points denote models with statistically significant incremental value but clear year-scaling issues, as shown in **Supplementary Figure 1**. **D**) Scatterplot summarizing model quality by jointly comparing generalization and incremental value. The x-axis shows the train-test C-index gap, with values closer to zero indicating better generalization, while the y-axis shows the test-set ΔC-index beyond covariates, with positive values indicating added predictive utility. Points are colored by modality and labeled by organ/system. Filled circle points denote the clocks that were selected for downstream analyses.

**Figure 36: Model performance of MI EPOCH clocks across organs and modalities**

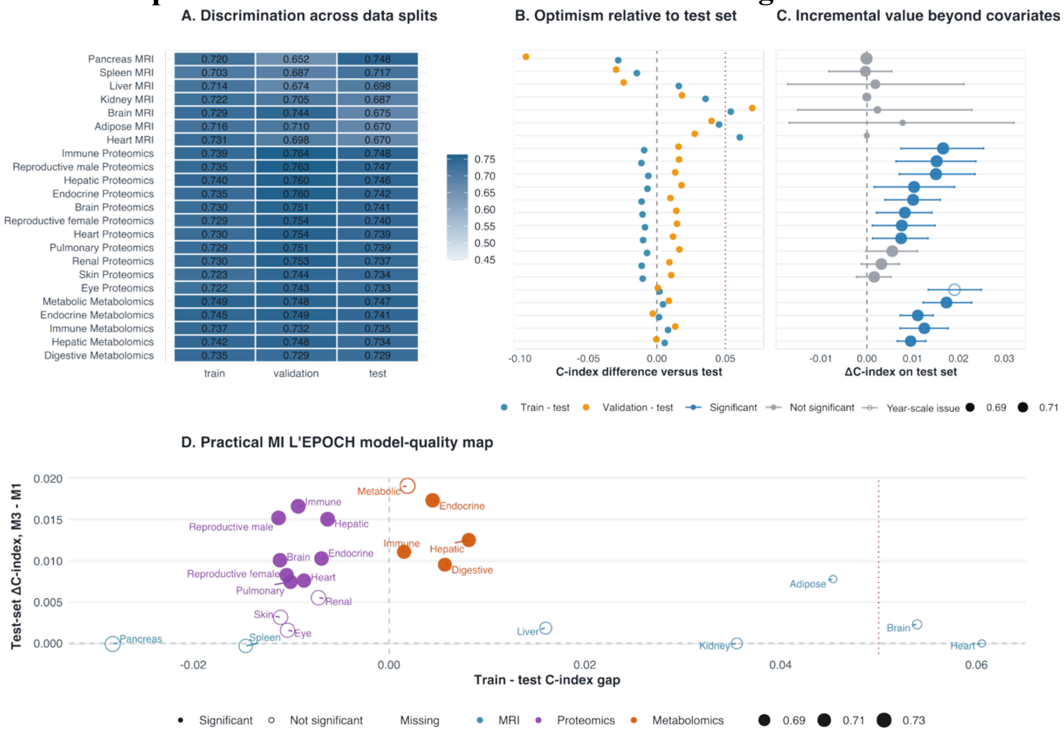


**A**) Heatmap showing C-index values for myocardial infarction (MI) EPOCH clocks across the training, validation, and held-out test datasets. Rows represent organ-specific MI EPOCH clocks, including MRI-, proteomics-, and metabolomics-derived models. **B**) Dot plot showing the difference in C-index between the training or validation datasets and the held-out test dataset. Values near zero indicate limited optimism and stable generalization, whereas positive values indicate higher apparent performance in training or validation than in the test set. **C**) Forest plot showing the change in test-set C-index after adding each MI EPOCH clock to the covariate-only model. Points indicate ΔC-index estimates, and horizontal bars indicate uncertainty intervals. Filled blue points denote models with statistically significant incremental value, whereas gray points indicate non-significant improvements. Open blue points denote models with statistically significant incremental value but clear year-scaling issues, as shown in **Supplementary Figure 1**. **D**) Scatterplot summarizing model quality by jointly comparing generalization and incremental value. The x-axis shows the train-test C-index gap, with values closer to zero indicating better generalization, while the y-axis shows the test-set ΔC-index beyond covariates, with positive values indicating added predictive utility. Points are colored by modality and labeled by organ/system. Filled circle points denote the clocks that were selected for downstream analyses.

**Figure 37: Model performance of stroke EPOCH clocks across organs and modalities**

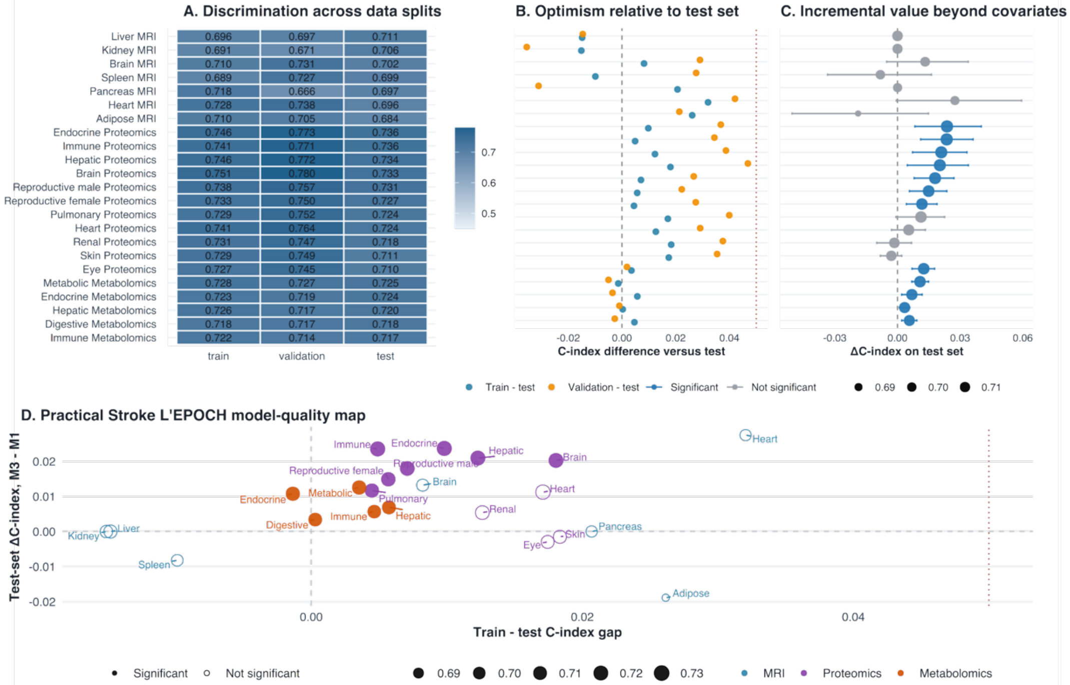


**A**) Heatmap showing C-index values for stroke EPOCH clocks across the training, validation, and held-out test datasets. Rows represent organ-specific stroke EPOCH clocks, including MRI-, proteomics-, and metabolomics-derived models. **B**) Dot plot showing the difference in C-index between the training or validation datasets and the held-out test dataset. Values near zero indicate limited optimism and stable generalization, whereas positive values indicate higher apparent performance in training or validation than in the test set. **C**) Forest plot showing the change in test-set C-index after adding each stroke EPOCH clock to the covariate-only model. Points indicate ΔC-index estimates, and horizontal bars indicate uncertainty intervals. Filled blue points denote models with statistically significant incremental value, whereas gray points indicate non-significant improvements. **D**) Scatterplot summarizing model quality by jointly comparing generalization and incremental value. The x-axis shows the train-test C-index gap, with values closer to zero indicating better generalization, while the y-axis shows the test-set ΔC-index beyond covariates, with positive values indicating added predictive utility. Points are colored by modality and labeled by organ/system. Filled circle points denote the clocks that were selected for downstream analyses.

**Figure 38: Comparing disease-specific EPOCH clocks against conventional mortality-related biomarkers for all-cause mortality**

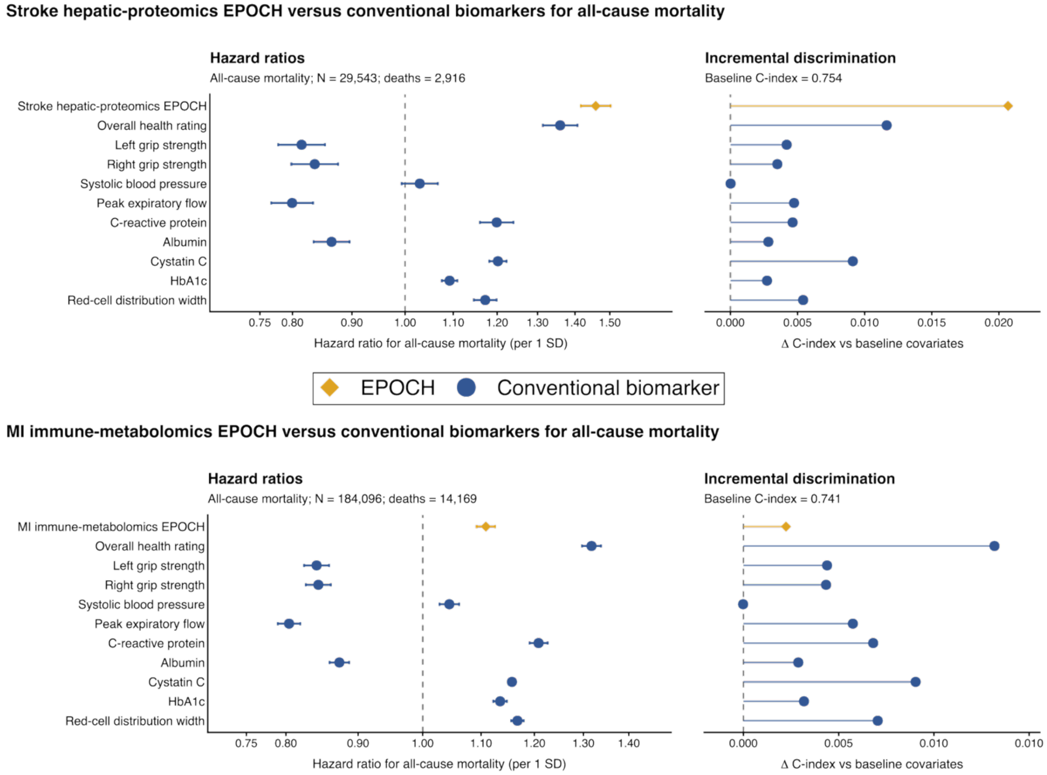


Comparison of two disease-specific EPOCH clocks with ten conventional mortality-related biomarkers for predicting all-cause mortality in UK Biobank. For each EPOCH analysis, the EPOCH clock and all ten conventional biomarkers were evaluated in the same overlapping participant population, enabling a matched comparison within each analysis. Landmark Cox proportional-hazards models were adjusted for age at assessment, sex, genetic ethnic grouping, assessment center, smoking status, and body mass index. **Upper panel**: Comparison of the stroke hepatic-proteomics EPOCH with overall health rating, left- and right-hand grip strength, systolic blood pressure, peak expiratory flow, C-reactive protein, albumin, cystatin C, HbA1c, and red-cell distribution width. **Lower panel**: Corresponding comparison for the myocardial infarction (MI) immune-metabolomics EPOCH, included as a contrasting disease-specific EPOCH with more modest mortality associations. Hazard ratios (HRs) and 95% confidence intervals for all-cause mortality per 1 s.d. higher predictor; dashed vertical lines indicate HR = 1. Incremental discrimination quantified as the change in concordance index (ΔC-index) after adding each predictor to the common covariate model; dashed vertical lines indicate ΔC-index = 0. EPOCH clocks are shown as yellow diamonds and conventional biomarkers as blue circles. The comparisons illustrate heterogeneity in mortality prognostic performance across disease-specific EPOCH clocks, with the stroke hepatic-proteomics EPOCH showing substantially greater incremental discrimination than the conventional biomarkers in its matched analysis, whereas the MI immune-metabolomics EPOCH shows a more modest improvement relative to several established mortality-related biomarkers.

**Table 1. First-generation clocks**

These clocks were first trained primarily on chronological age, then evaluated for disease or mortality in subsequent analyses.

| Study / clock | Data modality and scope | Machine learning model | Longitudinal survival setup? | Outcome evaluation |
|---|---|---|---|---|
| Hannum DNAm age[9] | Blood DNA methylation; chronological-age clock. | Elastic-net modeling of chronological age from methylation sites. | No. | Important foundation for molecular age acceleration, but outcome analyses are downstream validation. |
| Horvath multi-tissue DNAm age[10] | Multi-tissue DNA methylation; broad tissue applicability. | Predict chronological age across tissues and cell types. | No. | Different types of cancer are associated with the clock after clock construction. |
| Brain age[11] | *In vivo* imaging features from T1-weighted brain MRI | Gaussian Process regression to predict chronological age. | No. | Weaker grip strength, poorer lung function, slower walking speed, lower fluid intelligence, and mortality risk. |
| Plasma proteomic organ-aging clocks[12] or ProtBAG | Plasma proteome with organ-enriched proteins; organ-specific age models. | Bootstrap aggregated LASSO aging models. | No. | In downstream analyses, organ age predicts mortality, organ-specific functional decline, disease risk and progression. |
| MRIBAG[3] | MRI-derived brain, heart, liver, adipose, spleen, kidney and pancreas age gaps. | Organ-specific MRI models trained to predict chronological age. | No. | In downstream analyses, links to proteins, metabolites, genetics, disease and mortality. |
| MetBAG[5] | Plasma metabolomics; five organ/system metabolome age gaps. | Organ/system metabolite models trained to predict chronological age. | No. | Downstream links to cardiometabolic disease and mortality relevance. |

**Table 2. Second-generation and outcome-informed clocks**

These clocks are trained or anchored to disease, mortality, lifespan, longitudinal pace, or survival risk, which are often referred to as second-generation, or next-generation clocks.

| Study / clock | Data modality and scope | Machine learning model | Longitudinal survival setup? | Outcome evaluation |
|---|---|---|---|---|
| DNAm PhenoAge[13] | Clinical chemistry mortality-informed phenotypic age; DNAm version. | Two-step: clinical phenotypic age built from mortality-associated biomarkers, then DNAm model predicts phenotypic age. | Partly. Mortality follow-up informs the first-stage phenotypic-age measure, but final DNAm clock predicts the derived phenotype. | Outperforms earlier epigenetic age clocks for mortality, cancers, healthspan, physical function and Alzheimer's disease endpoints. |
| DNAm GrimAge / GrimAge2[14,15] | DNA methylation surrogates for plasma proteins, smoking pack-years, age and sex. | Elastic-net Cox model for time-to-death / all-cause mortality, transformed to an age-like scale. | Yes, combined both regression model and survival-based models. Uses longitudinal survival outcome and censoring in Cox modeling. | Strong mortality and healthspan prediction; GrimAge2 updates protein surrogates and performance. |
| DunedinPACE[16] | DNA methylation pace-of-aging biomarker. | Trained to reproduce a longitudinal Pace of Aging phenotype from repeated physiological measures. | No Cox survival training, but it is longitudinally anchored to rate of aging rather than chronological age. | Associated with morbidity, disability and mortality; adds information beyond GrimAge in some analyses. |
| Goeminne et al. mortality-based organ models[17] | UK Biobank plasma proteomics; conventional and organ-specific models. | First-generation chronological-age models plus mortality-based models trained with Cox elastic net to predict time to death. | Yes. Mortality models are Cox elastic-net models producing relative log mortality hazards. | They compare age-trained and mortality-trained scores largely through hazard-ratio/disease-association analyses. |
| DunedinPACNI[18] | Brain MRI biomarker estimating longitudinal pace of aging. | Trained to estimate a longitudinal pace-of-aging phenotype from a single MRI scan. | No direct Cox training, but outcome validation includes future disease and mortality. | Predicts cognitive impairment, brain atrophy, dementia conversion, frailty, future diseases and mortality. |
| Health Octo Tool[19] | Clinical history, routine tests, bodily system-specific clocks and functional metrics. | Aging/health-state framework incorporating system-specific age, walking speed and disability-related measures. | Not primarily a Cox clock in the same way as GrimAge or Goeminne; it is a clinical aging framework evaluated for mortality/disability. | Predicts disability, geriatric syndromes, physical performance and mortality. |
| Transcriptomic age and expected-mortality clocks[20] | Multi-species, multi-tissue transcriptomes from mammals. | Models of chronological age and expected mortality from gene expression. | Mortality/expected-mortality target. | Predicts time-to-death, chronic disease, lifespan-modulating interventions and rejuvenation signatures. |

**Table 3: The characteristics of participants consolidated via the MULTI study**

| Data type | BAG/Omics | Study | Country | N | Age | Sex (Female) | Event follow-up years |
|---|---|---|---|---|---|---|---|
| Individual | MRI | UKBB[b] | UK | 43,135 | 64.15± 7.53 | 19,744/53% | 5-16 |
| | Proteomics | | | 50,316 | 64.16± 8.04 | 28,581/54% | |
| | Metabolomics | | | 274,247 | 56.56± 8.08 | 147,994/54% | |
| | Genetics | | | 342,341 | 56.80± 7.91 | 181,440/53% | |
| | Mortality & disease diagnosis | | | 494,951 | 56.53± 8.09 | 268,970/54% | |
| Summary | Genetics | FinnGen | Finnland | 521 | NA[a] | NA[a] | NA[a] |
| | | PGC | Global | 6 | NA[a] | NA[a] | NA[a] |

[a]FinnGen and PGC only provide summary-level GWAS statistics; individual-level age and sex information are unavailable.

**Table 4: LDSC statistics for scrutinizing the GWAS of the 22 mortality EPOCH clocks**

| Mortality EPOCH clock | Modality | h2 mean | h2 std | Intercept | Intercept std | Ratio | Ratio std | Lambda gc | Mean chi2 |
|---|---|---|---|---|---|---|---|---|---|
| Adipose mri | MRI | 0.2384 | 0.0375 | 1.0253 | 0.0073 | 0.2247 | 0.0653 | 1.0895 | 1.1124 |
| Brain mri | MRI | 0.2503 | 0.0246 | 1.0088 | 0.0084 | 0.0554 | 0.0525 | 1.1301 | 1.1594 |
| Heart mri | MRI | 0.1574 | 0.0276 | 1.0048 | 0.0083 | 0.0532 | 0.093 | 1.0741 | 1.0895 |
| Kidney mri | MRI | 0.0867 | 0.021 | 1.0018 | 0.0073 | 0.0394 | 0.1553 | 1.0405 | 1.0469 |
| Liver mri | MRI | 0.1745 | 0.0495 | 1.0097 | 0.0079 | 0.1293 | 0.1055 | 1.0557 | 1.0751 |
| Pancreas mri | MRI | 0.2495 | 0.0338 | 1.0036 | 0.0086 | 0.0331 | 0.0793 | 1.0895 | 1.1087 |
| Spleen mri | MRI | 0.2177 | 0.036 | 1.0244 | 0.0082 | 0.2141 | 0.0722 | 1.071 | 1.1138 |
| Brain proteomics | Proteomics | 0.1499 | 0.0228 | 1.0297 | 0.0097 | 0.2035 | 0.0662 | 1.1082 | 1.1459 |
| Endocrine proteomics | Proteomics | 0.1486 | 0.0243 | 1.0334 | 0.009 | 0.2275 | 0.0611 | 1.1113 | 1.1468 |
| Eye proteomics | Proteomics | 0.1748 | 0.0225 | 1.04 | 0.0091 | 0.2254 | 0.0513 | 1.1238 | 1.1776 |
| Heart proteomics | Proteomics | 0.0634 | 0.015 | 1.0074 | 0.007 | 0.1286 | 0.1231 | 1.0588 | 1.0572 |
| Hepatic proteomics | Proteomics | 0.1549 | 0.0215 | 1.0134 | 0.0078 | 0.0957 | 0.0553 | 1.0988 | 1.1402 |
| Immune proteomics | Proteomics | 0.1499 | 0.0199 | 1.0174 | 0.0078 | 0.1279 | 0.0569 | 1.0957 | 1.1363 |
| Pulmonary proteomics | Proteomics | 0.1335 | 0.021 | 1.0379 | 0.0091 | 0.2681 | 0.0642 | 1.105 | 1.1412 |
| Renal proteomics | Proteomics | 0.1528 | 0.0327 | 1.02 | 0.0082 | 0.1457 | 0.0598 | 1.0988 | 1.1375 |
| Reproductive female proteomics | Proteomics | 0.106 | 0.0173 | 1.0346 | 0.008 | 0.293 | 0.0675 | 1.0957 | 1.1182 |
| Reproductive male proteomics | Proteomics | 0.1392 | 0.0259 | 1.0165 | 0.0084 | 0.1318 | 0.067 | 1.0895 | 1.1254 |
| Skin proteomics | Proteomics | 0.2581 | 0.0751 | 1.0572 | 0.0094 | 0.2151 | 0.0353 | 1.1428 | 1.2661 |
| Digestive metabolomics | Metabolomics | 0.082 | 0.0104 | 1.0412 | 0.0131 | 0.1122 | 0.0358 | 1.207 | 1.3675 |
| Endocrine metabolomics | Metabolomics | 0.1029 | 0.0132 | 1.0699 | 0.0145 | 0.1452 | 0.03 | 1.2697 | 1.4813 |
| Hepatic metabolomics | Metabolomics | 0.0808 | 0.0104 | 1.0468 | 0.0123 | 0.1172 | 0.0308 | 1.2168 | 1.3995 |
| Immune metabolomics | Metabolomics | 0.0925 | 0.0138 | 1.0366 | 0.0142 | 0.0855 | 0.0331 | 1.2038 | 1.4282 |

**Table 5: GCTA statistics for comparative SNP-based heritability estimates between the 22 mortality EPOCH clocks and corresponding BAGs**

| Clock class | Clock name | Method type | V_G | V_G_SE | V_e | V_e_SE | Vp | Vp_SE | h2 | h2_SE | logL | logL0 | LRT | Pvalue |
|---|---|---|---|---|---|---|---|---|---|---|---|---|---|---|
| Mortality_L_EPOCH | adipose_mri | GCTA_REML_hsq | 0.499742 | 0.035525 | 0.491617 | 0.033916 | 0.991359 | 0.010528 | 0.504098 | 0.034621 | -8902.681 | -9022.287 | 239.213 | 2.92E-54 |
| Mortality_L_EPOCH | brain_mri | GCTA_REML_hsq | 0.40284 | 0.023964 | 0.596169 | 0.022853 | 0.999009 | 0.008565 | 0.40324 | 0.023167 | -13798.949 | -13970.986 | 344.075 | 4.13E-77 |
| Mortality_L_EPOCH | heart_mri | GCTA_REML_hsq | 0.228165 | 0.023477 | 0.768045 | 0.023612 | 0.99621 | 0.008618 | 0.229033 | 0.023316 | -13368.679 | -13423.861 | 110.363 | 4.08E-26 |
| Mortality_L_EPOCH | kidney_mri | GCTA_REML_hsq | 0.142835 | 0.023407 | 0.838591 | 0.024079 | 0.981426 | 0.008609 | 0.145538 | 0.02375 | -12774.945 | -12794.73 | 39.571 | 1.58E-10 |
| Mortality_L_EPOCH | liver_mri | GCTA_REML_hsq | 0.355118 | 0.034851 | 0.640293 | 0.03418 | 0.99541 | 0.01051 | 0.356755 | 0.034403 | -8988.468 | -9050.259 | 123.582 | 5.20E-29 |
| Mortality_L_EPOCH | pancreas_mri | GCTA_REML_hsq | 0.396418 | 0.030725 | 0.60157 | 0.029793 | 0.997988 | 0.009837 | 0.397217 | 0.030035 | -10366.663 | -10467.172 | 201.019 | 6.26E-46 |
| Mortality_L_EPOCH | spleen_mri | GCTA_REML_hsq | 0.499607 | 0.032291 | 0.488385 | 0.030691 | 0.987993 | 0.010024 | 0.505679 | 0.031468 | -9727.669 | -9877.562 | 299.788 | 1.83E-67 |
| Mortality_L_EPOCH | Brain_proteomics | GCTA_REML_hsq | 0.332041 | 0.018094 | 0.658232 | 0.01768 | 0.990273 | 0.007367 | 0.335303 | 0.01779 | -18051.935 | -18263.79 | 423.71 | 1.90E-94 |
| Mortality_L_EPOCH | Endocrine_proteomics | GCTA_REML_hsq | 0.309924 | 0.017843 | 0.677026 | 0.017567 | 0.98695 | 0.007335 | 0.314022 | 0.017658 | -18004.262 | -18197.889 | 387.255 | 1.64E-86 |
| Mortality_L_EPOCH | Eye_proteomics | GCTA_REML_hsq | 0.377252 | 0.018331 | 0.616151 | 0.017645 | 0.993404 | 0.007408 | 0.379758 | 0.017829 | -18065.193 | -18341.849 | 553.312 | 1.20E-122 |
| Mortality_L_EPOCH | Heart_proteomics | GCTA_REML_hsq | 0.109188 | 0.016306 | 0.88294 | 0.017364 | 0.992128 | 0.007325 | 0.110055 | 0.016394 | -18215.985 | -18243.2 | 54.43 | 8.05E-14 |
| Mortality_L_EPOCH | Hepatic_proteomics | GCTA_REML_hsq | 0.282564 | 0.017642 | 0.710507 | 0.017561 | 0.993072 | 0.007368 | 0.284536 | 0.017423 | -18147.305 | -18309.032 | 323.454 | 1.28E-72 |
| Mortality_L_EPOCH | Immune_proteomics | GCTA_REML_hsq | 0.276533 | 0.017354 | 0.713641 | 0.017323 | 0.990174 | 0.007344 | 0.279278 | 0.017199 | -18100.286 | -18276.484 | 352.395 | 6.38E-79 |
| Mortality_L_EPOCH | Pulmonary_proteomics | GCTA_REML_hsq | 0.364442 | 0.018096 | 0.627803 | 0.0175 | 0.992245 | 0.007393 | 0.36729 | 0.017658 | -18057.827 | -18354.587 | 593.52 | 2.15E-131 |
| Mortality_L_EPOCH | Renal_proteomics | GCTA_REML_hsq | 0.359405 | 0.018285 | 0.636502 | 0.017713 | 0.995906 | 0.007419 | 0.360882 | 0.017799 | -18129.814 | -18367.447 | 475.267 | 1.14E-105 |
| Mortality_L_EPOCH | Reproductive_female_proteomics | GCTA_REML_hsq | 0.261989 | 0.017695 | 0.730926 | 0.01773 | 0.992916 | 0.007361 | 0.263858 | 0.017529 | -18160.379 | -18294.989 | 269.219 | 8.40E-61 |
| Mortality_L_EPOCH | Reproductive_male_proteomics | GCTA_REML_hsq | 0.276205 | 0.017638 | 0.711546 | 0.017579 | 0.987751 | 0.007329 | 0.27963 | 0.017527 | -18045.255 | -18207.128 | 323.747 | 1.11E-72 |
| Mortality_L_EPOCH | Skin_proteomics | GCTA_REML_hsq | 0.543153 | 0.018516 | 0.439842 | 0.016838 | 0.982996 | 0.007418 | 0.552549 | 0.017516 | -17634.854 | -18349.167 | 1428.626 | 0 |
| Mortality_L_EPOCH | Digestive_metabolomics | GCTA_HEreg | NA | NA | NA | NA | NA | NA | 0.136109 | 0.00500645 | NA | NA | NA | 9.33E-163 |
| Mortality_L_EPOCH | Endocrine_metabolomics | GCTA_HEreg | NA | NA | NA | NA | NA | NA | 0.155201 | 0.00493927 | NA | NA | NA | 1.02E-216 |
| Mortality_L_EPOCH | Hepatic_metabolomics | GCTA_HEreg | NA | NA | NA | NA | NA | NA | 0.12477 | 0.00595125 | NA | NA | NA | 1.36E-97 |
| Mortality_L_EPOCH | Immune_metabolomics | GCTA_HEreg | NA | NA | NA | NA | NA | NA | 0.169462 | 0.00499235 | NA | NA | NA | 1.48E-252 |
| Aging_BAG | adipose_mri | GCTA_REML_hsq | 0.376904 | 0.034238 | 0.621718 | 0.033417 | 0.998622 | 0.010378 | 0.377424 | 0.033602 | -9319.176 | -9385.567 | 132.782 | 5.05E-31 |
| Aging_BAG | brain_mri | GCTA_REML_hsq | 0.475008 | 0.021218 | 0.525049 | 0.019642 | 1.000058 | 0.008138 | 0.474981 | 0.020076 | -15446.389 | -15783.017 | 673.255 | 9.79E-149 |
| Aging_BAG | heart_mri | GCTA_REML_hsq | 0.283515 | 0.023346 | 0.71077 | 0.023098 | 0.994285 | 0.008485 | 0.285144 | 0.02309 | -13746.074 | -13829.818 | 167.489 | 1.31E-38 |
| Aging_BAG | kidney_mri | GCTA_REML_hsq | 0.428816 | 0.024984 | 0.571246 | 0.02382 | 1.000062 | 0.008726 | 0.428789 | 0.024107 | -13323.895 | -13494.659 | 341.528 | 1.48E-76 |
| Aging_BAG | liver_mri | GCTA_REML_hsq | 0.392579 | 0.034176 | 0.609123 | 0.033266 | 1.001702 | 0.010402 | 0.391912 | 0.033382 | -9367.126 | -9444.699 | 155.145 | 6.51E-36 |
| Aging_BAG | pancreas_mri | GCTA_REML_hsq | 0.449763 | 0.030282 | 0.546832 | 0.028993 | 0.996595 | 0.009694 | 0.4513 | 0.029413 | -10663.49 | -10793.69 | 260.4 | 7.02E-59 |

| | | | | | | | | | | | | | |
|---|---|---|---|---|---|---|---|---|---|---|---|---|---|
| Aging_BAG | spleen_mri | GCTA_REML_hsq | 0.328403 | 0.030738 | 0.671673 | 0.030247 | 1.000076 | 0.009888 | 0.328378 | 0.030221 | -10296.273 | -10365.133 | 137.721 | 4.19E-32 |
| Aging_BAG | Brain_proteomics | GCTA_REML_hsq | 0.314173 | 0.017923 | 0.683772 | 0.017649 | 0.997946 | 0.007414 | 0.31482 | 0.017538 | -18216.738 | -18427.058 | 420.64 | 8.85E-94 |
| Aging_BAG | Endocrine_proteomics | GCTA_REML_hsq | 0.221297 | 0.017529 | 0.775766 | 0.017829 | 0.997063 | 0.007379 | 0.221949 | 0.017378 | -18269.948 | -18360.982 | 182.069 | 8.56E-42 |
| Aging_BAG | Eye_proteomics | GCTA_REML_hsq | 0.236692 | 0.017995 | 0.760757 | 0.018169 | 0.997449 | 0.007388 | 0.237297 | 0.017806 | -18263.964 | -18365.679 | 203.43 | 1.86E-46 |
| Aging_BAG | Heart_proteomics | GCTA_REML_hsq | 0.141889 | 0.016778 | 0.852395 | 0.017606 | 0.994284 | 0.007345 | 0.142705 | 0.016798 | -18248.86 | -18290.628 | 83.534 | 3.13E-20 |
| Aging_BAG | Hepatic_proteomics | GCTA_REML_hsq | 0.5011 | 0.018963 | 0.493639 | 0.017522 | 0.994739 | 0.007484 | 0.50375 | 0.017958 | -17929.076 | -18378.959 | 899.767 | 5.51E-198 |
| Aging_BAG | Immune_proteomics | GCTA_REML_hsq | 0.356034 | 0.017715 | 0.63111 | 0.017183 | 0.987144 | 0.007351 | 0.360671 | 0.017389 | -17967.286 | -18278.237 | 621.902 | 1.44E-137 |
| Aging_BAG | Pulmonary_proteomics | GCTA_REML_hsq | 0.41853 | 0.018322 | 0.576118 | 0.017403 | 0.994649 | 0.007435 | 0.420782 | 0.017656 | -18041.994 | -18418.814 | 753.641 | 3.24E-166 |
| Aging_BAG | Renal_proteomics | GCTA_REML_hsq | 0.413501 | 0.018571 | 0.582549 | 0.017664 | 0.99605 | 0.007445 | 0.415141 | 0.017897 | -18075.274 | -18387.546 | 624.543 | 3.84E-138 |
| Aging_BAG | Reproductive_female_proteomics | GCTA_REML_hsq | 0.286113 | 0.017935 | 0.705476 | 0.017805 | 0.991589 | 0.00736 | 0.28854 | 0.017733 | -18117.11 | -18270.625 | 307.029 | 4.85E-69 |
| Aging_BAG | Reproductive_male_proteomics | GCTA_REML_hsq | 0.414835 | 0.018404 | 0.579152 | 0.017498 | 0.993988 | 0.00743 | 0.417345 | 0.017762 | -18030.635 | -18364.822 | 668.373 | 1.13E-147 |
| Aging_BAG | Skin_proteomics | GCTA_REML_hsq | 0.470461 | 0.017986 | 0.51137 | 0.01676 | 0.981831 | 0.007364 | 0.479167 | 0.017331 | -17726.111 | -18377.414 | 1302.605 | 1.53E-285 |
| Aging_BAG | Digestive_metabolomics | GCTA_HEreg | NA | NA | NA | NA | NA | NA | 0.205105 | 0.00535569 | NA | NA | NA | 6.986e-321 |
| Aging_BAG | Endocrine_metabolomics | GCTA_HEreg | NA | NA | NA | NA | NA | NA | 0.295909 | 0.0115617 | NA | NA | NA | 1.78E-144 |
| Aging_BAG | Hepatic_metabolomics | GCTA_HEreg | NA | NA | NA | NA | NA | NA | 0.319407 | 0.00731584 | NA | NA | NA | 0 |
| Aging_BAG | Immune_metabolomics | GCTA_HEreg | NA | NA | NA | NA | NA | NA | 0.224936 | 0.00636435 | NA | NA | NA | 1.28E-273 |
| Aging_BAG | Metabolic_metabolomics | GCTA_HEreg | NA | NA | NA | NA | NA | NA | 0.176803 | 0.00682369 | NA | NA | NA | 5.11E-148 |

#### Table 6: SBayesS statistics for the three key genetic parameters

| Mortality clock | Modality | Pi | Pi_se | S | S_se | h2_mean | h2_se |
|---|---|---|---|---|---|---|---|
| adipose_mri | MRI | 0.026012 | 0.017163 | -0.650344 | 0.219951 | 0.298235 | 0.024278 |
| brain_mri | MRI | 0.066864 | 0.004532 | -0.163869 | 0.098401 | 0.246919 | 0.009705 |
| heart_mri | MRI | 0.043667 | 0.005097 | -0.585068 | 0.190131 | 0.146673 | 0.012299 |
| kidney_mri | MRI | 0.039815 | 0.003372 | -0.925658 | 0.27452 | 0.080489 | 0.011204 |
| liver_mri | MRI | 0.000838 | 0.002199 | -0.958123 | 0.500171 | 0.075013 | 0.058459 |
| pancreas_mri | MRI | 0.049204 | 0.005129 | -0.432533 | 0.114201 | 0.230162 | 0.015746 |
| spleen_mri | MRI | 0.01611 | 0.01549 | -0.638674 | 0.188755 | 0.272618 | 0.032609 |
| Brain_proteomics | Proteomics | 0.001277 | 0.000297 | -0.931309 | 0.093114 | 0.127909 | 0.01166 |
| Endocrine_proteomics | Proteomics | 0.000913 | 0.000267 | -0.460664 | 0.158001 | 0.124851 | 0.012308 |
| Eye_proteomics | Proteomics | 0.003467 | 0.000405 | -0.65117 | 0.129446 | 0.183131 | 0.009818 |
| Heart_proteomics | Proteomics | 0.030886 | 0.002713 | -0.35229 | 0.140626 | 0.066025 | 0.006941 |
| Hepatic_proteomics | Proteomics | 0.000354 | 0.000108 | -1.317431 | 0.142189 | 0.079039 | 0.007282 |
| Immune_proteomics | Proteomics | 0.002383 | 0.000364 | -0.905775 | 0.113667 | 0.135935 | 0.008777 |
| Pulmonary_proteomics | Proteomics | 0.000884 | 0.000223 | -1.203857 | 0.142913 | 0.124568 | 0.010873 |
| Renal_proteomics | Proteomics | 0.000262 | 0.000142 | -1.040232 | 0.308863 | 0.114029 | 0.010232 |
| Reproductive_female_proteomics | Proteomics | 0.002228 | 0.000421 | -1.126391 | 0.091279 | 0.121501 | 0.009732 |
| Reproductive_male_proteomics | Proteomics | 0.000296 | 7.00E-05 | -1.199914 | 0.178442 | 0.080852 | 0.006521 |
| Skin_proteomics | Proteomics | 0.000553 | 8.40E-05 | -0.402161 | 0.147199 | 0.188847 | 0.007002 |
| Digestive_metabolomics | Metabolomics | 0.000929 | 0.00012 | -1.329301 | 0.13323 | 0.076325 | 0.002052 |
| Endocrine_metabolomics | Metabolomics | 0.003651 | 0.000243 | -0.962163 | 0.056371 | 0.105835 | 0.002252 |
| Hepatic_metabolomics | Metabolomics | 0.003752 | 0.000311 | -1.00761 | 0.061381 | 0.074246 | 0.002418 |
| Immune_metabolomics | Metabolomics | 0.000806 | 6.20E-05 | -1.129161 | 0.144726 | 0.110888 | 0.001689 |

**Table 7: Landmark survival analysis to predict subsequent mortality risk for the 4 metabolomics mortality EPOCH clocks' longitudinal changes**

| Organ label | Model | Term | Estimate | STD.error | Statistic | P.value | CI.low | CI.high |
|---|---|---|---|---|---|---|---|---|
| Endocrine | baseline_only | clock_acceleration_years_0_0 | 1.07671559 | 0.00834924 | 8.85294072 | 8.52E-19 | 1.05923939 | 1.09448014 |
| Endocrine | baseline_only | chronological_age_0_0 | 1.12278974 | 0.00627352 | 18.4611635 | 4.24E-76 | 1.1090686 | 1.13668064 |
| Endocrine | baseline_plus_delta | clock_acceleration_years_0_0 | 1.11644216 | 0.00970267 | 11.3522332 | 7.23E-30 | 1.09541151 | 1.13787658 |
| **Endocrine** | **baseline_plus_delta** | **delta_accel_years_1_minus_0** | **1.07236897** | **0.00888422** | **7.86453087** | **3.70E-15** | **1.05385772** | **1.09120537** |
| Endocrine | baseline_plus_delta | chronological_age_0_0 | 1.12137684 | 0.00624023 | 18.3578589 | 2.86E-75 | 1.10774523 | 1.1351762 |
| Digestive | baseline_only | clock_acceleration_years_0_0 | 1.07553402 | 0.01028416 | 7.08053297 | 1.44E-12 | 1.05407197 | 1.09743307 |
| Digestive | baseline_only | chronological_age_0_0 | 1.11886956 | 0.00603279 | 18.6180526 | 2.29E-77 | 1.10571789 | 1.13217766 |
| Digestive | baseline_plus_delta | clock_acceleration_years_0_0 | 1.08647462 | 0.01147366 | 7.22856921 | 4.88E-13 | 1.06231469 | 1.11118402 |
| **Digestive** | **baseline_plus_delta** | **delta_accel_years_1_minus_0** | **1.01896044** | **0.00956218** | **1.96429291** | **0.04949613** | **1.00004139** | **1.0382374** |
| Digestive | baseline_plus_delta | chronological_age_0_0 | 1.1191385 | 0.0060306 | 18.6646612 | 9.60E-78 | 1.10598841 | 1.13244494 |
| Hepatic | baseline_only | clock_acceleration_years_0_0 | 1.09100817 | 0.00906579 | 9.60779107 | 7.41E-22 | 1.07179368 | 1.11056714 |
| Hepatic | baseline_only | chronological_age_0_0 | 1.11624159 | 0.00605445 | 18.1630426 | 1.01E-73 | 1.10307397 | 1.12956638 |
| Hepatic | baseline_plus_delta | clock_acceleration_years_0_0 | 1.10944493 | 0.01036092 | 10.0241867 | 1.19E-23 | 1.08714261 | 1.13220478 |
| **Hepatic** | **baseline_plus_delta** | **delta_accel_years_1_minus_0** | **1.03058362** | **0.00889885** | **3.38529753** | **0.00071101** | **1.01276461** | **1.04871614** |
| Hepatic | baseline_plus_delta | chronological_age_0_0 | 1.11612517 | 0.00604515 | 18.173741 | 8.33E-74 | 1.10297904 | 1.12942799 |
| Immune | baseline_only | clock_acceleration_years_0_0 | 1.0956017 | 0.01247468 | 7.31912385 | 2.50E-13 | 1.06913915 | 1.12271923 |
| Immune | baseline_only | chronological_age_0_0 | 1.11576639 | 0.00590751 | 18.5427455 | 9.33E-77 | 1.10292198 | 1.12876039 |
| Immune | baseline_plus_delta | clock_acceleration_years_0_0 | 1.10103238 | 0.01359502 | 7.07967206 | 1.44E-12 | 1.07208197 | 1.13076457 |
| **Immune** | **baseline_plus_delta** | **delta_accel_years_1_minus_0** | **1.01051646** | **0.01144408** | **0.91414506** | **0.36064063** | **0.9881029** | **1.03343843** |
| Immune | baseline_plus_delta | chronological_age_0_0 | 1.11578534 | 0.00590644 | 18.5489862 | 8.31E-77 | 1.10294303 | 1.12877719 |

**Table 8: Landmark survival analysis to predict subsequent disease risk for the 4 metabolomics mortality EPOCH clocks' longitudinal changes**

| Endpoint label | Organ label | Delta column | N | Ncase | N_noncase | event_rate | followup_years_max | delta_beta | delta_se | delta_hr | delta_ci_lo | delta_ci_hi | delta_p |
|---|---|---|---|---|---|---|---|---|---|---|---|---|---|
| All-cause dementia | Endocrine | delta_clock_age_1_minus_0 | 13451 | 176 | 13275 | 0.01308453 | 10.329911 | 0.13146287 | 0.08111635 | 1.14049556 | 0.97285019 | 1.33703024 | 0.10508834 |
| All-cause dementia | Digestive | delta_clock_age_1_minus_0 | 14279 | 192 | 14087 | 0.01344632 | 10.329911 | 0.03542867 | 0.08148696 | 1.03606374 | 0.88312743 | 1.21548492 | 0.66372419 |
| All-cause dementia | Hepatic | delta_clock_age_1_minus_0 | 14036 | 188 | 13848 | 0.01339413 | 10.329911 | 0.14375249 | 0.08137729 | 1.1545983 | 0.98437634 | 1.35425567 | 0.07731302 |
| All-cause dementia | Immune | delta_clock_age_1_minus_0 | 14556 | 198 | 14358 | 0.01360264 | 10.329911 | -0.042714 | 0.07956178 | 0.95818536 | 0.81983258 | 1.11988619 | 0.59136004 |
| Asthma | Endocrine | delta_clock_age_1_minus_0 | 11863 | 272 | 11591 | 0.02292843 | 10.329911 | 0.14401229 | 0.06622401 | 1.1548983 | 1.01431471 | 1.31496671 | 0.02965835 |
| Asthma | Digestive | delta_clock_age_1_minus_0 | 12595 | 306 | 12289 | 0.02429536 | 10.329911 | 0.07100998 | 0.06314515 | 1.07359194 | 0.94861286 | 1.21503692 | 0.26077909 |
| Asthma | Hepatic | delta_clock_age_1_minus_0 | 12381 | 302 | 12079 | 0.02439221 | 10.329911 | 0.0306635 | 0.06401182 | 1.03113847 | 0.90955514 | 1.16897426 | 0.63191814 |
| Asthma | Immune | delta_clock_age_1_minus_0 | 12847 | 316 | 12531 | 0.02459718 | 10.329911 | 0.0210441 | 0.06044029 | 1.02126709 | 0.90717594 | 1.14970694 | 0.72770498 |
| COPD | Endocrine | delta_clock_age_1_minus_0 | 13263 | 327 | 12936 | 0.02465506 | 10.329911 | 0.28134898 | 0.05451797 | 1.32491589 | 1.19064331 | 1.4743308 | 2.46E-07 |
| COPD | Digestive | delta_clock_age_1_minus_0 | 14077 | 355 | 13722 | 0.02521844 | 10.329911 | 0.14171623 | 0.05796617 | 1.15224963 | 1.02850113 | 1.29088746 | 0.01449288 |
| COPD | Hepatic | delta_clock_age_1_minus_0 | 13837 | 343 | 13494 | 0.02478861 | 10.329911 | 0.14522534 | 0.05805879 | 1.1563001 | 1.03192925 | 1.29566046 | 0.01237209 |
| COPD | Immune | delta_clock_age_1_minus_0 | 14349 | 359 | 13990 | 0.02501917 | 10.329911 | 0.10388164 | 0.05719424 | 1.10946913 | 0.99181461 | 1.24108047 | 0.06932506 |
| Myocardial infarction | Endocrine | delta_clock_age_1_minus_0 | 13136 | 405 | 12731 | 0.0308313 | 10.329911 | 0.2547439 | 0.05095517 | 1.29013117 | 1.16750824 | 1.42563314 | 5.75E-07 |
| Myocardial infarction | Digestive | delta_clock_age_1_minus_0 | 13945 | 432 | 13513 | 0.03097885 | 10.329911 | 0.08416726 | 0.05288981 | 1.08781083 | 0.98069206 | 1.20662993 | 0.11152634 |
| Myocardial infarction | Hepatic | delta_clock_age_1_minus_0 | 13711 | 427 | 13284 | 0.03114288 | 10.329911 | 0.1734233 | 0.05324723 | 1.18936946 | 1.07149913 | 1.32020612 | 0.00112618 |
| Myocardial infarction | Immune | delta_clock_age_1_minus_0 | 14218 | 440 | 13778 | 0.03094669 | 10.329911 | 0.12278877 | 0.05178354 | 1.13064557 | 1.02152136 | 1.25142701 | 0.01773076 |
| Stroke | Endocrine | delta_clock_age_1_minus_0 | 13277 | 279 | 12998 | 0.02101378 | 10.329911 | -0.0276761 | 0.066115 | 0.97270335 | 0.85448056 | 1.10728302 | 0.67550426 |
| Stroke | Digestive | delta_clock_age_1_minus_0 | 14090 | 289 | 13801 | 0.020511 | 10.329911 | 0.06420881 | 0.06584861 | 1.06631504 | 0.93720386 | 1.21321284 | 0.32951189 |
| Stroke | Hepatic | delta_clock_age_1_minus_0 | 13851 | 283 | 13568 | 0.02043174 | 10.329911 | -0.1038314 | 0.06660426 | 0.9013773 | 0.79106455 | 1.027073 | 0.11901307 |
| Stroke | Immune | delta_clock_age_1_minus_0 | 14364 | 295 | 14069 | 0.02053745 | 10.329911 | -0.0830516 | 0.06394248 | 0.9203037 | 0.81189942 | 1.04318205 | 0.19399616 |

**Table 9: Detailed statistics for the within-organ and cross-organ temporal coupling analysis**

**Cross-organ modeling delta changes of clocks**:

| model | outcome_type | outcome_system | predictor_system | edge_type | outcome | predictor | beta | se_HC3 | ci_lower | ci_upper | p | n | r2 |
|---|---|---|---|---|---|---|---|---|---|---|---|---|---|
| simultaneous_baseline_predictors | annualized_change | Endocrine | Endocrine | within_system | Endocrine_delta_per_year | baseline_Endocrine_z | -0.473709 | 0.01502712 | -0.5031622 | -0.4442559 | 4.12E-218 | 13594 | 0.18505207 |
| simultaneous_baseline_predictors | annualized_change | Endocrine | Digestive | cross_system | Endocrine_delta_per_year | baseline_Digestive_z | 0.06218897 | 0.01081353 | 0.04099444 | 0.08338349 | 8.87E-09 | 13594 | 0.18505207 |
| simultaneous_baseline_predictors | annualized_change | Endocrine | Hepatic | cross_system | Endocrine_delta_per_year | baseline_Hepatic_z | 0.0369144 | 0.01316342 | 0.0111141 | 0.06271471 | 0.00504233 | 13594 | 0.18505207 |
| simultaneous_baseline_predictors | annualized_change | Endocrine | Immune | cross_system | Endocrine_delta_per_year | baseline_Immune_z | -0.028689 | 0.01153348 | -0.0512946 | -0.0060834 | 0.01286618 | 13594 | 0.18505207 |
| simultaneous_baseline_predictors | annualized_change | Digestive | Endocrine | cross_system | Digestive_delta_per_year | baseline_Endocrine_z | 0.07121907 | 0.01324633 | 0.04525626 | 0.09718188 | 7.59E-08 | 13594 | 0.18414331 |
| simultaneous_baseline_predictors | annualized_change | Digestive | Digestive | within_system | Digestive_delta_per_year | baseline_Digestive_z | -0.4794791 | 0.0115408 | -0.5020991 | -0.4568592 | 0 | 13594 | 0.18414331 |
| simultaneous_baseline_predictors | annualized_change | Digestive | Hepatic | cross_system | Digestive_delta_per_year | baseline_Hepatic_z | 0.01555548 | 0.01256001 | -0.0090621 | 0.0401731 | 0.21553361 | 13594 | 0.18414331 |
| simultaneous_baseline_predictors | annualized_change | Digestive | Immune | cross_system | Digestive_delta_per_year | baseline_Immune_z | 0.00923718 | 0.01100969 | -0.0123418 | 0.03081617 | 0.40146634 | 13594 | 0.18414331 |
| simultaneous_baseline_predictors | annualized_change | Hepatic | Endocrine | cross_system | Hepatic_delta_per_year | baseline_Endocrine_z | 0.13457839 | 0.01753422 | 0.10021131 | 0.16894546 | 1.65E-14 | 13594 | 0.18741344 |
| simultaneous_baseline_predictors | annualized_change | Hepatic | Digestive | cross_system | Hepatic_delta_per_year | baseline_Digestive_z | 0.04503582 | 0.01146148 | 0.02257132 | 0.06750031 | 8.52E-05 | 13594 | 0.18741344 |
| simultaneous_baseline_predictors | annualized_change | Hepatic | Hepatic | within_system | Hepatic_delta_per_year | baseline_Hepatic_z | -0.5667359 | 0.02347493 | -0.6127468 | -0.5207251 | 9.02E-129 | 13594 | 0.18741344 |
| simultaneous_baseline_predictors | annualized_change | Hepatic | Immune | cross_system | Hepatic_delta_per_year | baseline_Immune_z | -0.010155 | 0.01219143 | -0.0340502 | 0.01374019 | 0.40486523 | 13594 | 0.18741344 |
| simultaneous_baseline_predictors | annualized_change | Immune | Endocrine | cross_system | Immune_delta_per_year | baseline_Endocrine_z | 0.04417497 | 0.01137318 | 0.02188354 | 0.0664664 | 0.00010269 | 13594 | 0.1446605 |
| simultaneous_baseline_predictors | annualized_change | Immune | Digestive | cross_system | Immune_delta_per_year | baseline_Digestive_z | 0.01044826 | 0.00876703 | -0.0067351 | 0.02763165 | 0.23335274 | 13594 | 0.1446605 |
| simultaneous_baseline_predictors | annualized_change | Immune | Hepatic | cross_system | Immune_delta_per_year | baseline_Hepatic_z | -0.0202888 | 0.01089594 | -0.0416448 | 0.00106727 | 0.06259626 | 13594 | 0.1446605 |
| simultaneous_baseline_predictors | annualized_change | Immune | Immune | within_system | Immune_delta_per_year | baseline_Immune_z | -0.3266283 | 0.00950791 | -0.3452638 | -0.3079928 | 1.26E-258 | 13594 | 0.1446605 |

**Within-organ modeling of baseline clocks predicting follow-up clocks**:

| model | outcome_type | outcome_system | predictor_system | edge_type | outcome | predictor | beta | se_HC3 | ci_lower | ci_upper | p | n | r2 |
|---|---|---|---|---|---|---|---|---|---|---|---|---|---|
| lagged_followup_sensitivity | instance1_level | Endocrine | Endocrine | within_system | Endocrine_t1 | baseline_Endocrine_z | 1.77092832 | 0.05846403 | 1.65633883 | 1.88551781 | 1.51E-201 | 13594 | 0.23487269 |
| lagged_followup_sensitivity | instance1_level | Endocrine | Digestive | cross_system | Endocrine_t1 | baseline_Digestive_z | 0.26120907 | 0.04355585 | 0.1758396 | 0.34657854 | 2.01E-09 | 13594 | 0.23487269 |
| lagged_followup_sensitivity | instance1_level | Endocrine | Hepatic | cross_system | Endocrine_t1 | baseline_Hepatic_z | 0.12408045 | 0.05151442 | 0.02311219 | 0.22504871 | 0.01601144 | 13594 | 0.23487269 |
| lagged_followup_sensitivity | instance1_level | Endocrine | Immune | cross_system | Endocrine_t1 | baseline_Immune_z | -0.0929655 | 0.04546088 | -0.1820689 | -0.0038622 | 0.0408591 | 13594 | 0.23487269 |
| lagged_followup_sensitivity | instance1_level | Digestive | Endocrine | cross_system | Digestive_t1 | baseline_Endocrine_z | 0.28095927 | 0.05212303 | 0.17879813 | 0.38312042 | 7.03E-08 | 13594 | 0.15747084 |
| lagged_followup_sensitivity | instance1_level | Digestive | Digestive | within_system | Digestive_t1 | baseline_Digestive_z | 1.25779428 | 0.04656575 | 1.16652541 | 1.34906314 | 1.09E-160 | 13594 | 0.15747084 |
| lagged_followup_sensitivity | instance1_level | Digestive | Hepatic | cross_system | Digestive_t1 | baseline_Hepatic_z | 0.03818928 | 0.04947788 | -0.0587874 | 0.13516592 | 0.44020589 | 13594 | 0.15747084 |
| lagged_followup_sensitivity | instance1_level | Digestive | Immune | cross_system | Digestive_t1 | baseline_Immune_z | 0.06210585 | 0.04283809 | -0.0218568 | 0.14606852 | 0.14711962 | 13594 | 0.15747084 |
| lagged_followup_sensitivity | instance1_level | Hepatic | Endocrine | cross_system | Hepatic_t1 | baseline_Endocrine_z | 0.51444241 | 0.07204714 | 0.37323001 | 0.65565481 | 9.31E-13 | 13594 | 0.18135688 |

| | | | | | | | | | | | | | |
|---|---|---|---|---|---|---|---|---|---|---|---|---|---|
| lagged_followup_sensitivity | instance1_level | Hepatic | Digestive | cross_system | Hepatic_t1 | baseline_Digestive_z | 0.18697719 | 0.04676031 | 0.09532699 | 0.27862739 | 6.37E-05 | 13594 | 0.18135688 |
| lagged_followup_sensitivity | instance1_level | Hepatic | Hepatic | within_system | Hepatic_t1 | baseline_Hepatic_z | 1.22954053 | 0.10011613 | 1.03331291 | 1.42576815 | 1.14E-34 | 13594 | 0.18135688 |
| lagged_followup_sensitivity | instance1_level | Hepatic | Immune | cross_system | Hepatic_t1 | baseline_Immune_z | -0.0236449 | 0.04893146 | -0.1195506 | 0.07226073 | 0.62893569 | 13594 | 0.18135688 |
| lagged_followup_sensitivity | instance1_level | Immune | Endocrine | cross_system | Immune_t1 | baseline_Endocrine_z | 0.16832631 | 0.04388096 | 0.08231964 | 0.25433299 | 0.00012507 | 13594 | 0.18302482 |
| lagged_followup_sensitivity | instance1_level | Immune | Digestive | cross_system | Immune_t1 | baseline_Digestive_z | 0.05469737 | 0.03498989 | -0.0138828 | 0.12327755 | 0.11799766 | 13594 | 0.18302482 |
| lagged_followup_sensitivity | instance1_level | Immune | Hepatic | cross_system | Immune_t1 | baseline_Hepatic_z | -0.0829062 | 0.04240559 | -0.1660212 | 0.00020875 | 0.05057394 | 13594 | 0.18302482 |
| lagged_followup_sensitivity | instance1_level | Immune | Immune | within_system | Immune_t1 | baseline_Immune_z | 1.29842958 | 0.03731214 | 1.22529778 | 1.37156137 | 2.51E-265 | 13594 | 0.18302482 |

### Table 10: Disease endpoints from FinnGen and PGC

#### FinnGen DEs

| DE | Name | Category | Num_cases | Num_controls |
|---|---|---|---|---|
| RX_PARACETAMOL_NSAID | Paracetamol of NSAID medication | Drug purchase endpoints | 314977 | 62300 |
| RX_ANTIHYP | Antihypertensive medication - note that there are other indications | Drug purchase endpoints | 199546 | 177731 |
| FG_CVD | Cardiovascular diseases (excluding rheumatic etc) | Cardiometabolic endpoints | 185353 | 191924 |
| PAIN | Pain (limb, back, neck, head abdominally) | Miscellaneous, not yet classified endpoints | 171922 | 204598 |
| K11_CARIES_1_ONLYAVO | Dental caries 1, only avohilmo | XI Diseases of the digestive system (K11_) | 161113 | 216164 |
| M13_ARTHROPATHIES | Arthropathies | XIII Diseases of the musculoskeletal system and connective tissue (M13_) | 136415 | 240862 |
| RX_STATIN | Statin medication | IX Diseases of the circulatory system (I9_) | 127169 | 250108 |
| K11_ORAL | Diseases of oral cavity, salivary glands and jaws | XI Diseases of the digestive system (K11_) | 118043 | 259234 |
| PULM_INFECTIONS | COPD/asthma/ILD related infections | Interstitial lung disease endpoints | 117312 | 259965 |
| I9_HYPTENS | Hypertension | IX Diseases of the circulatory system (I9_) | 111581 | 265626 |
| ANTIDEPRESSANTS | Depression medications | Comorbidities of Neurological endpoints | 106785 | 88536 |
| O15_DELIV_SPONT | Single spontaneous delivery | XV Pregnancy, childbirth and the puerperium (O15_) | 106627 | 92306 |
| M13_DORSOPATHY | Dorsopathies | XIII Diseases of the musculoskeletal system and connective tissue (M13_) | 106313 | 270964 |
| N14_FEMALEGENNONINF | Noninflammatory disorders of female genital tract | XIV Diseases of the genitourinary system (N14_) | 103306 | 107564 |
| M13_SOFTTISSUE | Soft tissue disorders | XIII Diseases of the musculoskeletal system and connective tissue (M13_) | 102065 | 275212 |
| KRA_PSY_ANYMENTAL | Any mental disorder | Psychiatric endpoints from Katri Räikkönen | 99751 | 277526 |
| M13_ARTHROSIS_INCLAVO | Artrosis, including avohilmo | XIII Diseases of the musculoskeletal system and connective tissue (M13_) | 96500 | 280777 |
| AUTOIMMUNE | Autoimmune diseases | Diseases marked as autimmune origin | 96150 | 281127 |
| J10_UPPERDIS | Other diseases of upper respiratory tract | X Diseases of the respiratory system (J10_) | 93935 | 283342 |
| FALLS | Falls/tendency to fall | Miscellaneous, not yet classified endpoints | 92857 | 284420 |
| I9_HYPTENSESS | Hypertension, essential | IX Diseases of the circulatory system (I9_) | 92462 | 265626 |
| G6_EPIPAROX | Episodal and paroxysmal disorders | VI Diseases of the nervous system (G6_) | 89440 | 287837 |
| K11_GINGIVITIS_PERIODONTAL | Gingivitis and periodontal diseases | XI Diseases of the digestive system (K11_) | 87497 | 259234 |
| K11_PULPITIS_1_ONLYAVO | Dental pulpitis 1, only avohilmo | XI Diseases of the digestive system (K11_) | 81713 | 295564 |
| M13_OTHERJOINT | Other joint disorders | XIII Diseases of the musculoskeletal system and connective tissue (M13_) | 81676 | 240862 |
| M13_DORSOPATHYOTH | Other dorsopathies, not elsewhere classified | XIII Diseases of the musculoskeletal system and connective tissue (M13_) | 79212 | 270964 |
| FG_OTHHEART | Other heart diseases | Cardiometabolic endpoints | 77711 | 191924 |
| RX_CROHN_1STLINE | First line medication for Crohn's disease | Drug purchase endpoints | 77497 | 299780 |
| C3_CANCER_EXALLC | Malignant neoplasm (controls excluding all cancers) | II Neoplasms, from cancer register (ICD-O-3) | 77325 | 287137 |
| K11_INTESTOTH | Other diseases of intestines | XI Diseases of the digestive system (K11_) | 75346 | 301931 |
| M13_ARTHROSIS | Arthrosis | XIII Diseases of the musculoskeletal system and connective tissue (M13_) | 73710 | 240862 |
| J10_UPPERINFEC | Acute upper respiratory infections | X Diseases of the respiratory system (J10_) | 69111 | 308166 |
| CARDIAC_ARRHYTM | Cardiac arrhythmias | Comorbidities of COPD | 67035 | 220224 |
| J10_LOWCHRON | Chronic lower respiratory diseases | X Diseases of the respiratory system (J10_) | 65991 | 311286 |
| RX_N05C | Use of hypnotics and sedatives | V Mental and behavioural disorders (F5_) | 65814 | 311463 |
| I9_IHD | Ischaemic heart disease, wide definition | IX Diseases of the circulatory system (I9_) | 63744 | 313533 |
| J10_INFLUPNEU | Influenza and pneumonia | X Diseases of the respiratory system (J10_) | 62604 | 314673 |
| O15_PREG_ABORT | Pregnancy with abortive outcome | XV Pregnancy, childbirth and the puerperium (O15_) | 61248 | 149622 |
| I9_CVD_HARD | Hard cardiovascular diseases | Cardiometabolic endpoints | 61240 | 316037 |
| H7_CATARACTSENILE | Senile cataract | VII Diseases of the eye and adnexa (H7_) | 59522 | 312864 |
| M13_DORSALGIA | Dorsalgia | XIII Diseases of the musculoskeletal system and connective tissue (M13_) | 59438 | 270964 |
| J10_PNEUMONIA | All pneumoniae | X Diseases of the respiratory system (J10_) | 58174 | 319103 |
| RX_CODEINE_TRAMADOL | Codeine or tramadol medication | Drug purchase endpoints | 57823 | 319454 |
| T2D | Type 2 diabetes, definitions combined | Diabetes endpoints | 57698 | 308252 |
| K11_OESSTODUO | Diseases of oesophagus, stomach and duodenum | XI Diseases of the digestive system (K11_) | 56890 | 320387 |
| E4_THYROID | Disorders of the thyroid gland | IV Endocrine, nutritional and metabolic diseases (E4_) | 56574 | 320703 |
| BRONCHITIS | Bronchitis | Asthma and related endpoints | 55222 | 322055 |
| K11_HERNIA | Hernia | XI Diseases of the digestive system (K11_) | 54691 | 322586 |
| KELA_DIAB_INSUL_EXMORE | Diabetes, insulin treatment (Kela reimbursement) (more control exclusions) | Diabetes endpoints | 53892 | 308249 |
| AUTOIMMUNE_NONTHYROID | Autoimmune diseases excluding thyroid diseases | Diseases marked as autimmune origin | 53213 | 280251 |
| I9_DISVEINLYMPH | Diseases of veins, lymphatic vessels and lymph nodes, not elsewhere classified | IX Diseases of the circulatory system (I9_) | 53156 | 324121 |
| E4_METABOLIA | Metabolic disorders | IV Endocrine, nutritional and metabolic diseases (E4_) | 53127 | 324150 |
| AUTOIMMUNE_NONTHYROID_STRICT | Autoimmune diseases excluding thyroid diseases, strict definition | Diseases marked as autimmune origin | 51863 | 275890 |
| N14_URINOTH | Other diseases of urinary system | XIV Diseases of the genitourinary system (N14_) | 49276 | 328001 |
| F5_DEPRESSION_DYSTHYMIA | Depression or dysthymia | V Mental and behavioural disorders (F5_) | 48847 | 225483 |
| O15_COMPLIC_LAB_DELIV | Complications of labour and delivery | XV Pregnancy, childbirth and the puerperium (O15_) | 48093 | 162777 |
| F5_MOOD | Mood [affective] disorders | V Mental and behavioural disorders (F5_) | 48085 | 329192 |
| I9_CORATHER | Coronary atherosclerosis | IX Diseases of the circulatory system (I9_) | 47550 | 313400 |
| N14_MALEGEN | Diseases of male genital organs | XIV Diseases of the genitourinary system (N14_) | 47110 | 119297 |
| G6_NERPLEX | Nerve, nerve root and plexus disorders | VI Diseases of the nervous system (G6_) | 46900 | 330377 |
| E4_DM2NASCOMP | Type 2 diabetes with other specified/multiple/unspecified complications | IV Endocrine, nutritional and metabolic diseases (E4_) | 46373 | 308280 |
| I9_AF | Atrial fibrillation and flutter | IX Diseases of the circulatory system (I9_) | 45766 | 191924 |
| H8_OTHEREAR | Other disorders of ear | VIII Diseases of the ear and mastoid process (H8_) | 45541 | 331736 |
| AB1_OTHER_BACTERIAL | Other bacterial diseases | I Certain infectious and parasitic diseases (AB1_) | 44934 | 332343 |
| M13_ARTHROSIS_KNEE | Gonarthrosis | XIII Diseases of the musculoskeletal system and connective tissue (M13_) | 44688 | 240862 |
| SLEEP | Sleep disorders (combined) | Neurological endpoints | 44299 | 329251 |
| I9_CHD | Major coronary heart disease event | IX Diseases of the circulatory system (I9_) | 43518 | 333759 |
| J10_CHRONTONSADEN | Chronic diseases of tonsils and adenoids | X Diseases of the respiratory system (J10_) | 43325 | 283342 |
| F5_DEPRESSIO | Depression | V Mental and behavioural disorders (F5_) | 43280 | 329192 |
| M13_SOFTTISSUEOTH | Other soft tissue disorders, not elsewhere classified | XIII Diseases of the musculoskeletal system and connective tissue (M13_) | 43190 | 275212 |
| J10_ASTHMA_EXMORE | Asthma (more control exclusions) | X Diseases of the respiratory system (J10_) | 42163 | 202399 |

| | | | | |
|---|---|---|---|---|
| O15_MATERN_CARE | Maternal care related to the fetus and amniotic cavity and possible delivery problems | XV Pregnancy, childbirth and the puerperium (O15_) | 41941 | 168929 |
| M13_LOWBACKPAINORANDSCIATICA | Lower back pain or/and sciatica | XIII Diseases of the musculoskeletal system and connective tissue (M13_) | 41815 | 335462 |
| G6_SLEEPAPNO_INCLAVO | Sleep apnoea, including avohilmo | VI Diseases of the nervous system (G6_) | 41704 | 335573 |
| J10_ASTHMACOPDKELA | Asthma/COPD (KELA code 203) | X Diseases of the respiratory system (J10_) | 41495 | 311286 |
| E4_HYTHY_AI_STRICT | Hypothyroidism, strict autoimmune | IV Endocrine, nutritional and metabolic diseases (E4_) | 40926 | 274069 |
| AB1_INTESTINAL_INFECTIONS | Intestinal infectious diseases | I Certain infectious and parasitic diseases (AB1_) | 40881 | 336396 |
| L12_DERMATITISECZEMA | Dermatitis and eczema | XII Diseases of the skin and subcutaneous tissue (L12_) | 40688 | 336589 |
| DEATH | Any death | Common endpoint | 40455 | 336822 |
| KRA_PSY_ANXIETY_EXMORE | Anxiety disorders (more control exclusions) | Psychiatric endpoints from Katri Räikkönen | 40191 | 277526 |
| C_STROKE | STROKE | Comorbidities of Diabetes | 39818 | 271817 |
| MIGRAINE_TRIPTAN | Migraine, single triptan purchase ok & required. ICD-code if available is included | Neurological endpoints | 39387 | 337890 |
| G6_SLEEPAPNO | Sleep apnoea | VI Diseases of the nervous system (G6_) | 38998 | 336659 |
| T2D_WIDE | Type 2 diabetes, wide definition | Diabetes endpoints | 38657 | 310131 |
| N14_RENALTUB | Renal tubulo-interstitial diseases | XIV Diseases of the genitourinary system (N14_) | 38335 | 338942 |
| M13_SPONDYLOPATHY | Spondylopathies | XIII Diseases of the musculoskeletal system and connective tissue (M13_) | 38276 | 270964 |
| E4_LIPOPROT | Disorders of lipoprotein metabolism and other lipidaemias | IV Endocrine, nutritional and metabolic diseases (E4_) | 37742 | 324150 |
| M13_INTERVERTEB | Other intervertebral disc disorders | XIII Diseases of the musculoskeletal system and connective tissue (M13_) | 37636 | 270964 |
| K11_CHOLELITH | Cholelithiasis | XI Diseases of the digestive system (K11_) | 37041 | 330903 |
| N14_MESNRUIRREG | Excessive, frequent and irregular menstruation | XIV Diseases of the genitourinary system (N14_) | 36824 | 107564 |
| J10_ACUTEUPPERINFEC | Acute upper respiratory infections of multiple and unspecified sites | X Diseases of the respiratory system (J10_) | 36445 | 308166 |
| O15_ABORT_MEDICAL | Medical abortion | XV Pregnancy, childbirth and the puerperium (O15_) | 36232 | 149622 |
| K11_CONSTIPATION | Constipation | XI Diseases of the digestive system (K11_) | 36022 | 341255 |
| BLEEDING | Bleeding | Miscellaneous, not yet classified endpoints | 35917 | 341360 |
| M13_KNEEDERANGEMENTS | Internal derangement of knee | XIII Diseases of the musculoskeletal system and connective tissue (M13_) | 35511 | 240862 |
| M13_SHOULDER | Shoulder lesions | XIII Diseases of the musculoskeletal system and connective tissue (M13_) | 34594 | 275212 |
| I9_ANGINA | Angina pectoris | IX Diseases of the circulatory system (I9_) | 34456 | 313400 |
| J10_ASTHMA_MAIN_EXMORE | Asthma (only as main-diagnosis) (more control exclusions) | X Diseases of the respiratory system (J10_) | 34343 | 202399 |
| N14_URETHRAOTH | Other disorders of urethra and urinary system | XIV Diseases of the genitourinary system (N14_) | 32905 | 328001 |
| H7_CHOROIDRETINA | Disorders of choroid and retina | VII Diseases of the eye and adnexa (H7_) | 32708 | 344569 |
| H7_LIDLACRIMALORBIT | Disorders of eyelid, lacrimal system and orbit | VII Diseases of the eye and adnexa (H7_) | 32593 | 344684 |
| H8_HL_SEN_NAS | Sensorineural hearing loss | VIII Diseases of the ear and mastoid process (H8_) | 32487 | 331736 |
| K11_HERING | Inguinal hernia | XI Diseases of the digestive system (K11_) | 32335 | 322586 |
| M13_POLYARTHROPATHIES | Polyarthropathies | XIII Diseases of the musculoskeletal system and connective tissue (M13_) | 32199 | 240862 |
| CD2_BENIGN_LEIOMYOMA_UTERI | Leiomyoma of uterus | II Neoplasms from hospital discharges (CD2_) | 31661 | 179209 |
| E4_PCOS_CONCORTIUM | Polycystic ovarian syndrome, consortium definition | IV Endocrine, nutritional and metabolic diseases (E4_) | 31548 | 179322 |
| O15_PREG_OTHER_MAT_DISORD | Other maternal disorders predominantly related to pregnancy | XV Pregnancy, childbirth and the puerperium (O15_) | 30971 | 179899 |
| K11_DIVERTIC | Diverticular disease of intestine | XI Diseases of the digestive system (K11_) | 30649 | 301931 |
| M13_LIMBPAIN | Pain in limb | XIII Diseases of the musculoskeletal system and connective tissue (M13_) | 30485 | 275212 |
| K11_PARODON_OPER | Parodontitis or operation codes | XI Diseases of the digestive system (K11_) | 30377 | 346900 |
| N14_PROSTHYPERPLA | Hyperplasia of prostate | XIV Diseases of the genitourinary system (N14_) | 30066 | 119297 |
| I9_VARICVE | Varicose veins | IX Diseases of the circulatory system (I9_) | 29539 | 324121 |
| AB1_GASTROENTERITIS_NOS | Diarrhoea and gastroenteritis of presumed infectious origin | I Certain infectious and parasitic diseases (AB1_) | 29401 | 336396 |
| M13_LOWBACKPAIN | Low back pain | XIII Diseases of the musculoskeletal system and connective tissue (M13_) | 29329 | 270964 |
| H7_CONJUNCTIVITIS | Conjunctivitis | VII Diseases of the eye and adnexa (H7_) | 28895 | 345122 |
| K11_APPENDACUT | Acute appendicitis | XI Diseases of the digestive system (K11_) | 28745 | 346283 |
| I9_OTHARR | Other arrhythmias | IX Diseases of the circulatory system (I9_) | 28079 | 286109 |
| K11_PARODON_OPER_DENTAL_OPER | Parodontitis or/and operation codes | XI Diseases of the digestive system (K11_) | 27833 | 349444 |
| JOINTPAIN | Pain in joint | XIII Diseases of the musculoskeletal system and connective tissue (M13_) | 27688 | 205355 |
| J10_TONSILLECTOMY | Tonsillectomy | X Diseases of the respiratory system (J10_) | 27449 | 349828 |
| D3_ANAEMIA | Anaemias | III Diseases of the blood and blood-forming organs and certain disorders involving the immune mechanism (D3_) | 27371 | 88536 |
| I9_HEARTFAIL | Heart failure,strict | IX Diseases of the circulatory system (I9_) | 27304 | 349973 |
| I9_HEARTFAIL_EXMORE | Heart failure,strict (more control exclusions) | IX Diseases of the circulatory system (I9_) | 27304 | 328105 |
| G6_OTHNEU | Other neurological diseases | VI Diseases of the nervous system (G6_) | 27026 | 350251 |
| I9_HEARTFAIL_ALLCAUSE | All-cause Heart Failure | IX Diseases of the circulatory system (I9_) | 26872 | 349361 |
| K11_CHOLECYSTECTOMY | Cholecystectomy | XI Diseases of the digestive system (K11_) | 26778 | 350499 |
| K11_REFLUX | Gastro-oesophageal reflux disease | Interstitial lung disease endpoints | 26184 | 320387 |
| K11_APPENDECTOMY | Appendectomy | XI Diseases of the digestive system (K11_) | 26013 | 351264 |
| E4_HYPERCHOL | Pure hypercholesterolaemia | IV Endocrine, nutritional and metabolic diseases (E4_) | 25928 | 324150 |
| I9_HEARTFAIL_AND_ANTIHYPERT | Heart failure and antihypertensive medication | Cardiometabolic endpoints | 25827 | 177731 |
| I9_STR | Stroke, excluding SAH | IX Diseases of the circulatory system (I9_) | 25398 | 339920 |
| K11_CARIES_2 | Dental caries 2 | XI Diseases of the digestive system (K11_) | 25095 | 352182 |
| H8_MIDDLEMASTOID | Diseases of middle ear and mastoid | VIII Diseases of the ear and mastoid process (H8_) | 24706 | 352571 |
| F5_ALLANXIOUS | All anxiety disorders | V Mental and behavioural disorders (F5_) | 24662 | 337577 |
| L12_OTHERSKINSUBCUTIS | Other disorders of skin and subcutaneous tissue | XII Diseases of the skin and subcutaneous tissue (L12_) | 24189 | 353088 |
| I9_MI_STRICT | Myocardial infarction, strict | IX Diseases of the circulatory system (I9_) | 24185 | 313400 |
| M13_MENISCUSDERANGEMENTS | Meniscus derangement | XIII Diseases of the musculoskeletal system and connective tissue (M13_) | 24158 | 240862 |
| M13_ROTATORCUFF | Rotator cuff syndrome | XIII Diseases of the musculoskeletal system and connective tissue (M13_) | 24061 | 275212 |
| N14_PYELONEPHR | Acute tubulo-interstitial nephritis | XIV Diseases of the genitourinary system (N14_) | 23912 | 338942 |
| H7_RETINALDISOTH | Other retinal disorders | VII Diseases of the eye and adnexa (H7_) | 23610 | 344569 |

| | | | | |
|---|---|---|---|---|
| KRA_PSY_SUBSTANCE_EXMORE | Substance abuse (more control exclusions) | Psychiatric endpoints from Katri Räikkönen | 23194 | 277526 |
| I9_REVASC | Coronary revascularization (ANGIO or CABG) | IX Diseases of the circulatory system (I9_) | 23139 | 313400 |
| N14_FEMALEGENINF | Inflammatory diseases of female pelvic organs | XIV Diseases of the genitourinary system (N14_) | 22575 | 188295 |
| G6_CARPTU | Carpal tunnel syndrome | VI Diseases of the nervous system (G6_) | 22426 | 330377 |
| M13_ARTHTROSIS_COX | Coxarthrosis, | XIII Diseases of the musculoskeletal system and connective tissue (M13_) | 22254 | 240862 |
| FG_DOAAC | Diseases of arteries, arterioles and capillaries (FINNGEN) | IX Diseases of the circulatory system (I9_) | 22224 | 191924 |
| APPENDACUT_NOCOMPLIC | Acute appendicitis, no complications | XI Diseases of the digestive system (K11_) | 21998 | 346283 |
| AUD_SWEDISH | Alcohol use disorder, Swedish definition | Alcohol related diseases | 21996 | 355281 |
| L12_DERMATITISNAS | Other and unspecified dermatitis | XII Diseases of the skin and subcutaneous tissue (L12_) | 21951 | 336589 |
| ASTHMA_INFECTIONS | Asthma-related infections | Asthma and related endpoints | 21927 | 335114 |
| I9_AF_REIMB | Atrial fibrillation and flutter with reimbursement | IX Diseases of the circulatory system (I9_) | 21700 | 191924 |
| N14_MENORRHAGIA | Menorrhagia | XIV Diseases of the genitourinary system (N14_) | 21496 | 107564 |
| ASTHMA_NAS | Asthma, unspecified (mode) | Asthma and related endpoints | 21392 | 210122 |
| E4_OBESITY | Obesity | IV Endocrine, nutritional and metabolic diseases (E4_) | 21375 | 355786 |
| L12_INFECT_SKIN | Infections of the skin and subcutaneous tissue | XII Diseases of the skin and subcutaneous tissue (L12_) | 21308 | 355969 |
| H7_ALLERGICCONJUNCTIVITIS | Allergic conjunctivitis | VII Diseases of the eye and adnexa (H7_) | 20958 | 356319 |
| C3_SKIN_EXALLC | Malignant neoplasm of skin (controls excluding all cancers) | II Neoplasms, from cancer register (ICD-O-3) | 20951 | 287137 |
| M13_ARTHROSIS_OTH | Other arthrosis | XIII Diseases of the musculoskeletal system and connective tissue (M13_) | 20949 | 240862 |
| I9_VHD | Valvular heart disease excluding rheumatic fever | IX Diseases of the circulatory system (I9_) | 20929 | 286109 |
| J10_LOWERINF | Other acute lower respiratory infections | X Diseases of the respiratory system (J10_) | 20873 | 356404 |
| I9_NONRHEVALV | Non-rheumatic valve diseases | IX Diseases of the circulatory system (I9_) | 20772 | 286109 |
| N14_OVARYCYST | Ovarian cyst | XIV Diseases of the genitourinary system (N14_) | 20750 | 107564 |
| K11_REIMB_202 | KELA_REIMBURSEMENT_202 | Gastrointestinal endpoints | 20730 | 356547 |
| J10_SINUSITIS | Acute sinusitis | X Diseases of the respiratory system (J10_) | 20520 | 308166 |
| M13_SYNOTEND | Disorders of synovium and tendon | XIII Diseases of the musculoskeletal system and connective tissue (M13_) | 20492 | 275212 |
| AB1_ERYSIPELAS | Erysipelas | I Certain infectious and parasitic diseases (AB1_) | 20470 | 332343 |
| RX_CROHN_2NDLINE | Second line medication for Crohn's disease | Drug purchase endpoints | 20353 | 356924 |
| CD2_BENIGN_COLORECANI | Benign neoplasm of colon, rectum, anus and anal canal | II Neoplasms from hospital discharges (CD2_) | 20336 | 356941 |
| M13_ARTHTROSIS_COX_PRIM_ICD10 | Coxarthrosis, primary | XIII Diseases of the musculoskeletal system and connective tissue (M13_) | 20056 | 357221 |
| ST19_FRACT_LOWER_LEG_INCLU_ANKLE | Fracture of lower leg, including ankle | XIX Injury, poisoning and certain other consequences of external causes (ST19_) | 19994 | 323307 |
| M13_SPONDYLOSIS | Spondylosis | XIII Diseases of the musculoskeletal system and connective tissue (M13_) | 19892 | 270964 |
| ST19_FRACT_FOREA | Fracture of forearm | XIX Injury, poisoning and certain other consequences of external causes (ST19_) | 19577 | 351196 |
| J10_VOCALLARYNX | Diseases of vocal cords and larynx+other diseases of upper respiratory tract, no elsewhere classified | X Diseases of the respiratory system (J10_) | 19530 | 283342 |
| C3_BASAL_CELL_CARCINOMA_INCLAVO_EXALLC | Basal cell carcinoma, including avohilmo (controls excluding all cancers) | II Neoplasms, from cancer register (ICD-O-3) | 19522 | 286768 |
| H7_SCLERACORNEA | Disorders of sclera, cornea, iris and ciliary body | VII Diseases of the eye and adnexa (H7_) | 19463 | 357814 |
| I9_VTE | Venous thromboembolism | IX Diseases of the circulatory system (I9_) | 19372 | 357905 |
| I9_K_CARDIAC | Death due to cardiac causes | IX Diseases of the circulatory system (I9_) | 19295 | 357982 |
| M13_OSTEOCHONDRO | Osteopathies and chondropathies | XIII Diseases of the musculoskeletal system and connective tissue (M13_) | 19263 | 358014 |
| M13_ARTHROSIS_KNEE_PRIM_KNEESURG | Gonarthrosis, primary, with knee surgery | XIII Diseases of the musculoskeletal system and connective tissue (M13_) | 19126 | 337196 |
| M13_SPINSTENOSIS | Spinal stenosis | XIII Diseases of the musculoskeletal system and connective tissue (M13_) | 18989 | 270964 |
| C3_BASAL_CELL_CARCINOMA_EXALLC | Basal cell carcinoma (controls excluding all cancers) | II Neoplasms, from cancer register (ICD-O-3) | 18982 | 287137 |
| H7_GLAUCOMA | Glaucoma | VII Diseases of the eye and adnexa (H7_) | 18902 | 358375 |
| AB1_BACTINF_NOS | Bacterial infection, other or unspecified | I Certain infectious and parasitic diseases (AB1_) | 18576 | 332343 |
| M13_SCIATICA | Sciatica+with lumbago | XIII Diseases of the musculoskeletal system and connective tissue (M13_) | 18569 | 270964 |
| G6_MIGRAINE | Migraine | VI Diseases of the nervous system (G6_) | 18477 | 287837 |
| I9_TIA | Transient ischemic attack | IX Diseases of the circulatory system (I9_) | 18398 | 342294 |
| RX_GLUCOSAMINE | Glucosamine medication | Drug purchase endpoints | 18341 | 358936 |
| J10_COPD | COPD | X Diseases of the respiratory system (J10_) | 18266 | 311286 |
| N14_CYSTITIS | Cystitis | XIV Diseases of the genitourinary system (N14_) | 18080 | 328001 |
| C3_OTHER_SKIN_EXALLC | Other malignant neoplasms of skin (=non-melanoma skin cancer) (controls excluding all cancers) | II Neoplasms, from cancer register (ICD-O-3) | 17958 | 287137 |
| F5_DEMENTIA_INCLAVO | Dementia, including avohilmo | V Mental and behavioural disorders (F5_) | 17901 | 359376 |
| H7_CATARACTOTHER | Other cataract | VII Diseases of the eye and adnexa (H7_) | 17699 | 312864 |
| K11_ENERCOLNONINF | Noninfective enteritis and colitis | XI Diseases of the digestive system (K11_) | 17350 | 359927 |
| N14_MENSIRREG | Irregular menses | XIV Diseases of the genitourinary system (N14_) | 17228 | 179322 |
| F5_DEPRESSION_RECURRENT | Recurrent or chronic depression | V Mental and behavioural disorders (F5_) | 17227 | 228647 |
| N14_FEMGENPROL | Female genital prolapse | XIV Diseases of the genitourinary system (N14_) | 17150 | 107564 |
| PRIM_KNEEARTHROSIS | Primary gonarthrosis, bilateral | Rheuma endpoints | 17009 | 332589 |
| O15_ABORT_SPONTAN | Spontaneous abortion | XV Pregnancy, childbirth and the puerperium (O15_) | 16906 | 149622 |
| N14_BREAST | Disorders of breast | XIV Diseases of the genitourinary system (N14_) | 16897 | 193973 |
| F5_ANXIETY | Other anxiety disorders | V Mental and behavioural disorders (F5_) | 16887 | 337577 |
| I9_HEARTFAIL_AND_OVERWEIGHT | Heart failure and bmi 25plus | Cardiometabolic endpoints | 16707 | 154410 |
| F5_DEMENTIA | Dementia | V Mental and behavioural disorders (F5_) | 16499 | 356660 |
| CD2_BENIGN_COLON | Benign neoplasm: Colon | II Neoplasms from hospital discharges (CD2_) | 16445 | 360832 |
| J10_CHRONSINUSITIS | Chronic sinusitis | X Diseases of the respiratory system (J10_) | 16395 | 283342 |
| DM_SEVERAL_COMPLICATIONS | Diabetes, several complications | Diabetes endpoints | 16320 | 271817 |
| K11_OTHFUNC | Other functional intestinal disorders | XI Diseases of the digestive system (K11_) | 16311 | 301931 |
| J10_PNEUMOBACT | Bacterial pneumoniae | X Diseases of the respiratory system (J10_) | 16244 | 314673 |
| K11_OTHDIG | Other diseases of the digestive system | XI Diseases of the digestive system (K11_) | 16222 | 361055 |
| L12_DISORDSKINAPPENDIX | Disorders of skin appendages | XII Diseases of the skin and subcutaneous tissue (L12_) | 16137 | 361140 |
| H7_OCUMUSCLE | Disorders of ocular muscles, binocular movement, accommodation and refraction | VII Diseases of the eye and adnexa (H7_) | 16040 | 361237 |
| ASTHMA_ACUTE_RESPIRATORY_INFECTIONS | Asthma-related acute respiratory infections | Asthma and related endpoints | 16018 | 335114 |
| G6_HEADACHE | Other headache syndromes | VI Diseases of the nervous system (G6_) | 15851 | 287837 |
| AUD | Alcohol use disorder, ICD-based | V Mental and behavioural disorders (F5_) | 15715 | 361562 |
| H7_VISUDISTURB | Visual disturbances | VII Diseases of the eye and adnexa (H7_) | 15707 | 360091 |

| | | | | |
|---|---|---|---|---|
| C3_BREAST_EXALLC | Malignant neoplasm of breast (controls excluding all cancers) | II Neoplasms, from cancer register (ICD-O-3) | 15680 | 167189 |
| F5_KELAMENT | Kela-code for severe mental illness | V Mental and behavioural disorders (F5_) | 15451 | 360114 |
| J10_PLEURA | Other diseases of pleura | X Diseases of the respiratory system (J10_) | 15441 | 361836 |
| J10_BRONCHITIS | Acute bronchitis | X Diseases of the respiratory system (J10_) | 15393 | 356404 |
| ABDOM_HERNIA | Hernia of abdominal wall | XI Diseases of the digestive system (K11_) | 15338 | 361939 |
| E4_ENDOGLAND | Disorders of other endocrine glands | IV Endocrine, nutritional and metabolic diseases (E4_) | 15289 | 361988 |
| H7_VITRBODYGLOBE | Disorders of vitreous body | VII Diseases of the eye and adnexa (H7_) | 15177 | 361552 |
| H7_MACULADEGEN | Degeneration of macula and posterior pole | VII Diseases of the eye and adnexa (H7_) | 15137 | 344569 |
| N14_ENDOMETRIOSIS | Endometriosis | XIV Diseases of the genitourinary system (N14_) | 15088 | 107564 |
| E4_OBESITYCAL | Obesity due to excess calories | IV Endocrine, nutritional and metabolic diseases (E4_) | 15045 | 355902 |
| I9_ATHSCLE | Atherosclerosis, excluding cerebral, coronary and PAD | IX Diseases of the circulatory system (I9_) | 15002 | 349539 |
| M13_ARTHTROSIS_COX_PRIM_HIPSURG | Coxarthrosis, primary, with hip surgery | XIII Diseases of the musculoskeletal system and connective tissue (M13_) | 14990 | 357221 |
| H8_VERTIGO | Disorders of vestibular function (Vertigo) | VIII Diseases of the ear and mastoid process (H8_) | 14918 | 359094 |
| O15_HYPTENSPREG | Pregnancy hypertension | XV Pregnancy, childbirth and the puerperium (O15_) | 14727 | 196143 |
| M13_HALLUXVALGUS | Hallux valgus (acquired) | XIII Diseases of the musculoskeletal system and connective tissue (M13_) | 14726 | 240862 |
| I9_ANGIO | Coronary angioplasty | IX Diseases of the circulatory system (I9_) | 14723 | 313400 |
| H8_MED_SUPP | Suppurative and unspecified otitis media | VIII Diseases of the ear and mastoid process (H8_) | 14406 | 352571 |
| KRA_PSY_DEMENTIA_EXMORE | Any dementia (more control exclusions) | Psychiatric endpoints from Katri Räikkönen | 14367 | 277526 |
| H7_EYELIDDIS | Other disorders of eyelid | VII Diseases of the eye and adnexa (H7_) | 14347 | 344684 |
| N14_POLYPFEMGEN | Polyp of the female genital tract | XIV Diseases of the genitourinary system (N14_) | 14324 | 107564 |
| O15_FALSE_LAB | False labour | XV Pregnancy, childbirth and the puerperium (O15_) | 14282 | 168929 |
| AB1_VIRAL_SKIN_MUCOUS_MEMBRANE | Viral infections characterized by skin and mucous membrane lesions | I Certain infectious and parasitic diseases (AB1_) | 14280 | 362997 |
| N14_RENFAIL | Renal failure | XIV Diseases of the genitourinary system (N14_) | 14100 | 363177 |
| M13_DORSALGIANAS | Other/unspecified dorsalgia | XIII Diseases of the musculoskeletal system and connective tissue (M13_) | 14075 | 270964 |
| F5_STRESSOTH | Other reaction to severe stress, and adjustment disorders | V Mental and behavioural disorders (F5_) | 13913 | 337577 |
| D3_ANAEMIA_IRONDEF | Iron deficiency anaemia | III Diseases of the blood and blood-forming organs and certain disorders involving the immune mechanism (D3_) | 13689 | 360528 |
| D3_ANAEMIANAS | Other and unspecified anaemias | III Diseases of the blood and blood-forming organs and certain disorders involving the immune mechanism (D3_) | 13600 | 362319 |
| L12_ATOPIC | Atopic dermatitis | XII Diseases of the skin and subcutaneous tissue (L12_) | 13473 | 336589 |
| M13_CERVICDISC | Cervical disc disorders | XIII Diseases of the musculoskeletal system and connective tissue (M13_) | 13394 | 270964 |
| G6_AD_WIDE | Alzheimer’s disease, wide definition | VI Diseases of the nervous system (G6_) | 13393 | 363884 |
| H7_LACRIMALSYSTEM | Disorders of lacrimal system | VII Diseases of the eye and adnexa (H7_) | 13326 | 344684 |
| I9_UAP | Unstable angina pectoris | IX Diseases of the circulatory system (I9_) | 13304 | 333375 |
| TEMPOROMANDIB_INCLAVO | Temporomandibular joint disorders, including avohilmo | XI Diseases of the digestive system (K11_) | 13282 | 363995 |
| C3_PROSTATE_EXALLC | Malignant neoplasm of prostate (controls excluding all cancers) | II Neoplasms, from cancer register (ICD-O-3) | 13216 | 119948 |
| L12_PAPULOSQUAMOUS | Papulosquamous disorders | XII Diseases of the skin and subcutaneous tissue (L12_) | 13206 | 364071 |
| J10_TONSILLITIS | Other and unspecified tonsillitis | X Diseases of the respiratory system (J10_) | 13190 | 308166 |
| N14_FEMALEINFERT | Female infertility | XIV Diseases of the genitourinary system (N14_) | 13142 | 107564 |
| NEURODEGOTH | Other degenerative diseases of the nervous system | Comorbidities of Gastrointestinal endpoints | 13086 | 212179 |
| KRA_PSY_SCHIZODEL_EXMORE | Schizophrenia or delusion (more control exclusions) | Psychiatric endpoints from Katri Räikkönen | 13061 | 277526 |
| GEST_DIABETES | Gestational diabetes (for exclusion) | Diabetes endpoints | 13039 | 197831 |
| I9_HEARTFAIL_AND_CHD | Heart failure and coronary heart disease | Cardiometabolic endpoints | 12872 | 333759 |
| I9_VEINSOTH | Other disorders of veins | IX Diseases of the circulatory system (I9_) | 12630 | 324121 |
| O15_DELIV_CAESAR | Single delivery by caesarean section | XV Pregnancy, childbirth and the puerperium (O15_) | 12596 | 92306 |
| M13_RHEUMA | Rheumatoid arthritis | XIII Diseases of the musculoskeletal system and connective tissue (M13_) | 12555 | 240862 |
| CD2_BENIGN_SKIN | Other benign neoplasms of skin | II Neoplasms from hospital discharges (CD2_) | 12341 | 364936 |
| E4_HYPERLIPNAS | Hyperlipidaemia, other/unspecified | IV Endocrine, nutritional and metabolic diseases (E4_) | 12304 | 324150 |
| AB1_OTHER_SEPSIS | Other septicaemia | I Certain infectious and parasitic diseases (AB1_) | 12301 | 332343 |
| O15_OTHER_MATERN_DIS_ELSEWHERE | Other maternal diseases classifiable elsewhere but complicating pregnancy, childbirth and the puerperium | XV Pregnancy, childbirth and the puerperium (O15_) | 12297 | 198180 |
| K11_PULP_PERIAPICAL | Diseases of pulp and periapical tissues | XI Diseases of the digestive system (K11_) | 12078 | 259234 |
| ASTHMA_PNEUMONIA | Asthma-related pneumonia | Asthma and related endpoints | 12044 | 335114 |
| HYPOTHY_REIMB | Hypothyroidism, drug reimbursement | IV Endocrine, nutritional and metabolic diseases (E4_) | 12040 | 88531 |
| H7_VITROTH | Other and unspecified disorders of vitreous body | VII Diseases of the eye and adnexa (H7_) | 12004 | 361552 |
| K11_DIAHER | Diaphragmatic hernia | XI Diseases of the digestive system (K11_) | 11757 | 322586 |
| ST19_FRACT_SHOUL_UPPER_ARM | Fracture of shoulder and upper arm | XIX Injury, poisoning and certain other consequences of external causes (ST19_) | 11750 | 345173 |
| M13_SYSTCONNECT | Systemic connective tissue disorders | XIII Diseases of the musculoskeletal system and connective tissue (M13_) | 11744 | 365533 |
| G6_EPLEPSY | Epilepsy | VI Diseases of the nervous system (G6_) | 11740 | 287837 |
| O15_CONCEPT_ABNORM | Other abnormal products of conception | XV Pregnancy, childbirth and the puerperium (O15_) | 11716 | 149622 |
| J10_SEPTDEV | Deviated nasal septum | X Diseases of the respiratory system (J10_) | 11680 | 283342 |
| Z21_PROCREATIVE_MANAG | Procreative management | XXI Factors influencing health status and contact with health services (Z21_) | 11666 | 300151 |
| R18_RETEN_URINE | Retention of urine | XVIII Symptoms, signs and abnormal clinical and laboratory findings, not elsewhere classified (R18_) | 11657 | 343006 |
| H7_KERATITIS | Keratitis | VII Diseases of the eye and adnexa (H7_) | 11611 | 357814 |
| N14_POSTMENBLEED | Postmenopausal bleeding | XIV Diseases of the genitourinary system (N14_) | 11582 | 107564 |
| I9_SEQULAE | Sequelae of cerebrovascular disease | IX Diseases of the circulatory system (I9_) | 11408 | 342673 |
| ST19_FRACT_WRIST_HAND_LEVEL | Fracture at wrist and hand level | XIX Injury, poisoning and certain other consequences of external causes (ST19_) | 11394 | 336418 |
| N14_FIOTHNAS | Female infertility, cervigal, vaginal, other or unspecified origin | XIV Diseases of the genitourinary system (N14_) | 11348 | 107564 |
| H7_RETINALDETACH | Retinal detachments and breaks | VII Diseases of the eye and adnexa (H7_) | 11230 | 344569 |
| L12_ACTINKERA | Actinic keratosis | XII Diseases of the skin and subcutaneous tissue (L12_) | 11081 | 364831 |
| RHEU_ARTHRITIS_OTH | Other arthritis (FG) | Rheuma endpoints | 11033 | 285035 |
| E4_OBESITYNAS | Obesity, other/unspecified | IV Endocrine, nutritional and metabolic diseases (E4_) | 11025 | 355902 |
| ALLERG_RHINITIS | Allergic rhinitis | Comorbidities of Asthma | 11009 | 359149 |
| I9_CABG | Coronary artery bypass grafting | IX Diseases of the circulatory system (I9_) | 10938 | 313400 |
| AB1_SEXUAL_TRANSMISSION | Infections with a predominantly sexual mode of transmission | I Certain infectious and parasitic diseases (AB1_) | 10937 | 366340 |
| J10_CHRONRHINITIS | Chronic rhinitis, nasopharyngitis and pharyngitis | X Diseases of the respiratory system (J10_) | 10868 | 283342 |
| K11_LIVER | Diseases of liver | XI Diseases of the digestive system (K11_) | 10827 | 366450 |
| KRA_PSY_ANYMENTAL_SUICID_PREG_NERV_EXMORE | Any mental disorder, or suicide (or attempt), or psychic disorders complicating pregnancy, partum or puerperium or nerve system disorders (more control exclusions) | Psychiatric endpoints from Katri Räikkönen | 10811 | 276857 |
| H7_GLAUCSUSP | Glaucoma suspect | VII Diseases of the eye and adnexa (H7_) | 10758 | 358375 |
| K11_OTHDISDIG | Other diseases of the digestive system | XI Diseases of the digestive system (K11_) | 10589 | 361055 |

| | | | | |
|---|---|---|---|---|
| M13_ENTESOPATHYOTH | Other enthesopathies | XIII Diseases of the musculoskeletal system and connective tissue (M13_) | 10581 | 275212 |
| DM_RETINOPATHY_EXMORE | Diabetic retinopathy (more control exclusions) | Diabetes endpoints | 10413 | 308633 |
| COPD_LATER | Later onset COPD | COPD and related endpoints | 10404 | 161813 |
| F5_BEHAVE | Behavioural syndromes associated with physiological disturbances and physical factors | V Mental and behavioural disorders (F5_) | 10401 | 366876 |
| N14_ABNORMALBLEED | Other abnormal uterine and caginal bleeding | XIV Diseases of the genitourinary system (N14_) | 10319 | 107564 |
| ASTHMA_OBESITY | Obesity related asthma | Comorbidities of Asthma | 10306 | 335114 |
| DENTAL_TMD | TMD related pain | Dental endpoints | 10303 | 366974 |
| O15_PREEC_OR_FETGRO | Pre-eclampsia or poor fetal growth | XV Pregnancy, childbirth and the puerperium (O15_) | 10297 | 200573 |
| H8_EXTERNAL | Diseases of external ear | VIII Diseases of the ear and mastoid process (H8_) | 10272 | 367005 |
| D3_ANAEMIA_IRONDEF_NAS | Other and unspecified iron deficiency | III Diseases of the blood and blood-forming organs and certain disorders involving the immune mechanism (D3_) | 10208 | 360528 |
| L12_URTICARIA | Urticaria | XII Diseases of the skin and subcutaneous tissue (L12_) | 10175 | 364583 |
| KRA_PSY_PERSON_EXMORE | Personality disorders (more control exclusions) | Psychiatric endpoints from Katri Räikkönen | 10012 | 277522 |
| I9_CONDUCTIO | Conduction disorders | IX Diseases of the circulatory system (I9_) | 9949 | 286109 |
| K11_OTHANRECT | Other diseases of anus and rectum | XI Diseases of the digestive system (K11_) | 9932 | 301931 |
| I9_NONISCHCARDMYOP | Non-ischemic cardiomyopathy | IX Diseases of the circulatory system (I9_) | 9926 | 303607 |
| F5_ALCOHOL_DEPENDENCE | Alcohol dependence | V Mental and behavioural disorders (F5_) | 9876 | 353355 |
| K11_DENTOFACIAL_ANOMALIES | Dentofacial anomalies [including malocclusion] | XI Diseases of the digestive system (K11_) | 9866 | 259234 |
| H8_SUP_ACUTE | Acute suppurative otitis media | VIII Diseases of the ear and mastoid process (H8_) | 9850 | 352571 |
| G6_XTRAPYR | Extrapyramidal and movement disorders | VI Diseases of the nervous system (G6_) | 9833 | 367444 |
| AB1_OTHER_VIRAL | Other viral diseases | I Certain infectious and parasitic diseases (AB1_) | 9805 | 367472 |
| O15_MATERN_CARE_OTHER | Maternal care for other known or suspected fetal problems | XV Pregnancy, childbirth and the puerperium (O15_) | 9783 | 168929 |
| N14_CALCUKIDUR | Calculus of kidney and ureter | XIV Diseases of the genitourinary system (N14_) | 9713 | 366693 |
| C3_BREAST_ERPLUS_EXALLC | Malignant neoplasm of breast, HER-positive (controls excluding all cancers) | II Neoplasms, from cancer register (ICD-O-3) | 9698 | 167017 |
| E4_FLUIDELECTRO | Other disorders of fluid, electrolyte and acid-base balance | IV Endocrine, nutritional and metabolic diseases (E4_) | 9694 | 324150 |
| M13_DEFORMDORSO | Deforming dorsopathies | XIII Diseases of the musculoskeletal system and connective tissue (M13_) | 9672 | 270964 |
| ALLERG_ASTHMA | Allergic asthma (mode) | Asthma and related endpoints | 9631 | 210122 |
| I9_PAROXTAC | Paroxysmal tachycardia | IX Diseases of the circulatory system (I9_) | 9604 | 191924 |
| K11_CHRONGASTR | Chronic gastritis | XI Diseases of the digestive system (K11_) | 9570 | 320387 |
| DM_RETINA_PROLIF | Proliferative diabetic retinopathy | Diabetes endpoints | 9511 | 362581 |
| E4_NONTOXIC_THYROID | Nontoxic goitre/thyroid nodule | IV Endocrine, nutritional and metabolic diseases (E4_) | 9485 | 367792 |
| O15_HAEMORRH_EARLY_PREG | Haemorrhage in early pregnancy | XV Pregnancy, childbirth and the puerperium (O15_) | 9433 | 179899 |
| CD2_BENIGN_LIPOMATOUS | Benign lipomatous neoplasm | II Neoplasms from hospital discharges (CD2_) | 9431 | 367846 |
| K11_IBS | Irritable bowel syndrome | XI Diseases of the digestive system (K11_) | 9323 | 301931 |
| G6_ALZHEIMER | Alzheimer disease | VI Diseases of the nervous system (G6_) | 9301 | 367976 |
| L12_PSORIASIS | Psoriasis | XII Diseases of the skin and subcutaneous tissue (L12_) | 9267 | 364071 |
| I9_PULMEMB | Pulmonary embolism | IX Diseases of the circulatory system (I9_) | 9243 | 367108 |
| RHEUMA_SEROPOS_WIDE | Seropositive rheumatoid arthritis, wide | Rheuma endpoints | 9243 | 368029 |
| N14_FEMALE_GENITAL_DYSPLASIA_ALL | All dysplastic lesions of the cervix uteri, vagina or vulva | XIV Diseases of the genitourinary system (N14_) | 9227 | 201643 |
| K11_GASTRODUOULC | Gastroduodenal ulcer | XI Diseases of the digestive system (K11_) | 9216 | 320387 |
| I9_CAVS_OPERATED | Calcific aortic valvular stenosis, operated | IX Diseases of the circulatory system (I9_) | 9153 | 368124 |
| O15_MATERN_CARE_PELVIC_ABNORM | Maternal care for known or suspected abnormality of pelvic organs | XV Pregnancy, childbirth and the puerperium (O15_) | 9149 | 168929 |
| RHEUMA_SEROPOS_OTH | Other/unspecified seropositive rheumatoid arthritis | Rheuma endpoints | 9139 | 368029 |
| I9_PHLETHROMBDVTLOW | DVT of lower extremities | IX Diseases of the circulatory system (I9_) | 9109 | 324121 |
| ST19_FRACT_RIBS_STERNUM_THORACIC_SPINE | Fracture of rib(s), sternum and thoracic spine | XIX Injury, poisoning and certain other consequences of external causes (ST19_) | 9094 | 362260 |
| N14_CHRONKIDNEYDIS | Chronic kidney disease | XIV Diseases of the genitourinary system (N14_) | 9073 | 363177 |
| DENTAL_TMD_FIBRO | TMD muscular pain linked with fibromyalgia | Dental endpoints | 8995 | 368282 |
| VWXY20_SUICI_OTHER_INTENTI_SELF_H | Suicide or other Intentional self-harm | Psychiatric endpoints from Katri Räikkönen | 8978 | 368299 |
| T1D_WIDE | Type 1 diabetes, wide definition | Diabetes endpoints | 8967 | 308373 |
| F5_ALCOHOLAC | Acute alcohol intoxication | V Mental and behavioural disorders (F5_) | 8957 | 353355 |
| H7_AMD | Age-related macular degeneration (whether dry or wet) | VII Diseases of the eye and adnexa (H7_) | 8913 | 348936 |
| H7_CONJUNCTIVITISACUNONATOPIC | Conjunctivitis (acute, non atopic) | VII Diseases of the eye and adnexa (H7_) | 8907 | 345122 |
| CD2_BENIGN_MELANOCYTIC | Melanocytic naevi | II Neoplasms from hospital discharges (CD2_) | 8900 | 368377 |
| K11_FISTULA | Colonic or urogenital fistula | XI Diseases of the digestive system (K11_) | 8879 | 368398 |
| K11_FUNCDYSP | Functional dyspepsia | XI Diseases of the digestive system (K11_) | 8875 | 320387 |
| H8_OTHHEARINGLOSS | Other hearing loss | VIII Diseases of the ear and mastoid process (H8_) | 8833 | 331736 |
| ST19_FRACT_FEMUR | Fracture of femur | XIX Injury, poisoning and certain other consequences of external causes (ST19_) | 8766 | 360928 |
| N14_OTHNONINFUTER | Other noninflammatory disorders of uterus, except cervix | XIV Diseases of the genitourinary system (N14_) | 8752 | 107564 |
| OTHER_SYSTCON_FG | Other systemic involvement of connective tissue (FG) | Rheuma endpoints | 8695 | 285035 |
| M13_IMPINGEMENT | Impingement syndrome of shoulder | XIII Diseases of the musculoskeletal system and connective tissue (M13_) | 8676 | 275212 |
| K11_ILEUS | Paralytic ileus and intestinal obstruction | XI Diseases of the digestive system (K11_) | 8660 | 301931 |
| O15_COMPLIC_PUERP | Complications predominantly related to the puerperium | XV Pregnancy, childbirth and the puerperium (O15_) | 8603 | 202267 |
| PRIM_COXARTHROSIS | Primary coxarthrosis, bilateral | Rheuma endpoints | 8597 | 355023 |
| K11_HAEMORR | Haemorrhoids and perianal venous thrombosis | XI Diseases of the digestive system (K11_) | 8583 | 301931 |
| J10_INFLUENZA | All influenza | X Diseases of the respiratory system (J10_) | 8555 | 314673 |
| H7_BLEPHAROCHALASIS | Blepharochalasis | VII Diseases of the eye and adnexa (H7_) | 8547 | 344684 |
| K11_UMBHER | Umbilical hernia | XI Diseases of the digestive system (K11_) | 8510 | 322586 |
| O15_PRETERM | Preterm labour and delivery | XV Pregnancy, childbirth and the puerperium (O15_) | 8507 | 162777 |
| O15_GESTAT_HYPERT | Gestational [pregnancy-induced] hypertension | XV Pregnancy, childbirth and the puerperium (O15_) | 8502 | 194266 |
| M13_GOUT | Gout | XIII Diseases of the musculoskeletal system and connective tissue (M13_) | 8489 | 240862 |
| I9_HYPTENSHD | Hypertensive Heart Disease | IX Diseases of the circulatory system (I9_) | 8387 | 265626 |

| | | | | |
|---|---|---|---|---|
| M13_EARLY_LUMBAR_PROLAPSE | Early lumbar disc prolapse, operated | XIII Diseases of the musculoskeletal system and connective tissue (M13_) | 8351 | 368926 |
| O15_LABOUR_FETAL_STRESS | Labour and delivery complicated by fetal stress [distress] | XV Pregnancy, childbirth and the puerperium (O15_) | 8324 | 162777 |
| H8_NONSUPPNAS | Nonsuppurative otitis media | VIII Diseases of the ear and mastoid process (H8_) | 8315 | 352571 |
| H8_BPV | Benign paroxysmal vertigo | VIII Diseases of the ear and mastoid process (H8_) | 8280 | 359094 |
| O15_POSTPART_HEAMORRH | Postpartum haemorrhage | XV Pregnancy, childbirth and the puerperium (O15_) | 8249 | 162777 |
| O15_POSTPART_HEAMORRH_ALLW | Postpartum haemorrhage | XV Pregnancy, childbirth and the puerperium (O15_) | 8249 | 202621 |
| O15_POSTPART_HEAMORRH_LB | Postpartum haemorrhage | XV Pregnancy, childbirth and the puerperium (O15_) | 8249 | 146016 |
| M13_SOFTOVERUSE | Soft tissue disorders related to use, overuse and pressure | XIII Diseases of the musculoskeletal system and connective tissue (M13_) | 8236 | 275212 |
| AB1_MYCOSES | Mycoses | I Certain infectious and parasitic diseases (AB1_) | 8179 | 369098 |
| THYROTOXICOSIS | Thyrotoxicosis | Comorbidities of Gastrointestinal endpoints | 8173 | 367407 |
| L12_ABSCESS_CUT | Cutaneous abscess, furuncle and carbuncle | XII Diseases of the skin and subcutaneous tissue (L12_) | 8157 | 355969 |
| K11_EMBIMPACT_TEETH_INCLAVO | Embedded and impacted teeth , including avohilmo | XI Diseases of the digestive system (K11_) | 8139 | 369138 |
| N14_CERVICAL_DYSPLASIA_ALL | All dysplastic lesions of the cervix uteri | XIV Diseases of the genitourinary system (N14_) | 8025 | 202845 |
| K11_OTHENTERCOL | Other noninfective gastroenteritis and colitis | XI Diseases of the digestive system (K11_) | 7988 | 359927 |
| M13_FIBROBLASTIC | Fibroblastic disorders | XIII Diseases of the musculoskeletal system and connective tissue (M13_) | 7952 | 275212 |
| G6_MIGRAINE_WITH_AURA | Migraine with aura | VI Diseases of the nervous system (G6_) | 7917 | 287837 |
| O15_LABOUR_LONG | Long labour | XV Pregnancy, childbirth and the puerperium (O15_) | 7866 | 162777 |
| DM_KETOACIDOSIS | Diabetic ketoacidosis | Diabetes endpoints | 7841 | 271817 |
| H7_GLAUCOMA_POAG | Primary open-angle glaucoma, strict | VII Diseases of the eye and adnexa (H7_) | 7756 | 358375 |
| H7_REFRAACCOMMODIS | Disorders of refraction and accommodation | VII Diseases of the eye and adnexa (H7_) | 7737 | 361237 |
| K11_CARIES_DENTIN | Dentinal caries | XI Diseases of the digestive system (K11_) | 7680 | 259234 |
| O15_DELIV_FORCEP_VAC | Single delivery by forceps and vacuum extractor | XV Pregnancy, childbirth and the puerperium (O15_) | 7630 | 92306 |
| K11_VENTHER | Ventral hernia | XI Diseases of the digestive system (K11_) | 7628 | 322586 |
| K11_IBD_STRICT | Inflammatory bowel disease, strict (require KELA) | XI Diseases of the digestive system (K11_) | 7625 | 369652 |
| K11_KELAIBD | IBD patients in KELA-register | XI Diseases of the digestive system (K11_) | 7625 | 359927 |
| AB1_VIRAL_OTHER_INTEST_INFECTIONS | Viral and other specified intestinal infections | I Certain infectious and parasitic diseases (AB1_) | 7613 | 336396 |
| ST19_FRACT_FOOT_ANKLE | Fracture of foot, except ankle | XIX Injury, poisoning and certain other consequences of external causes (ST19_) | 7593 | 351392 |
| N14_ENDOMETRIOSIS_ASRM_STAGE3_4 | Endometriosis ASRM stages 3,4 | XIV Diseases of the genitourinary system (N14_) | 7574 | 203296 |
| CD2_PRIMARY_LYMPHOID_HEMATOPOIETIC_EXALLC | Primary_lymphoid and hematopoietic malignant neoplasms (controls excluding all cancers) | II Neoplasms from hospital discharges (CD2_) | 7519 | 299952 |
| K11_OTHGASTR | Other gastritis (incl. Duodenitis) | XI Diseases of the digestive system (K11_) | 7514 | 320387 |
| J10_PERITONSABSC | Peritonsillar abscess | X Diseases of the respiratory system (J10_) | 7510 | 283342 |
| F5_PSYCHOTH | Other and unspecified nonorganic psychotic disorders | V Mental and behavioural disorders (F5_) | 7508 | 364160 |
| E4_DMNAS | Unspecified diabetes | IV Endocrine, nutritional and metabolic diseases (E4_) | 7455 | 308280 |
| O15_LABOUR_MALPOS | Obstructed labour due to malposition and malpresentation of fetus | XV Pregnancy, childbirth and the puerperium (O15_) | 7414 | 162777 |
| I9_AORTANEUR | Aortic aneurysm | IX Diseases of the circulatory system (I9_) | 7395 | 349539 |
| M13_CERVICOBRACHSDR | Cervicobrachial syndrome | XIII Diseases of the musculoskeletal system and connective tissue (M13_) | 7353 | 270964 |
| DM_HYPOGLYC | Diabetic hypoglycemia | Diabetes endpoints | 7332 | 271817 |
| H7_VISUDISTURBSUB | Subjective visual disturbances | VII Diseases of the eye and adnexa (H7_) | 7313 | 360091 |
| H7_IRIDOCYCLITIS | Iridocyclitis | VII Diseases of the eye and adnexa (H7_) | 7306 | 357814 |
| M13_OSTEOPOROSIS | Osteoporosis | XIII Diseases of the musculoskeletal system and connective tissue (M13_) | 7300 | 358014 |
| M13_GANGLION | Ganglion | XIII Diseases of the musculoskeletal system and connective tissue (M13_) | 7270 | 275212 |
| CPAP | Continuous positive airway pressure | Interstitial lung disease endpoints | 7247 | 164315 |
| O15_PRE_OR_ECLAMPSIA | Pre-eclampsia or eclampsia | XV Pregnancy, childbirth and the puerperium (O15_) | 7212 | 194266 |
| H7_STRABOTH | Other strabismus | VII Diseases of the eye and adnexa (H7_) | 7179 | 361237 |
| H8_TINNITUS | Tinnitus | VIII Diseases of the ear and mastoid process (H8_) | 7175 | 331736 |
| O15_MEMBR_PREMAT_RUPT | Premature rupture of membranes | XV Pregnancy, childbirth and the puerperium (O15_) | 7147 | 168929 |
| NONALLERG_ASTHMA_EXMORE | Non-allergic asthma (mode) (more control exclusions) | Asthma and related endpoints | 7143 | 202399 |
| M13_ENTESOPATHYLOW | Enthesopathies of lower limb, excluding foot | XIII Diseases of the musculoskeletal system and connective tissue (M13_) | 7125 | 270964 |
| COPD_EARLY | Early onset COPD | COPD and related endpoints | 7079 | 359794 |
| I9_PHLETHROM | Phlebitis and thrombophlebitis (not including DVT) | IX Diseases of the circulatory system (I9_) | 7037 | 324121 |
| M13_MYALGIA | Myalgia | XIII Diseases of the musculoskeletal system and connective tissue (M13_) | 7024 | 275212 |
| CD2_BENIGN_COLON_NOS | Benign neoplasm: Colon, unspecified | II Neoplasms from hospital discharges (CD2_) | 7013 | 370264 |
| F5_BIPO | Bipolar affective disorders | V Mental and behavioural disorders (F5_) | 7006 | 329192 |
| N14_FEMGENPAIN | Pain and other conditions associated with female genital organs and menstrual cycle | XIV Diseases of the genitourinary system (N14_) | 6976 | 107564 |
| ST19_FRACT_SKULL_FACIAL_BONES | Fracture of skull and facial bones | XIX Injury, poisoning and certain other consequences of external causes (ST19_) | 6907 | 329318 |
| D3_BLOODOTHER | Other diseases of blood and blood-forming organs | III Diseases of the blood and blood-forming organs and certain disorders involving the immune mechanism (D3_) | 6877 | 370400 |
| N14_PHIMOSIS | Redundant prepuce, phimosis and paraphimosis | XIV Diseases of the genitourinary system (N14_) | 6859 | 119297 |
| K11_DISORD_TEETH | Other disorders of teeth and supporting structures | XI Diseases of the digestive system (K11_) | 6850 | 259234 |
| O15_PREG_MATERN_CARE | Maternal care for other conditions predominantly related to pregnancy | XV Pregnancy, childbirth and the puerperium (O15_) | 6845 | 179899 |
| FE | Focal epilepsy | VI Diseases of the nervous system (G6_) | 6842 | 365537 |
| H7_RETINOPATHYDIAB | Diabetic retinopathy | VII Diseases of the eye and adnexa (H7_) | 6818 | 344569 |
| F5_PERSMOOD | Persistent mood disorders | V Mental and behavioural disorders (F5_) | 6756 | 329192 |
| APPENDACUT_COMPLIC | Acute appendicitis, with complications | XI Diseases of the digestive system (K11_) | 6747 | 346283 |
| G6_MIGRAINE_NO_AURA | Migraine without aura | VI Diseases of the nervous system (G6_) | 6730 | 287837 |
| TRAUMBRAIN_NONCONCUS | severe traumatic brain injury, does not include concussion | Neurological endpoints | 6687 | 370590 |
| M13_CERVICALGIA | Cervicalgia | XIII Diseases of the musculoskeletal system and connective tissue (M13_) | 6684 | 270964 |
| L12_CELLULITIS | Cellulitis | XII Diseases of the skin and subcutaneous tissue (L12_) | 6683 | 355969 |

| | | | | |
|---|---|---|---|---|
| O15_PREECLAMPS | Pre-eclampsia | XV Pregnancy, childbirth and the puerperium (O15_) | 6663 | 194266 |
| F5_SUBSNOALCO | Substance use, excluding alcohol | V Mental and behavioural disorders (F5_) | 6605 | 353355 |
| M13_OTHERBONE | Other disorders of bone | XIII Diseases of the musculoskeletal system and connective tissue (M13_) | 6568 | 358014 |
| H8_EXTOTITIS | Otitis externa | VIII Diseases of the ear and mastoid process (H8_) | 6547 | 367005 |
| H7_IRIDOCYC_ANTER | Anterior Iridocyclitis | VII Diseases of the eye and adnexa (H7_) | 6536 | 370741 |
| I9_INTRACRA | Nontraumatic intracranial haemmorrhage | IX Diseases of the circulatory system (I9_) | 6530 | 342673 |
| F5_SCHZPHR | Schizophrenia | V Mental and behavioural disorders (F5_) | 6515 | 364160 |
| C3_COLORECTAL_EXALLC | Colorectal cancer (controls excluding all cancers) | II Neoplasms, from cancer register (ICD-O-3) | 6509 | 287137 |
| AD_LO_EXMORE | Alzheimer's disease (Late onset) (more control exclusions) | Neurological endpoints | 6489 | 170429 |
| K11_PARODON_PERIAPIC_CHRONIC | Chronic periapical parodontitis | XI Diseases of the digestive system (K11_) | 6466 | 259234 |
| RX_INFERTILITY | Medical treatment for female infertility | Drug purchase endpoints | 6446 | 204424 |
| J10_RESPOTHER | Other diseases of the respiratory system | X Diseases of the respiratory system (J10_) | 6381 | 370896 |
| L12_URTICA_NAS | Other and unspecified urticaria | XII Diseases of the skin and subcutaneous tissue (L12_) | 6381 | 364583 |
| E4_PARATHYRO | Disorders of parathyroid gland | IV Endocrine, nutritional and metabolic diseases (E4_) | 6362 | 361988 |
| M13_PATELLAOTH | Other disorders of patella | XIII Diseases of the musculoskeletal system and connective tissue (M13_) | 6333 | 240862 |
| SPONDYLOARTHRITIS | Spondyloarthritis | Comorbidities of Interstitial lung disease endpoints | 6333 | 341455 |
| NIV | Non-invasive ventilation | Interstitial lung disease endpoints | 6315 | 164315 |
| ALCOHOLACUTE10 | Acute alcohol intoxication | Alcohol related diseases | 6289 | 369838 |
| K11_SALIVARY | Diseases of salivary glands | XI Diseases of the digestive system (K11_) | 6269 | 259234 |
| K11_ANOMALI_DENTAL_ARCH_RELATIONS1_INCLAVO | Anomalies of dental arch relationship, including avohilmo | XI Diseases of the digestive system (K11_) | 6260 | 371017 |
| J10_NASALPOLYP | Nasal polyp | X Diseases of the respiratory system (J10_) | 6255 | 283342 |
| E4_DM1NASCOMP | Type 1 diabetes with other specified/multiple/unspecified complications | IV Endocrine, nutritional and metabolic diseases (E4_) | 6234 | 308280 |
| K11_ACUTPANC | Acute pancreatitis | XI Diseases of the digestive system (K11_) | 6223 | 330903 |
| FE_STRICT | Focal epilepsy, strict definition | VI Diseases of the nervous system (G6_) | 6213 | 365534 |
| ST19_FRACT_LUMBAR_SPINE_PELVIS | Fracture of lumbar spine and pelvis | XIX Injury, poisoning and certain other consequences of external causes (ST19_) | 6209 | 364504 |
| M13_ARTHROSIS_POLY | Polyarthrosis | XIII Diseases of the musculoskeletal system and connective tissue (M13_) | 6185 | 240862 |
| H7_IRIDOACUTE | Acute and subacute iridocyclitis | VII Diseases of the eye and adnexa (H7_) | 6166 | 357814 |
| F5_BEHEMOCHILD | Behavioural and emotional disorders with onset usually occurring in childhood and adolescence | V Mental and behavioural disorders (F5_) | 6160 | 371117 |
| RHEUMA_OTHER_WIDE | Other (seronegative) rheumatoid arthritis, wide | Rheuma endpoints | 6153 | 371124 |
| E4_GOITREMULTINOD | Nontoxic multinodular goitre | IV Endocrine, nutritional and metabolic diseases (E4_) | 6149 | 320703 |
| POLLENALLERG | Pollen allergy | Comorbidities of Asthma | 6118 | 368409 |
| N14_HYPERTROPHYBREAST_BOTH | Hypertrophy of breast in women and men | XIV Diseases of the genitourinary system (N14_) | 6105 | 359487 |
| DRY_AMD | Dry age-related macular degeneration (includes geographic atrophy) | VII Diseases of the eye and adnexa (H7_) | 6065 | 251042 |
| D3_IMMUNEMECHANISM | Certain disorders involving the immune mechanism | III Diseases of the blood and blood-forming organs and certain disorders involving the immune mechanism (D3_) | 6022 | 371255 |
| N14_FEMALE_GENITAL_HSIL | HSIL lesion of the cervix uteri, vagina or vulva | XIV Diseases of the genitourinary system (N14_) | 5996 | 204874 |
| C3_BREAST_ERNEG_EXALLC | Malignant neoplasm of breast, HER-negative (controls excluding all cancers) | II Neoplasms, from cancer register (ICD-O-3) | 5965 | 167017 |
| O15_MAT_CARE_MALPRESENT | Maternal care for known or suspected malpresentation of fetus | XV Pregnancy, childbirth and the puerperium (O15_) | 5957 | 168929 |
| K11_GULC | Gastric ulcer | XI Diseases of the digestive system (K11_) | 5935 | 320387 |
| K11_OTHILEUS | Other or unspecified ileus, impaction or obstruction | XI Diseases of the digestive system (K11_) | 5925 | 301931 |
| N14_ACUTERENFAIL | Acute renal failure | XIV Diseases of the genitourinary system (N14_) | 5908 | 363177 |
| I9_CARDMYO | Cardiomyopathy | IX Diseases of the circulatory system (I9_) | 5874 | 286109 |
| N14_ENDOMETRIOSIS_OVARY | Endometriosis of ovary | XIV Diseases of the genitourinary system (N14_) | 5867 | 107564 |
| ASTHMA_CHILD_EXMORE | Childhood asthma (age<16) (more control exclusions) | Asthma and related endpoints | 5865 | 202399 |
| AB1_VIRAL_NOS | Other viral diseases, not elsewhere classified | I Certain infectious and parasitic diseases (AB1_) | 5864 | 367472 |
| C3_BRONCHUS_LUNG_EXALLC | Malignant neoplasm of bronchus and lung (controls excluding all cancers) | II Neoplasms, from cancer register (ICD-O-3) | 5842 | 287137 |
| L12_SEBORRKERAT | Seborrhoeic keratosis | XII Diseases of the skin and subcutaneous tissue (L12_) | 5835 | 353088 |
| VWXY20_INTENTI_SELF_P_EXPOS_OTHER_UNSPE_CHEMIC_NOXIO_SUBST | Intentional self-poisoning by and exposure to other and unspecified chemicals and noxious substances | XX External causes of morbidity and mortality (VWXY20_) | 5811 | 371466 |
| K11_ORAL_LICHEN_PLANUS_WIDE | Oral lichen ruber planus, wide definition | XI Diseases of the digestive system (K11_) | 5791 | 371486 |
| ABDOM_HERNIA_POSTOP | Hernia of abdominal wall, postoperative | XI Diseases of the digestive system (K11_) | 5775 | 210314 |
| D3_COAGDEF_PURPUR_HAEMORRHAGIC | Coagulation defects, purpura and other haemorrhagic conditions | III Diseases of the blood and blood-forming organs and certain disorders involving the immune mechanism (D3_) | 5773 | 371504 |
| N14_ENDOMETRIOSIS_ASRM_STAGE1_2 | Endometriosis ASRM stages 1,2 | XIV Diseases of the genitourinary system (N14_) | 5769 | 205101 |
| N14_SALPHOOPH | Salpingitis and oophoritis | XIV Diseases of the genitourinary system (N14_) | 5768 | 188295 |
| M13_SPONDYLOLISTHESIS | Spondylolisthesis/Spondylolysis | XIII Diseases of the musculoskeletal system and connective tissue (M13_) | 5766 | 270964 |
| L12_PSORI_VULG | Psoriasis vulgaris | XII Diseases of the skin and subcutaneous tissue (L12_) | 5759 | 364071 |
| AB1_SEXUAL_TRANSMISSION_NOS | Other predominantly sexually transmitted diseases, not elsewhere classified | I Certain infectious and parasitic diseases (AB1_) | 5753 | 366340 |
| TEMPOROMANDIB | Temporomandibular joint disorders | XI Diseases of the digestive system (K11_) | 5668 | 205355 |
| O15_PREG_ECTOP | Ectopic pregnancy | XV Pregnancy, childbirth and the puerperium (O15_) | 5648 | 149622 |
| N14_GLOMERULAR | Glomerular diseases | XIV Diseases of the genitourinary system (N14_) | 5642 | 371635 |
| AB1_BACT_INTEST_OTH | Other bacterial intestinal infections | I Certain infectious and parasitic diseases (AB1_) | 5631 | 336396 |
| N14_ENDOMETRIOSIS_PELVICPERITONEUM | Endometriosis of pelvic peritoneum | XIV Diseases of the genitourinary system (N14_) | 5628 | 107564 |
| M13_HAMMERTOE | Other hammer toe(s) (acquired) | XIII Diseases of the musculoskeletal system and connective tissue (M13_) | 5625 | 240862 |
| CD2_BENIGN_RECTUM_ANAL | Benign neoplasm: Rectum, anus and anal canal | II Neoplasms from hospital discharges (CD2_) | 5613 | 371664 |
| E4_HYPERPARA | Hyperparathyroidism | IV Endocrine, nutritional and metabolic diseases (E4_) | 5590 | 361988 |
| G6_POLYOTHUNS | Other and unspecified polyneuropathies, also in other diseases | VI Diseases of the nervous system (G6_) | 5553 | 370790 |
| M13_RADICULOPATHY | Radiculopathy | XIII Diseases of the musculoskeletal system and connective tissue (M13_) | 5550 | 270964 |
| M13_ADHCAPSULITIS | Adhesive capsulitis of shoulder | XIII Diseases of the musculoskeletal system and connective tissue (M13_) | 5538 | 275212 |
| I9_AVBLOCK | AV-block | IX Diseases of the circulatory system (I9_) | 5536 | 286109 |

| | | | | |
|---|---|---|---|---|
| K11_EROSION_INCLAVO | Erosion of teeth, including avohilmo | XI Diseases of the digestive system (K11_) | 5521 | 371756 |
| H7_CORNULCER | Corneal ulcer | VII Diseases of the eye and adnexa (H7_) | 5476 | 357814 |
| L12_FOLLICULARCYST | Follicular cysts of skin and subcutaneous tissue | XII Diseases of the skin and subcutaneous tissue (L12_) | 5459 | 361140 |
| N14_CERVICAL_HSIL | HSIL lesion of the cervix uteri | XIV Diseases of the genitourinary system (N14_) | 5459 | 205411 |
| K11_IMPACTED_TEETH_INCLAVO | Impacted teeth , including avohilmo | XI Diseases of the digestive system (K11_) | 5435 | 371842 |
| F5_ALZHDEMENT | Dementia in Alzheimer disease | V Mental and behavioural disorders (F5_) | 5422 | 356660 |
| C3_COLORECTAL_ADENO_EXALLC | Colorectal adenocarcinoma (controls excluding all cancers) | II Neoplasms, from cancer register (ICD-O-3) | 5378 | 287137 |
| I9_SAHANEUR | Aneurysms, operations, SAH | IX Diseases of the circulatory system (I9_) | 5342 | 342673 |
| CHILDHOOD_ALLERGY | Childhood allergy (age < 16) | Asthma and related endpoints | 5337 | 371940 |
| CD2_BENIGN_OVARY | Benign neoplasm of ovary | II Neoplasms from hospital discharges (CD2_) | 5308 | 205562 |
| G6_ROOTPLEXOTHUNS | Other and unspecified nerve root and plexus disorders, also in other diseases | VI Diseases of the nervous system (G6_) | 5294 | 330377 |
| GOUT_NOS | Gout, unspecified | Rheuma endpoints | 5292 | 368788 |
| KRA_PSY_SLEEP_NONORG_EXMORE | Nonorganic sleeping disorders (more control exclusions) | Psychiatric endpoints from Katri Räikkönen | 5254 | 277526 |
| N14_HYDROCELE | Hydrocele | XIV Diseases of the genitourinary system (N14_) | 5221 | 119297 |
| N14_HYPERTROPHYBREAST | Hypertrophy of breast | XIV Diseases of the genitourinary system (N14_) | 5212 | 193973 |
| E4_DM1OPTH | Type 1 diabetes with ophthalmic complications | IV Endocrine, nutritional and metabolic diseases (E4_) | 5202 | 308280 |
| F5_SOMATOFORM | Somatoform disorder | V Mental and behavioural disorders (F5_) | 5164 | 337577 |
| E4_HYPOOSMNAT | Hypo-osmolality and hyponatraemia | IV Endocrine, nutritional and metabolic diseases (E4_) | 5137 | 324150 |
| INFLUENZA | All influenza (not pneumonia) | Asthma and related endpoints | 5136 | 314673 |
| M13_DUPUTRYEN | Palmar fascial fibromatosis [Dupuytren] | XIII Diseases of the musculoskeletal system and connective tissue (M13_) | 5128 | 275212[1] |
| N14_OTHFEMPELINF | Other female pelvic inflammatory diseases | XIV Diseases of the genitourinary system (N14_) | 5117 | 188295 |
| VOCALCORDDYS | Vocal cord dysfunction | COPD and related endpoints | 5087 | 139795 |
| CD2_BENIGN_OTHERDIGESTIVE | Benign neoplasm of other and ill-defined parts of digestive system | II Neoplasms from hospital discharges (CD2_) | 5062 | 372215 |
| K11_DISLIVOTH | Other diseases of liver | XI Diseases of the digestive system (K11_) | 5052 | 366450 |
| K11_UC_STRICT2 | Ulcerative colitis (strict definition, require KELA, min 2 HDR) | XI Diseases of the digestive system (K11_) | 5034 | 371530 |
| L12_LICHENPLANUS_INCLAVO | Lichen planus, including avohilmo | XII Diseases of the skin and subcutaneous tissue (L12_) | 5015 | 372262 |

### PGC DEs

| Author | DE | Download URL | PMID | Build # | Ancestry |
|---|---|---|---|---|---|
| Ditte Demontis | ADHD | https://figshare.com/articles/dataset/adhd2022/22564390 | 36702997 | GRCh37 | European |
| Douglas P. Wightman | AD | https://ctg.cncr.nl/software/summary_statistics | 34493870 | GRCh37 | European |
| Jakob Grove | ASD | https://figshare.com/articles/dataset/asd2019/14671989 | 30804558 | GRCh37 | European |
| Niamh Mullins | BIP | https://figshare.com/articles/dataset/PGC3_bipolar_disorder_GWAS_summary_statistics/14102594 | 34002096 | GRCh37 | European |
| OCGAS | OCD | https://figshare.com/articles/dataset/ocd2018/14672103 | 28761083 | GRCh37 | European |
| Vassily Trubetskoy | SCZ | https://figshare.com/articles/dataset/scz2022/19426775 | 35396580 | GRCh37 | European |

### References


1. Wen, J. *et al.* Convolutional neural networks for classification of Alzheimer's disease: Overview and reproducible evaluation. *Medical Image Analysis* **63**, 101694 (2020).
2. Wen, J. *et al.* The genetic architecture of multimodal human brain age. *Nat Commun* **15**, 2604 (2024).
3. The MULTI Consortium *et al.* MRI-based multi-organ clocks for healthy aging and disease assessment. *Nat Med* 1–11 (2025) doi:10.1038/s41591-025-03999-8.
4. Wen, J. Refining the generation, interpretation and application of multi-organ, multi-omics biological aging clocks. *Nat Aging* 1–17 (2025) doi:10.1038/s43587-025-00928-9.
5. The MULTI Consortium *et al.* Multi-organ metabolome biological age implicates cardiometabolic conditions and mortality risk. *Nat Commun* **16**, 4871 (2025).
6. Wen, J. *et al.* Subtyping Brain Diseases from Imaging Data. in *Machine Learning for Brain Disorders* (ed. Colliot, O.) 491–510 (Springer US, New York, NY, 2023). doi:10.1007/978-1-0716-3195-9_16.
7. Wen, J. *et al.* Neuroimaging endophenotypes reveal underlying mechanisms and genetic factors contributing to progression and development of four brain disorders. *Nat. Biomed. Eng* 1–18 (2025) doi:10.1038/s41551-025-01412-w.
8. The MULTI Consortium *et al.* Multi-organ AI endophenotypes chart the heterogeneity of brain, eye and heart pan-disease. *Nat. Mental Health* 1–28 (2026) doi:10.1038/s44220-025-00560-x.
9. Hannum, G. *et al.* Genome-wide Methylation Profiles Reveal Quantitative Views of Human Aging Rates. *Molecular Cell* **49**, 359–367 (2013).
10. Horvath, S. DNA methylation age of human tissues and cell types. *Genome Biology* **14**, 3156 (2013).

11. Cole, J. H. *et al.* Brain age predicts mortality. *Mol Psychiatry* **23**, 1385–1392 (2018).

12. Oh, H. S.-H. *et al.* Organ aging signatures in the plasma proteome track health and disease. *Nature* **624**, 164–172 (2023).

13. Levine, M. E. *et al.* An epigenetic biomarker of aging for lifespan and healthspan. *Aging (Albany NY)* **10**, 573–591 (2018).

14. Lu, A. T. *et al.* DNA methylation GrimAge strongly predicts lifespan and healthspan. *Aging (Albany NY)* **11**, 303–327 (2019).

15. Lu, A. T. *et al.* DNA methylation GrimAge version 2. *Aging (Albany NY)* **14**, 9484–9549 (2022).

16. Belsky, D. W. *et al.* DunedinPACE, a DNA methylation biomarker of the pace of aging. *Elife* **11**, e73420 (2022).

17. Goeminne, L. J. E. *et al.* Plasma protein-based organ-specific aging and mortality models unveil diseases as accelerated aging of organismal systems. *Cell Metab* **37**, 205-222.e6 (2025).

18. Whitman, E. T. *et al.* DunedinPACNI estimates the longitudinal Pace of Aging from a single brain image to track health and disease. *Nat Aging* **5**, 1619–1636 (2025).

19. Salimi, S. *et al.* Health octo tool matches personalized health with rate of aging. *Nat Commun* **16**, 4007 (2025).

20. Tyshkovskiy, A. *et al.* Universal transcriptomic hallmarks of mammalian ageing and mortality. *Nature* **654**, 173–188 (2026).
